# Landscape and Heterogeneity of Treatment Outcome Measures in Facioscapulohumeral Muscular Dystrophy: A Systematic Literature Review

**DOI:** 10.1101/2025.08.29.25334711

**Authors:** Lucia Zhu, Katy Eichinger, Lianne Barnieh, Rachel Beckerman, Adi Eldar-Lissai

**Author notes:** **Previous Presentation of the Work:** Part of this work was presented as a poster at ISPOR (International Society for Pharmacoeconomics and Outcomes Research) 2023, the FSHD International Research Congress 2023, and the International Annual Congress of the World Muscle Society 2023. **Ethical Publication Statement:** The authors confirm that they have read the Journal’s position on issues involved in ethical publication and affirm that this report is consistent with those guidelines. Data Availability: This study did not generate new primary datasets. **Ethics Approval:** This study did not involve human or animal subjects. **Patient Consent:** Not applicable. **Permission to Reproduce Material from Other Sources:** This study did not use copyrighted materials by other parties. **Clinical Trial Registration:** Not applicable.

## Abstract

Facioscapulohumeral muscular dystrophy is a rare genetic disorder that manifests as progressive weakening and loss of skeletal muscles, with the face, shoulder girdle, and upper arms typically affected in early stages. Currently, there are no approved therapies, and no consensus on the outcome measures used to assess disease progression or effects of new interventions. This systematic literature review identified treatment outcomes used to date, and the extent of their psychometric validation. Electronic searches were executed from inception to October 2022, supplemented with manual searches of relevant conferences. Eligible studies evaluated patients with the disease; reported on treatment outcomes or their validation; were clinical trials or observational studies; and published in English. Across the 65 studies identified reporting on treatment outcomes, 89 measures were used to assess outcomes following interventions in patients with the disease, including 58 efficacy/effectiveness, 6 safety, and 25 humanistic outcomes. Only 22/89 treatment outcomes were previously validated. Measures of motor function, upper/lower limb function, and muscle strength were the most frequently used efficacy and effectiveness measures. Significant heterogeneity exists among outcomes used. A total of 38 full text articles and 15 conference abstracts reported data on the humanistic burden in FSHD. The most reported domains included general QoL, followed by fatigue then pain. QoL was assessed in both adult and pediatric FSHD patients, with some instruments developed exclusively for use among children and adolescents (for example, Kidscreen for general QoL and the FSHD-HI Peds for disease-specific QoL). Based on the available evidence, it is evident that FSHD impacts QoL, with pain and/or fatigue impacting QoL. 18 studies were identified that reported on economic burden. While limited, the available evidence suggests that FSHD is associated with a substantial economic burden for patients, their caregivers and society.

Overall, the findings of this SLR indicate that despite being the second most prevalent form of muscular dystrophy, FSHD is not well characterized in the literature. Further effort is needed to build consensus around the most relevant measures, such that trials for emerging interventions are designed with the consistent evaluation of patient-relevant benefits in mind.

## Introduction

Facioscapulohumeral muscular dystrophy (FSHD) is a rare genetic muscle disorder that manifests itself as a progressive weakening and loss of skeletal muscles.^1^ FSHD is caused by aberrant expression of the gene Double Homeobox 4 (*DUX4*) in skeletal muscle, resulting in the inappropriate presence of *DUX4* protein. *DUX4*-driven gene expression is usually limited to the early embryonic development stage, after which the *DUX4* gene is silenced. In patients with FSHD, the *DUX4* gene is unsilenced as a result of a genetic mutation. The aberrant DUX4 expression leads to the death of muscle and its replacement by fat, resulting in skeletal muscle weakness and progressive disability.^2^ FSHD is divided into type 1 and type 2 based on genetic mutation, although both types are clinically alike.^3^

At disease onset, muscles of the face (facio), shoulder girdle (scapulo), and upper arms (humeral) are primarily affected. As the disease progresses, muscle weakness and loss spread to the trunk and lower body,^2^ although magnetic resonance imaging (MRI) studies have shown that muscles of the trunk and legs may also be affected early in the disease before observable muscle weakness.^4, 5^ FSHD leads to significant physical limitations, including facial weakness, difficulty using arms, and challenges with mobility. Approximately 20% of FSHD-affected individuals eventually require a wheelchair.^6^

Around 1 in 8,000 individuals worldwide are affected by FSHD,^7^ and the prevalence of FSHD in the United States (US) is between 16,000 to 38,000 individuals.^8^ It is estimated that two-thirds of cases are inherited, and the remaining one-third are caused by sporadic mutations that are not due to family history of the disease. FSHD equally affects both genders and all ethnic groups.^2^

There are currently no approved therapies for FSHD. Disease management focuses on supportive treatments with physical therapy playing a key role.^6^ To date, only three guidelines on the evaluation, diagnosis, or management of FSHD have been published.^9–11^ One is multinational and was issued in 2010 following the 171^st^ International Workshop on FSHD^11^; one was issued from the French Muscular Dystrophy Association in France in 2011^9^; and the most recent one was issued from the American Academy of Neurology in 2015.^10^ Overall, a holistic approach to FSHD management is recommended, including strength training, exercise, pain medications, orthopedic and ophthalmic care, social management, as well as surgical intervention in select patients.^9, 10, 12^

Currently, there is no consensus in the field on the outcome measures that should be used to assess disease progression, and validation of disease outcome measures is limited. Similarly, there is a lack of consensus on established endpoints for clinical trials designed to assess the treatment effects of new interventions. The objective of this systematic literature review (SLR) was to identify the measures used to assess treatment outcomes in patients with FSHD, highlighting outcome measures that have been previously validated.

## Methods

The SLR was conducted according to standard methodology, per the guideline provided by the University of York Centre for Reviews and Dissemination’s “Guidance for Undertaking Reviews in Health Care”.^13^ The results of this review are reported per the Preferred Reporting Items for Systematic Literature Reviews and Meta-Analyses (PRISMA) statement (2020).^14^

A comprehensive search was conducted in Embase (via Embase.com), MEDLINE (via PubMed.gov), and Cochrane (via cochranelibrary.com) databases. The search strategy was assembled by researching appropriate terms (both subject headings, i.e., Medical Subject Headings [MeSH] and Emtree terms, and text words as appropriate) for inclusion in the literature search strategy. The complete search strategy is presented in **Table Appendix 1** to **Table Appendix 3** of the **Supplementary Material A**. The searches were conducted from database inception to October 11, 2022, were not restricted by geographical location, and were restricted to publications in English. In addition, hand search of non-indexed conference proceedings from the Muscular Dystrophy Association (MDA), Muscle Study Group (MSG), and the FSHD Society International Research Congress (FSHD IRC) was conducted using free-text terms for the years available as of the search date, limited to the last three years prior the search (2020 to 2022).

Following removal of duplicates, two independent investigators screened all citations identified in the literature search using predetermined population, intervention, comparator, outcomes, and study design (PICOS) criteria, at both the title/abstract and full-text screening phases. A detailed list of the PICOS criteria is presented in **Table Appendix 4** of the **Supplementary Material A**. In brief, eligible studies included adults, adolescents, or children with FSHD type 1 or type 2; studies reporting on treatment outcome measures or validation of disease outcome measures; clinical trials, observational studies (including case series and case studies), or SLRs. Eligible studies were extracted by a single investigator and were independently verified by a second investigator.^13^ For all studies, the following elements were extracted: author, year, study design, geographic scope, intervention type and name, FSHD population (type 1, type 2, not specified), and proportion of pediatric patients. For treatment outcomes studies, the names of the treatment outcome measures used were extracted. For validation of disease outcome measures studies, the names of the validated outcomes along with the validation method used were extracted. At both the literature screening and data extraction phases, discrepancies between both investigators were discussed until a consensus was met, with unresolved conflicts decided by a third investigator.

Quality assessment of each included full-text article was conducted by a single investigator.^13^ Multiple tools were used, as applicable by study design, including the SIGN checklist for cohort studies^15^; the Downs and Black checklist for randomized and non-randomized trials^16^; the Joanna Briggs Institute (JBI) checklist for case series^17^; the JBI checklist for case studies^18^; the JBI checklist for qualitative research^19^; the JBI checklist for cross-sectional studies^18^; and the COSMIN tool for studies on reliability or measurement error of outcome measure instruments.^20^

## Results

### Study Selection and Characteristics

The electronic database searches, in addition to the hand searching of the conference websites, yielded 2,211 full-text articles and 754 conference abstracts after removal of duplicates. From these, a total of 1,990 full-text articles and 717 conference abstracts were excluded at the title and abstract screening stage. Following full-text review, 14 conference abstracts, 38 full-text articles, and 13 registers of clinical trials reporting on treatment outcome measures in FSHD were included, while 27 conference abstracts and 28 full-text articles reporting on validation of disease outcome measures were included. For each topic, a PRISMA flow diagram of studies identified and screened for inclusion is provided (**Figure 1-Figure 4**).

**Figure 1:**
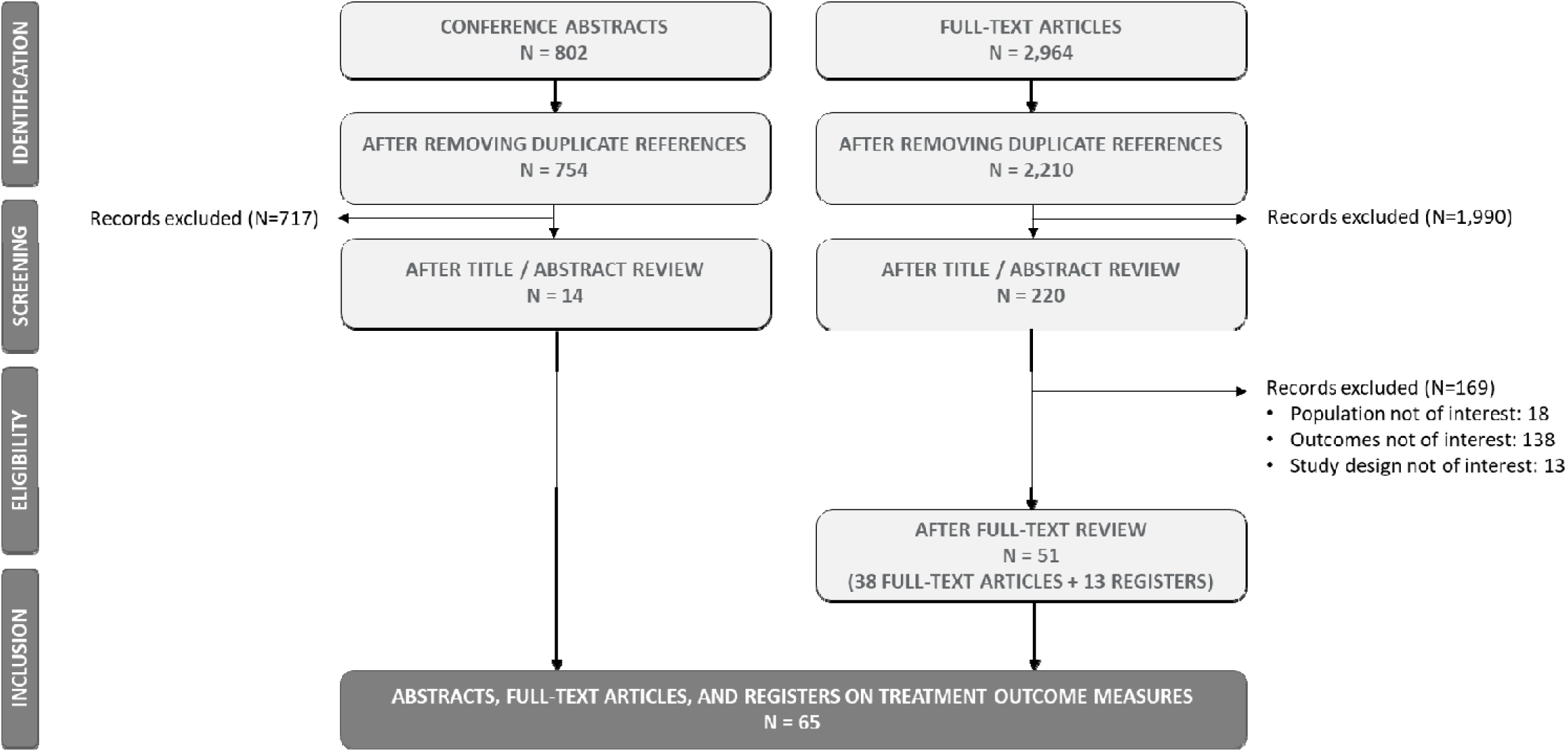
PRISMA Flow Diagram for Treatment Outcome Studies.

**Figure 2:**
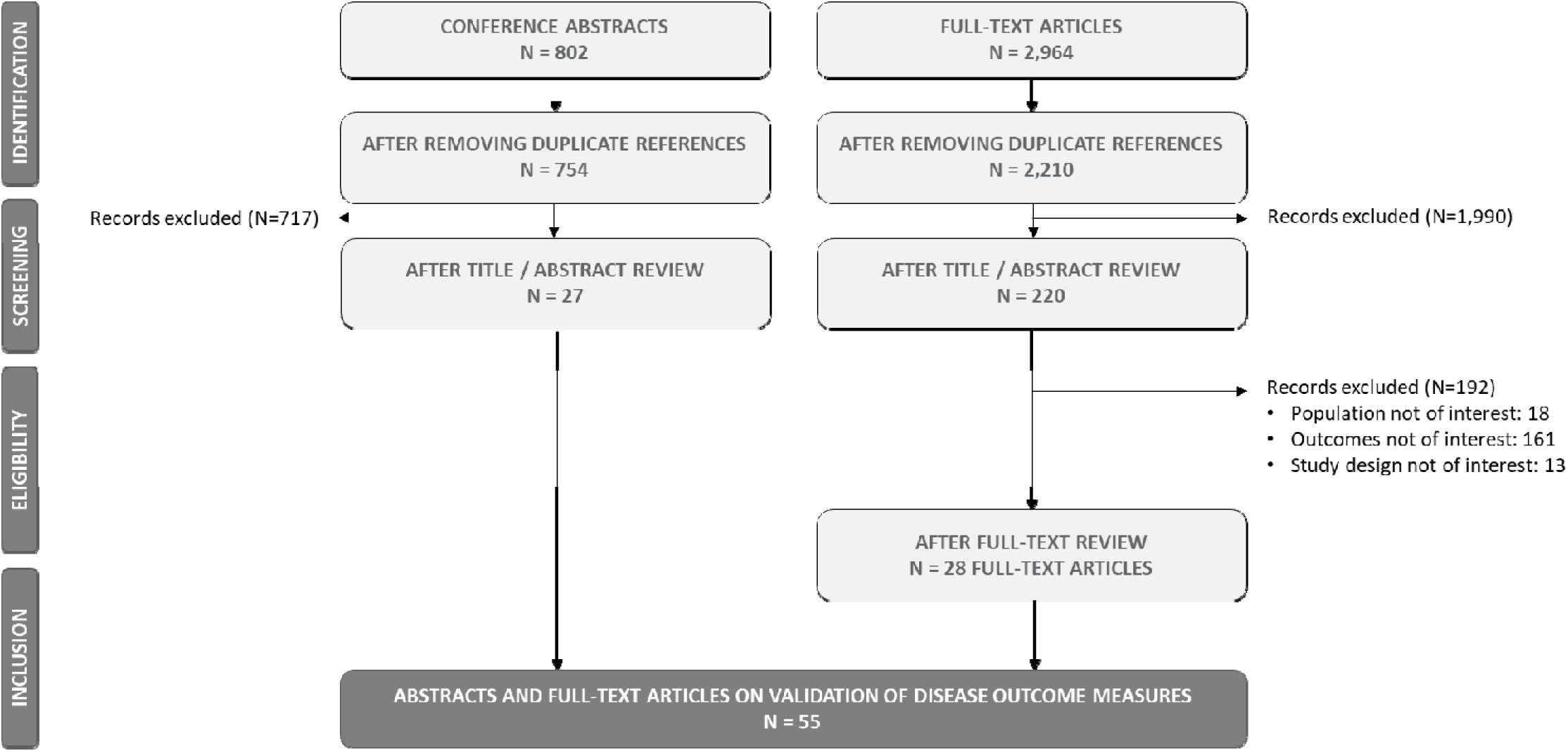
PRISMA Flow Diagram for Validation of Disease Outcome Measures Studies.

**Figure 3:**
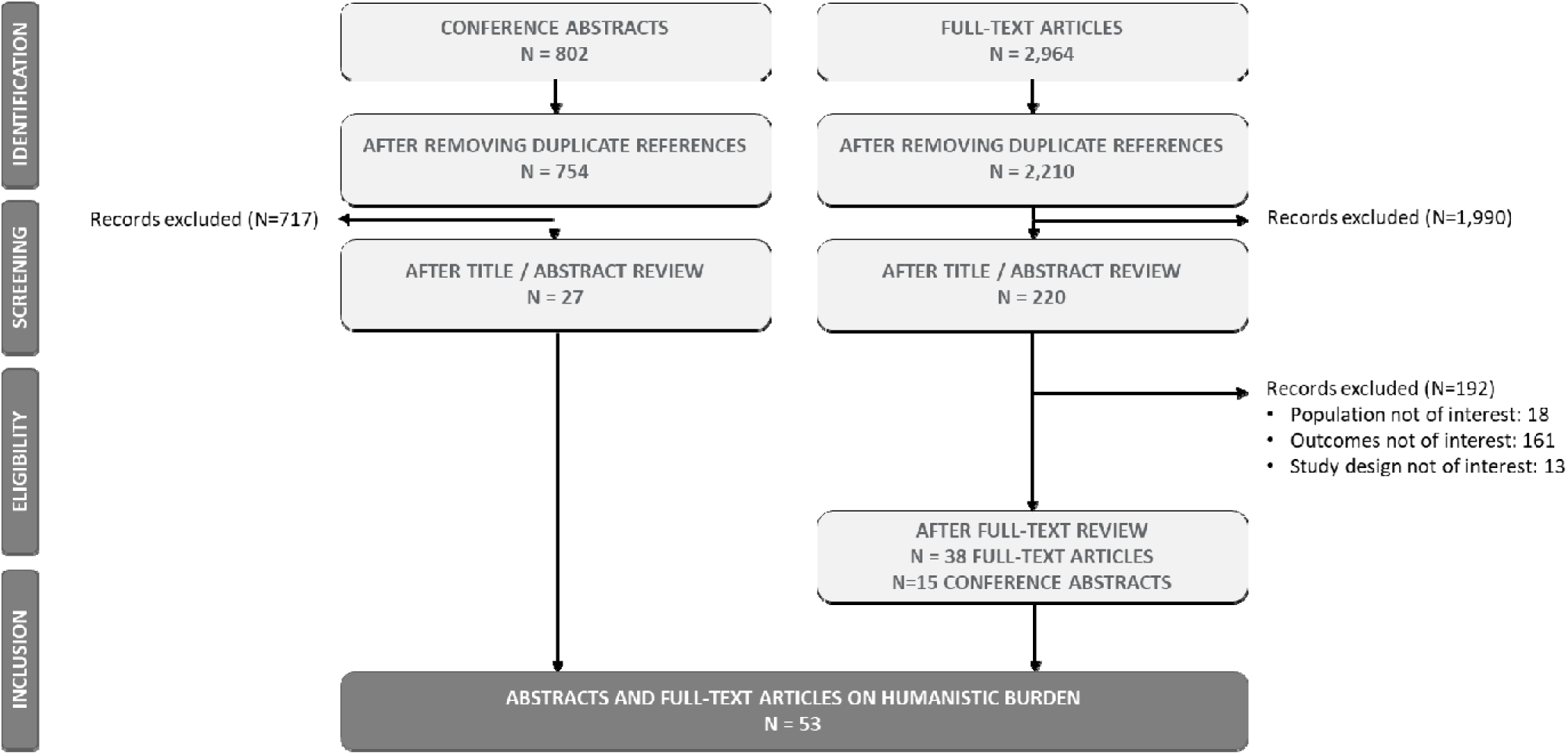
PRISMA Flow Diagram for Validation of Humanistic Burden Studies.

**Figure 4:**
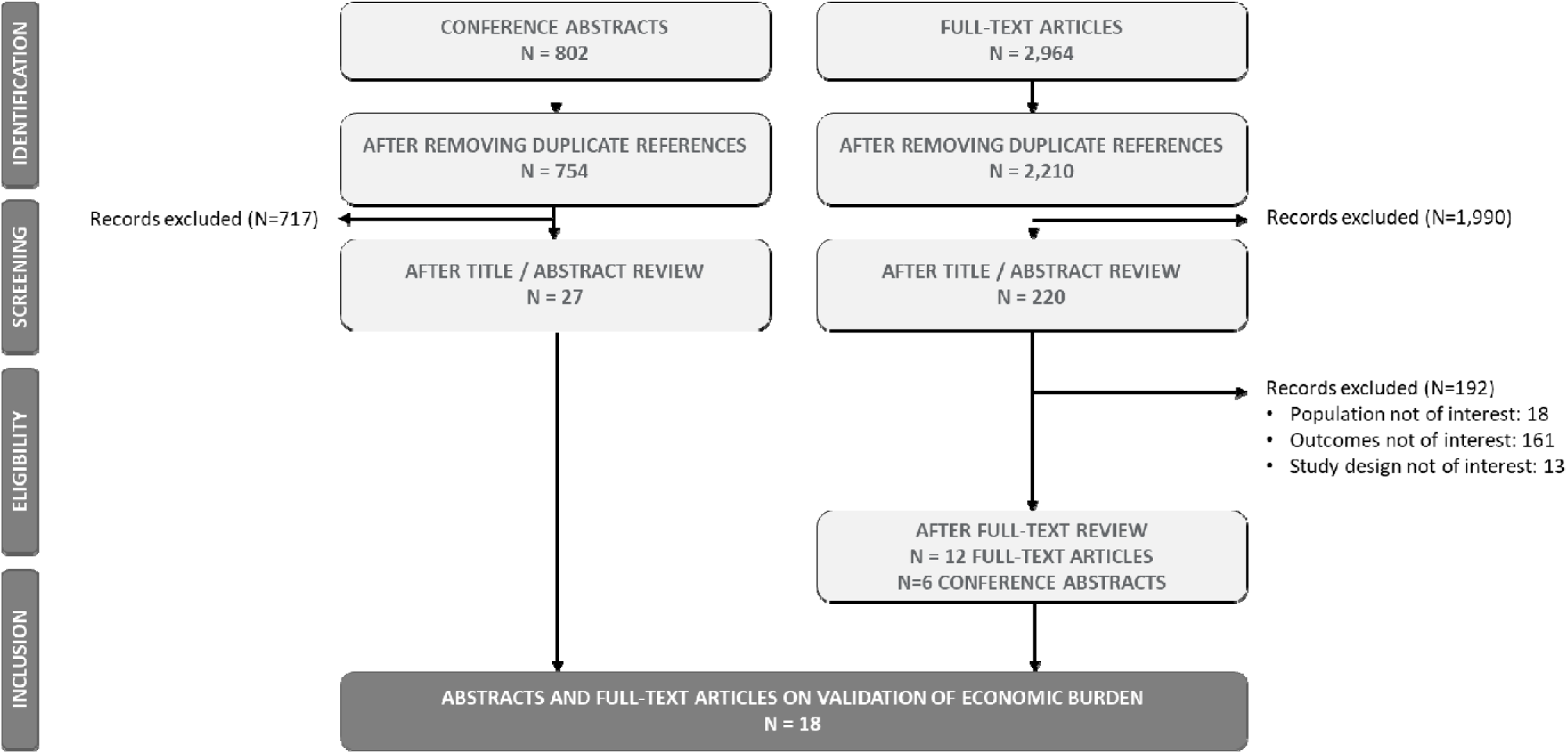
PRISMA Flow Diagram for Validation of Economic Burden Studies.

### Risk of Bias Assessment

Quality assessment was conducted for all full-text articles, with details per study presented in the **Supplementary Material B**. One full-text article reporting on treatment outcomes was not assessed as it was a narrative study.^21^

For treatment outcomes studies, the JBI checklists for case series and case studies were used for 15 studies and do not provide an overall study quality assessment score. Among the 19 randomized and non-randomized studies assessed with the Downs and Black checklist, the mean methodological quality score was 17.8 (range 12 to 24) out of a possible 28. Two studies were assessed using the SIGN cohort checklist; the overall quality was “acceptable” in one study^22^ but “low quality” in the other one.^23^ One qualitative study was assessed using the JBI checklist for qualitative studies.^24^ While the study was assessed as “include” in the overall appraisal, it did not include a statement addressing the role of the researcher and whether the beliefs and values have a potential influence on the study.

Most validation of disease outcome measures studies (25/28 studies) were assessed with the COSMIN tool for studies on reliability or measurement error of outcome measure instruments, which do not provide an overall study quality assessment score. The two cross-sectional studies assessed with the JBI checklist for cross-sectional studies were deemed to have sufficient quality to include.^25, 26^ Lastly, one study assessed with the SIGN cohort checklist had an “acceptable” overall quality.^27^

### Treatment Outcome Measures in FSHD

A total of 65 studies reported on treatment outcomes in patients with FSHD. Among full-text articles, the most common study design was observational studies (n=16), followed by clinical trials (n=13), case studies (n=8); of note, there was one qualitative study. The most frequently identified study design among conference abstracts was clinical trials (n=10), followed by observational studies (n=3) and a single case study. Among all full-text articles and conference abstracts, 9 studies exclusively assessed patients with FSHD type 1^28–36^ with sample sizes ranging from 1^35^ to 80^29^; the remaining studies did not specify the FSHD type. Only six studies included pediatric patients,^35–40^ of which three studies exclusively evaluated pediatric patients.^35–37^ Of note, two of these three were case studies with a total of three pediatric patients included.^35, 36^ Of the 13 registers of clinical trials, 2 were conducted in patients with FSHD type 1^41, 42^ and none were reported to include pediatric patients.

All included studies were categorized into one of four intervention types: pharmacological, physiological (exercise), surgical, or other. Of 65 studies identified, a total of 28 studies reported on pharmacological interventions, which included both disease-modifying therapies as well as nutritional supplements (details per study in **Table Appendix 5** of the **Supplementary Material A**). One study explored both pharmacological and physiological training and was included amongst the pharmacological interventions.^43^ Among the 18 studies included as physiological, interventions included strength training, high-intensity training, or aerobic conditioning. A total of 15 studies were identified exploring surgical interventions (either scapulothoracic arthrodesis or scapulopexy). Four studies met the inclusion criteria of the SLR but did not fit into the previous three categories: one explored the impact of a Chinese herbal medicine in a single patient with FSHD type 1^35^; one explored the impact of acupuncture in one patient with FSHD^44^; one assessed the intervention vibratory proprioceptive assistance among 8 patients^45^; and one explored the use of autologous stem cells in 13 patients.^46^

### An overview of the study design by intervention type is presented in Figure 3: PRISMA Flow Diagram for Validation of Humanistic Burden Studies

Figure 5. Most studies assessing pharmacological or physiological interventions were clinical trials, while observational studies were most predominant among studies evaluating surgical interventions.

**Figure 5:**
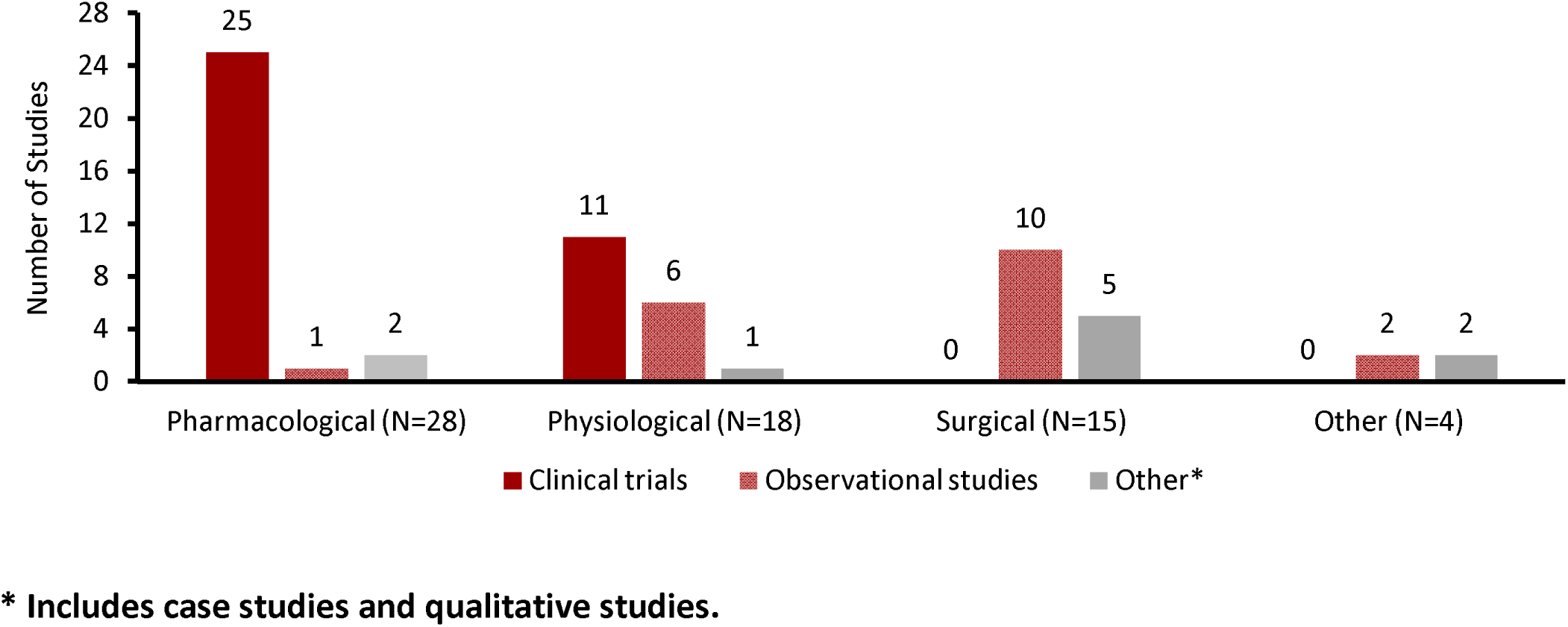
Study Design by Intervention for Treatment Outcomes Studies.

Treatment outcome measures used among the 65 studies included those used to evaluate efficacy and effectiveness, safety, and humanistic (i.e., patient-reported outcomes [PROs] such as patient functional status, quality of life (QoL), satisfaction, symptoms such as pain, fatigue, etc.) outcomes of an intervention. The list of outcomes reported in each study is presented in **Table Appendix 5** of the **Supplementary Material A.**

A total of 89 measures were identified across all studies. A summary of the number of outcomes identified by study design and intervention across all studies is presented in **Figure 6**, reflecting the high variety of measures being used in FSHD to assess treatment outcomes. Overall, the number of outcomes per study design and intervention reflected the number of studies identified in each group. As such, the overall heterogeneity in outcome measures across all studies was mainly driven by the high number of outcomes used in clinical trials of both pharmacological and physiological interventions (**Figure 6**), given the higher number of studies identified in these groups. However, when different study designs were compared, the number of outcomes in observational studies was proportionally higher than that in clinical trials and studies with other designs, although the difference was small (clinical trials: 68 outcomes from 36 studies; observational studies: 39 outcomes from 19 studies; other [case studies and qualitative studies]: 17 outcomes from 10 studies).

**Figure 6:**
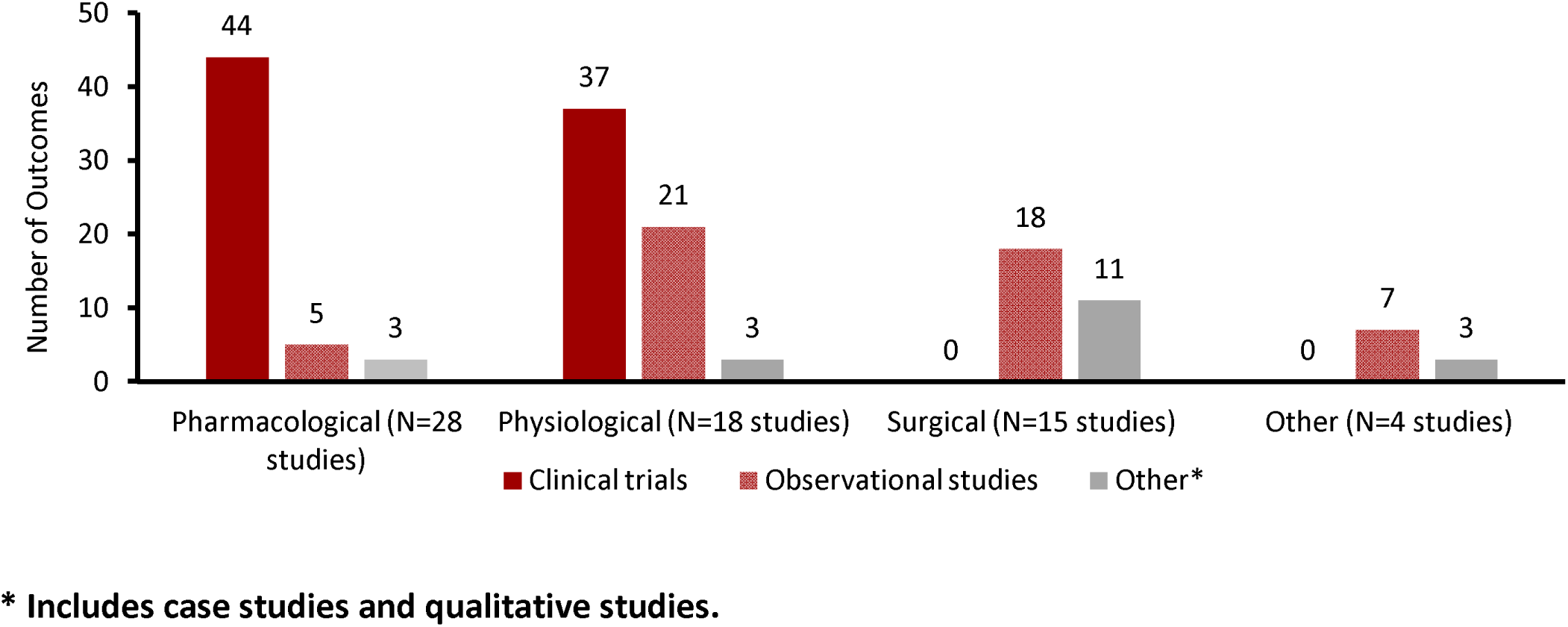
Total Number of Treatment Outcome Measures by Study Design and Intervention.

More than half of all outcome measures (50/89 outcomes) were used in at least 2 studies (**Figure 8**, **Figure 9**). Few measures were used in more than 10 studies; these included several muscle strength measures, respiratory capacity measures, shoulder motion measures, and the 6-minute walk test (6MWT) for efficacy and effectiveness assessment; and adverse events (AEs) and complications for safety evaluation (**Figure 8**). The most-used humanistic outcome was the Short Form 36 (SF-36), which was reported in 8 studies (**Figure 9**).

**Figure 7:**
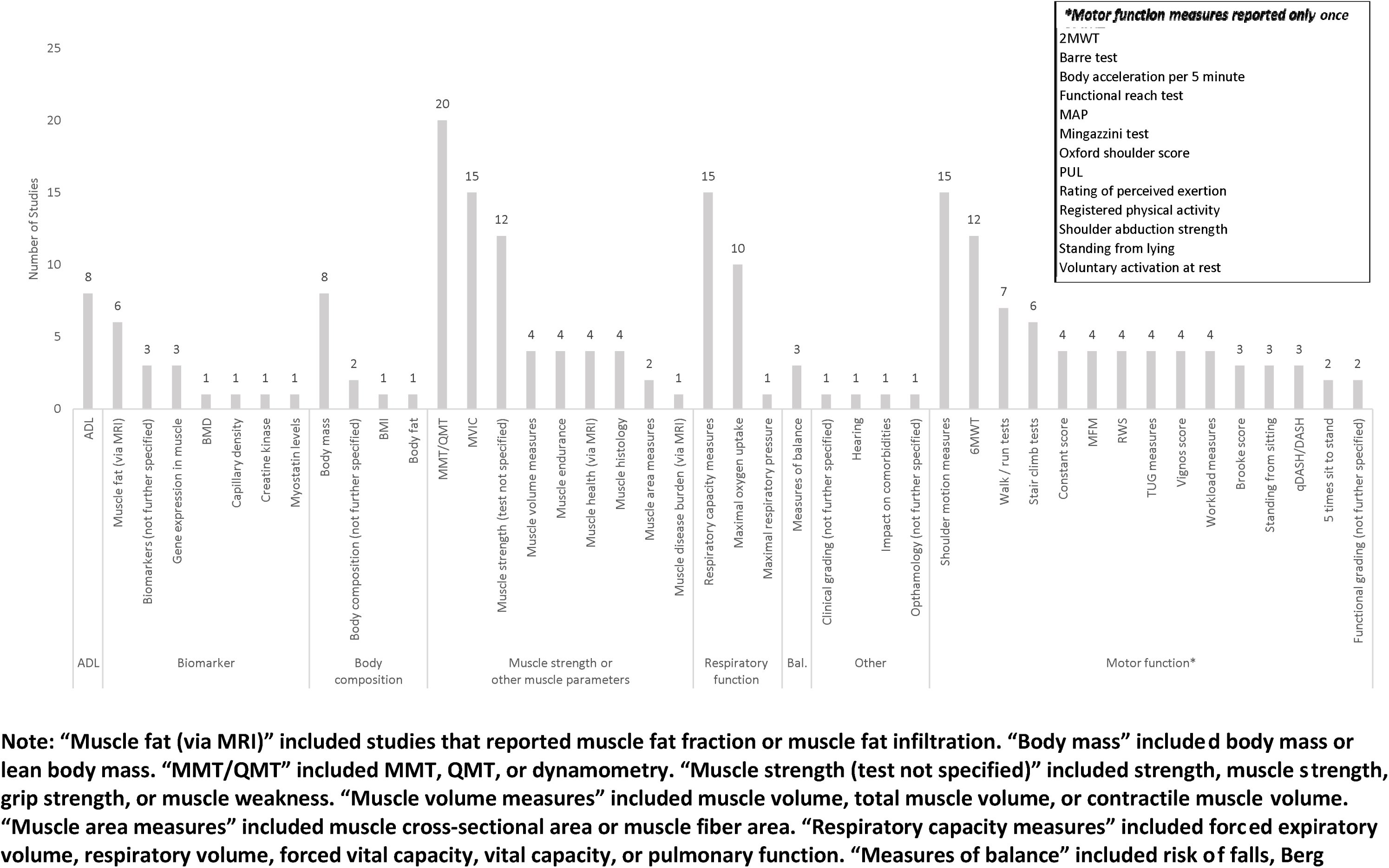
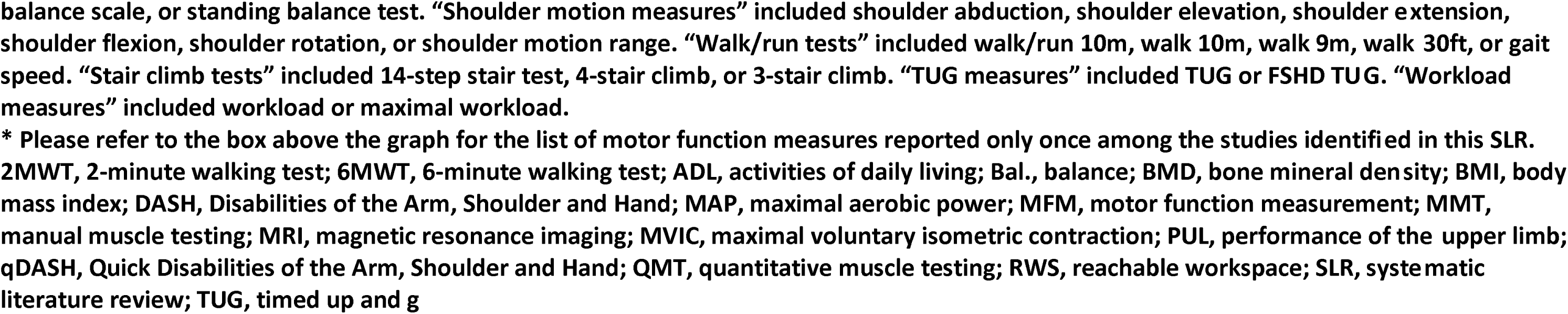
Number of Studies Reporting Individual Efficacy and Effectiveness Outcome Measures.

**Figure 8:**
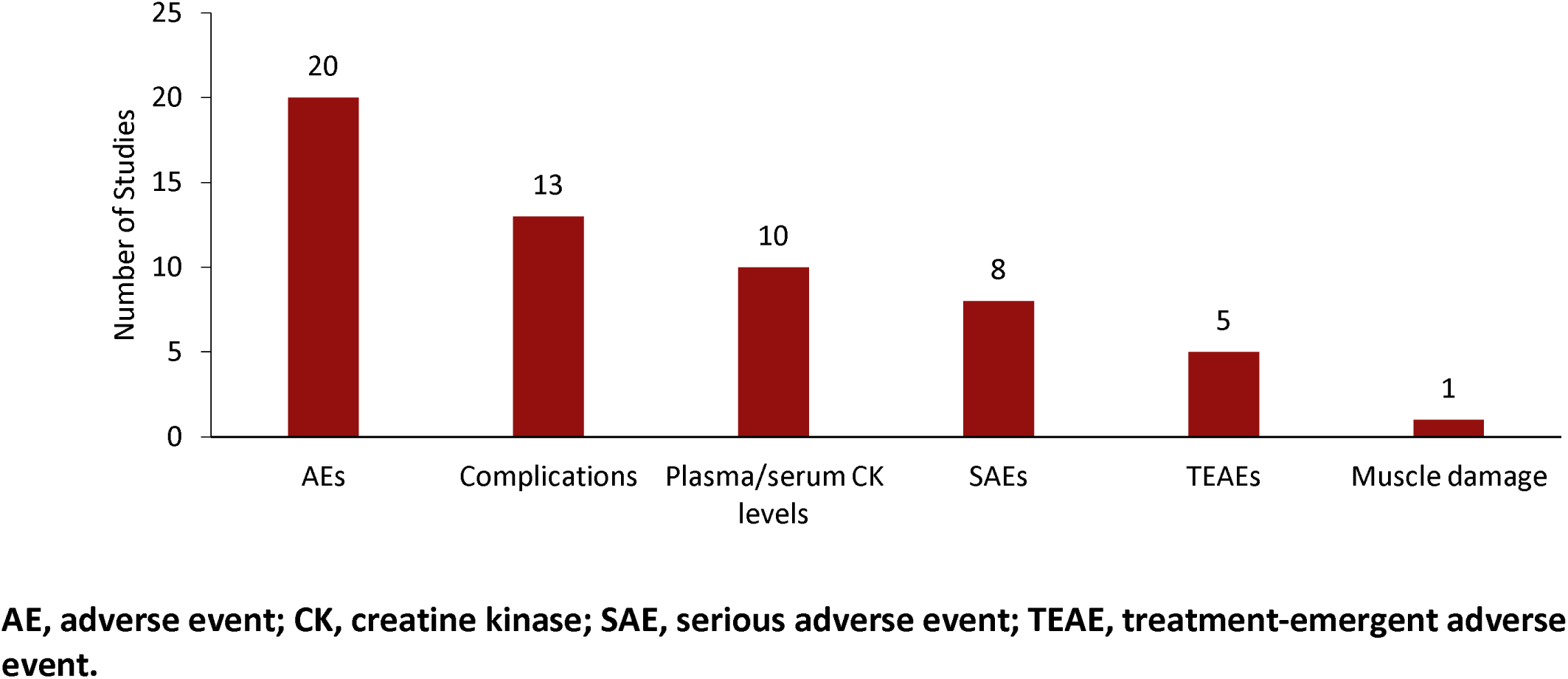
Number of Studies Reporting Individual Safety Outcome Measures.

**Figure 9:**
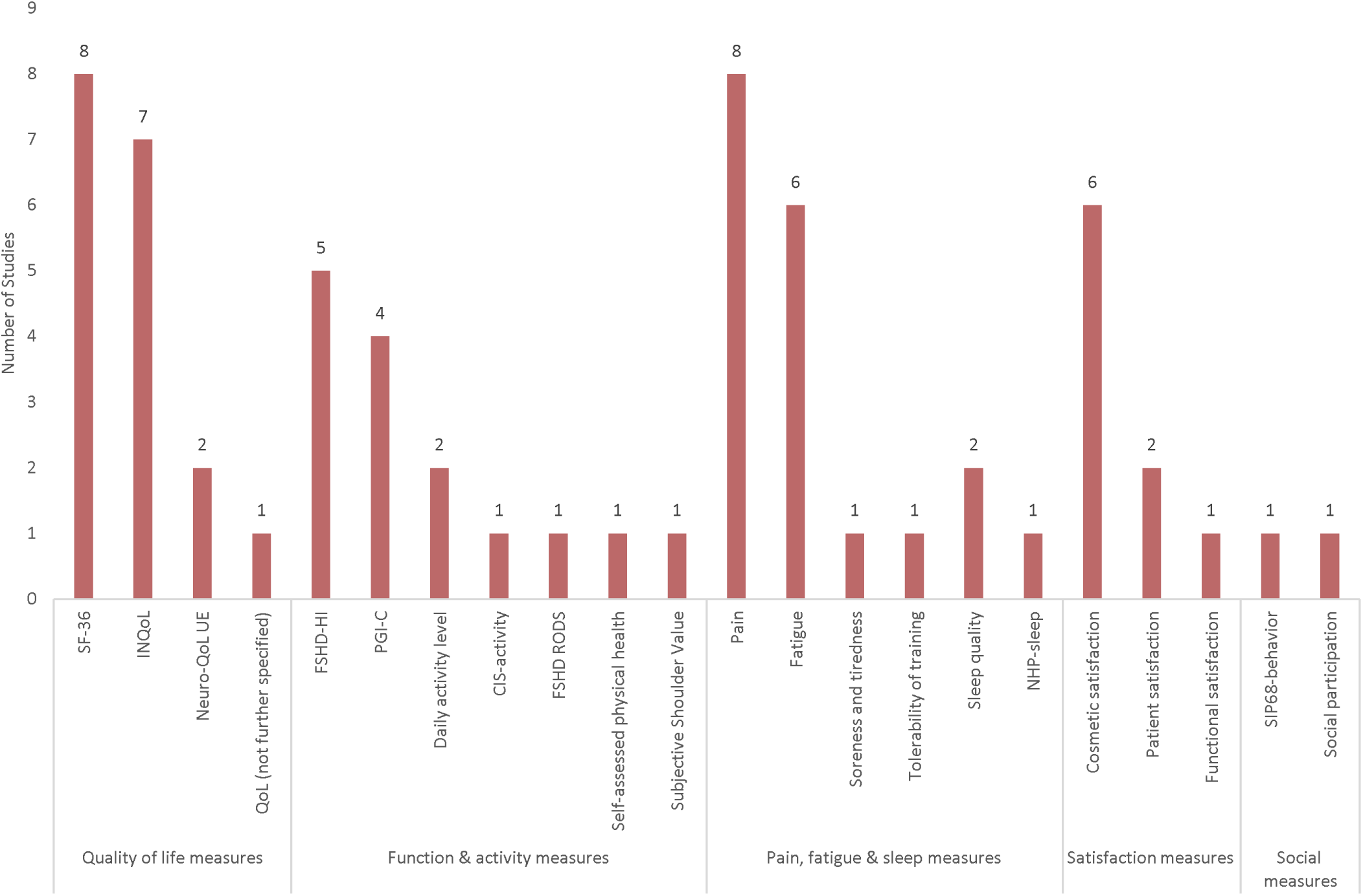
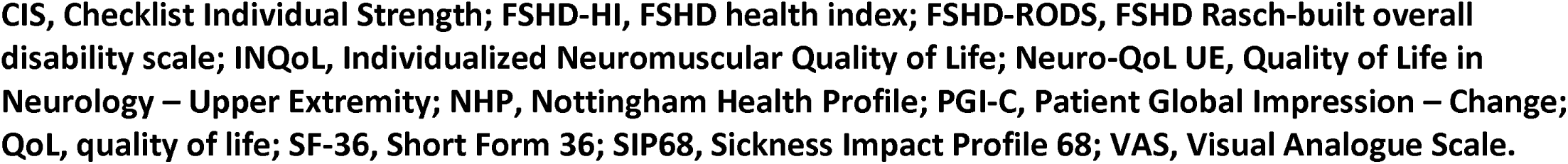
Number of Studies Reporting Individual Humanistic Outcome Measures.

**Figure 10:**
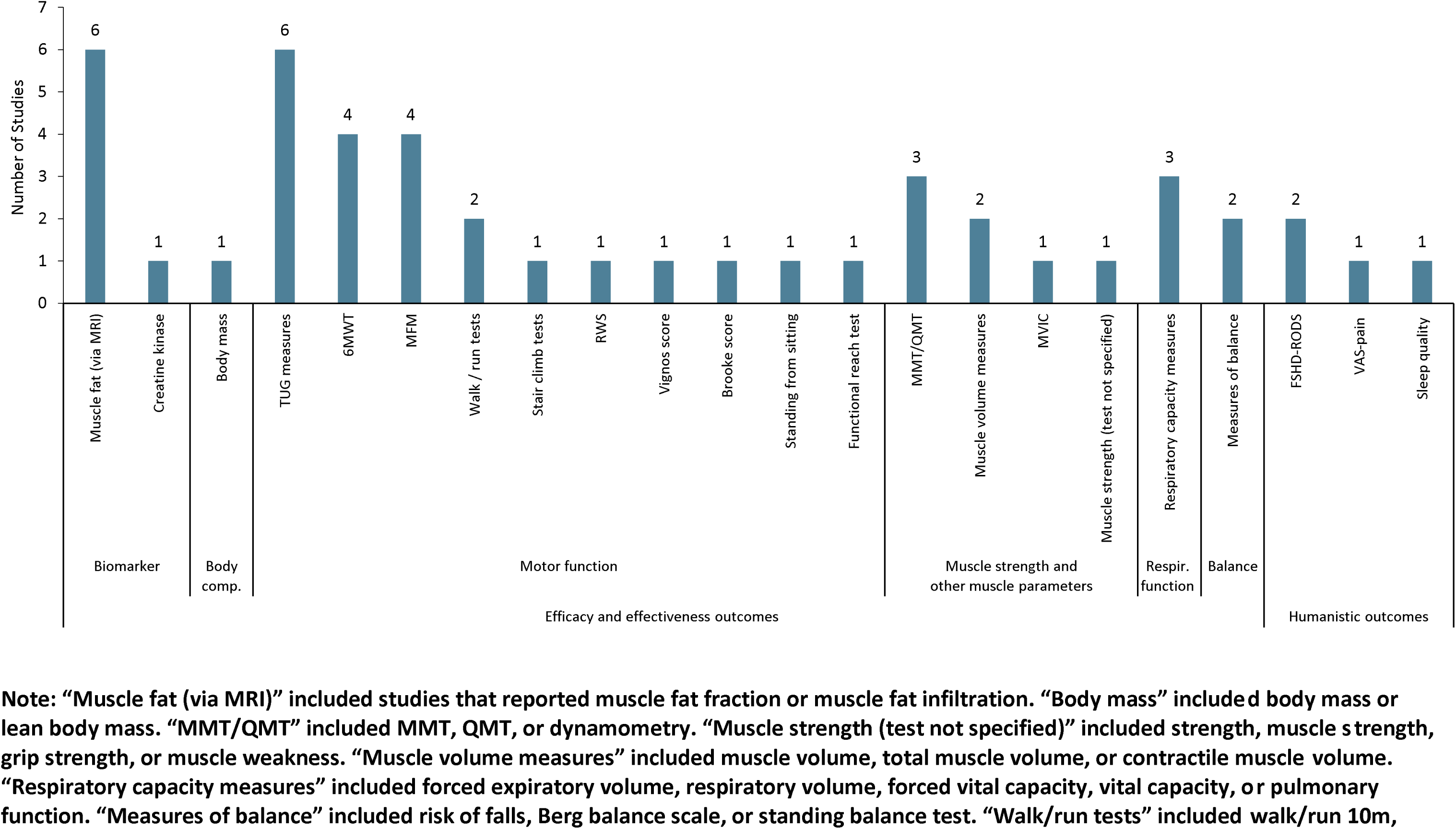
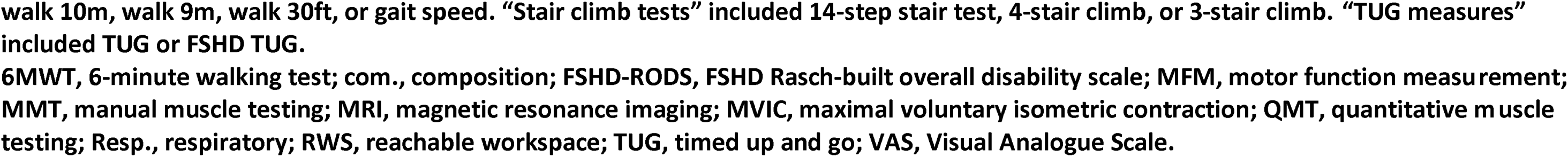
Number of Studies Validating Treatment Outcomes Identified in This Review.

Efficacy and effectiveness outcome measures were the most-commonly reported as compared to safety or humanistic outcomes (Error! Reference source not found.). A total of 58 measures were used to assess efficacy and effectiveness across studies, among which measures of motor function and muscle strength or other muscle parameters were the most frequently used measures. Frequently used measures of motor function include the 6MWT, walk/run tests and stair climb tests, while tests of shoulder motion, as well as reachable workspace (RWS) were used in multiple studies to evaluate upper limb function. Across studies, muscle strength was frequently measured via manual muscle testing (MMT)/quantitative manual testing (QMT) or maximal voluntary isometric contraction (MVIC). Respiratory function, mainly respiratory capacity, was also frequently measured. Among the six safety outcome measures identified across studies, proportion of patients experiencing any AEs or complication were the most reported measures (**Figure 8**). Frequency of AE occurrence was commonly assessed in clinical trials of pharmacological interventions, while complications were reported in studies using surgical procedures, which were mostly observational by study design.

Twenty-five measures assessed humanistic outcomes in the included studies, among which quality of life measured via different tools, patient satisfaction (including functional and cosmetic satisfaction), and pain- and fatigue-related scales were the most frequently used measures (**Figure 9**).

### Validation of Disease Outcome Measures in FSHD

A total of 55 studies reported on validation of disease outcome measures in patients with FSHD. Properties used to validate the outcome measures included correlation, Rasch analysis, factor analysis, reliability, responsiveness, and internal consistency. Details on which outcome measure was assessed in each study with which validation properties, along with brief definitions of each property, are presented in **Table Appendix 6**.

Among the 89 treatment outcomes identified in this SLR, only 22 of them were validated with at least 1 of the 6 properties, with correlation and reliability being the most reported ones and oftentimes both were used for the same outcome **Table Appendix 6**). Of the 22 validated treatment outcomes, most were efficacy and effectiveness outcomes, while only 3 were humanistic. Measures of timed up and go (TUG) and assessments of muscle fat were the most frequently assessed efficacy and effectiveness outcomes among validation studies, followed by 6MWT and motor function measurement (MFM), both measures of motor function.

### Humanistic Burden

An overview of the studies by quality of life (QoL) domain is provided in **Appendix Table 7**. Some studies did not report data but noted the use of an instrument in the Methods; thus, they are included in order to provide an overview of the domains noted to be measured. Among all studies, general QoL, as measured by a PRO instrument, was the most reported domain (n=21), followed by fatigue (n=14) then pain (n=12). A total of eight studies reported humanistic burden results qualitatively without the use of a PRO instrument. Detailed findings on humanistic burden are reported in **Supplementary Material C**.

### Economic Burden

The SLR identified 18 studies (12 full text articles and 6 conference abstracts) that reported at least one economic burden outcome and were included in the SLR (**Appendix Table 8, Appendix Table 9**). Of those that reported a geographic location, 9 studies were in the US, 6 in Europe (2 in Germany and 4 in the Netherlands), and 2 in the United Kingdom; 1 study did not report the geographic location. Two major study designs were used to assess economic burden: cohort (either retrospective or prospective) or survey, with most surveys being cross-sectional. Most studies did not detail the FSHD population included; of those which did (n=6), four were exclusively among those with Type 1 FSHD^135, 147–149^, and two were in a mixed FSHD population^127, 150^. Of the studies that reported the age of participants, all were among adults with mean/median age between 40 – 60 years of age; one study appeared to include pediatric patients according to the age range, but this was not specified^147^. Few studies reported the age at onset of FSHD, which was in the early 20s for all studies with the exception of Katz *et al.* which reported a median age of onset of 17 years^148^. None of the economic burden studies reported severity of disease and only three included information regarding D4Z4 pairs, repeats or contraction size.

Detailed outcomes of the topic “Economic Burden” are reported **Supplementary Material C**. Sample sizes across the studies ranged from 16 to 690. Outcomes reported were heterogenous. Per patient annual direct costs identified in the literature ranged from €9,270 (2010)^151^ to €21,256 (2018)^150^. Per patient annual societal costs ranged from €26,240 (2009)^152^ to €40,850 (2018).^153^ Konersman et al. reported that annual medical costs increased post-FSHD diagnosis; this increase post-diagnosis was much higher than for other diseases post-diagnosis, including connective tissues diseases.^154^ In a second conference abstract by Konersman et al., compared to matched controls, individuals with FSHD had higher costs and total number of prescriptions. Blokhuis et al. reported that total mean annual costs increased by mobility level ranging from €14,008 (2018) without any mobility aids to €63,765 (2018) among those who were unable to walk; further, the per-patient direct costs of illness were reported to be approximately five times higher than the mean per-capita health expenditure in the Netherlands.^150^

Healthcare resource use was reported in multiple studies. Up to 41% of patients^116^ reported use of ankle braces, leg braces, canes or walkers and up to 58.2%^155^ respondents noted that they used a wheelchair at some point during the day; full-time wheelchair was reported in up to 25.5%^127^ of FSHD individuals. Statland *et al.* found that among 343 individuals with FSHD who were device-free at baseline, after a mean follow-up of 6 years (range 1 – 9 years), 11% used an ankle brace, leg brace, cane or walker and 22% used a wheelchair at some point during the day.^116^ A second study by Statland et al. found that the use of wheelchairs, ankle braces, canes, walkers, breathing machines and hearing aids was higher for patients with large D4Z4 contractions (≤18 kb) versus medium or small contractions.^149^ Blokhius *et al.* reported that in the preceding 3 months, 58%, 53% and 26% of FSHD patients had a physical therapy, general practitioner or rehabilitation physician visit, respectively.^150^ Employment among individuals with FSHD and the impact of FSHD on employment was reported in 4 studies. Among 574 individuals with FSHD, Hilbert *et al.* found that 50% of all adult members reported reduced employment opportunities. Fowler *et al.* reported that among the 16 individuals with FSHD included in their study, 9 were employed; of the 7 unemployed, 6 had been employed in the past. In a study in the Netherlands, among employed individuals with FSHD, 12% reported absenteeism in the past 4 weeks, with a median absence duration of 152 days. One study reported that respondents taking 6 or more medications per day were significantly more likely to be unemployed (75.4%) than those taking 0 – 5 medications (41.9%) (p<0.001).^157^

## Discussion

FSHD is a highly heterogeneous disease with a wide phenotypic spectrum,^47^ thus requiring the use of a variety of outcome measures to understand disease progression and assess treatment benefit, in which the choice of the outcome measures may be dictated by the goal of the treatment and the type of intervention. This SLR identified a large number of measures used to assess outcomes following pharmacological, physiological, surgical, or other interventions in patients with FSHD, suggesting a lack of consensus in the field regarding how to best measure treatment benefit or disease burden in FSHD. This heterogeneity in the use of outcome measures was present in all study design groups, including clinical trials, observational studies, and case studies, although the heterogeneity was slightly higher among observational studies. Consistent with the heterogeneity observed among clinical trials included in the SLR, a variety of outcomes are being used in 7 currently ongoing clinical trials assessing interventions in FSHD (with all of them being pharmacological),^41, 48–53^ including 15 different efficacy outcomes, 8 safety, and 8 humanistic, reinforcing the need for achieving consensus on the use of outcome measures in FSHD. In line with this observation and with the findings of this SLR, there is not a defined set of efficacy outcome measures required or recommended by the Food and Drug Administration (FDA) for developing drug therapies for muscular dystrophies similar to FSHD (e.g. Duchenne muscular dystrophy and related dystrophinopathies including Becker muscular dystrophy).^54^

Efficacy and effectiveness outcomes were the most-commonly reported measures as compared to safety or humanistic outcomes, thus contributing more to the overall heterogeneity. While most (79%) efficacy and effectiveness outcomes appeared to be reported in fewer than 5 studies, half of the safety measures were used in at least 10 studies, suggesting that there is a higher consensus on using standard safety and tolerability assessments in patients with FSHD receiving treatment. Nevertheless, some safety outcomes reported are very broad in nature (e.g., frequency of any AE or complication) and may explain the lower observed heterogeneity in this outcome category. In terms of humanistic outcomes, the findings suggest that although there seems to be general consensus on what to measure (e.g., QoL, patient satisfaction, pain, fatigue), there are a variety of different tools or scales being used across studies. Recent guidance from the FDA indicate that, in muscular dystrophies like Duchenne muscular dystrophy or Becker muscular dystrophy, disease-specific PROs are better suited for clinical trials to measure disease burden and relevant therapeutic gains over time, with an improved ability to show whether a therapy has patient-relevance.^55^ Across the 25 humanistic outcomes identified in this SLR, only 2 of them were FSHD-specific PROs, revealing an opportunity for improvement in selecting more relevant PROs in future studies.

Outcome measures of motor function and muscle strength were the most frequently used to assess efficacy and effectiveness among the clinical trials and observational studies identified. These findings are in line with a recent SLR by Aguirre et al. summarizing the clinical benefits of emerging treatments in FSHD focusing on evidence from 11 clinical trials,^56^ all of which were captured also in our SLR. Measures of muscle strength and upper and lower limb function were frequent outcomes used to assess efficacy outcomes of pharmacological therapies among the clinical trials identified by Aguirre et al., consistent with the trends observed in our SLR which is broader in scope.

Examples of frequently used measures identified in this SLR include the 6MWT and walk/run tests for motor function, shoulder motion measures for upper limb function, and MMT/QMT and MVIC for muscle strength. RWS, one of the few measures that evaluates upper limb function, was used in only four clinical trials, likely because it is a relatively new measure in FSHD (2015)^57^ and most studies identified in this SLR were conducted before its development. However, given that RWS provides a quantitative measure of upper limb function,^57^ one of the most affected parts of the body in patients with FSHD,^2^ the use of RWS may observe an increase as new studies arise. Indeed, two recently initiated clinical trials not captured in this SLR include RWS as outcome.^51, 52^

Given that motor function and muscle strength and endurance outcomes provide quantitative measures of key aspects that determine disease progression, they may represent an opportunity to build consensus around both what and how to measure treatment benefit in FSHD. Like in other muscular dystrophies, combining several of these outcome measures may allow to assess change of function over time across the many areas of the body affected by FSHD. In FSHD, previous lack of consensus on the importance of some outcome domains over others has led to the development of the FSHD Composite Outcome Measure (FSHD-COM) for use in future clinical trials.^58^ In this context, selection of specific measures to be included as part of a composite outcome measure but also as standalone outcomes should consider the type of intervention and the particularities of the study population. For instance, as FSHD typically manifests first in the shoulder and upper arms (in addition to the face),^2^ the use of upper limb function measures like RWS may allow for detection of disease progression and evaluation of therapy effects at early stages of the disease. Importantly, selection of specific measures should also consider the clinical heterogeneity of FSHD. When selecting measures of motor function, combining outcomes like RWS, which measures upper limb function, with the TUG, which measures lower limb function as well as mobility (e.g., sit-to-stand, walking, turning balance), should be considered to capture the full picture of disease progression.

The humanistic burden of patients with FSHD was reported in 38 full text articles and 15 conference abstracts. While the majority of the studies were observational in nature with a single point in time of measurement, some clinical trials did include an assessment of one or more PROs. Multiple instruments across the domains of general QoL, disease-specific QoL, depression, anxiety, pain, sleep, activities of daily living, disability/function, fatigue, neurological/psychological and symptom/illness perception were identified.

Further, some studies reported a qualitative assessment of QoL without the use of an instrument. The most reported domains included general QoL, followed by fatigue then pain. QoL was assessed in both adult and pediatric FSHD patients, with some instruments developed exclusively for use among children and adolescents (for example, Kidscreen for general QoL and the FSHD-HI Peds for disease-specific QoL). Based on the available evidence, it is evident that FSHD impacts QoL, with pain and/or fatigue impacting QoL.

Limited economic burden evidence. A total of 18 studies (12 full text articles and 6 conference abstracts) reported on the economic burden among FSHD patients. Economic outcomes reported across the studies were heterogenous including medical and societal costs, number of prescriptions and healthcare resource use, such as wheelchair and hearing aid use and specialist visits. While al range of per patient annual direct and societal costs were identified in the literature, which was reported to be approximately five times higher than the mean per-capita health expenditure in the Netherlands.^150^ Nearly half of FSHD patients reported use of either ankle braces, leg braces, canes or walkers and over half noting that they used a wheelchair at some point during the day. In addition, a large proportion of patients with FSHD have reduced employment opportunities. While limited, the available evidence suggests that FSHD is associated with a substantial economic burden for patients, their caregivers and society.

Finally, outcome measures used in FSHD clinical trials should be in general reliable and valid to allow implementation across multiple sites in a clinical trial.^59^ As the field anticipates more trials during the upcoming years, reliable and practical outcome measures will be needed.^60^ This SLR found that only around 25% of the treatment outcomes identified had been previously validated, suggesting that more effort is needed not only on establishing consensus on which outcome measures should be used but also in assessing their validity and reliability.

## Strengths and Limitations

The major strength of this SLR relies on the broad and comprehensive search conducted in three major medical literature databases (PubMed, Embase, and Cochrane CENTRAL). No limits other than English language were applied, which allowed the strategy to capture all available published evidence indexed in the three databases from inception to the search date. In addition, multiple conference proceedings relevant to the field were searched to supplement the databases searches.

Furthermore, the inclusion criteria of this SLR were broad, without limits to the population (FSHD type, adult/pediatric population), intervention (any type), comparators (any), or study design (clinical trials and observational studies, including case studies with one single patient, as well as other published SLRs on the topic). Meanwhile, the SLR by Aguirre et al.^56^ only focused on treatment outcomes of pharmacological interventions from clinical trials. As such, this SLR captured all 11 studies identified in the SLR by Aguirre et al.^56^ and 54 additional studies.

There are some limitations applicable to all SLRs that should be acknowledged. Articles and conference abstracts that were published after the search date were not captured. Further, publications that were published close to the search date but were not yet indexed in the databases at the time of the search may have not been captured by the search. The SLR is also limited by the use of published data. There is a risk of publication bias as some studies fail to be published while others are published only in abstract form, which present limited information. However, regardless of the type of publication, all information was captured and reported on in this SLR.

## Conclusion

Despite being the second most prevalent form of muscular dystrophy and the lack of approved therapies, consensus on outcome measures assessing disease progression and treatment outcomes in FSHD has not yet been reached in the field, and validation of currently used measured is limited. This SLR identified a high number of measures used to assess treatment outcomes and patient burden following interventions in patients with FSHD, but only one fourth of them were previously validated. Significant heterogeneity in included outcome assessments was also evident in clinical trials. Therefore, further effort is needed to build consensus around the treatment outcome measures most relevant to FSHD patients, such that clinical trials for new interventions can be designed with the evaluation of patient-relevant benefits in mind and emerging innovations can be consistently evaluated.

## Data Availability

All data produced in the present study are available upon reasonable request to the authors

## Abbreviations

10MWT: 10-minute walk test
2MWT: 2-minute walk test
6MWT: 6-minute walk test
ADL: activities of daily living
AE: adverse event
BMD: bone mineral density
BMI: body mass index
CIS: Checklist Individual Strength
CK: creatine kinase
CSS: clinical severity score
D: dimension
DASH: Disabilities of the Arm, Shoulder and Hand
DEXA: dual energy X-ray absorptiometry
DUX4: Double Homeobox 4
EIM: electrical impedance myography
eTUG: electronic timed up and go
FCS: FSHD clinical score
FDA: Food and Drug Administration
FFS: Facial Function Scale
FSHD: facioscapulohumeral muscular dystrophy
FSHD IRC: FSHD Society International Research Congress
FSHD-COM: FSHD composite outcome measure
FSHD-HI: FSHD health index
FSHD-RODS: FSHD Rasch-built overall disability scale
ft: feet
FVC: forced vital capacity
IFF: infiltrated fat fraction
INQoL: Individualized Neuromuscular Quality of Life
IOPI: Iowa Oral Performance Instrument
IPT: isokinetic peak torque
JBI: Joanna Briggs Institute
LE: lower extremity
LMV: lower muscle volume
MAP: maximal aerobic power
MDA: Muscular Dystrophy Association
MFF: muscle fat fraction
MFI: muscle fat infiltration
MFM: motor function measurement
MMSE: Mini-Mental State Examination
MMT: manual muscle testing
MoCA: Montreal Cognitive Assessment
MPI: mean pixel intensity
MRC: Medical Research Council
MRI: magnetic resonance imaging
MSG: Muscle Study Group
MUS: muscle ultrasound
MVC: maximum voluntary contraction
MVIC: maximal voluntary isometric contraction
Neuro-QoL UE: Quality of Life in Neurology – Upper Extremity
NHP: Nottingham Health Profile
NR: not reported
NS: not specified
ODI: oxygen desaturation index
oTUG: optimized timed up and go
PGI-C: Patient Global Impression – Change
PICOS: Population, Intervention, Comparator, Outcome, Study design
PRISMA: Preferred Reporting Items for Systematic Literature Reviews and Meta-Analyses
PRO: patient-reported outcome
PROMIS-PF: Patient-Reported Outcomes Measurement Information System Physical Function
PUL: performance of the upper limb
qDASH: Quick Disabilities of the Arm, Shoulder and Hand
QMT: quantitative muscle testing
QoL: quality of life
RCT: randomized controlled trial
RSA: relative surface area
RWS: reachable work space
SAE: serious adverse event
SES: Self-efficacy Scale
SF-36: Short Form 36
SIP68: Sickness Impact Profile 68
SLR: systematic literature review
SpO2: oxygen saturation
TEAE: treatment emergent adverse event
TUG: timed up and go
UK: United Kingdom
US: United States
VAS: Visual Analogue Scale
VO2: oxygen consumption
Wmax: maximal workload

## Supplementary Material A

**Table Appendix 1:**
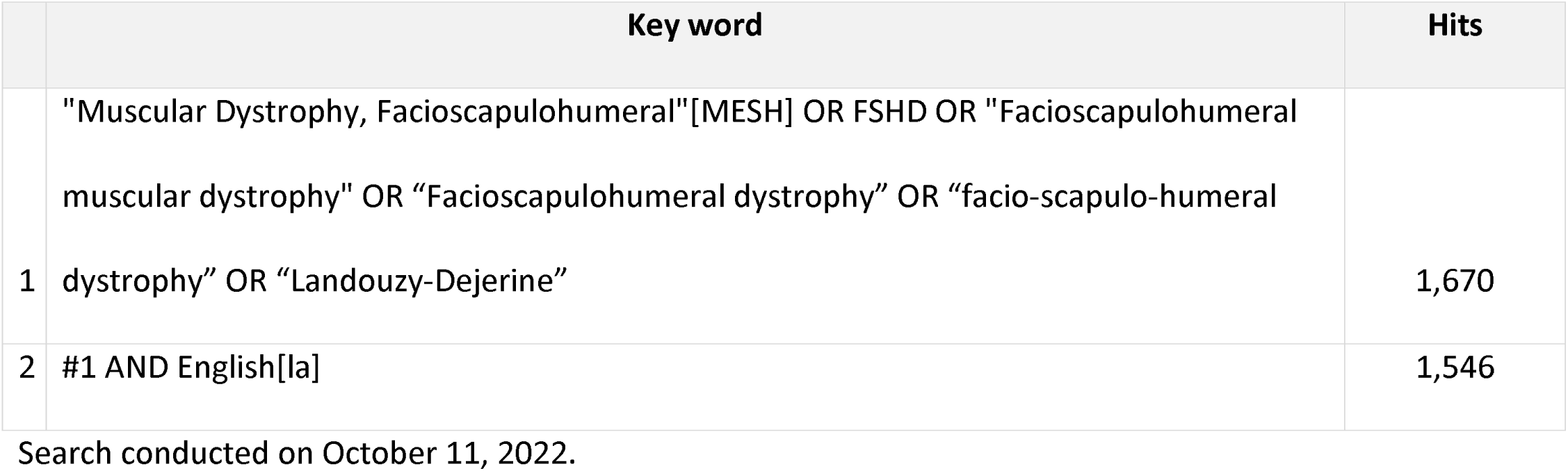
PubMed / MEDLINE Search Strategy.

**Table Appendix 2:**
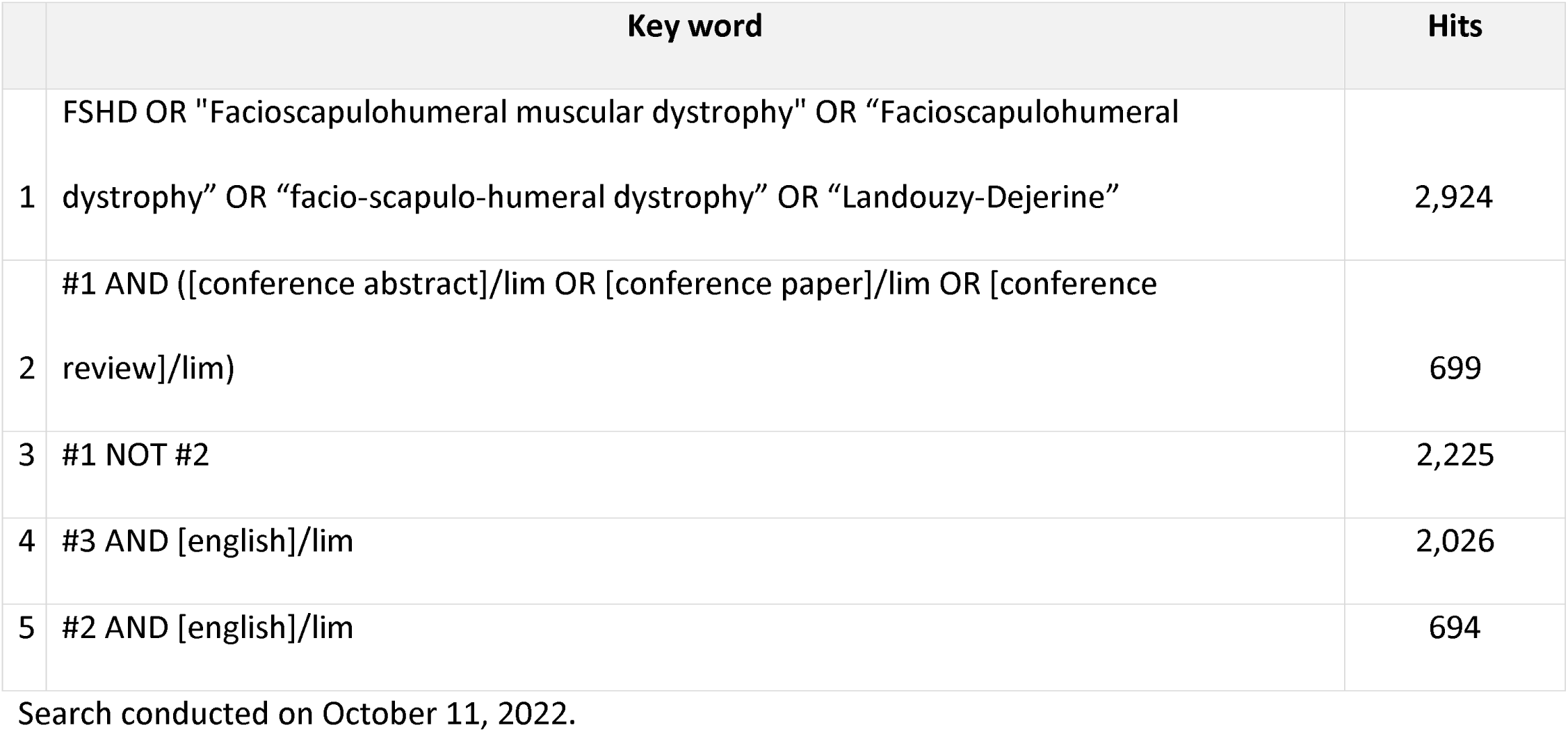
Embase Search Strategy.

**Table Appendix 3:**
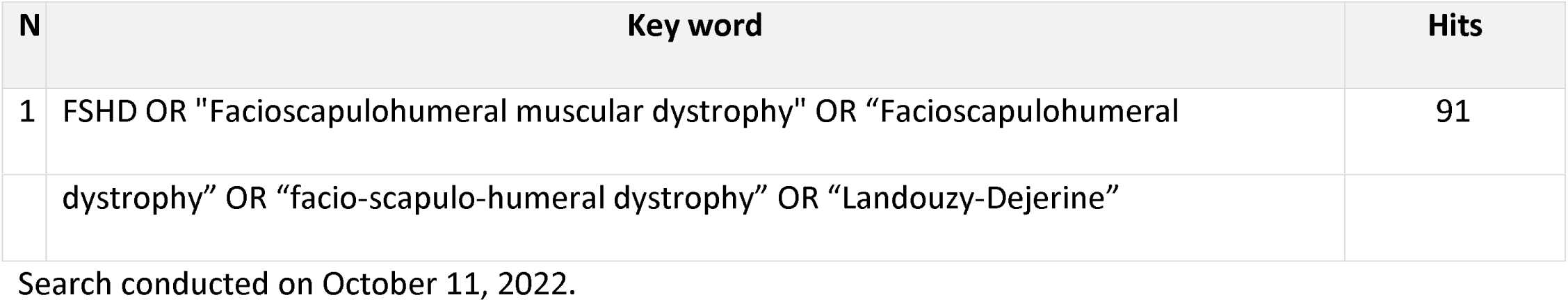
Cochrane CENTRAL Search Strategy.

Search conducted on October 11, 2022. Table Appendix 3: Cochrane CENTRAL Search Strategy
| N | Key word | Hits |
| --- | --- | --- |
| 1 | FSHD OR "Facioscapulohumeral muscular dystrophy" OR "Facioscapulohumeral | 91 |
|  | dystrophy” OR “facio-scapulo-humeral dystrophy” OR “Landouzy-Dejerine” |  |

Search conducted on October 11, 2022.

**Table Appendix 4:** PICOS Criteria.

| **Criteria** | **Inclusion** | **Exclusion** |
| --- | --- | --- |
| **Population** | Adults, children/adolescents with FSHD, type 1 and 2 | Patients with conditions/muscular dystrophies other than FSHD<br>Mixed populations not presenting stratified FSHD data |
| **Intervention** | Any intervention | No intervention |
| **Comparator** | No criteria | No criteria |
| **Outcomes** | Any type of measures that assess treatment outcomes<br>Validation of disease outcome measures (any method/property used) | Outcomes other than those described in the inclusion criteria |
| **Study Design** | Clinical trials of investigational therapies or interventions (i.e., physical therapy)<br>Observational studies/real-world evidence studies (e.g., prospective cohort and cross-sectional studies, retrospective analyses of real-world databases, disease registries, case series) | Pre-clinical/non-human studies<br>Narrative reviews, commentaries, letters, notes, editorials |

| Criteria | Inclusion | Exclusion |
| --- | --- | --- |
|  | • Systematic literature reviews<br>• Case reports/studies<br>• Trial registers |  |
| Limits | • Language: English<br>• Timeframe:<br>○ Full-text articles and indexed conference abstracts: no limit<br>○ Hand-searched conference abstracts: since January 1, 2019 | • Conference abstracts published before January 1, 2019 for hand searched conferences only |

**FSHD, facioscapulohumeral muscular dystrophy; PICOS, Population, Intervention, Comparator, Outcome, Study design**

**Table Appendix 5:**
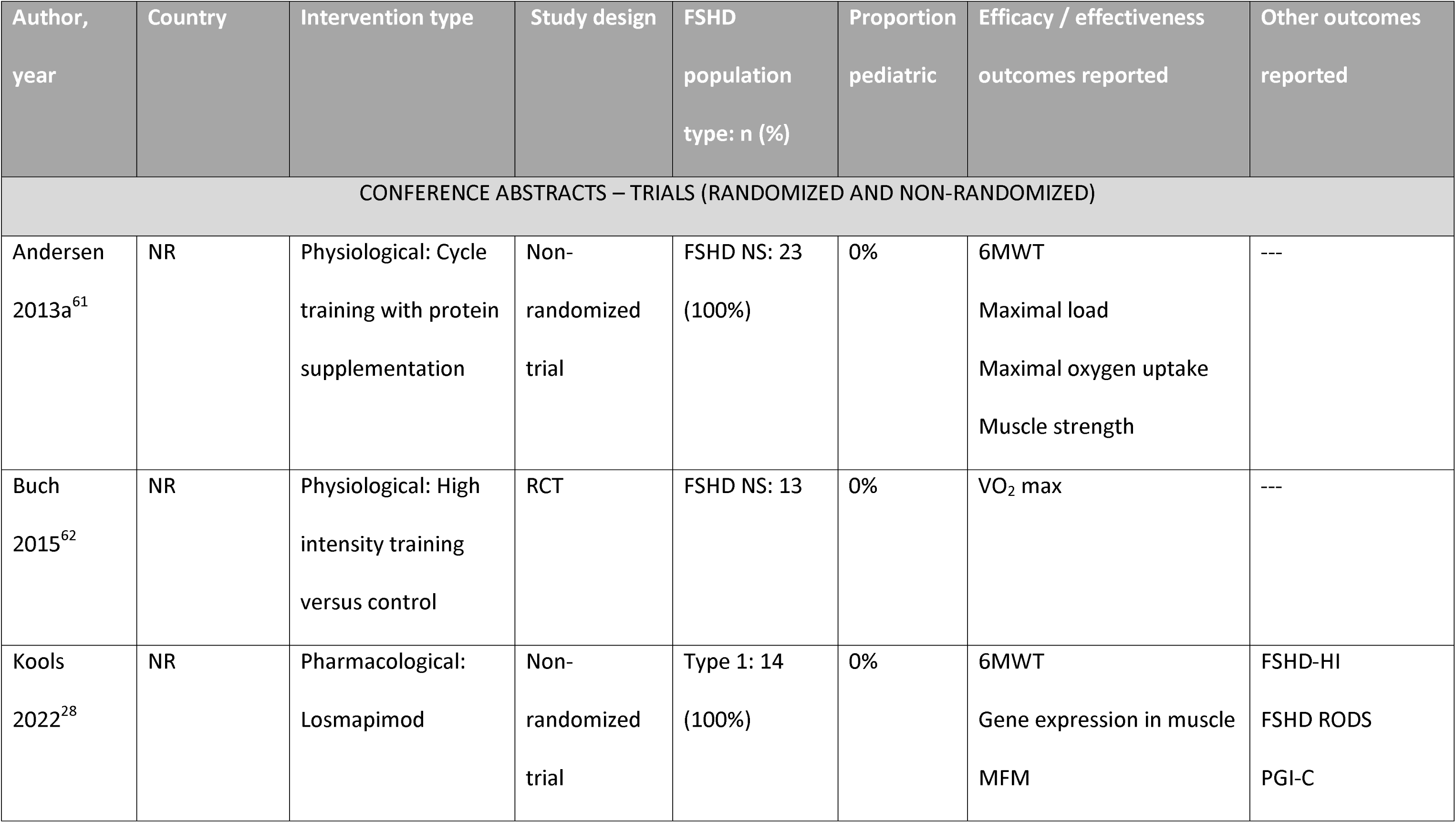
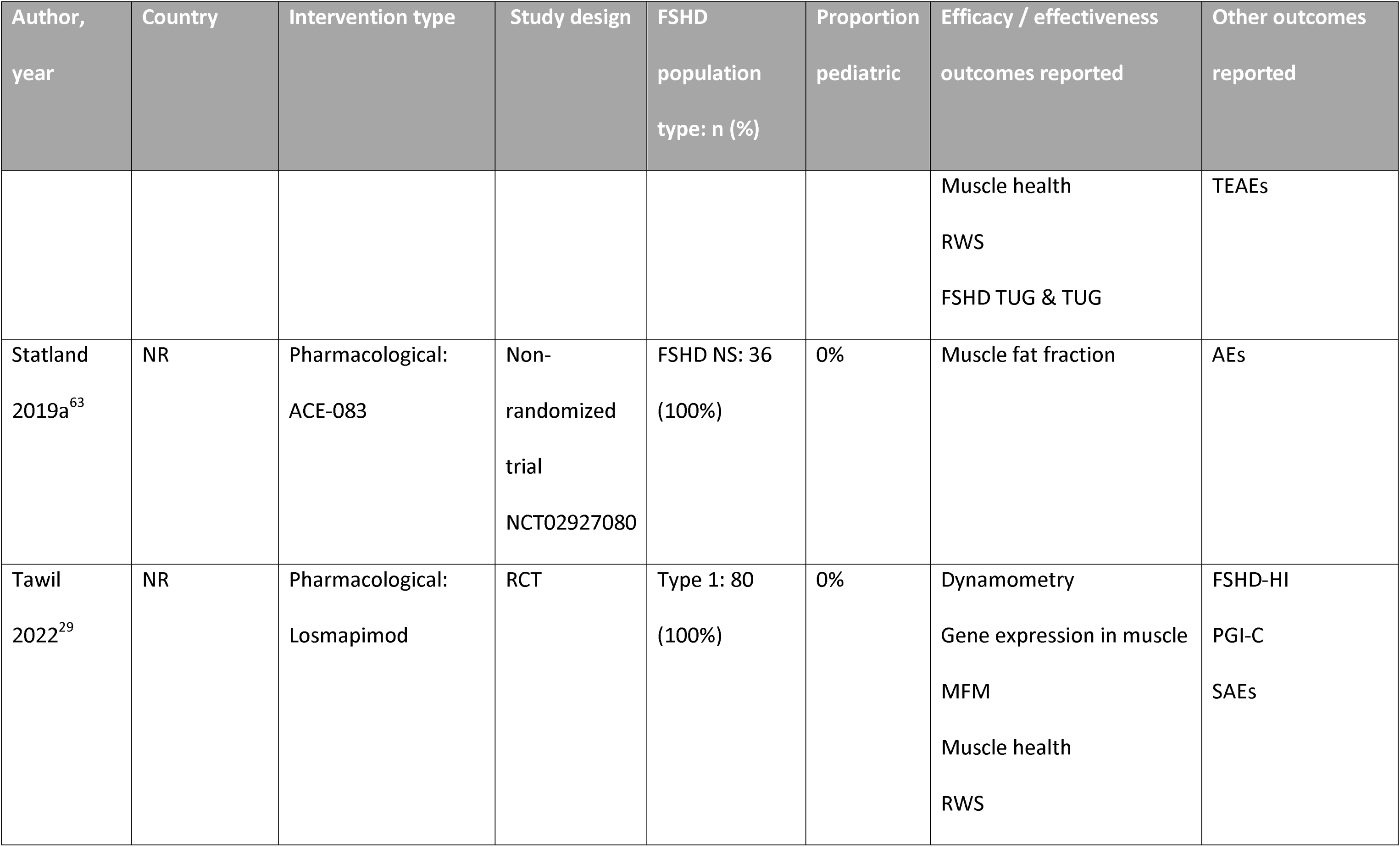
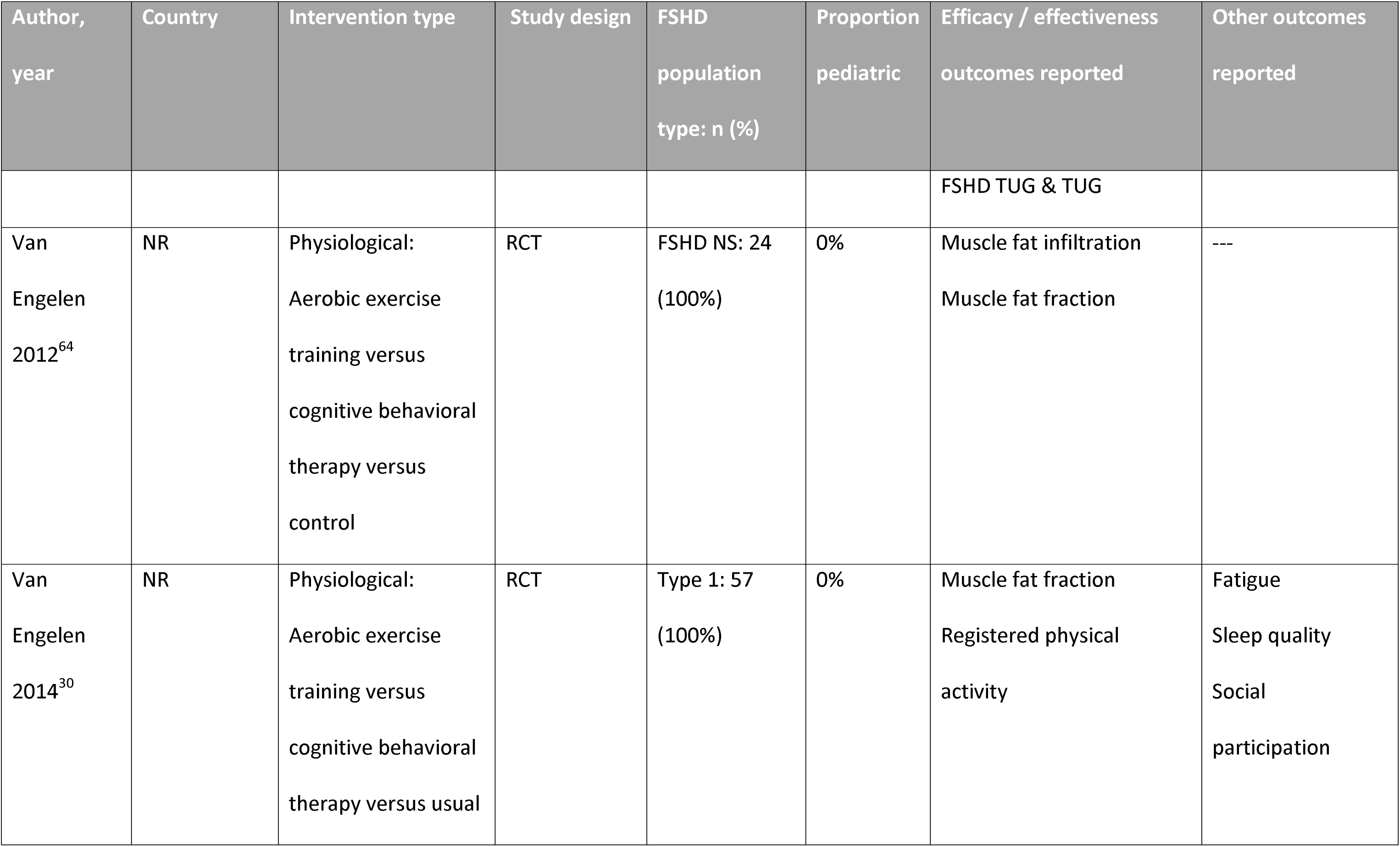
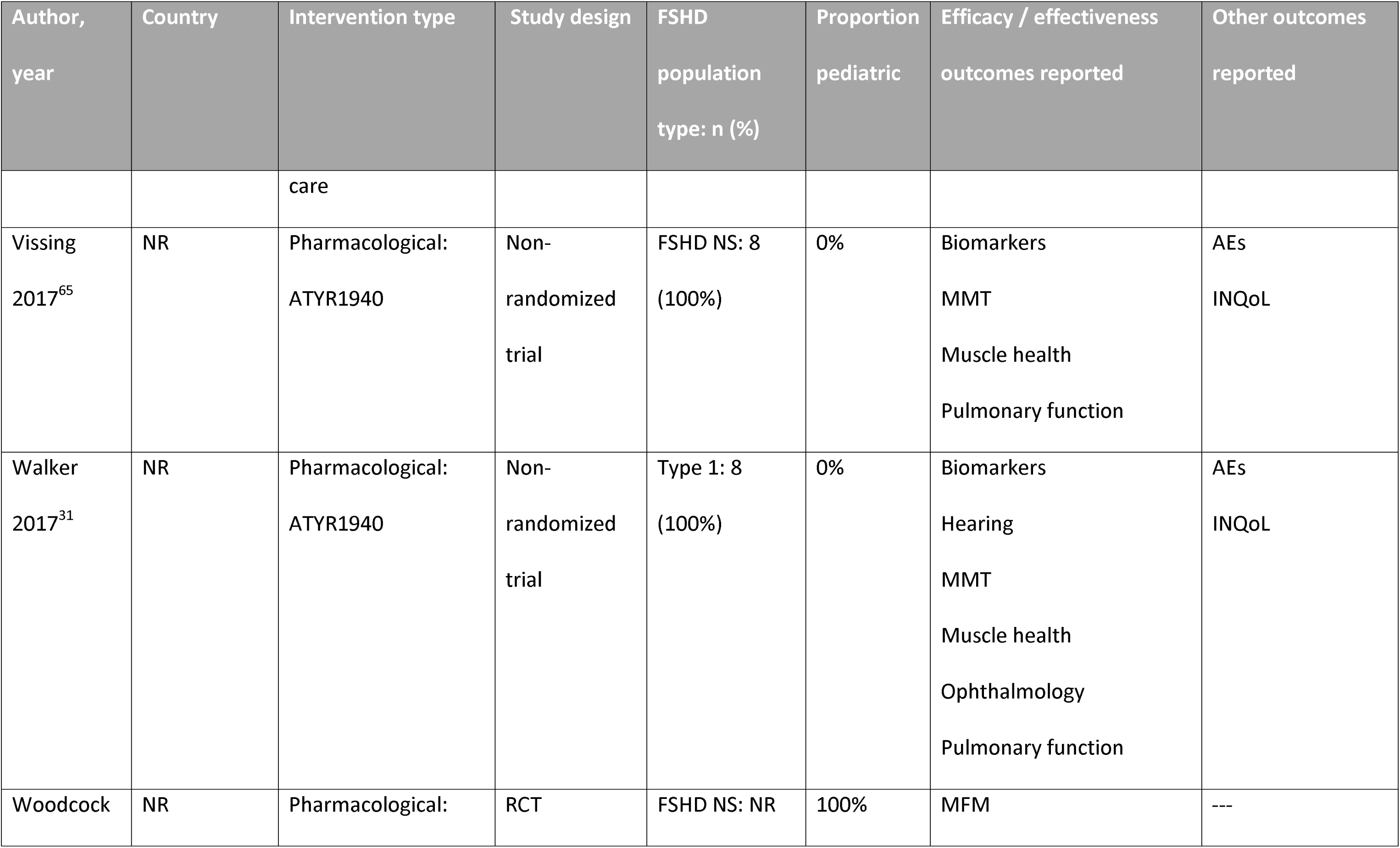
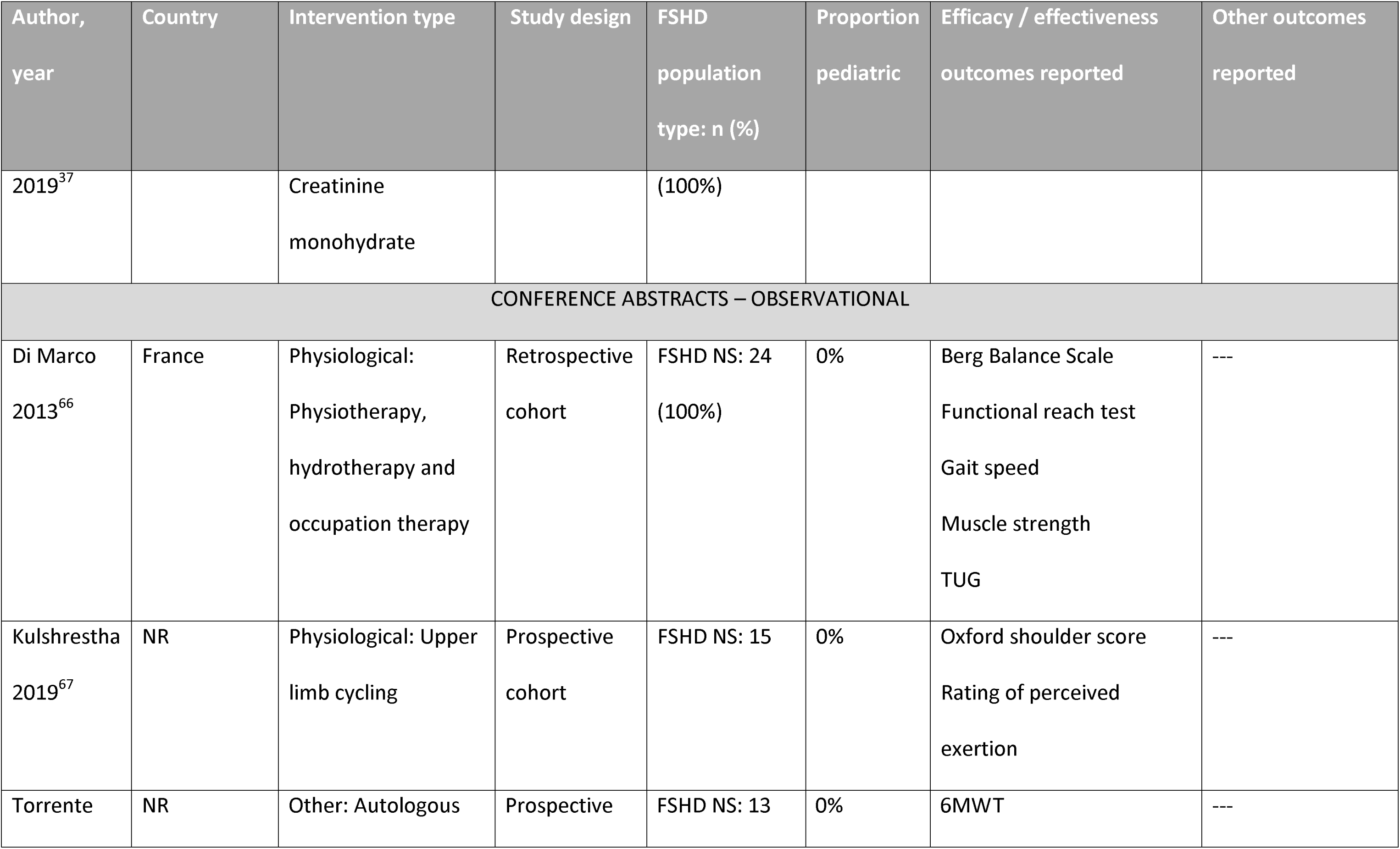
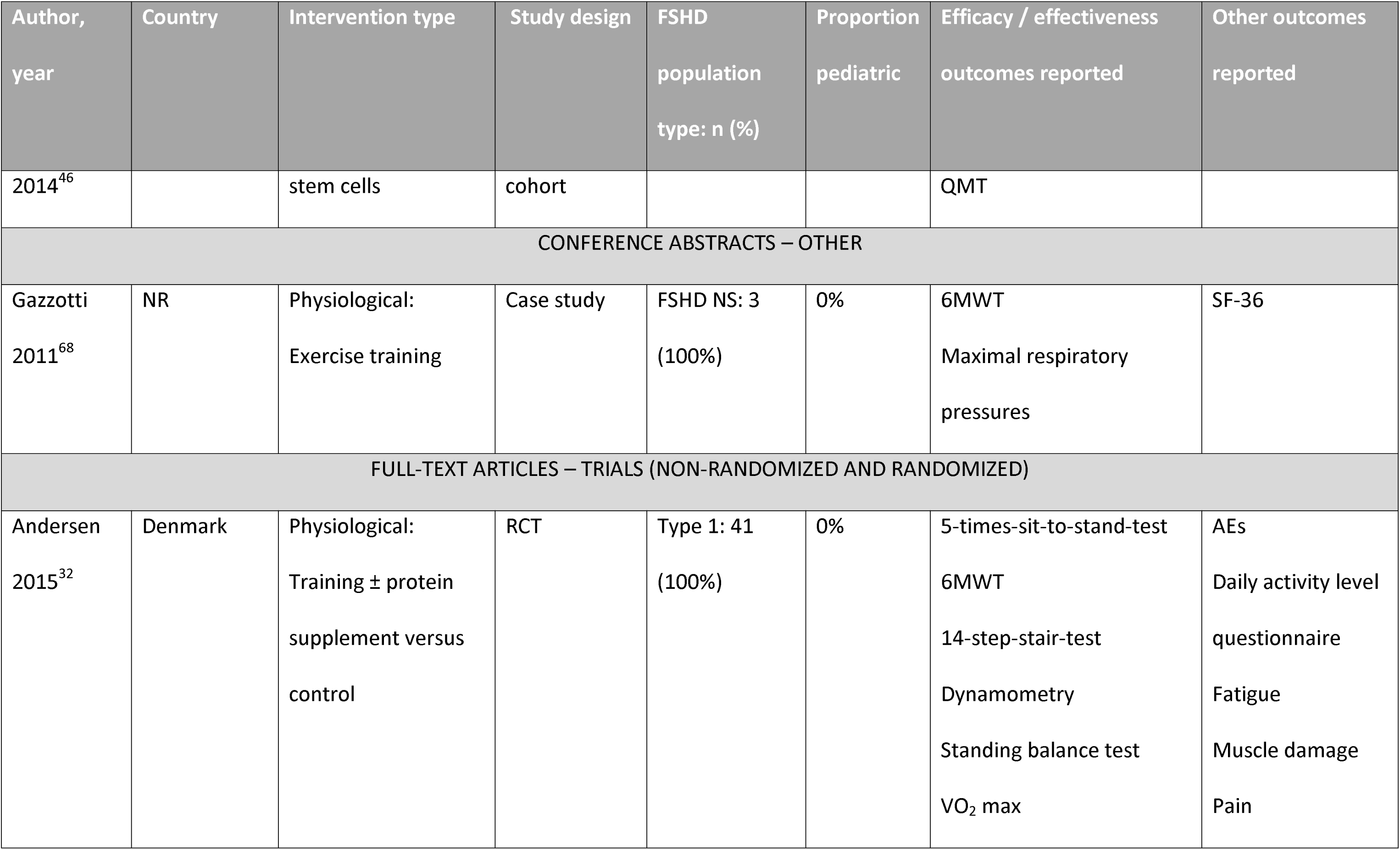
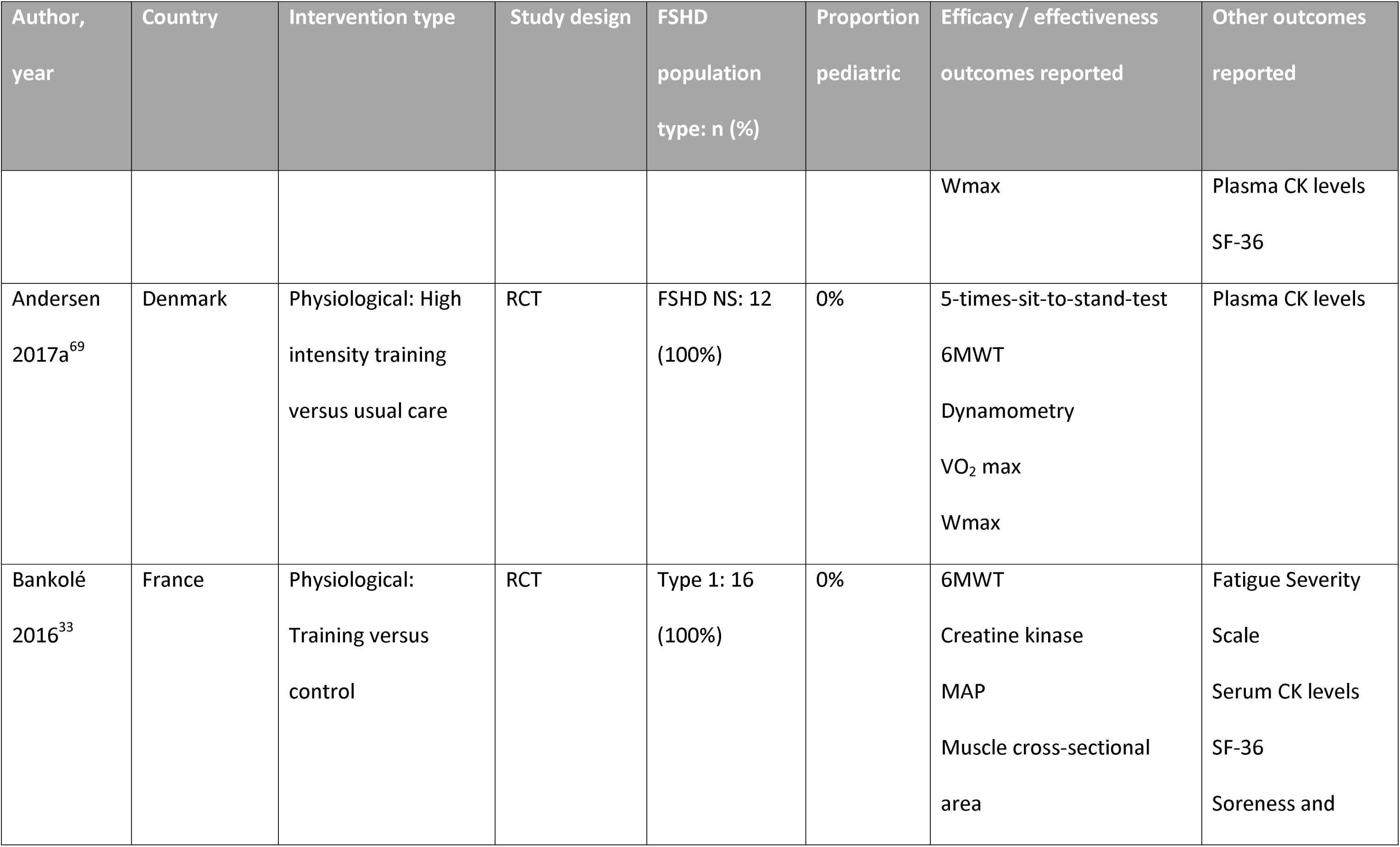
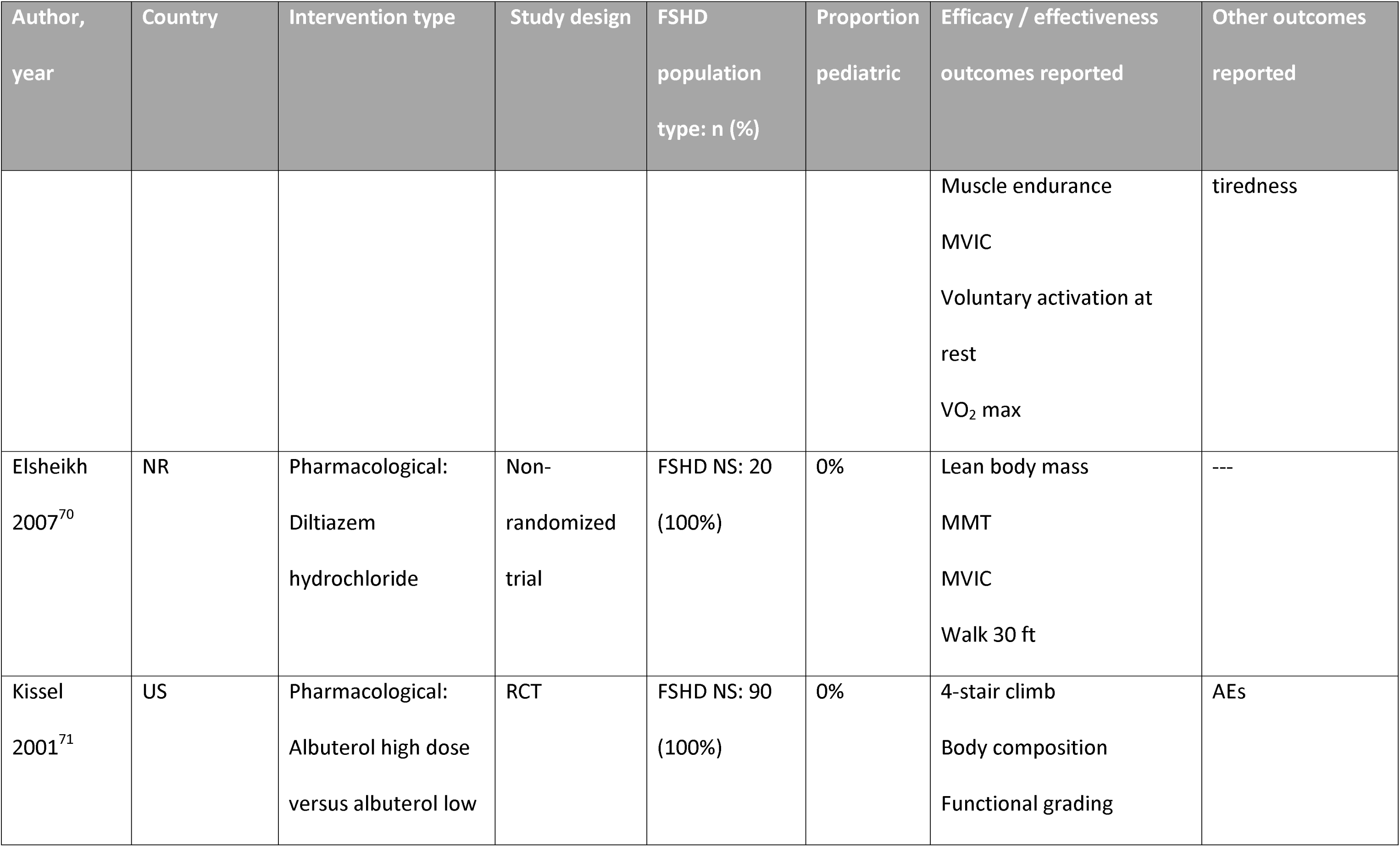
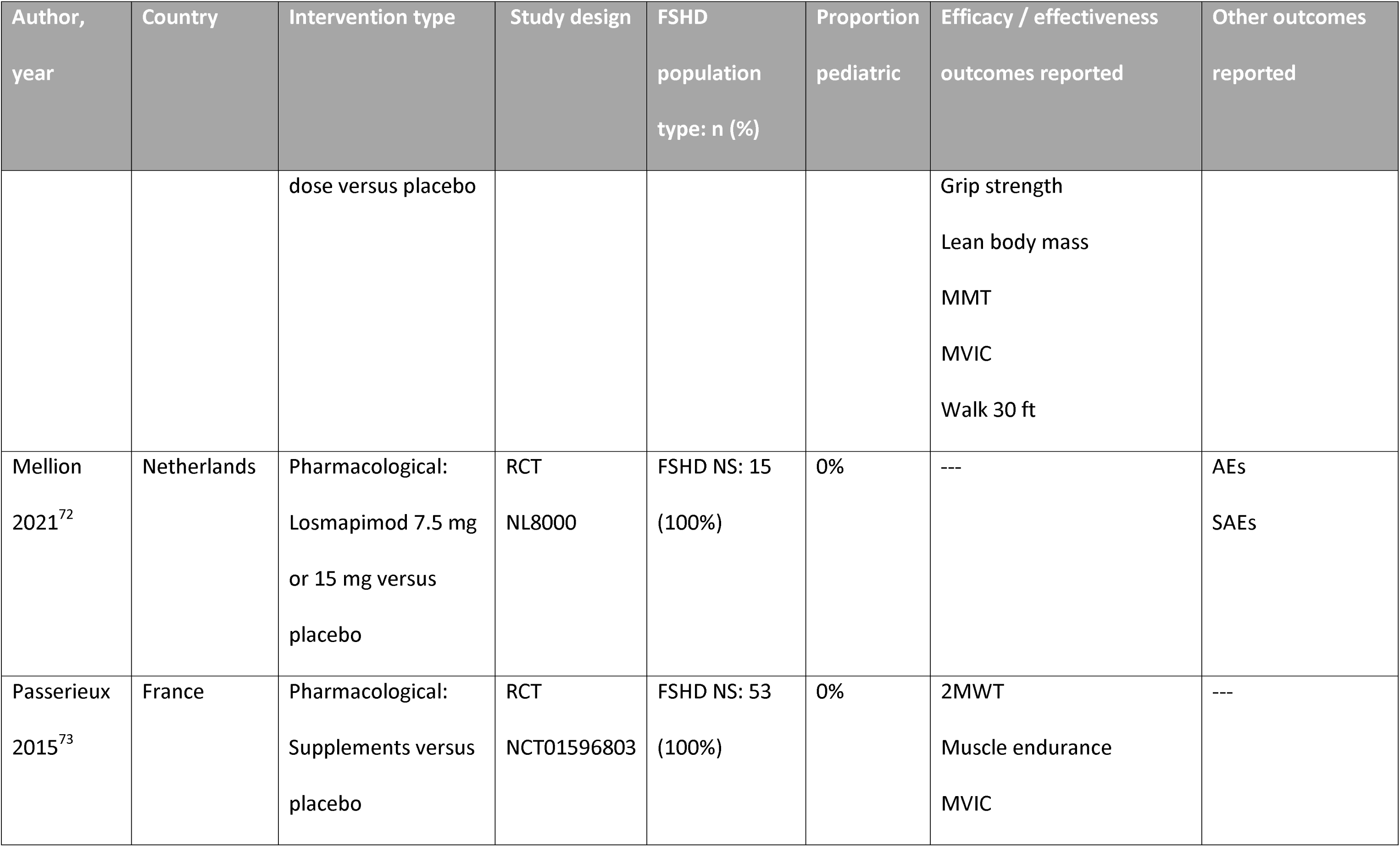
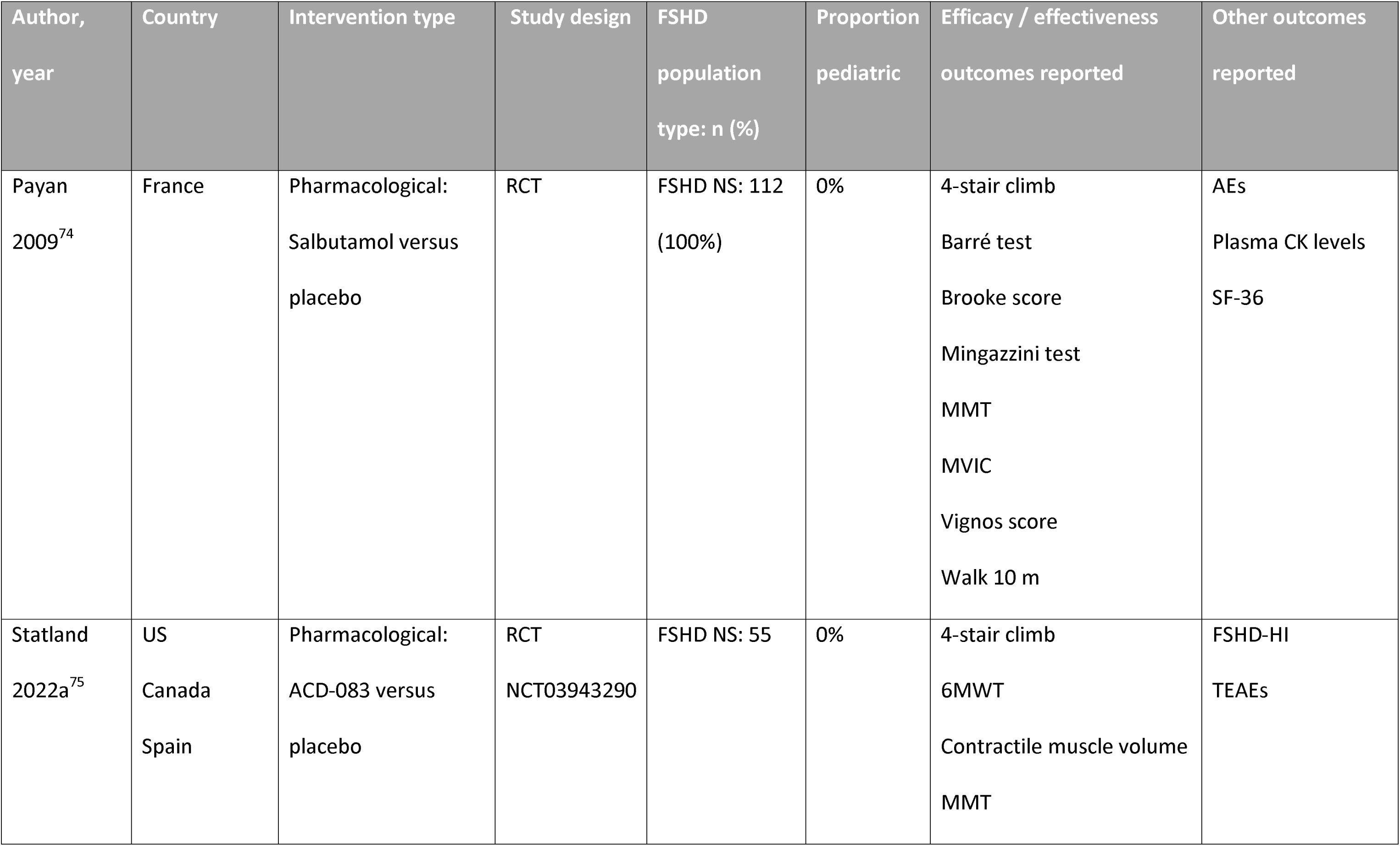
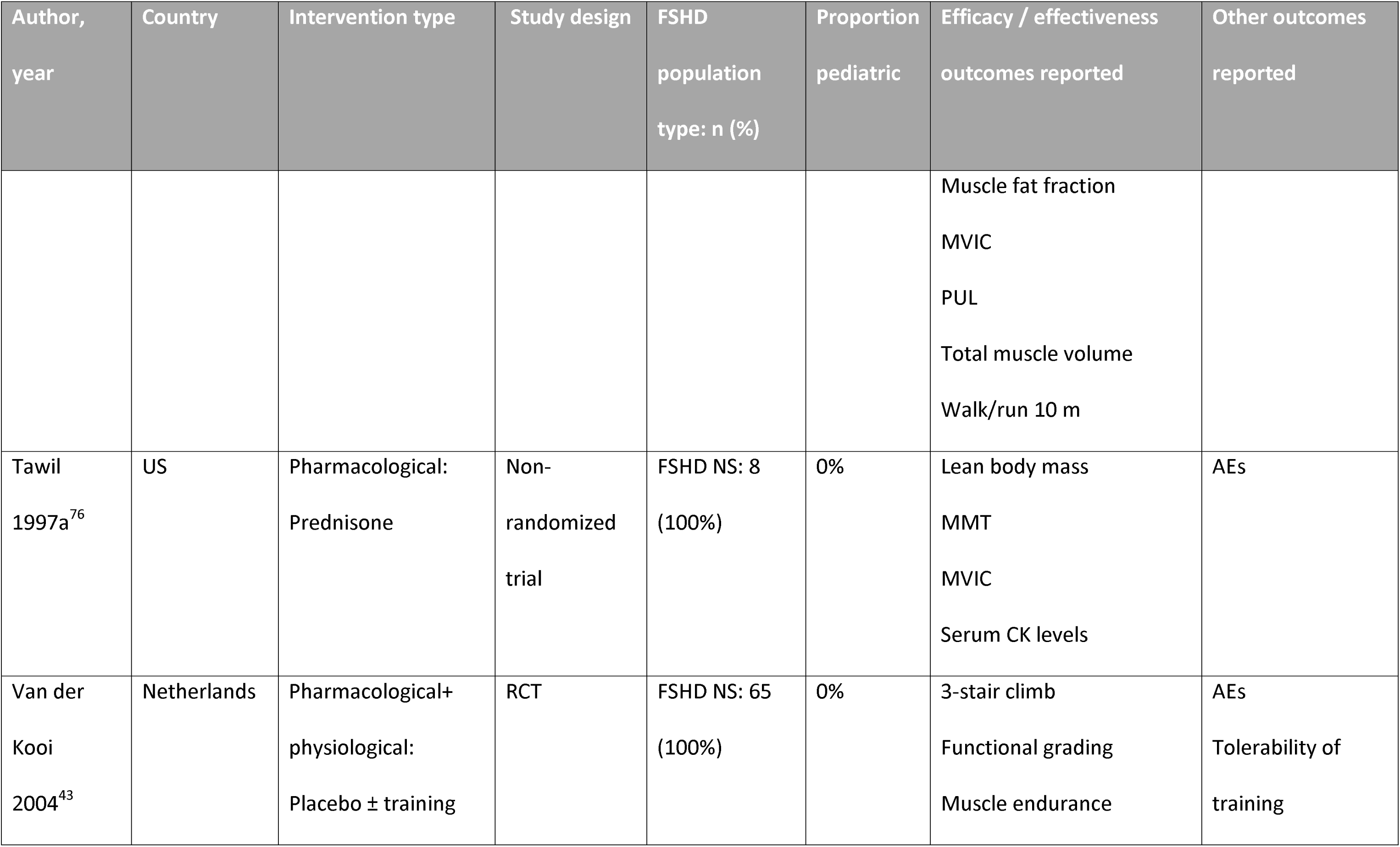
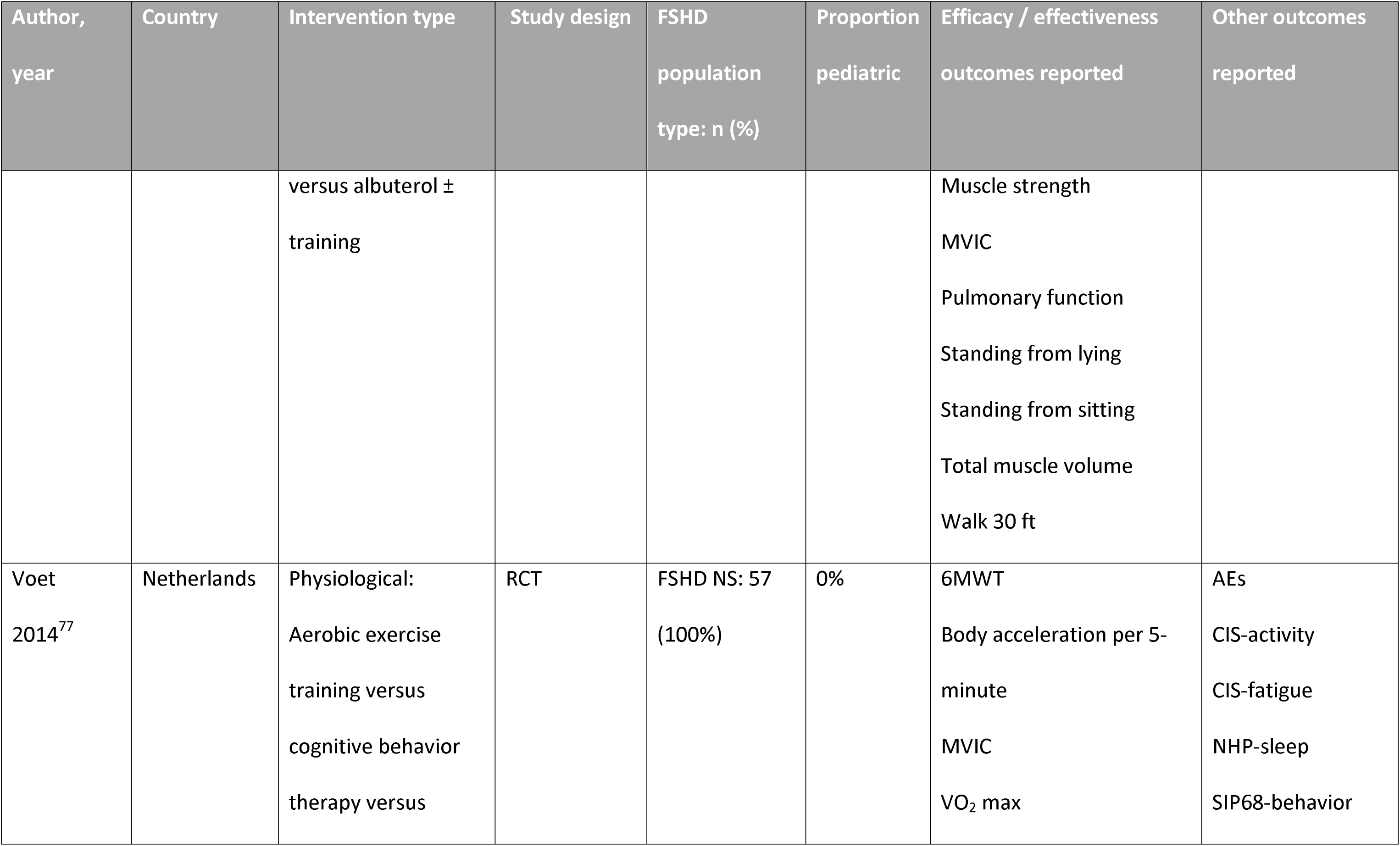
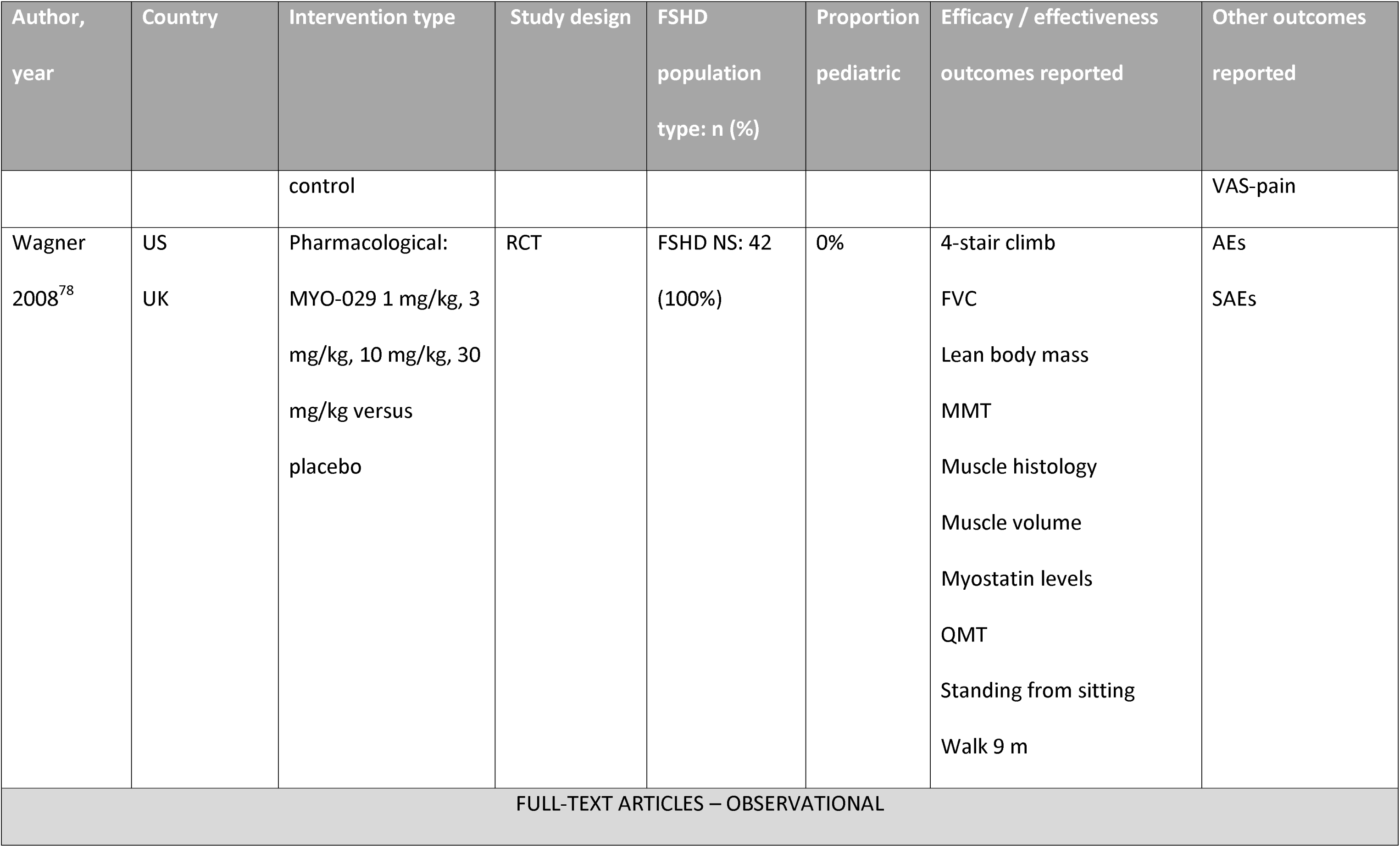
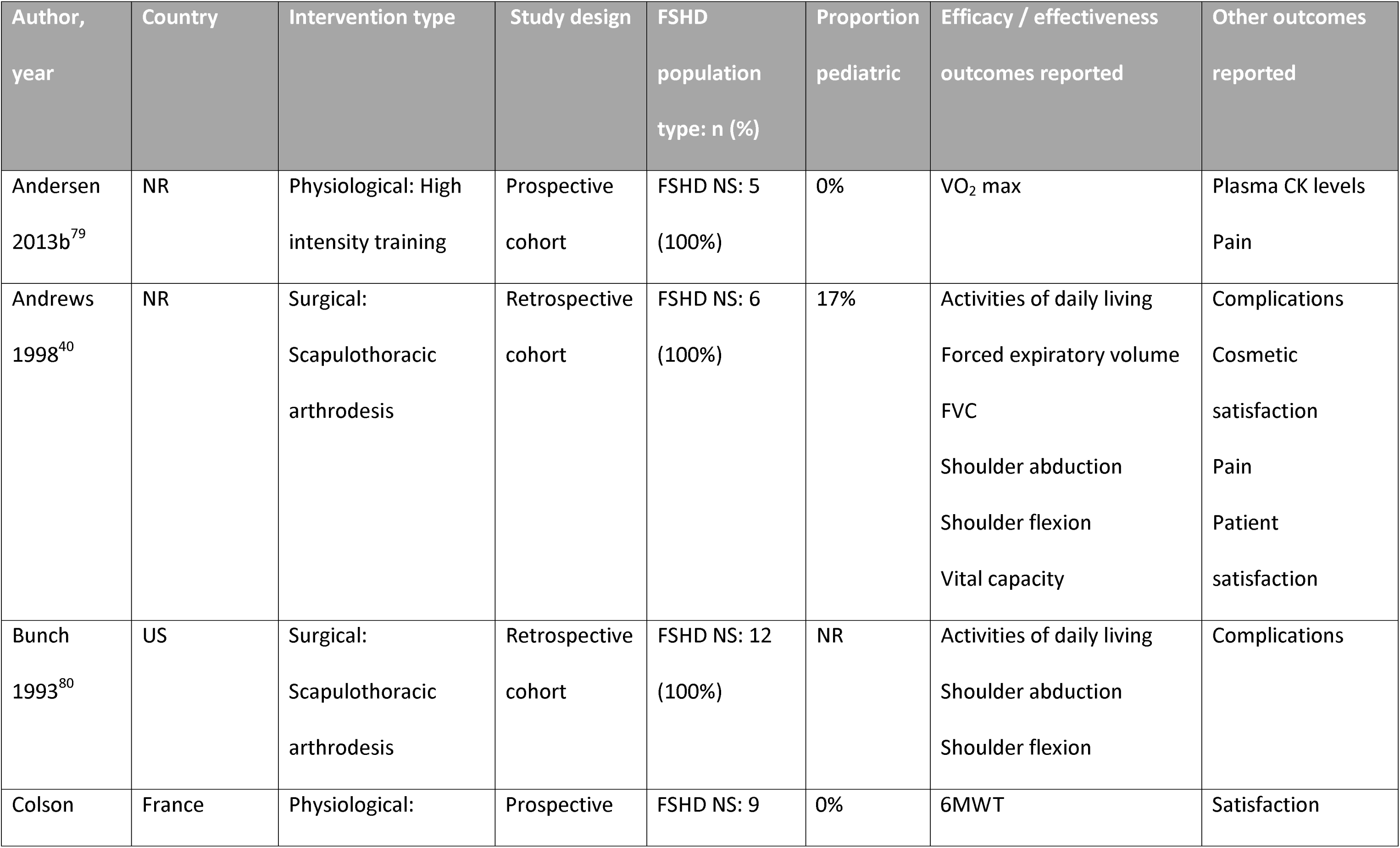
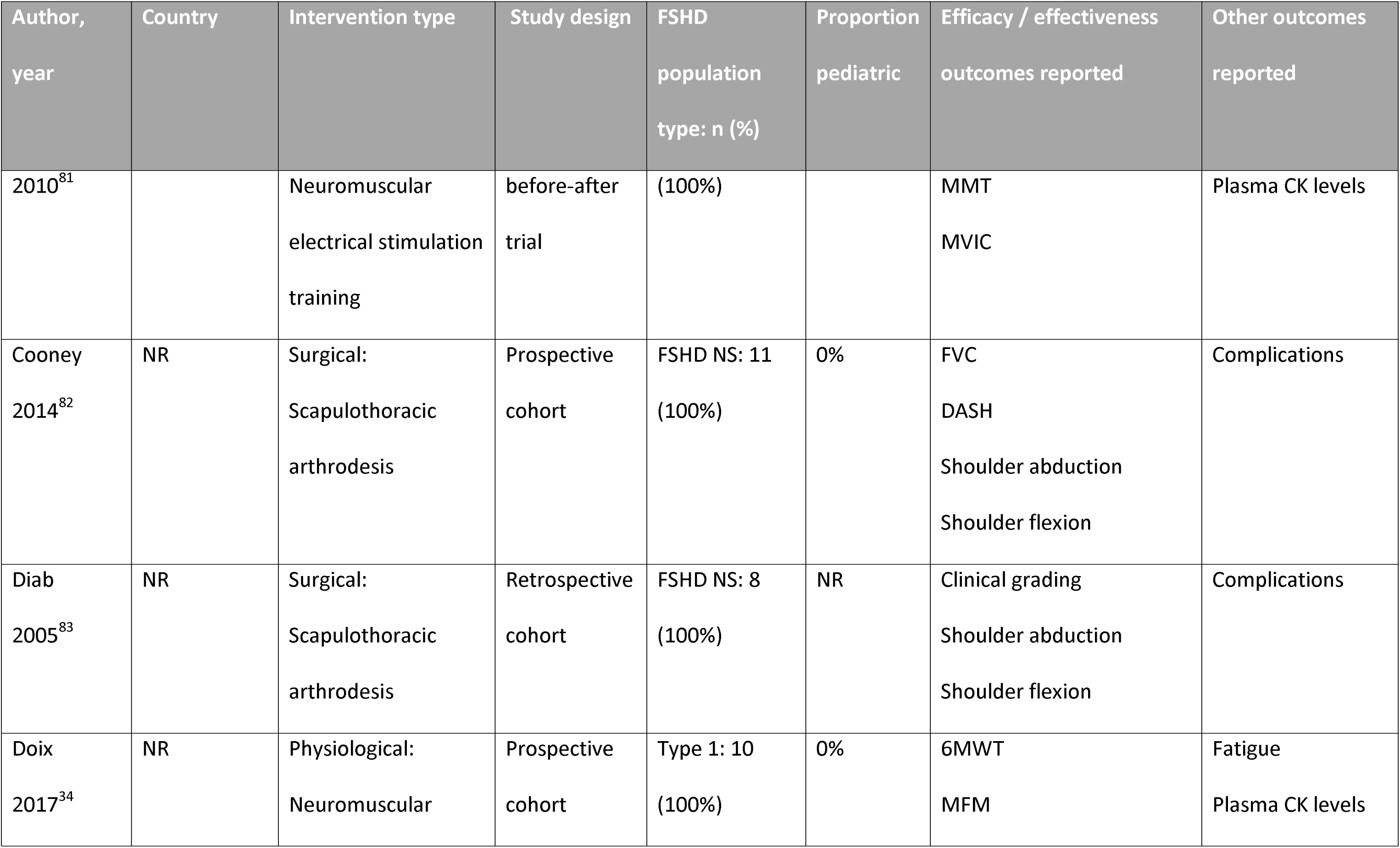
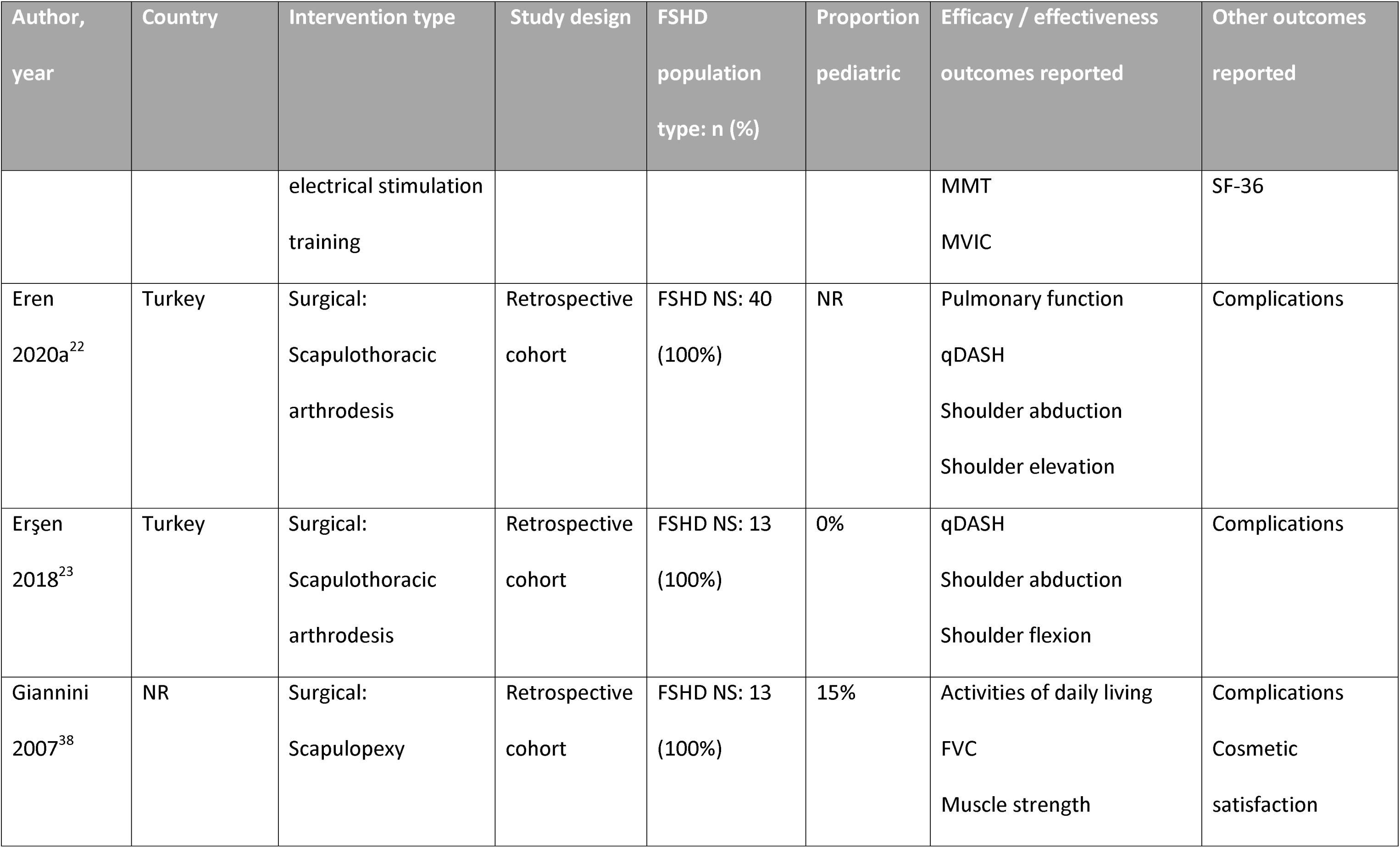
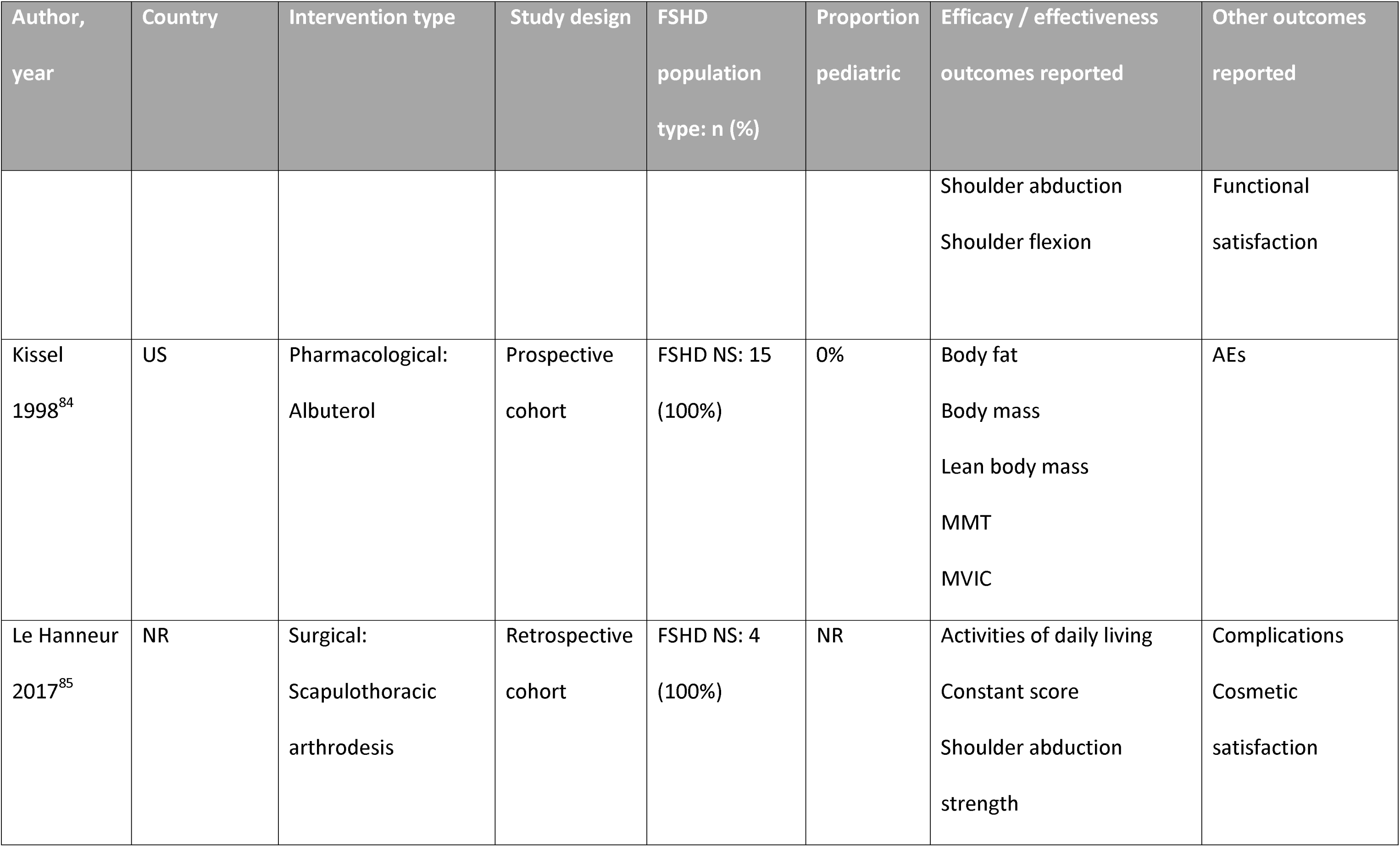
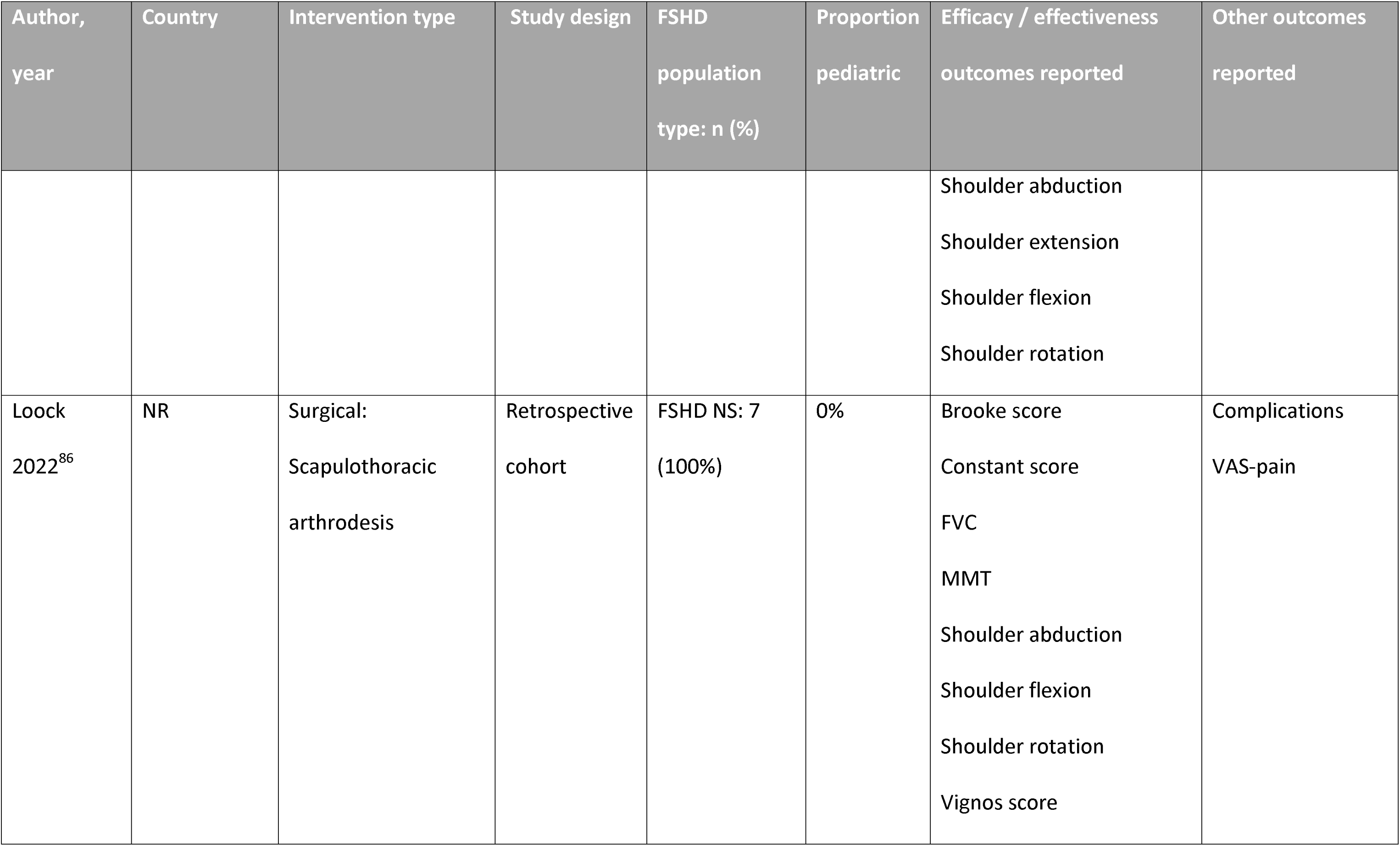
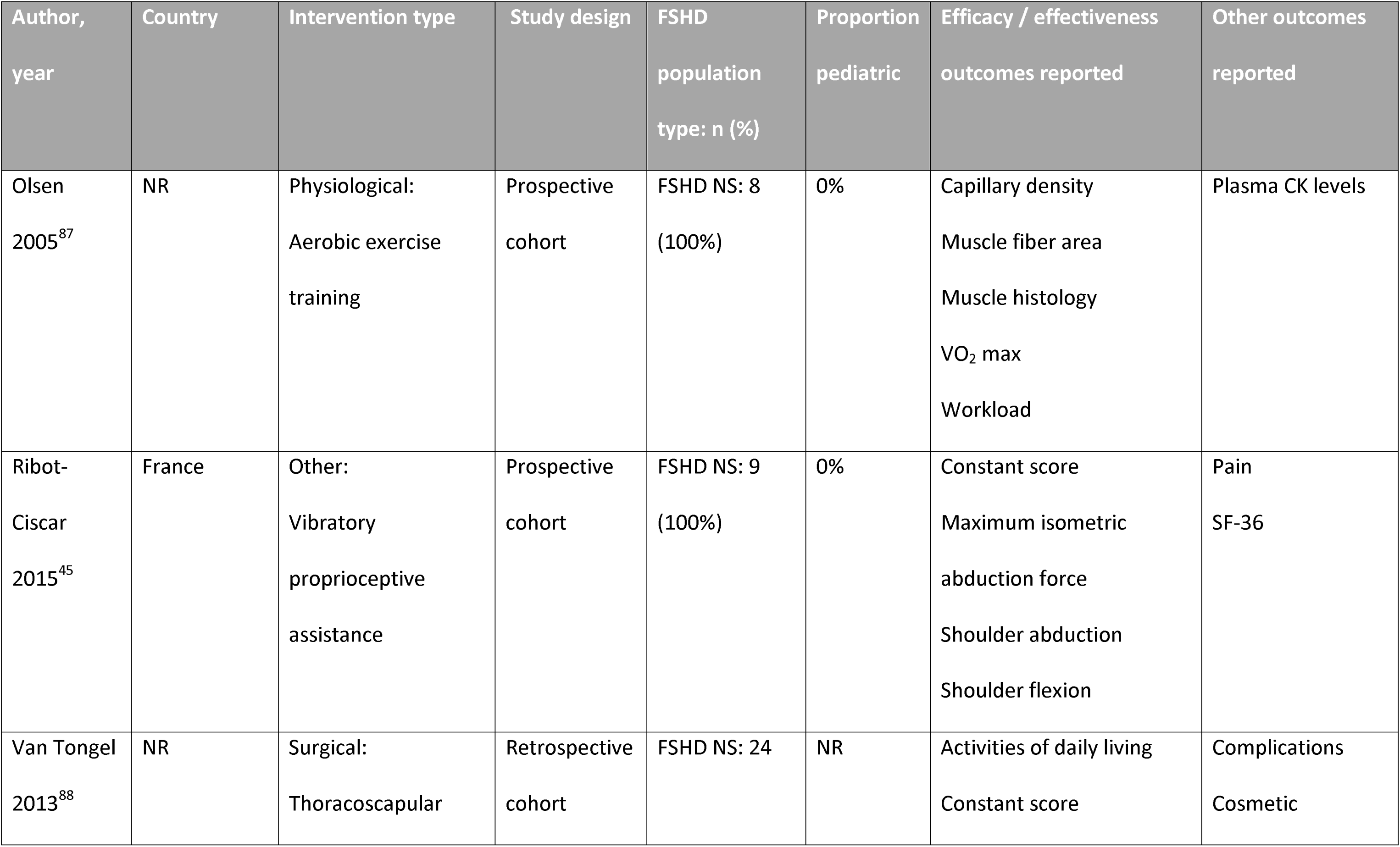
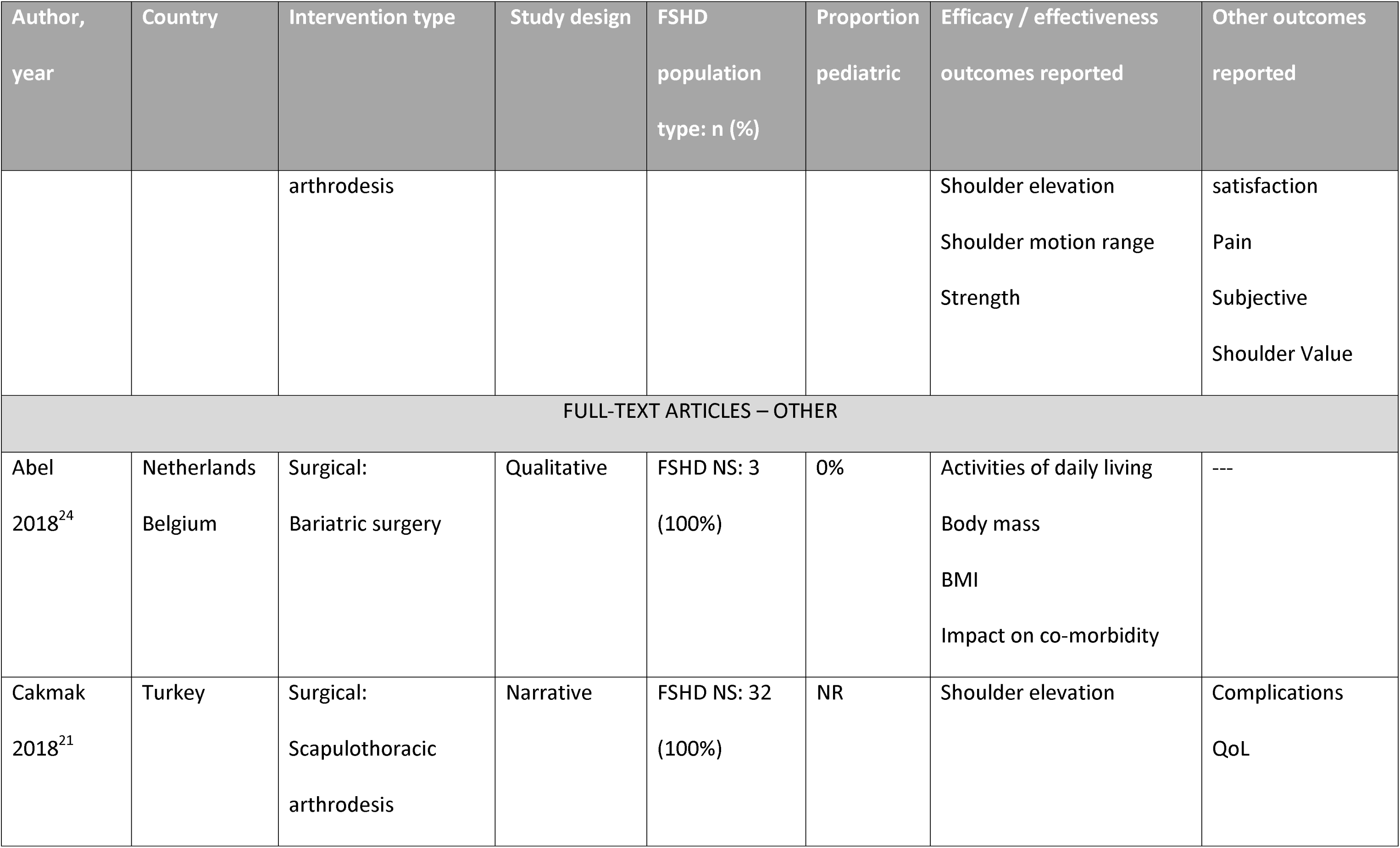
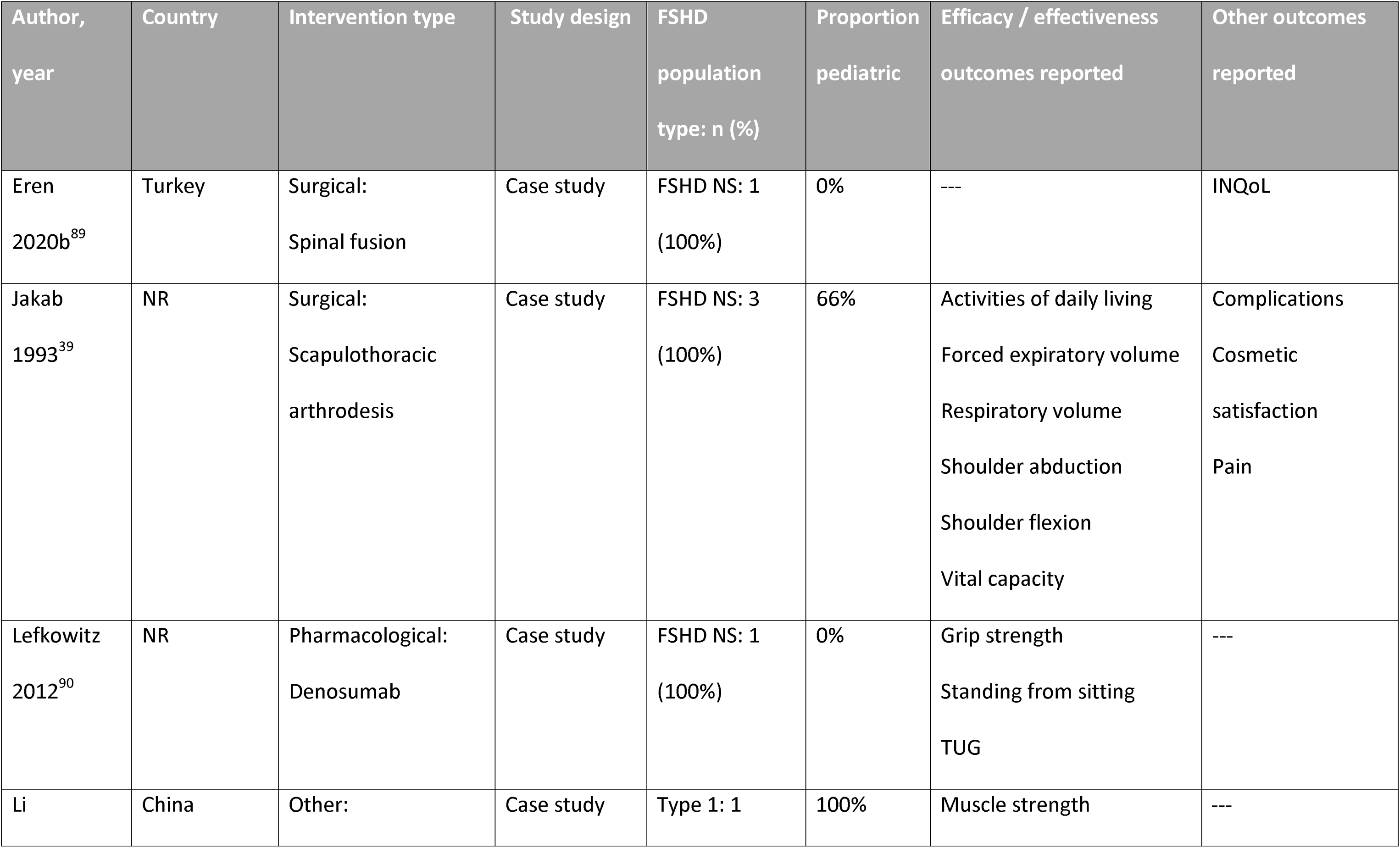
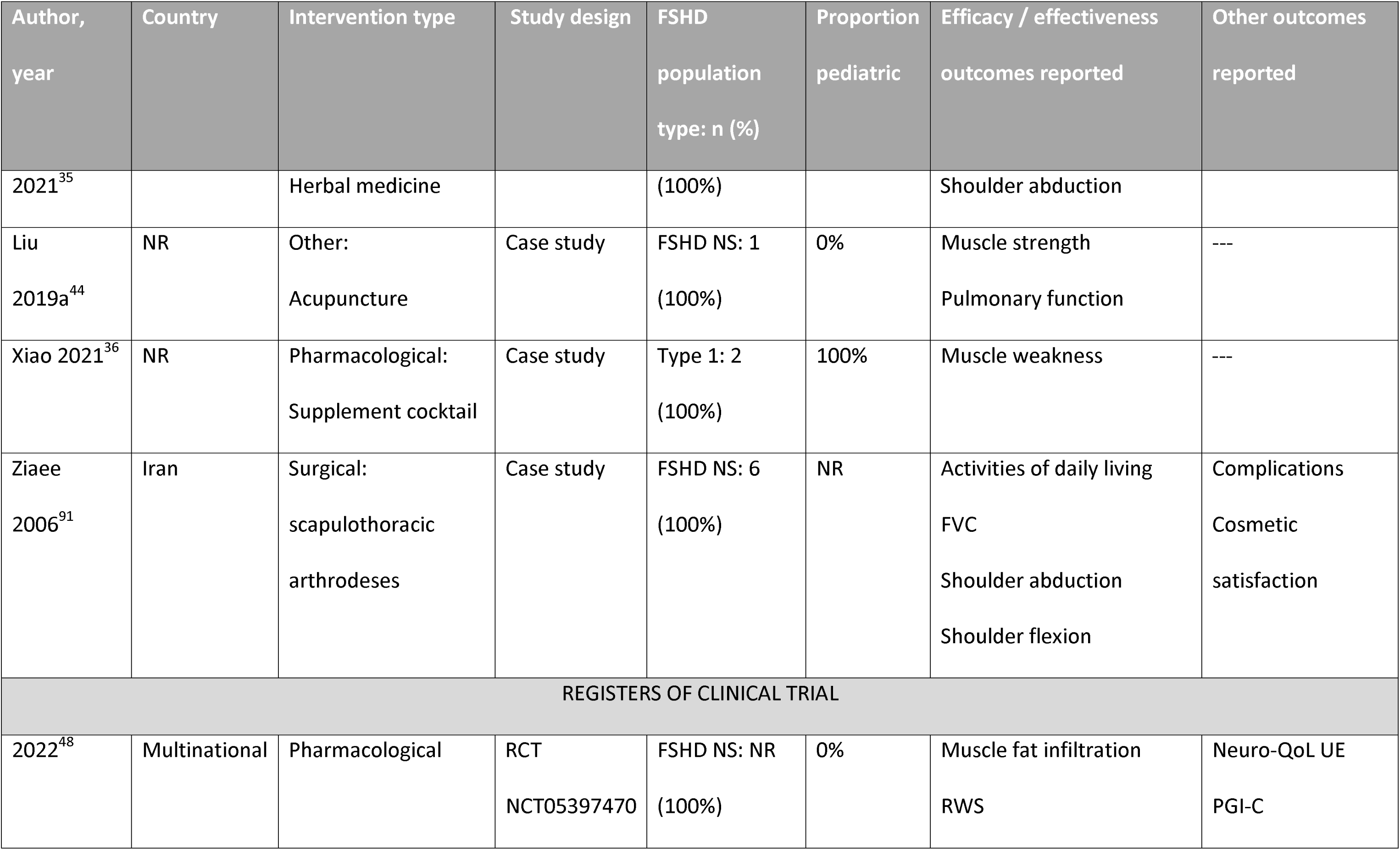
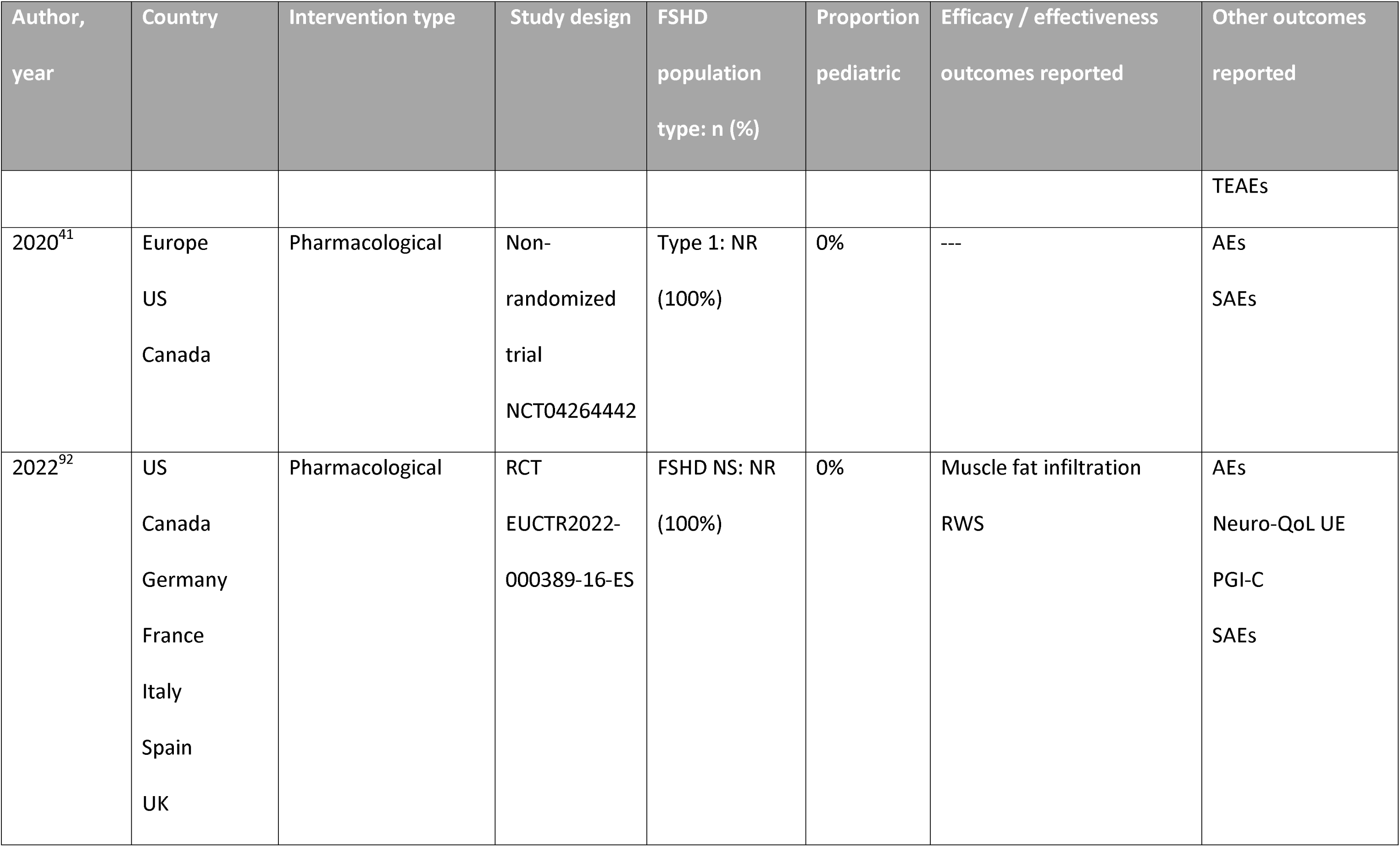
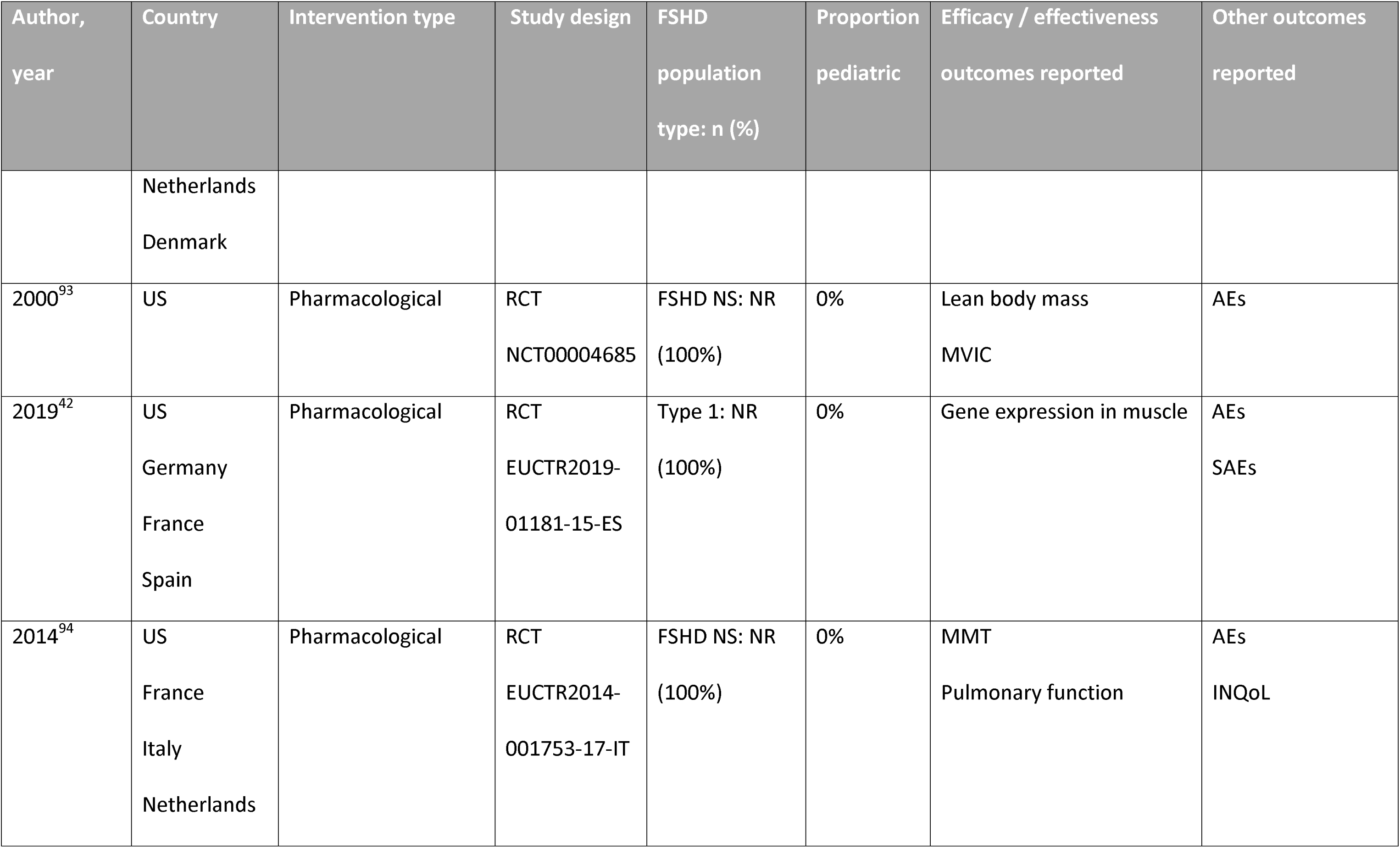
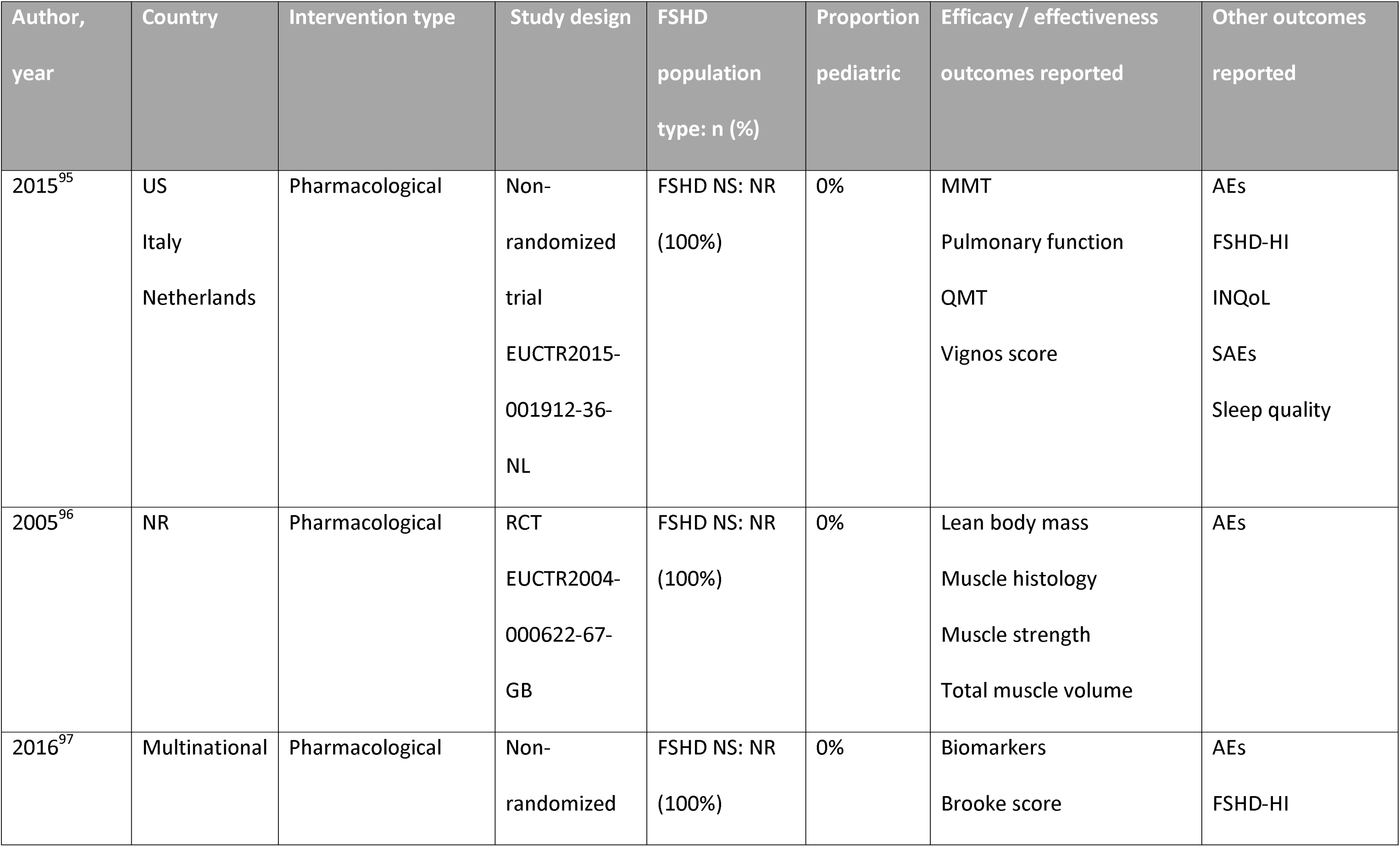
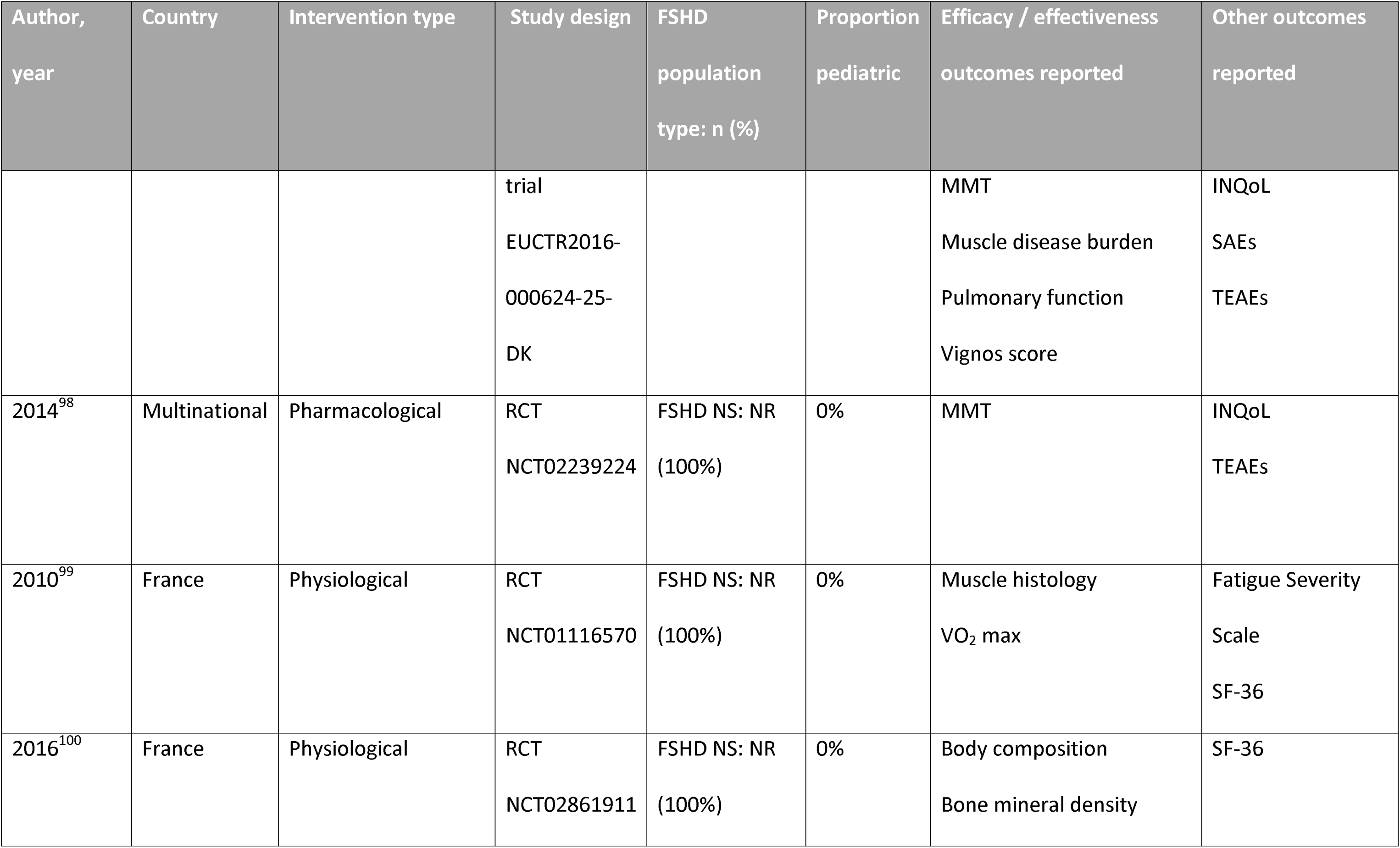
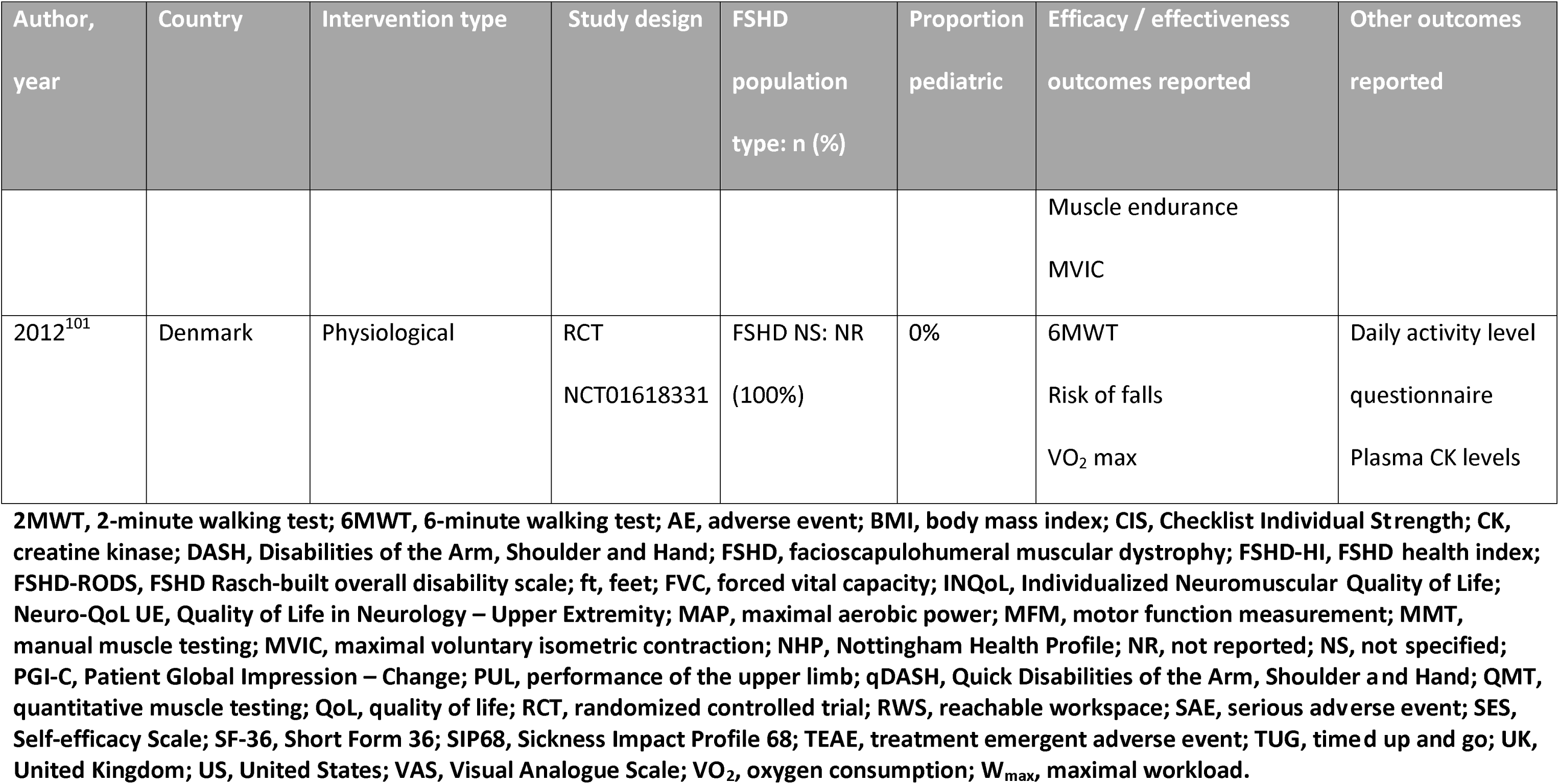
Study Characteristics and Treatment Outcomes Reported in Included Studies.

**Table Appendix 6:**
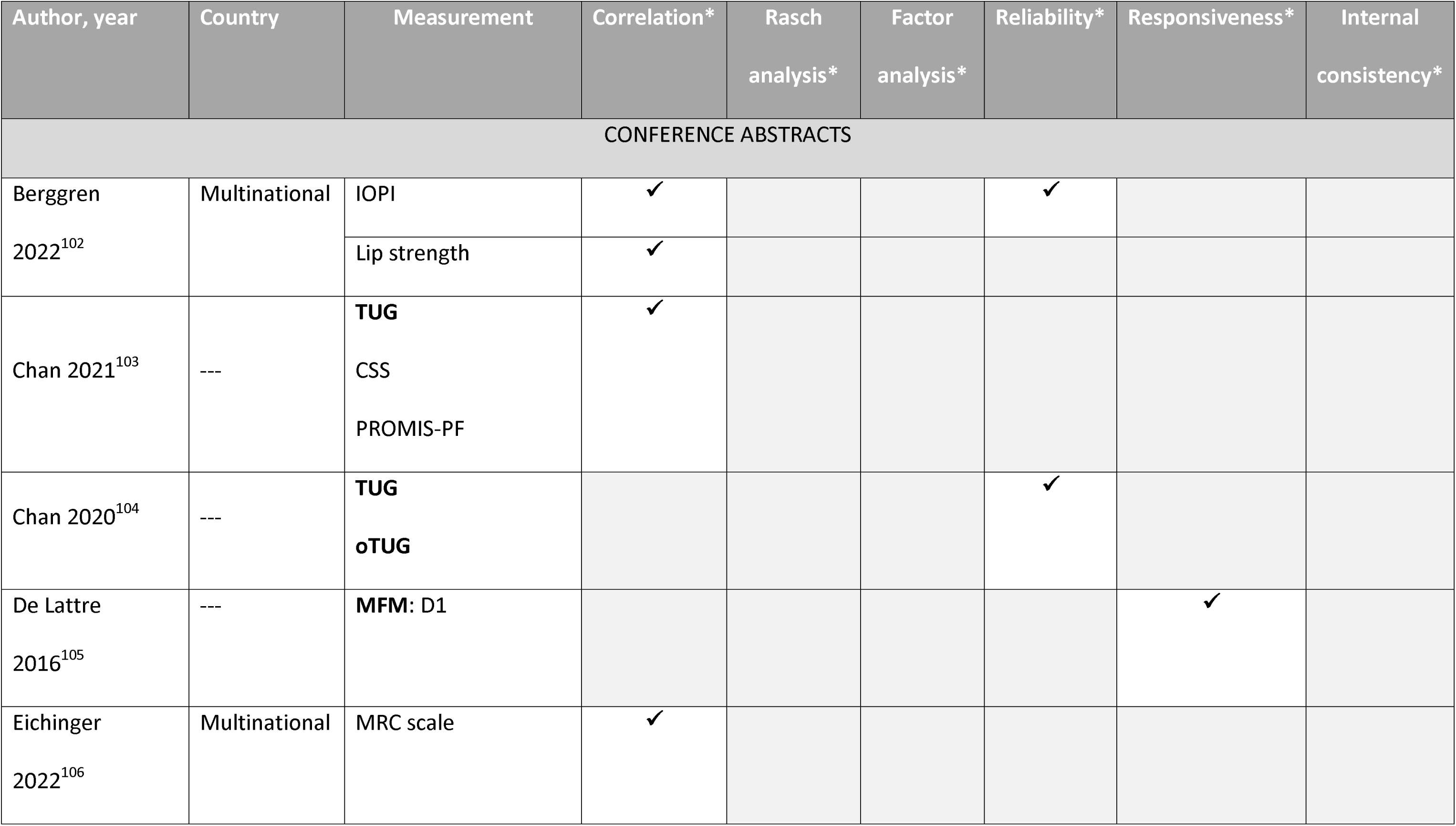
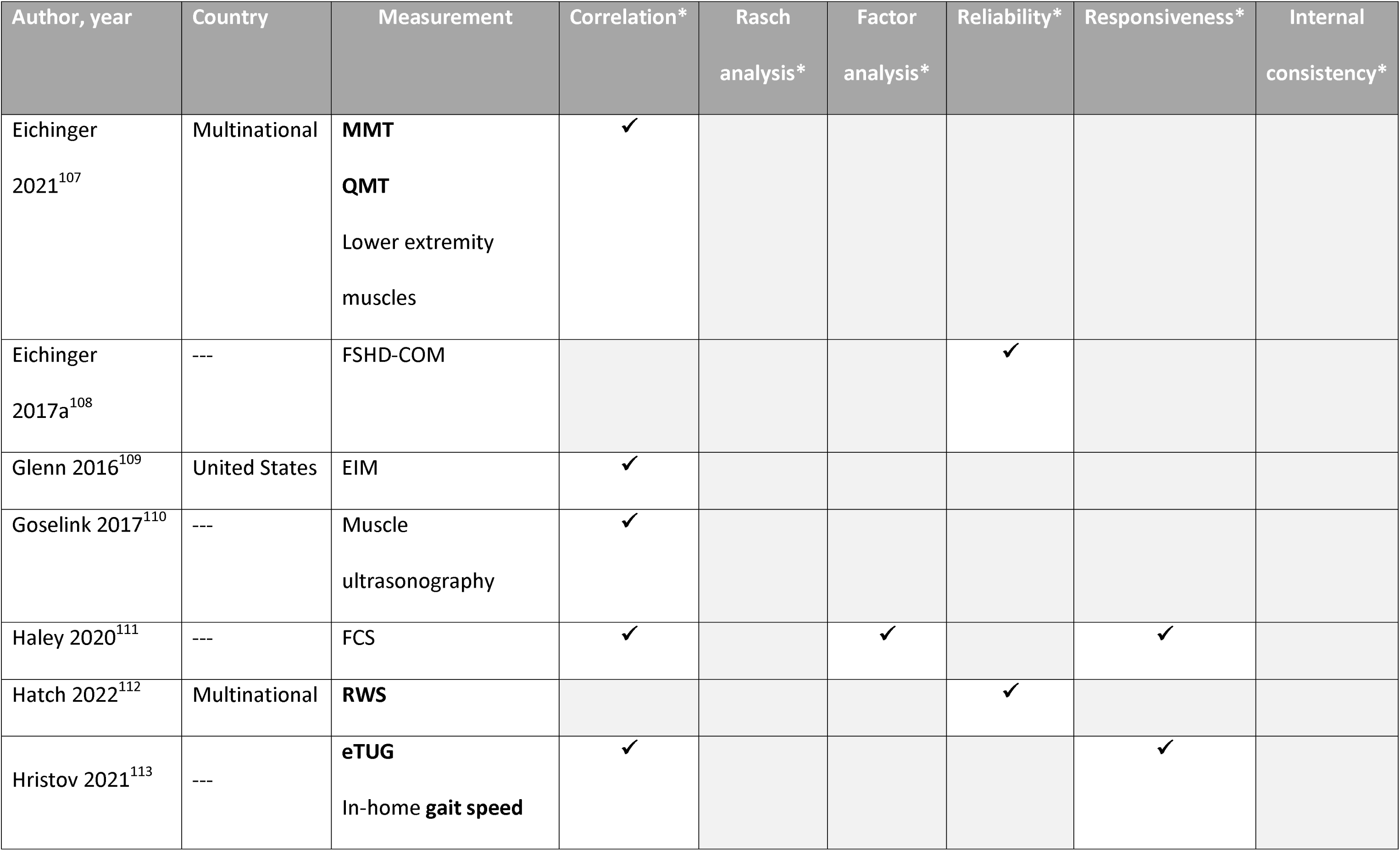
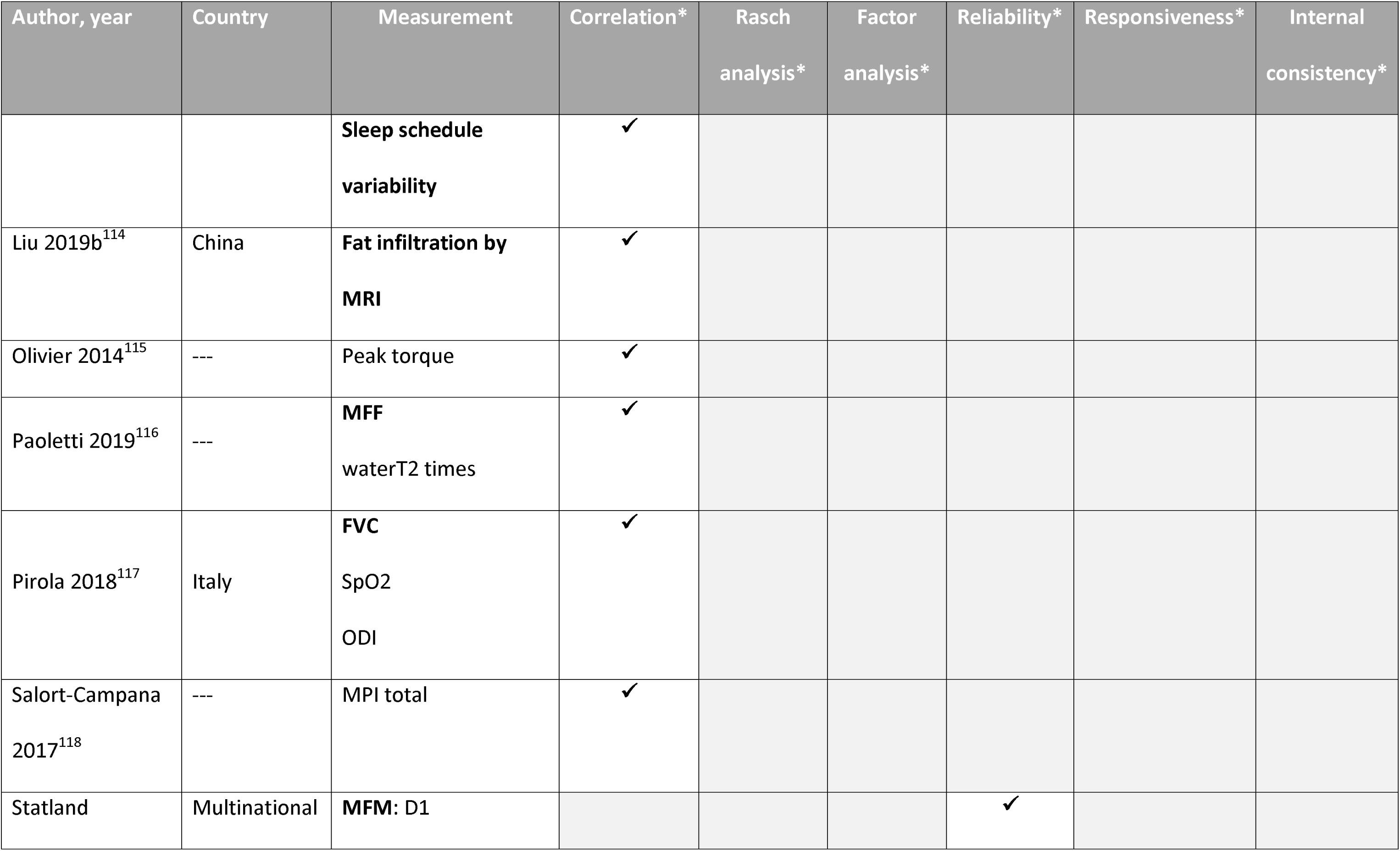
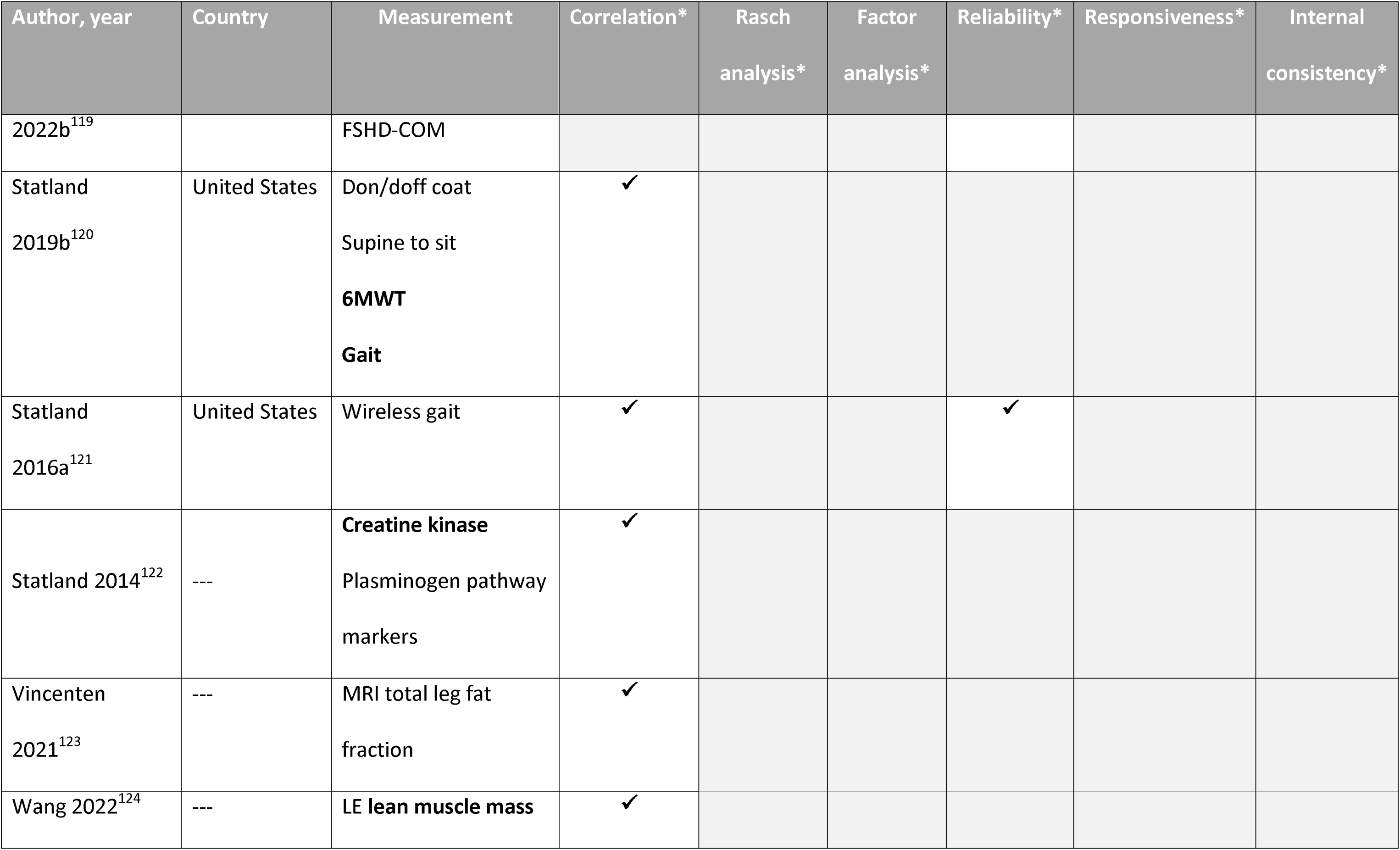
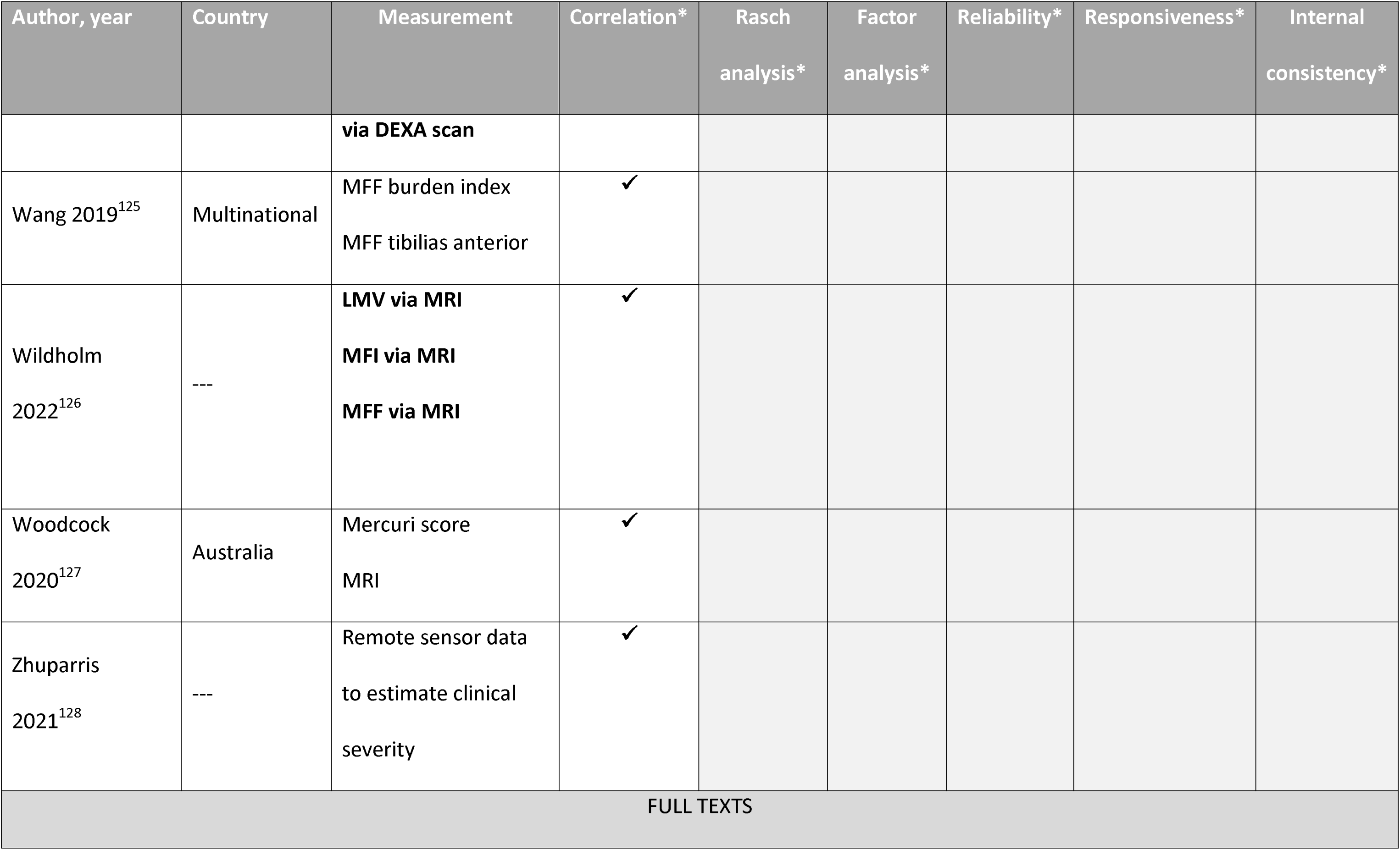
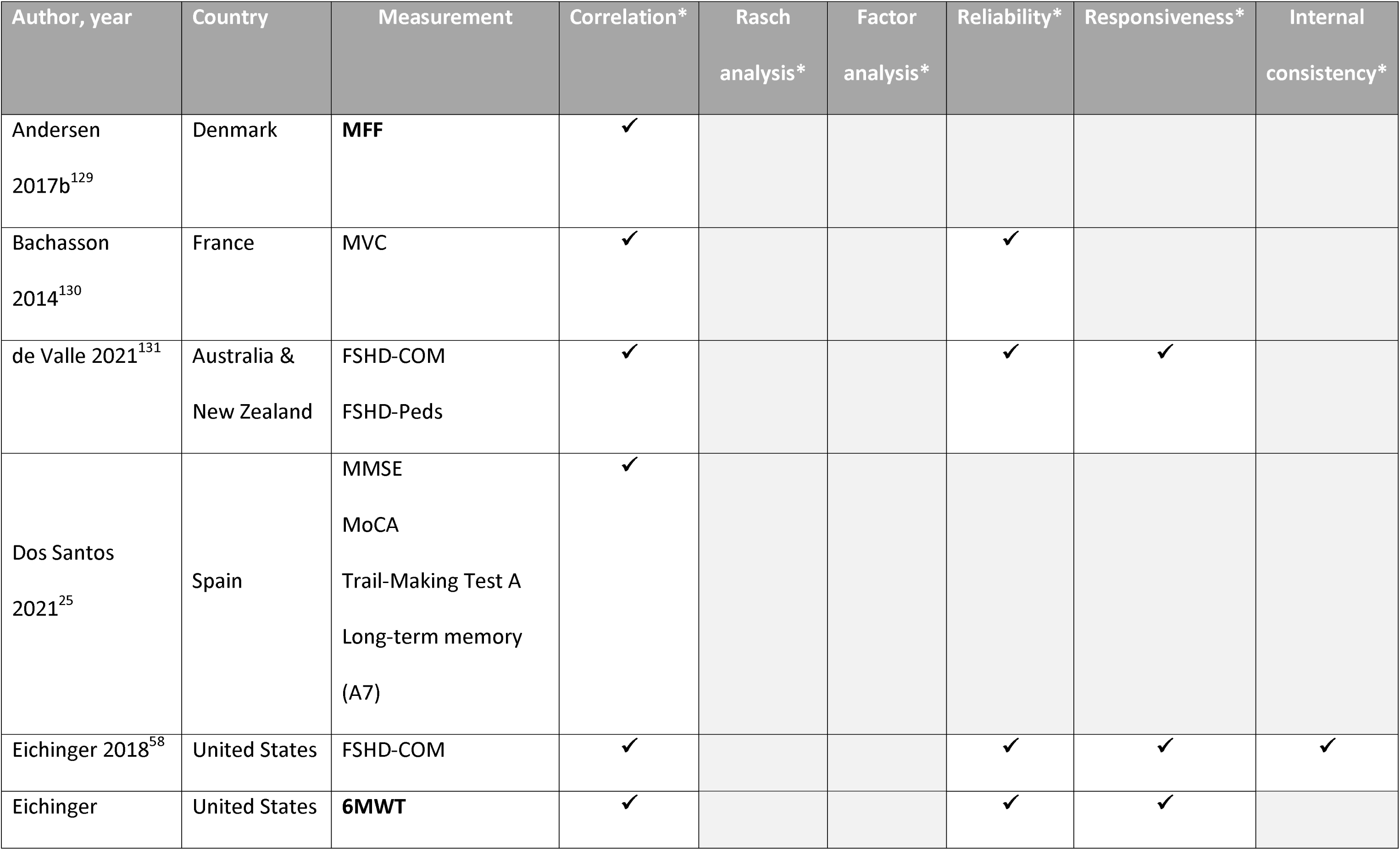
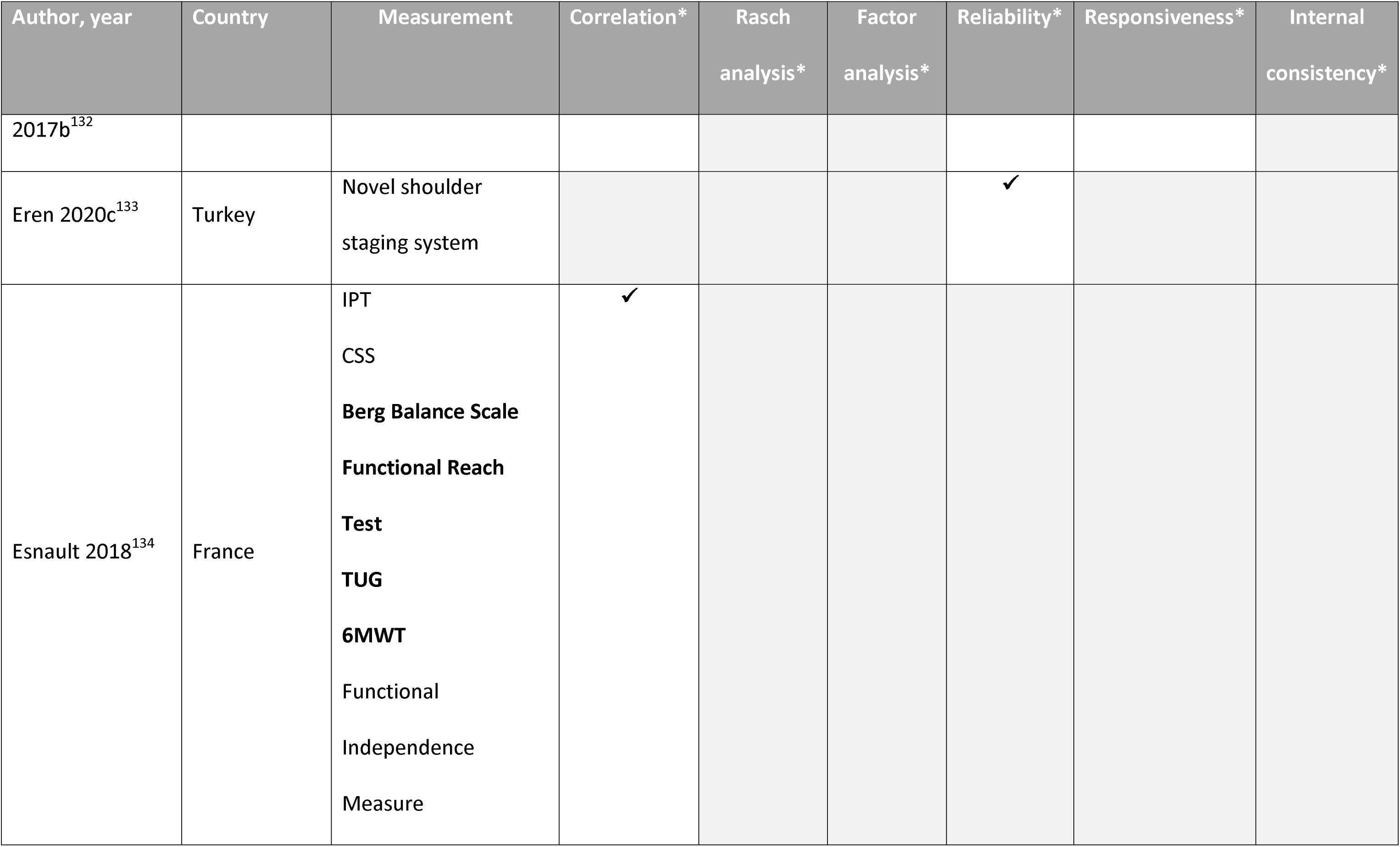
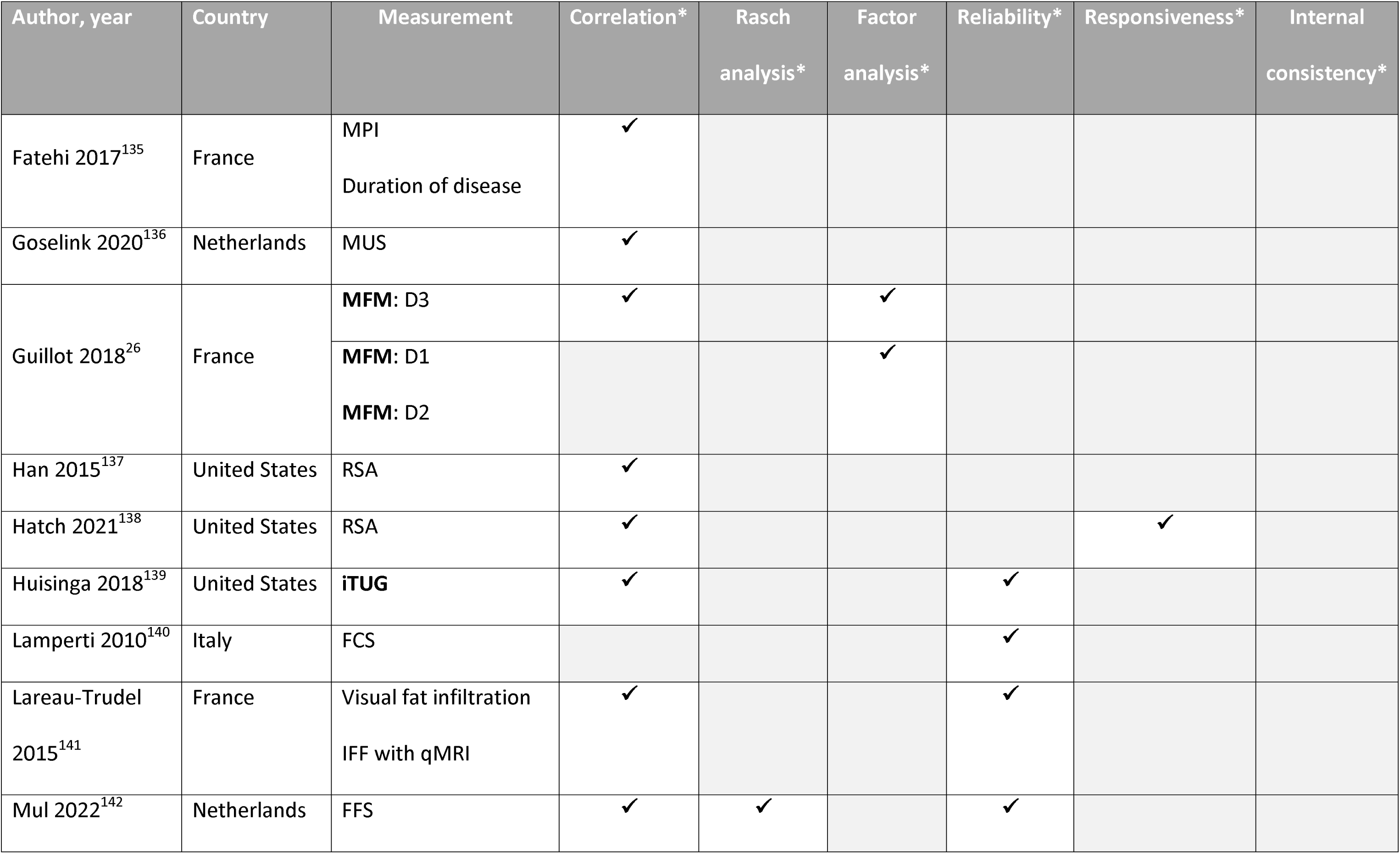
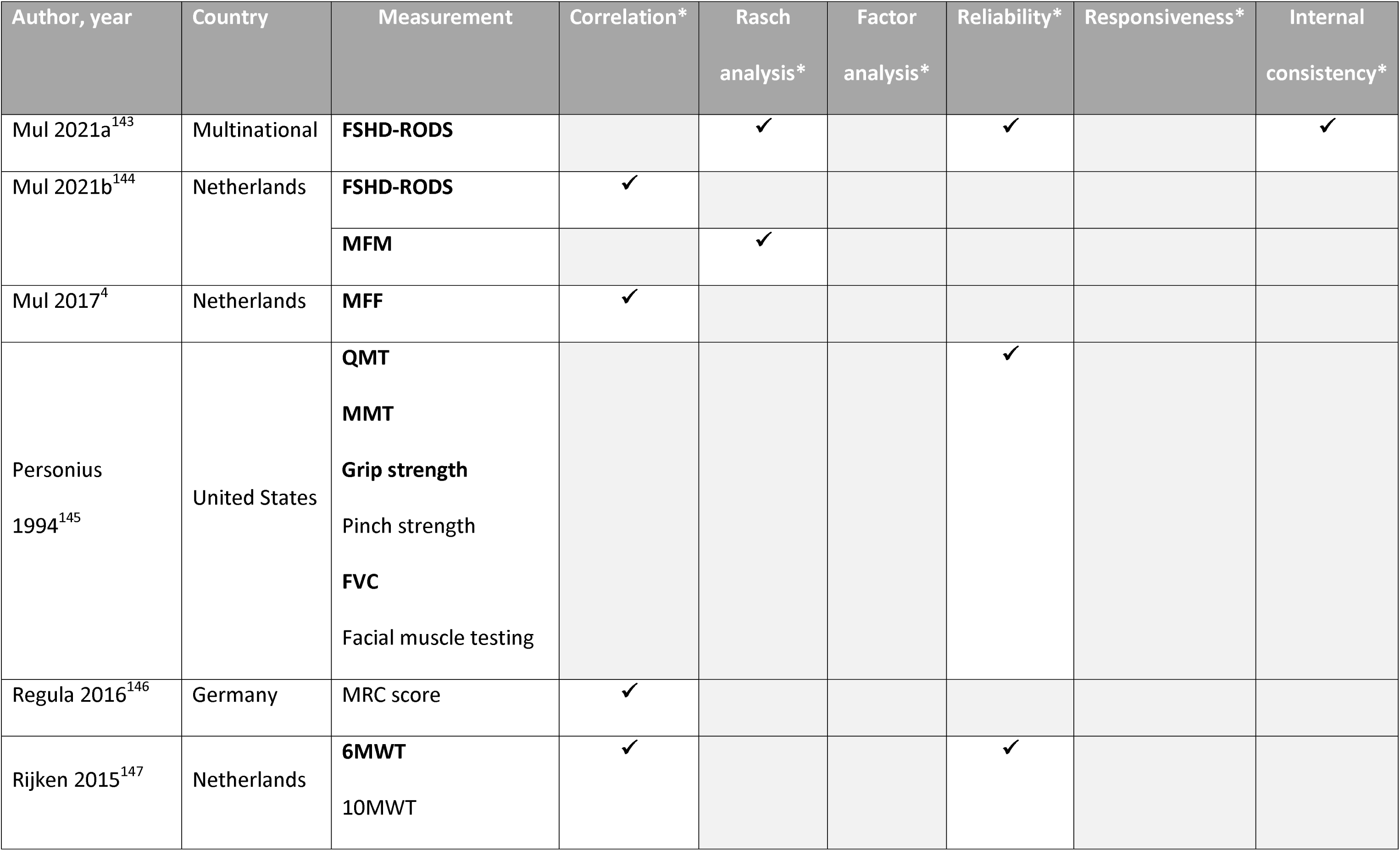
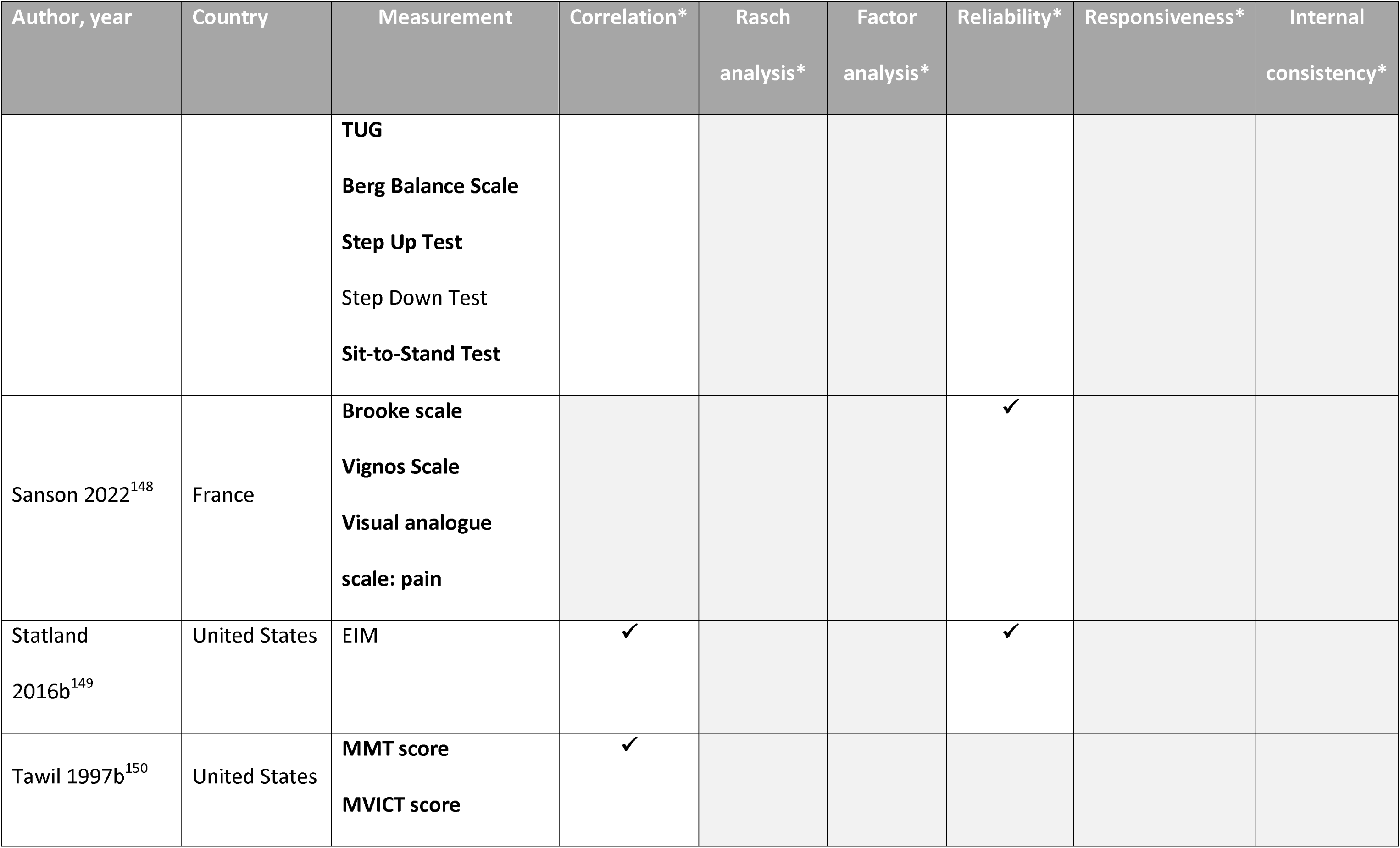
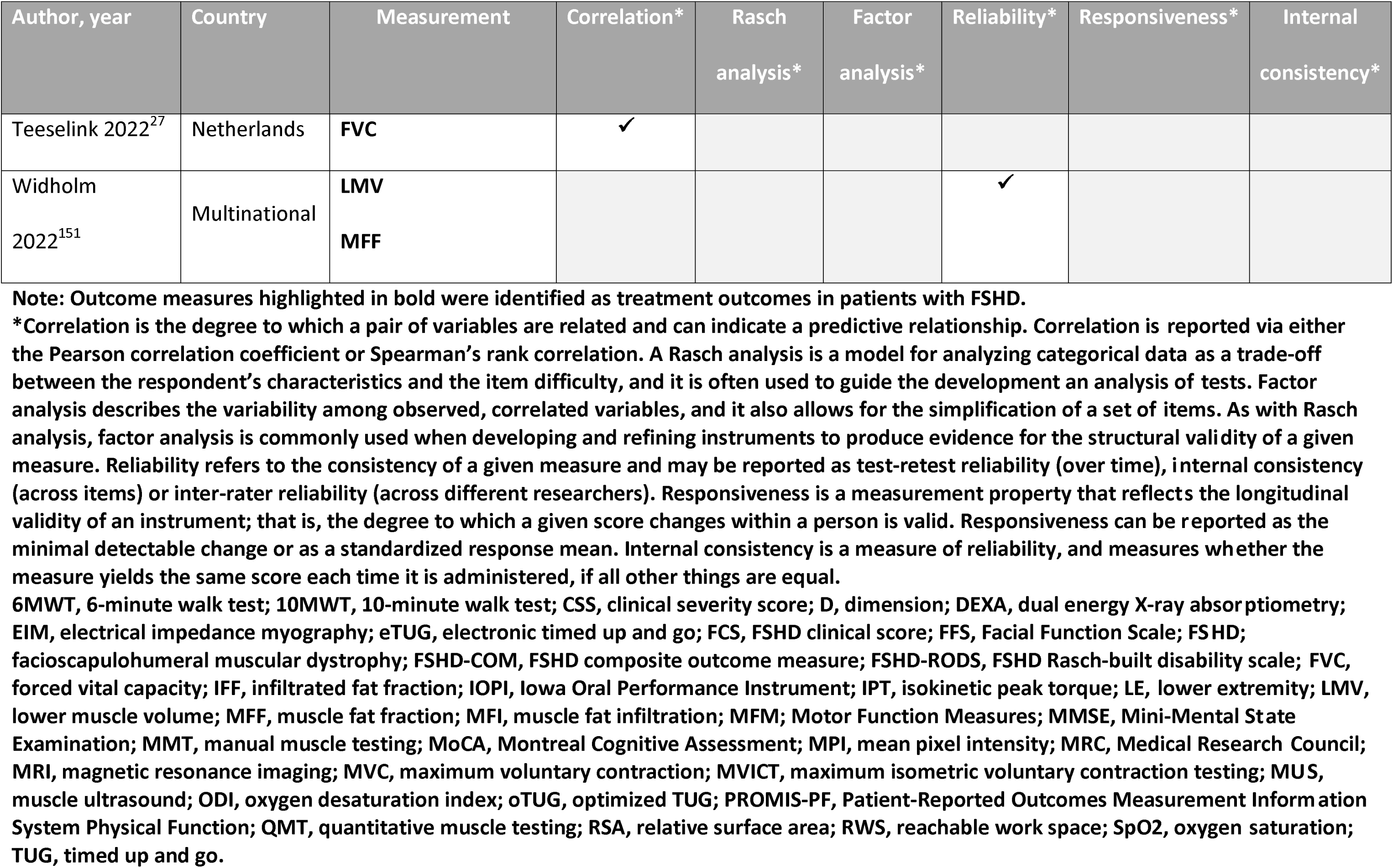
Validation Methods of Measures Reported in “Validation of Disease Outcome Measures” Studies.

| Author, year | Country | Measurement | Correlation* | Rasch<br>analysis* | Factor<br>analysis* | Reliability* | Responsiveness* | Internal<br>consistency* |
| --- | --- | --- | --- | --- | --- | --- | --- | --- |
| CONFERENCE ABSTRACTS |  |  |  |  |  |  |  |  |
| Berggren<br>2022<sup>102</sup> | Multinational | IOPI | ✓ |  |  | ✓ |  |  |
|  |  | Lip strength | ✓ |  |  |  |  |  |
| Chan 2021<sup>103</sup> | --- | **TUG**<br>CSS<br>PROMIS-PF | ✓ |  |  |  |  |  |
| Chan 2020<sup>104</sup> | --- | **TUG**<br>**oTUG** |  |  |  | ✓ |  |  |
| De Lattre<br>2016<sup>105</sup> | --- | **MFM: D1** |  |  |  |  | ✓ |  |
| Eichinger<br>2022<sup>106</sup> | Multinational | MRC scale | ✓ |  |  |  |  |  |
| Eichinger<br>2021<sup>107</sup> | Multinational | **MMT**<br>**QMT**<br>Lower extremity<br>muscles | ✓ |  |  |  |  |  |
| Eichinger<br>2017a<sup>108</sup> | --- | FSHD-COM |  |  |  | ✓ |  |  |
| Glenn 2016<sup>109</sup> | United States | EIM | ✓ |  |  |  |  |  |
| Goselink 2017<sup>110</sup> | --- | Muscle<br>ultrasonography | ✓ |  |  |  |  |  |
| Haley 2020<sup>111</sup> | --- | FCS | ✓ |  | ✓ |  | ✓ |  |
| Hatch 2022<sup>112</sup> | Multinational | **RWS** |  |  |  | ✓ |  |  |
| Hristov 2021<sup>113</sup> | --- | **eTUG**<br>In-home **gait speed** | ✓ |  |  |  | ✓ |  |
|  |  | **Sleep schedule**<br>**variability** | ✓ |  |  |  |  |  |
| Liu 2019b<sup>114</sup> | China | **Fat infiltration by**<br>**MRI** | ✓ |  |  |  |  |  |
| Olivier 2014<sup>115</sup> | --- | Peak torque | ✓ |  |  |  |  |  |
| Paoletti 2019<sup>116</sup> | --- | **MFF**<br>waterT2 times | ✓ |  |  |  |  |  |
| Pirola 2018<sup>117</sup> | Italy | **FVC**<br>SpO2<br>ODI | ✓ |  |  |  |  |  |
| Salort-Campana<br>2017<sup>118</sup> | --- | MPI total | ✓ |  |  |  |  |  |
| Statland | Multinational | **MFM: D1** |  |  |  | ✓ |  |  |
| 2022b<sup>119</sup> |  | FSHD-COM |  |  |  |  |  |  |
| Statland<br>2019b<sup>120</sup> | United States | Don/doff coat<br>Supine to sit<br>**6MWT**<br>**Gait** | ✓ |  |  |  |  |  |
| Statland<br>2016a<sup>121</sup> | United States | Wireless gait | ✓ |  |  | ✓ |  |  |
| Statland 2014<sup>122</sup> | --- | **Creatine kinase**<br>Plasminogen pathway<br>markers | ✓ |  |  |  |  |  |
| Vincenten<br>2021<sup>123</sup> | --- | MRI total leg fat<br>fraction | ✓ |  |  |  |  |  |
| Wang 2022<sup>124</sup> | --- | LE lean muscle mass **via DEXA scan** | ✓ |  |  |  |  |  |
| Wang 2019<sup>125</sup> | Multinational | MFF burden index<br>MFF tibialis anterior | ✓ |  |  |  |  |  |
| Wildholm<br>2022<sup>126</sup> | --- | **LMV via MRI**<br>**MFI via MRI**<br>**MFF via MRI** | ✓ |  |  |  |  |  |
| Woodcock<br>2020<sup>127</sup> | Australia | Mercuri score<br>MRI | ✓ |  |  |  |  |  |
| Zhuparris<br>2021<sup>128</sup> | --- | Remote sensor data<br>to estimate clinical<br>severity | ✓ |  |  |  |  |  |
| FULL TEXTS |  |  |  |  |  |  |  |  |
| Andersen<br>2017<sup>b</sup><sup>129</sup> | Denmark | **MFF** | ✓ |  |  |  |  |  |
| Bachasson<br>2014<sup>130</sup> | France | MVC | ✓ |  |  | ✓ |  |  |
| de Valle 2021<sup>131</sup> | Australia &<br>New Zealand | FSHD-COM<br>FSHD-Peds | ✓ |  |  | ✓ | ✓ |  |
| Dos Santos<br>2021<sup>25</sup> | Spain | MMSE<br>MoCA<br>Trail-Making Test A<br>Long-term memory<br>(A7) | ✓ |  |  |  |  |  |
| Eichinger 2018<sup>58</sup> | United States | FSHD-COM | ✓ |  |  | ✓ | ✓ | ✓ |
| Eichinger | United States | **6MWT** | ✓ |  |  | ✓ | ✓ |  |
| 2017b<sup>132</sup> |  |  |  |  |  |  |  |  |
| Eren 2020c<sup>133</sup> | Turkey | Novel shoulder<br>staging system |  |  |  | ✓ |  |  |
| Esnault 2018<sup>134</sup> | France | IPT<br>CSS<br>**Berg Balance Scale**<br>**Functional Reach**<br>**Test**<br>**TUG**<br>**6MWT**<br>Functional<br>Independence<br>Measure | ✓ |  |  |  |  |  |
| Fatehi 2017<sup>135</sup> | France | MPI<br>Duration of disease | ✓ |  |  |  |  |  |
| Goselink 2020<sup>136</sup> | Netherlands | MUS | ✓ |  |  |  |  |  |
| Guillot 2018<sup>26</sup> | France | **MFM: D3** | ✓ |  | ✓ |  |  |  |
|  |  | **MFM: D1** |  |  | ✓ |  |  |  |
|  |  | **MFM: D2** |  |  |  |  |  |  |
| Han 2015<sup>137</sup> | United States | RSA | ✓ |  |  |  |  |  |
| Hatch 2021<sup>138</sup> | United States | RSA | ✓ |  |  |  | ✓ |  |
| Huisinga 2018<sup>139</sup> | United States | **iTUG** | ✓ |  |  | ✓ |  |  |
| Lamperti 2010<sup>140</sup> | Italy | FCS |  |  |  | ✓ |  |  |
| Lareau-Trudel<br>2015<sup>141</sup> | France | Visual fat infiltration<br>IFF with qMRI | ✓ |  |  | ✓ |  |  |
| Mul 2022<sup>142</sup> | Netherlands | FFS | ✓ | ✓ |  | ✓ |  |  |
| Mul 2021a<sup>143</sup> | Multinational | **FSHD-RODS** |  | ✓ |  | ✓ |  | ✓ |
| Mul 2021b<sup>144</sup> | Netherlands | **FSHD-RODS** | ✓ |  |  |  |  |  |
|  |  | **MFM** |  | ✓ |  |  |  |  |
| Mul 2017<sup>4</sup> | Netherlands | **MFF** | ✓ |  |  |  |  |  |
| Personius<br>1994<sup>145</sup> | United States | **QMT** |  |  |  | ✓ |  |  |
|  |  | **MMT** |  |  |  |  |  |  |
|  |  | **Grip strength** |  |  |  |  |  |  |
|  |  | Pinch strength |  |  |  |  |  |  |
|  |  | **FVC** |  |  |  |  |  |  |
|  |  | Facial muscle testing |  |  |  |  |  |  |
| Regula 2016<sup>146</sup> | Germany | MRC score | ✓ |  |  |  |  |  |
| Rijken 2015<sup>147</sup> | Netherlands | **6MWT** | ✓ |  |  | ✓ |  |  |
|  |  | 10MWT |  |  |  |  |  |  |
|  |  | **TUG**<br>**Berg Balance Scale**<br>**Step Up Test**<br>Step Down Test<br>**Sit-to-Stand Test** |  |  |  |  |  |  |
| Sanson 2022<sup>148</sup> | France | **Brooke scale**<br>**Vignos Scale**<br>**Visual analogue**<br>**scale: pain** |  |  |  | ✓ |  |  |
| Statland<br>2016b<sup>149</sup> | United States | EIM | ✓ |  |  | ✓ |  |  |
| Tawil 1997b<sup>150</sup> | United States | **MMT score**<br>**MVICT score** | ✓ |  |  |  |  |  |
| Teeselink 2022<sup>27</sup> | Netherlands | **FVC** | ✓ |  |  |  |  |  |
| Widholm<br>2022<sup>151</sup> | Multinational | **LMV**<br>**MFF** |  |  |  | ✓ |  |  |

**Note: Outcome measures highlighted in bold were identified as treatment outcomes in patients with FSHD.**
\*Correlation is the degree to which a pair of variables are related and can indicate a predictive relationship. Correlation is reported via either the Pearson correlation coefficient or Spearman's rank correlation. A Rasch analysis is a model for analyzing categorical data as a trade-off between the respondent's characteristics and the item difficulty, and it is often used to guide the development an analysis of tests. Factor analysis describes the variability among observed, correlated variables, and it also allows for the simplification of a set of items. As with Rasch analysis, factor analysis is commonly used when developing and refining instruments to produce evidence for the structural validity of a given measure. Reliability refers to the consistency of a given measure and may be reported as test-retest reliability (over time), internal consistency (across items) or inter-rater reliability (across different researchers). Responsiveness is a measurement property that reflects the longitudinal validity of an instrument; that is, the degree to which a given score changes within a person is valid. Responsiveness can be reported as the minimal detectable change or as a standardized response mean. Internal consistency is a measure of reliability, and measures whether the measure yields the same score each time it is administered, if all other things are equal.
6MWT, 6-minute walk test; 10MWT, 10-minute walk test; CSS, clinical severity score; D, dimension; DEXA, dual energy X-ray absorptiometry; EIM, electrical impedance myography; eTUG, electronic timed up and go; FCS, FSHD clinical score; FFS, Facial Function Scale; FSHD; facioscapulohumeral muscular dystrophy; FSHD-COM, FSHD composite outcome measure; FSHD-RODS, FSHD Rasch-built disability scale; FVC, forced vital capacity; IFF, infiltrated fat fraction; IOPI, Iowa Oral Performance Instrument; IPT, isokinetic peak torque; LE, lower extremity; LMV, lower muscle volume; MFF, muscle fat fraction; MFI, muscle fat infiltration; MFM; Motor Function Measures; MMSE, Mini-Mental State Examination; MMT, manual muscle testing; MoCA, Montreal Cognitive Assessment; MPI, mean pixel intensity; MRC, Medical Research Council; MRI, magnetic resonance imaging; MVC, maximum voluntary contraction; MVICT, maximum isometric voluntary contraction testing; MUS, muscle ultrasound; ODI, oxygen desaturation index; oTUG, optimized TUG; PROMIS-PF, Patient-Reported Outcomes Measurement Information System Physical Function; QMT, quantitative muscle testing; RSA, relative surface area; RWS, reachable work space; SpO2, oxygen saturation; TUG, timed up and go.

**Table Appendix 7:**
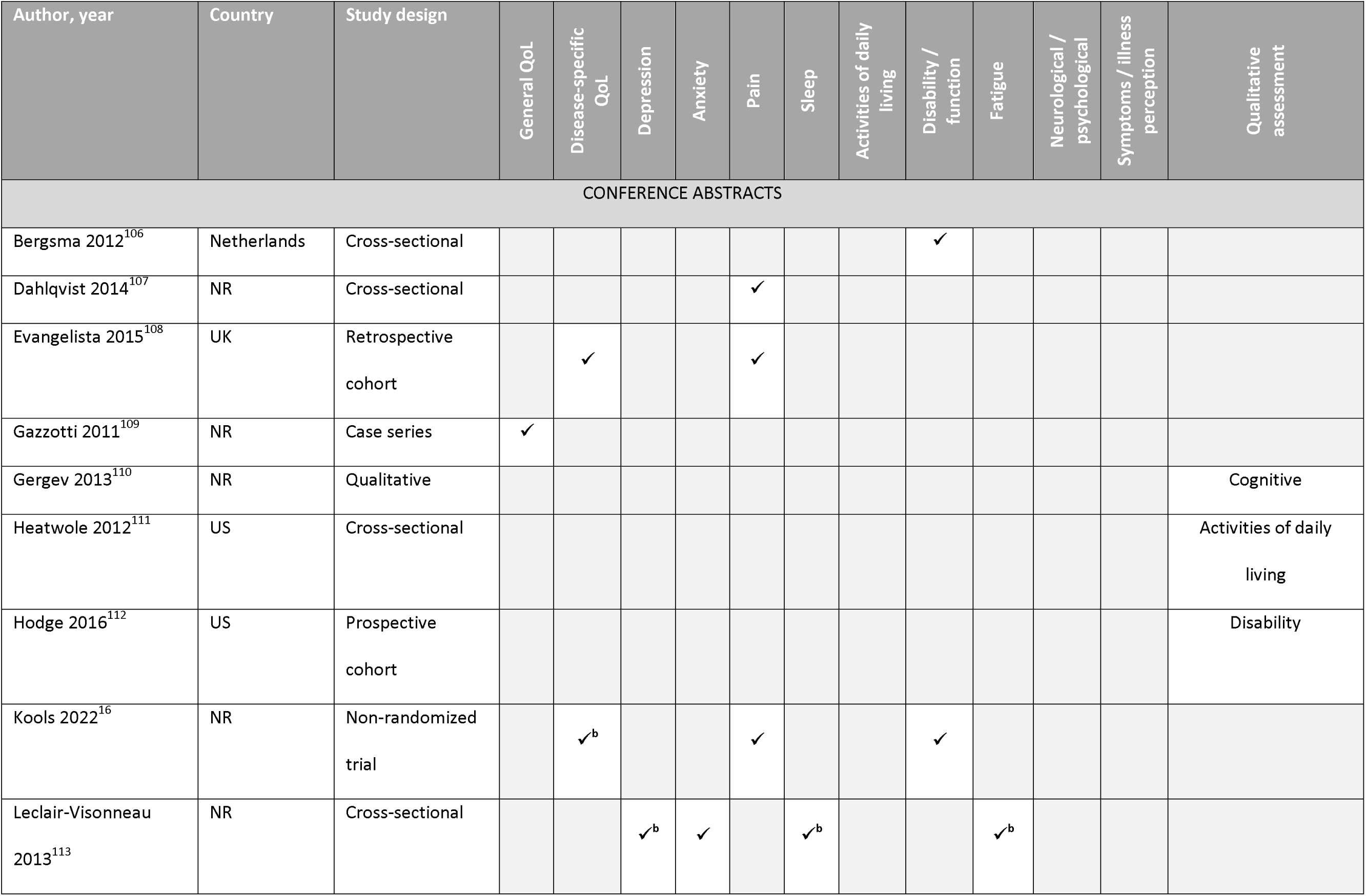
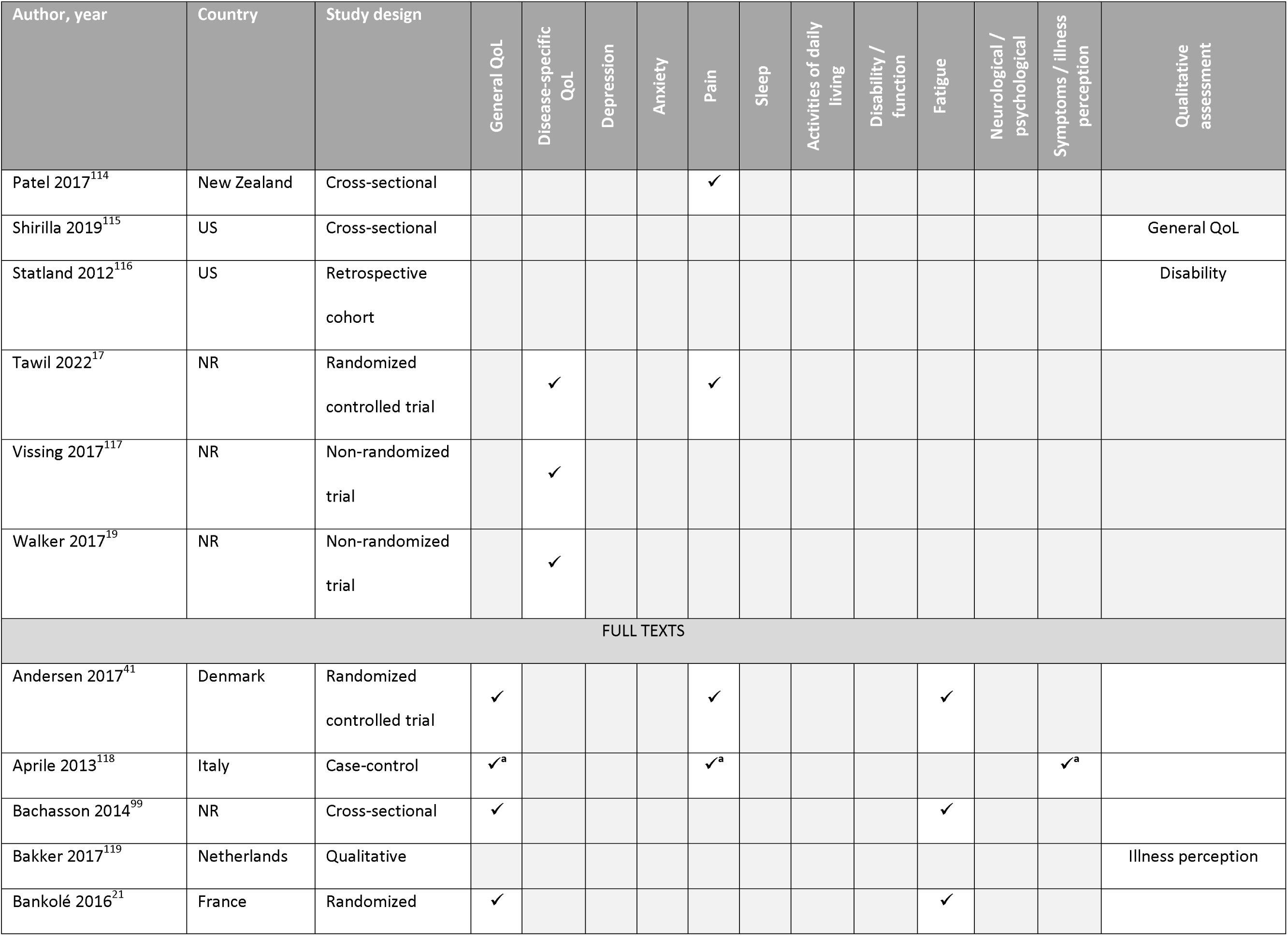
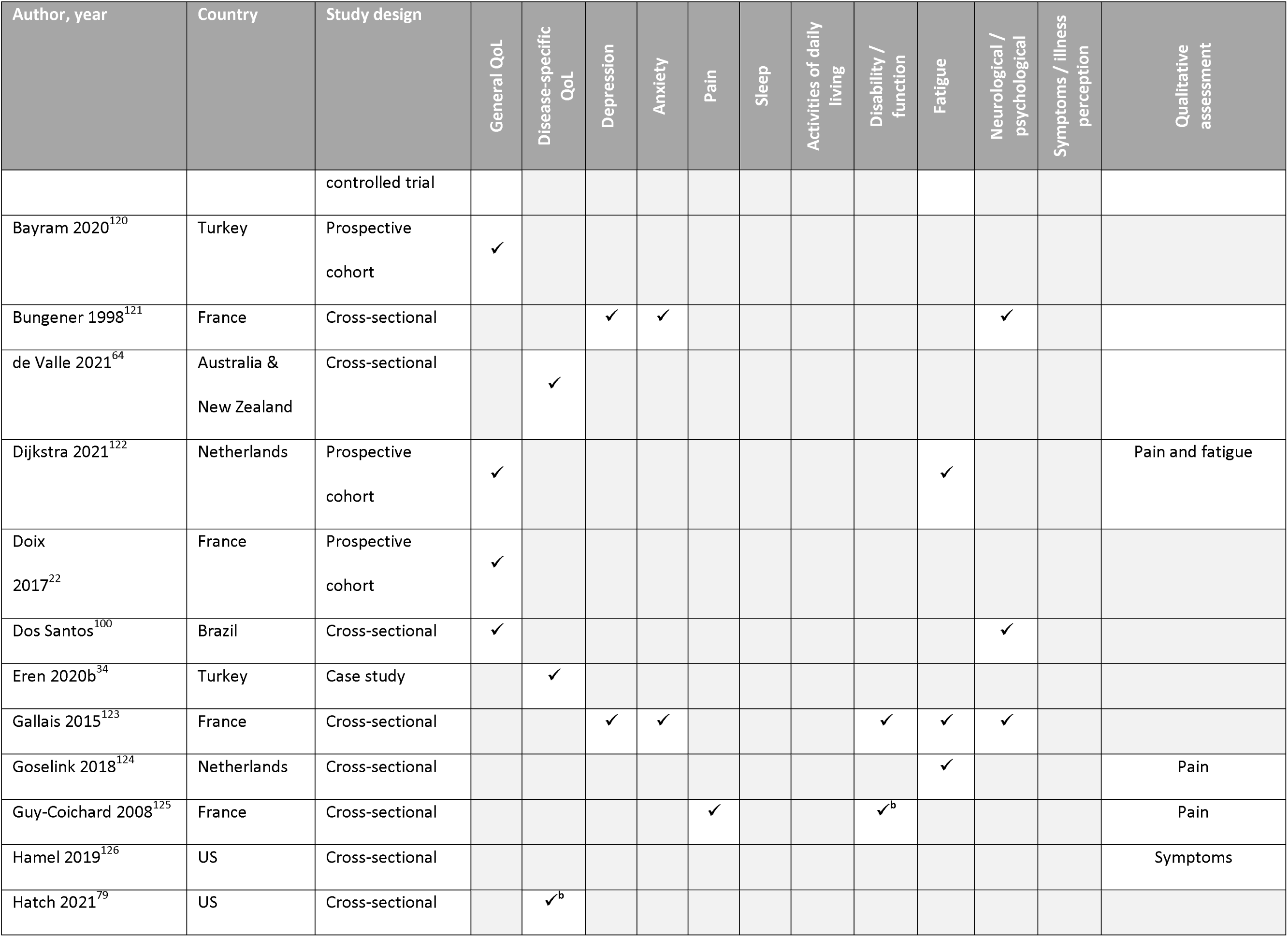
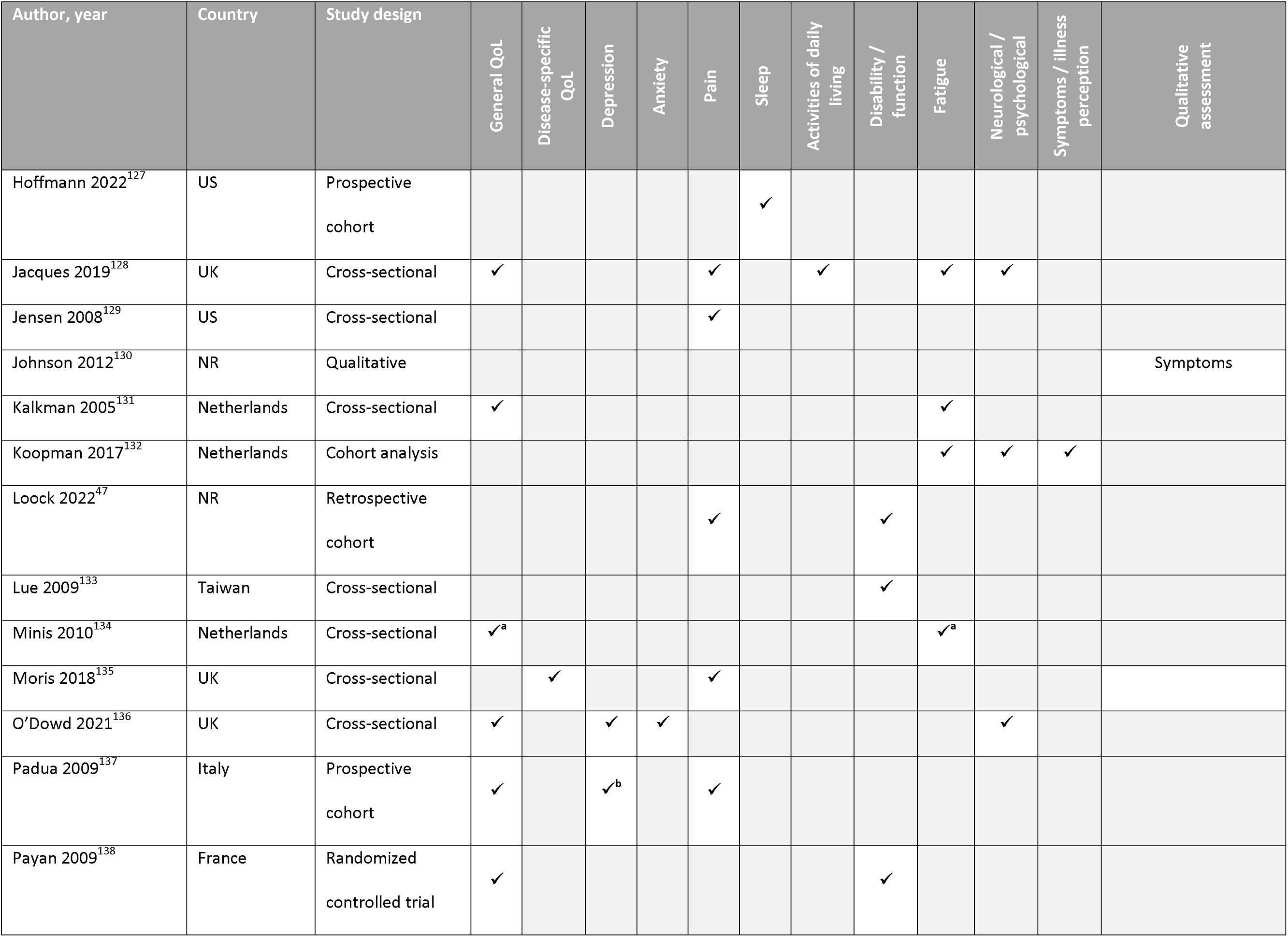
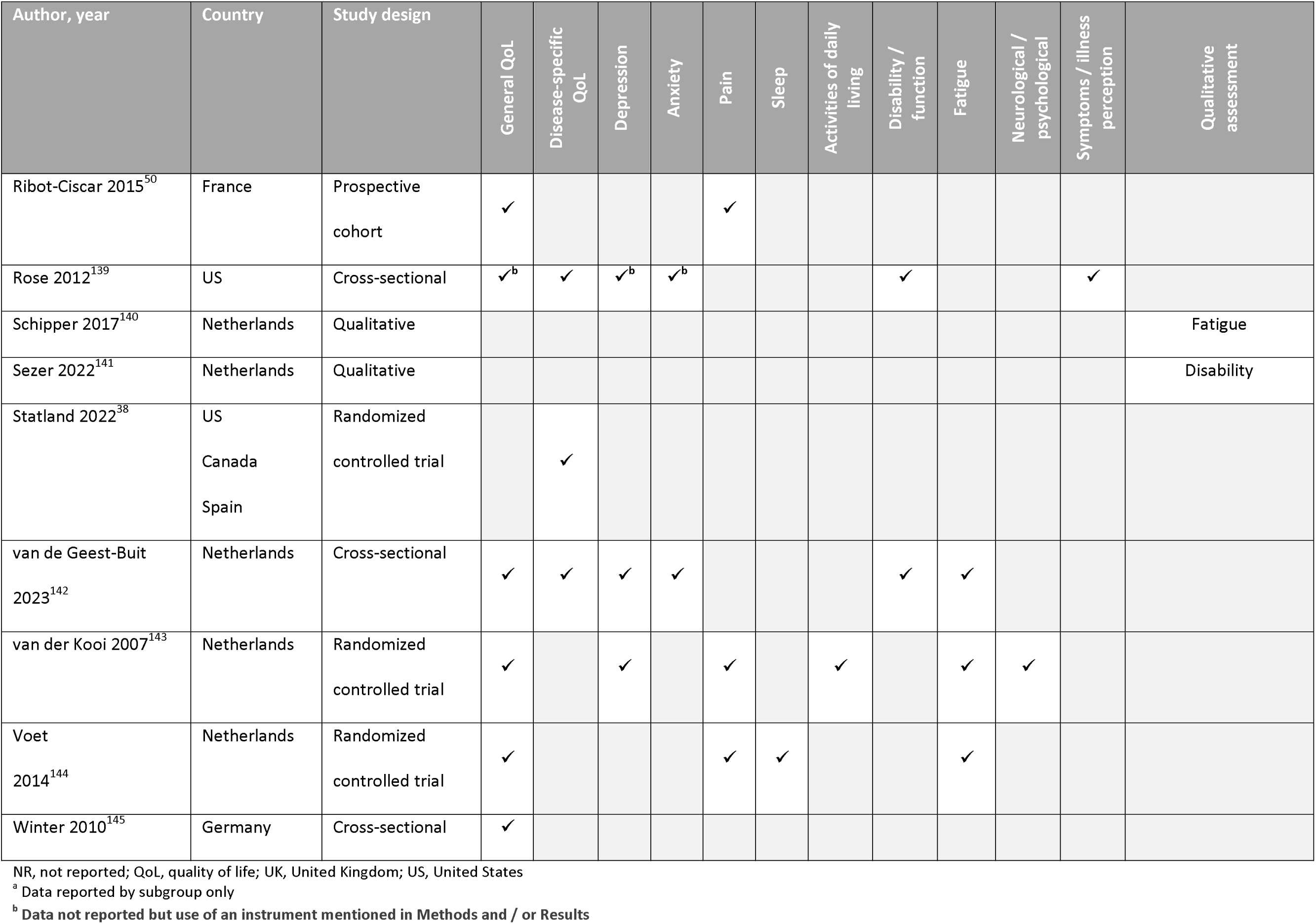
Summary of Identified Humanistic Burden Studies.

**Table Appendix 8:**
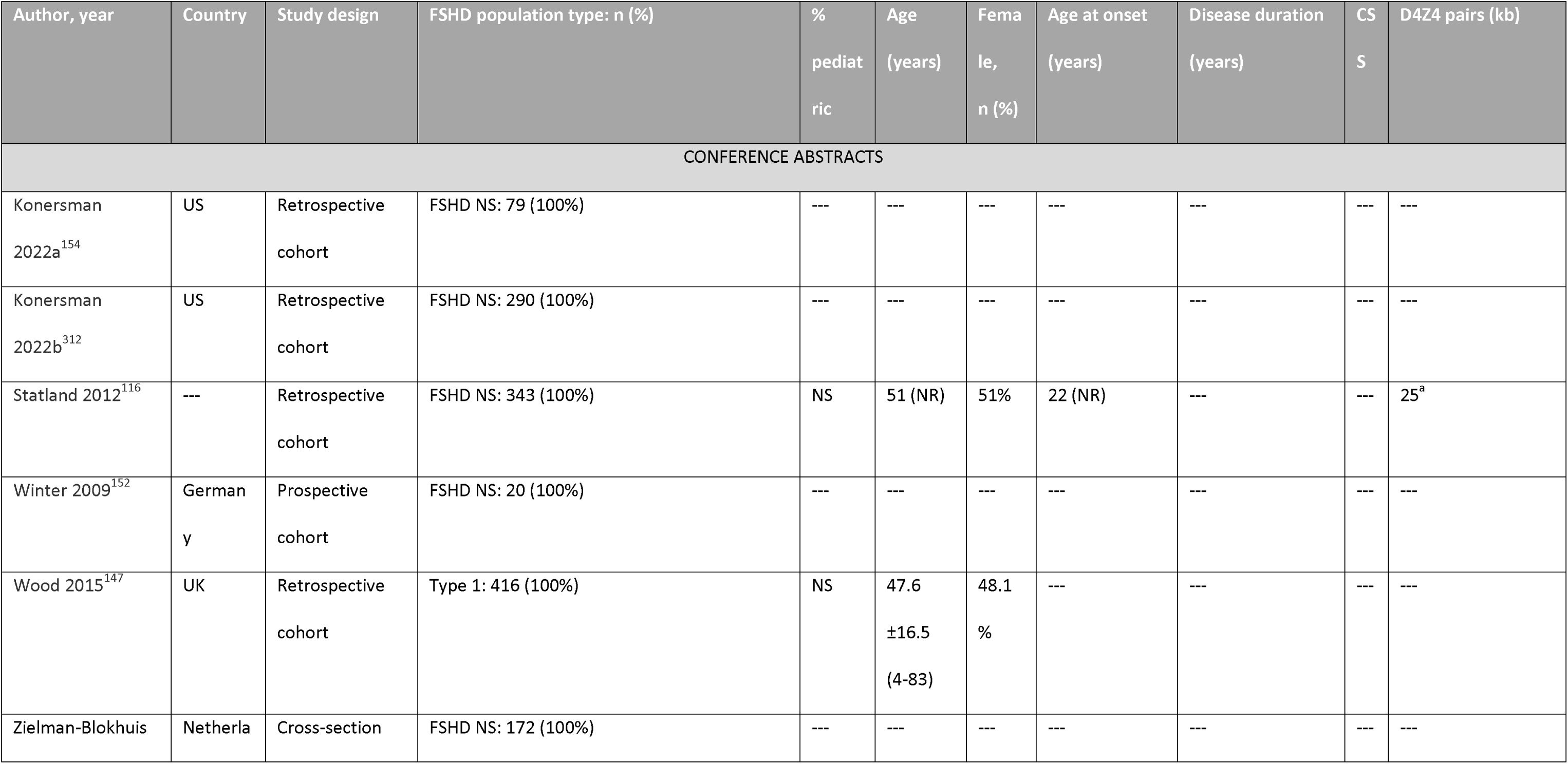
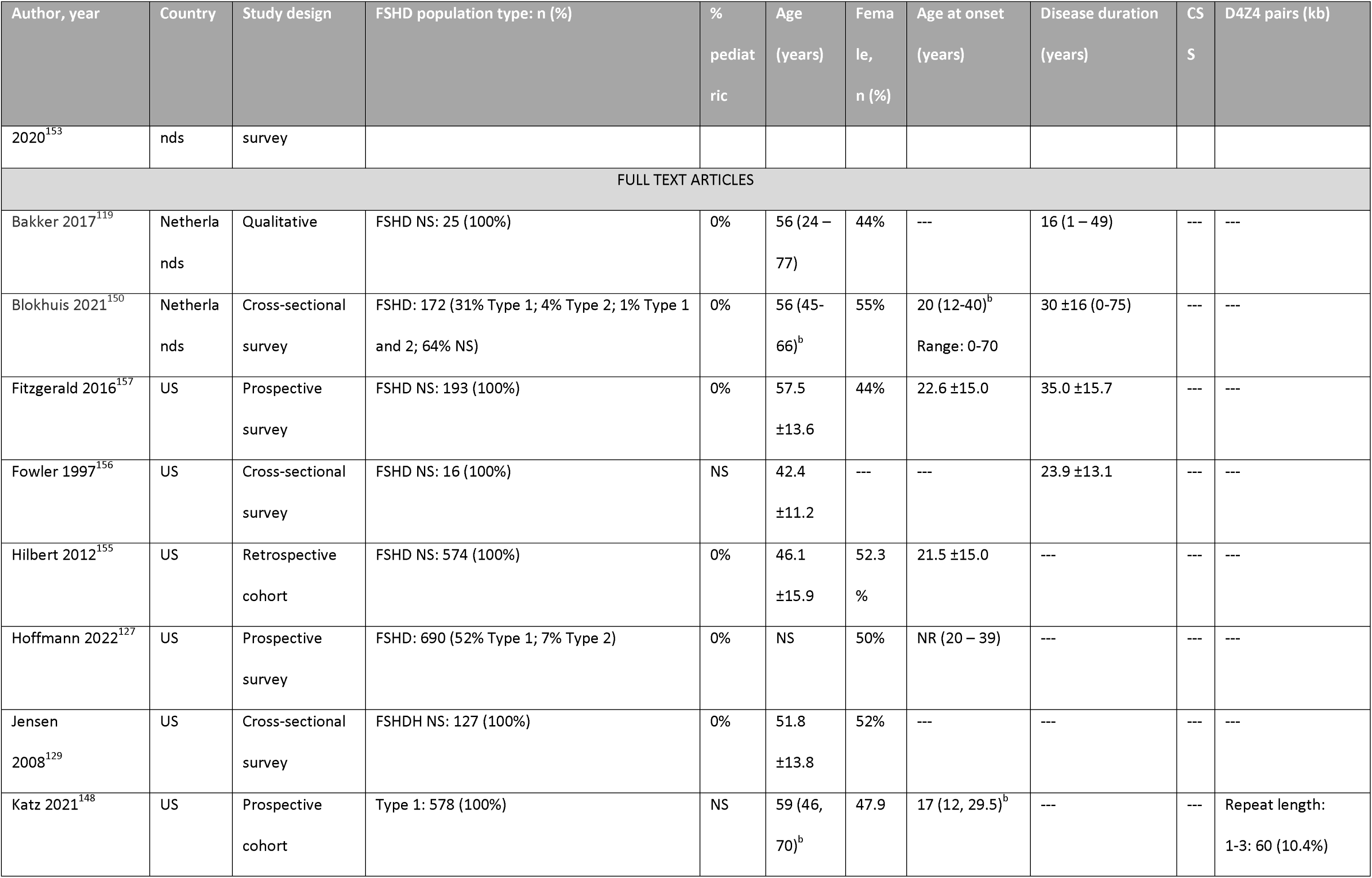
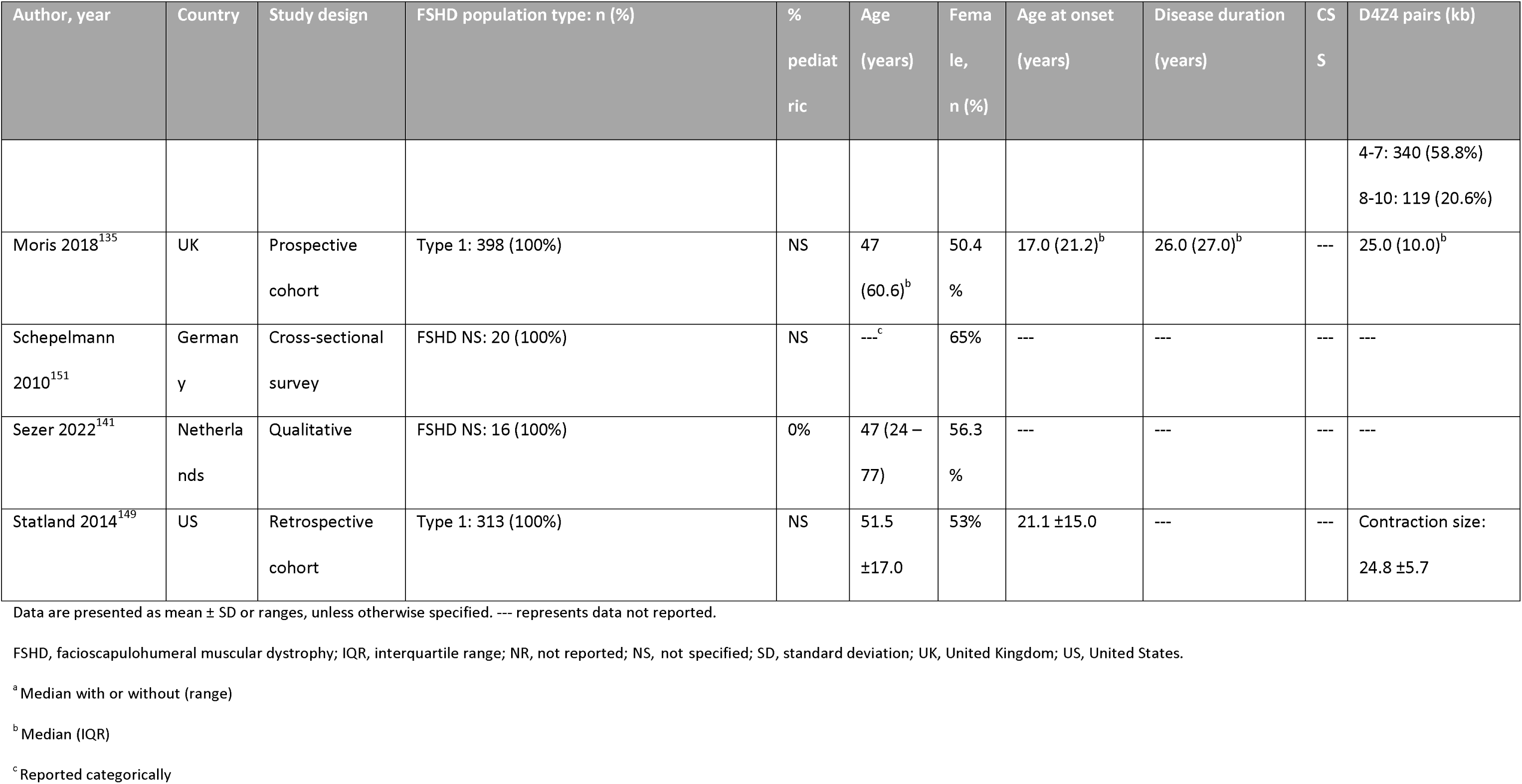
Summary of Identified Economic Burden Studies.

**Table Appendix 9:**
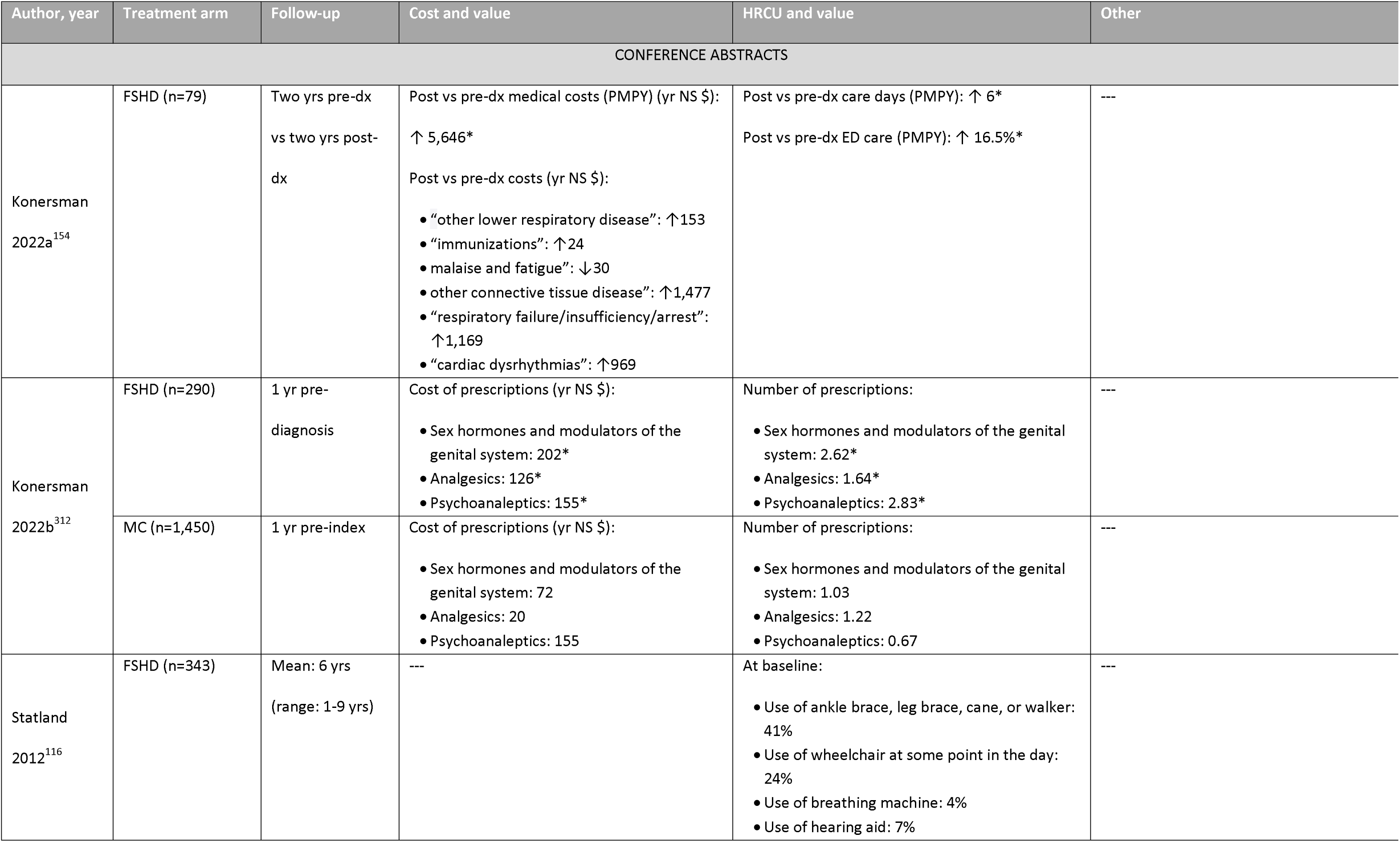
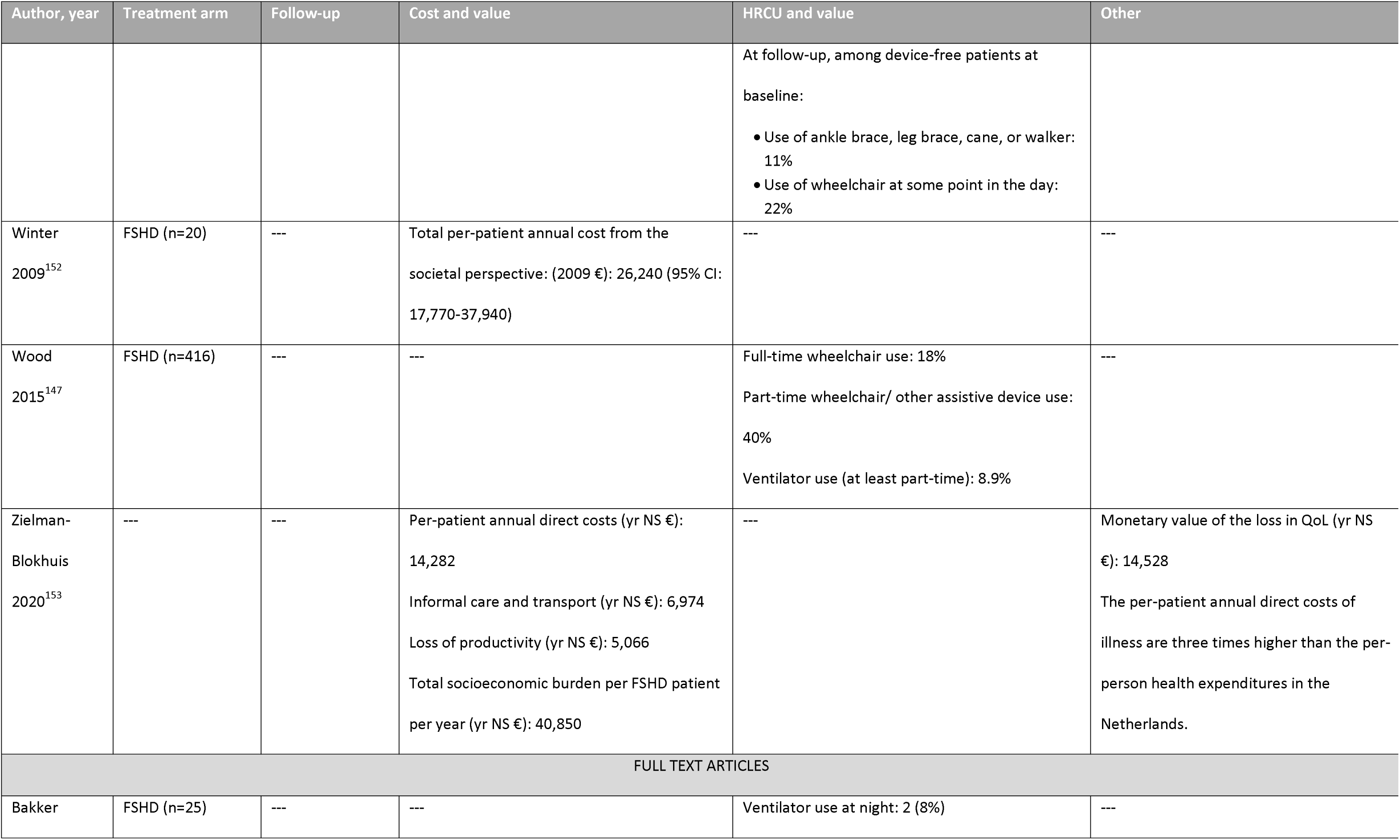
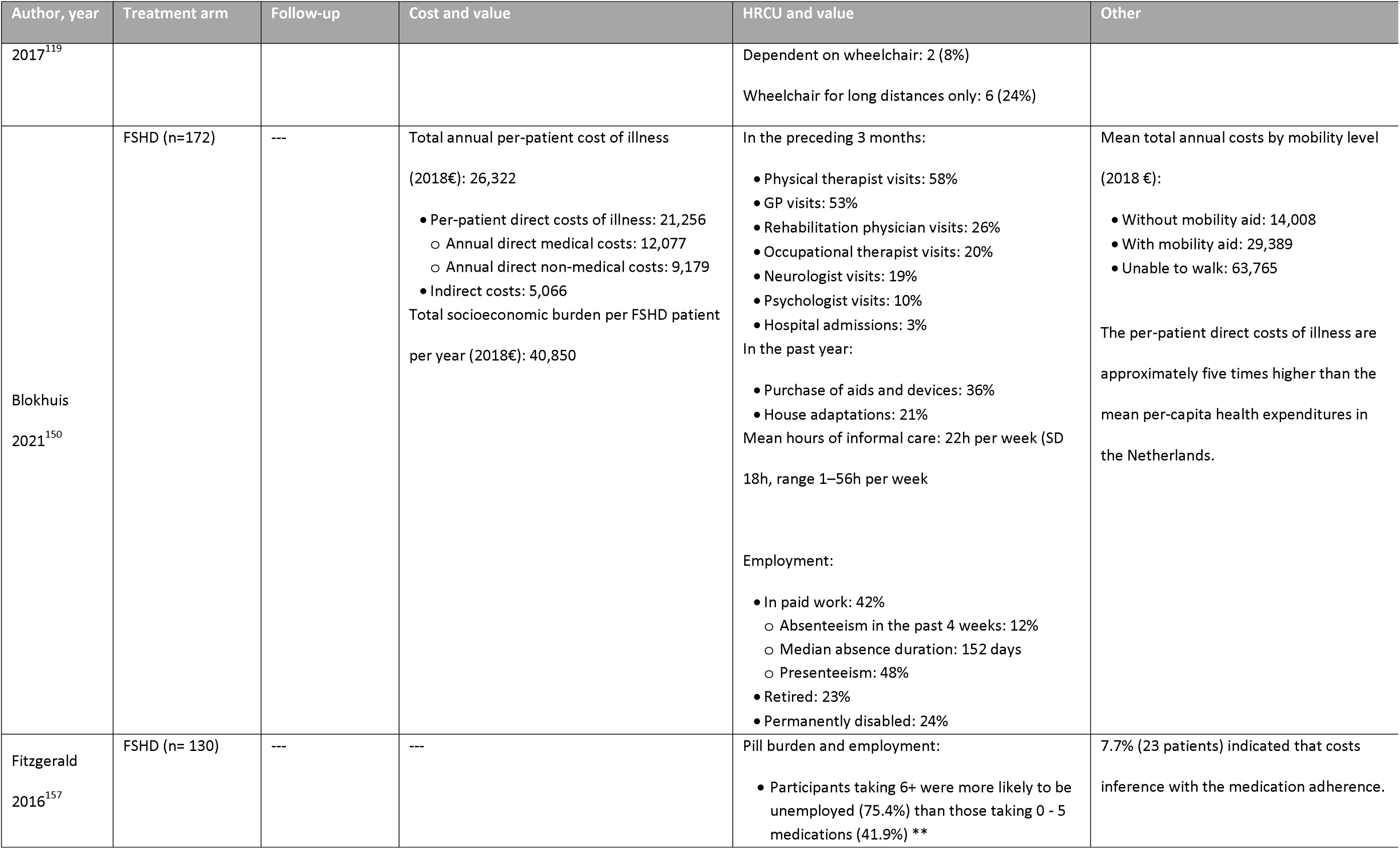
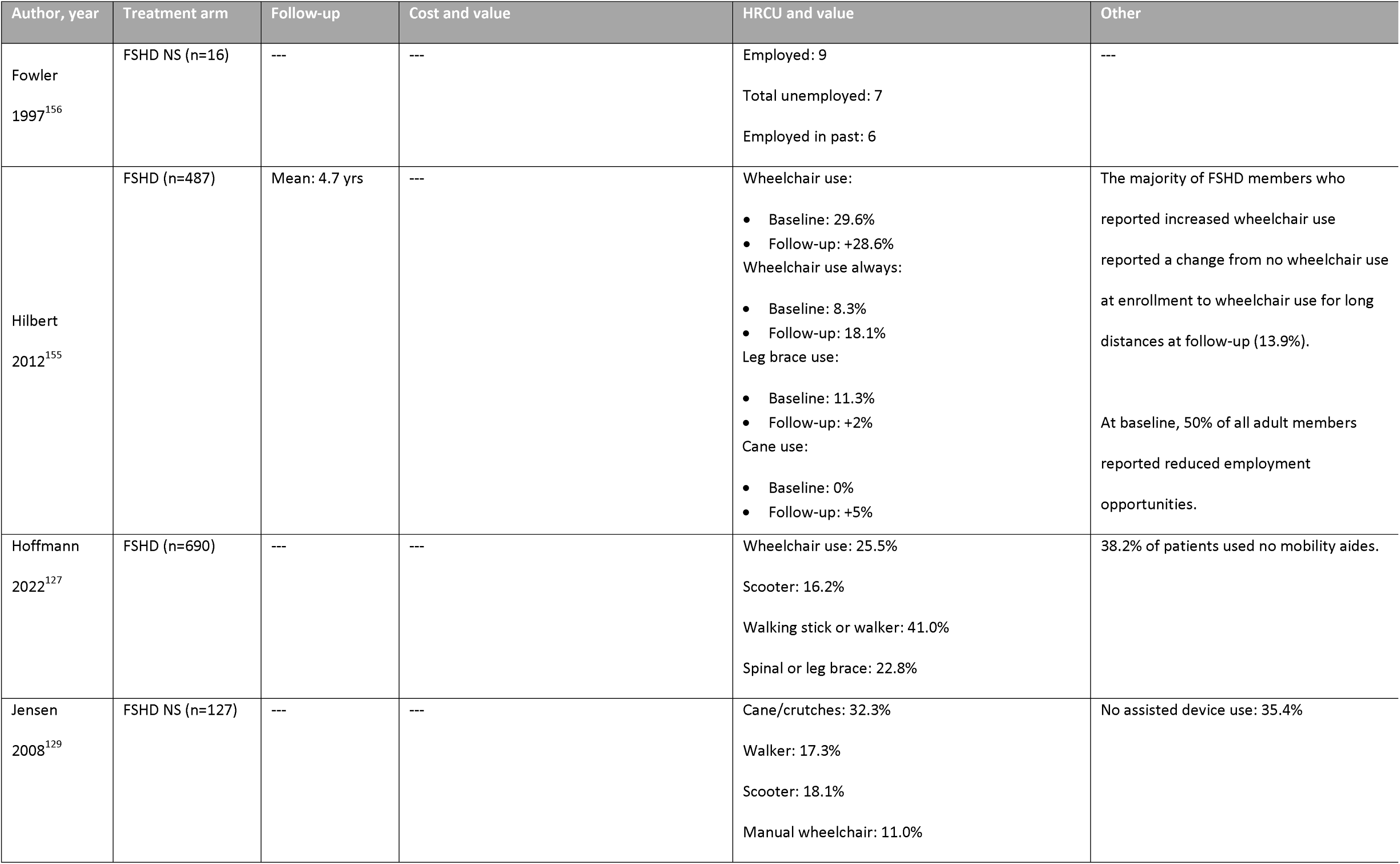
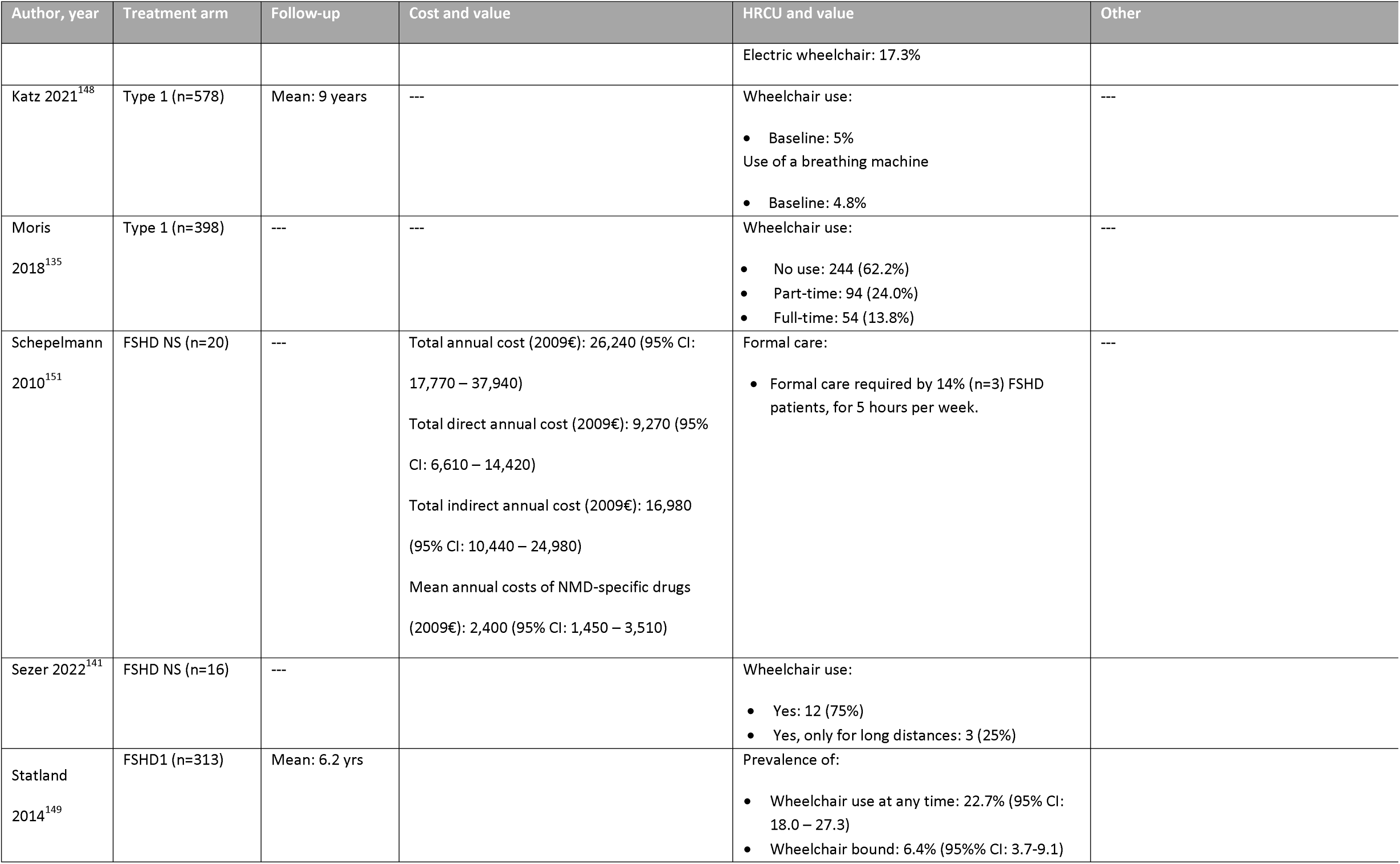
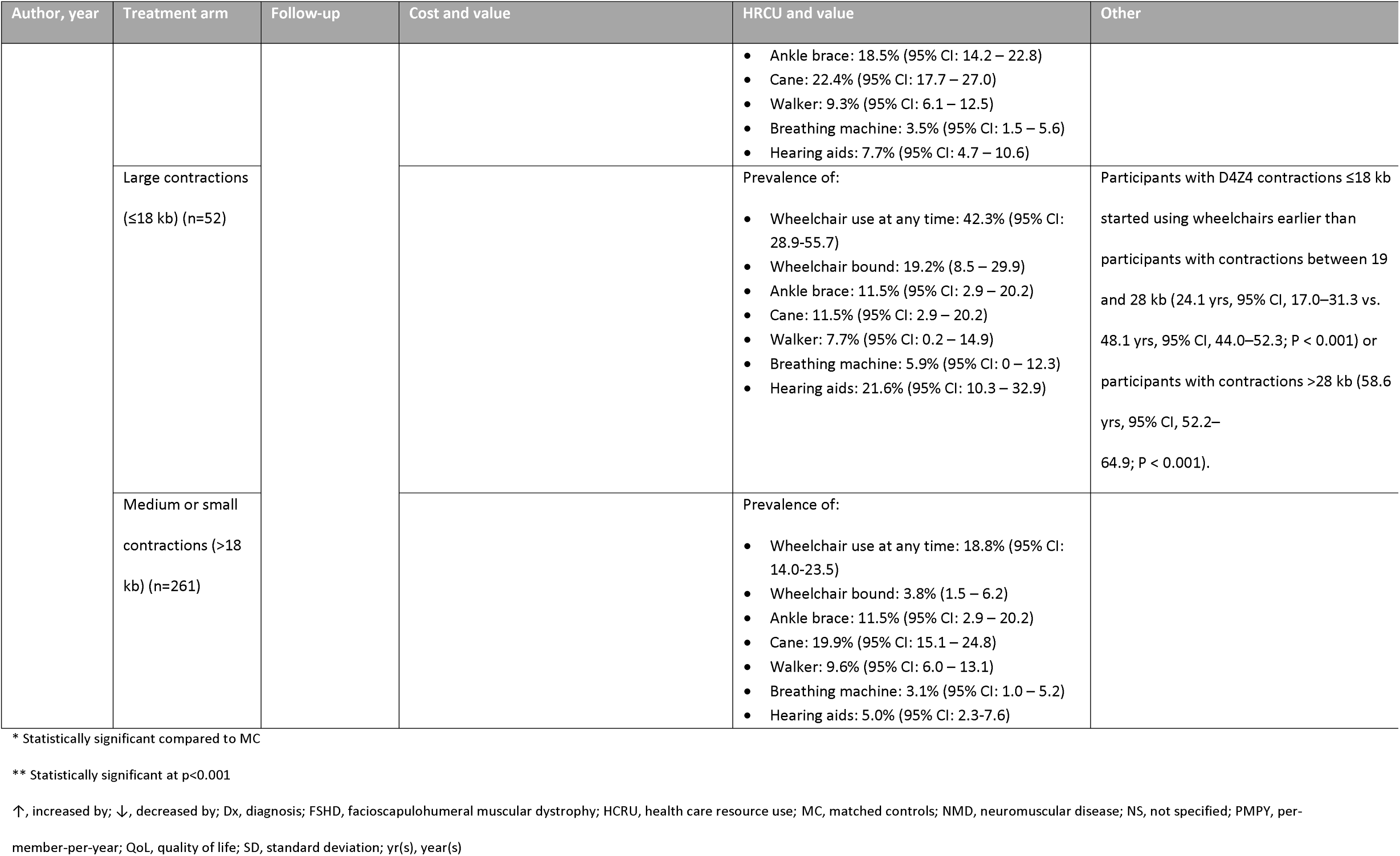
Outcomes of Identified Economic Burden Studies.

