## Supplemental C for "Landscape and Heterogeneity of Treatment Outcome Measures in Facioscapulohumeral Muscular Dystrophy: A Systematic Literature Review"

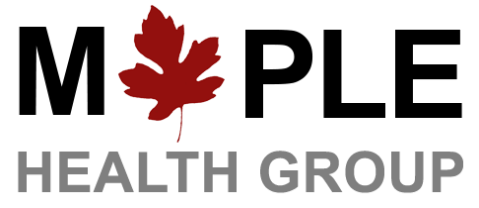

**Prepared for: Adi Eldar-Lissai**

**Author Contact Information**:

**Version 5**

January 25, 2024

**Systematic Literature Review of Facioscapulohumeral Muscular Dystrophy**

**REPORT**

**Abbreviations**

| **6MWT** | 6-minute walking test |
| --- | --- |
| **BDI** | Beck Depression Inventory |
| **BMI** | body mass index |
| **BPI** | Brief Pain Inventory |
| **BUE** | Brooke Upper Extremity |
| **CCEF** | Comprehensive Clinical Evaluation Form |
| **CI** | confidence interval |
| **CIS** | Checklist Individual Strength |
| **CMV** | contractile muscle volume |
| **CSS** | Clinical Severity Score |
| **CUE** | Capabilities of Upper Extremity |
| **DOA** | Daily Observed Activity |
| **DOF** | Daily Observed Fatigue |
| **DOP** | Daily Observed Pain |
| **EHD** | Depressive Mood Scale |
| **ESS** | Epworth Sleepiness Scale |
| **eTUG** | expanded Timed Up-and-Go |
| **FSHD** | facioscapulohumeral muscular dystrophy |
| **FSHD-COM** | FSHD Composite Outcome Measure |
| **FSHD-COM Peds** | FSHD Composite Outcome Measure pediatrics |
| **FSHD-HI** | FSHD health index |
| **FSHD-RODS** | FSHD Rasch-built overall disability scale |
| **FSS** | Fatigue Severity Scale |
| **GSE** | General Self-Efficacy Scale |
| **HADS** | Hospital Anxiety Depression Scale |
| **HAQ** | Health Assessment Questionnaire |
| **HDRS** | Hamilton Depression Rating Scale |
| **ICQ** | Illness Cognitions Questionnaire |
| **IMQ** | Illness Management Questionnaire |
| **INQoL** | Individualized Neuromuscular Quality of Life |
| **IPQ-R** | Illness Perception Questionnaire - Revised |
| **iTUG** | instrumented Timed Up-and-Go |
| **JBI** | Johanna Briggs Institute |
| **LARS** | Lille Apathy Rating Scale |
| **MADRS** | Montgomery-Asberg Depression Rating Scale |
| **MeSH** | Medical Subject Heading |
| **MFM** | Motor Function Measure |
| **MINI** | Mini-International Neuropsychiatric Interview |
| **MMT** | manual muscle testing |
| **MPQ** | McGill Pain Questionnaire |
| **MRC** | Medical Research Council |
| **MRI** | magnetic resonance imaging |
| **MVIC** | maximal voluntary isometric contraction |
| **NEADL** | Nottingham Extended Activities of Daily Living |
| **NeuroQoL** | Quality of Life in Neurological Disease |
| **NGS** | next generations sequencing |
| **NHP** | Nottingham Health Profile |
| **NMES** | neuromuscular electrical stimulation |
| **NHP** | Nottingham Health Profile |
| **NOS** | Newcastle Ottawa Scale |
| **Peds QL NMM** | Pediatric Quality of Life Neuromuscular Module |
| **PCR** | polymerase chain reaction |
| **PFGE** | pulsed field gel electrophoresis |
| **PGIC** | Patient Global Impression Change |
| **PICOS** | Population, Intervention, Comparator, Outcome, Study design |
| **PRISMA** | Preferred Reporting Items for Systematic Literature Reviews and Meta-Analyses |
| **PRO** | patient-reported outcome |
| **PSQI** | Pittsburg Sleep Quality Index |
| **QoL** | quality of life |
| **RCT** | randomized controlled trial |
| **SAAS** | Social Appearance Anxiety Scale |
| **SD** | standard deviation |
| **SES** | Self-efficacy Scale |
| **SF-36** | Short Form 36 |
| **SIAS** | Social Interaction Anxiety Scale |
| **SIP** | Sickness Impact Profile |
| **SLR** | systematic literature review |
| **STAI** | State-Trait Anxiety Inventory |
| **TMT** | Trail Making Test |
| **TMV** | total muscle volume |
| **US** | United States |
| **VAS** | visual analogue scale |
| **VUE** | Vignos Lower Extremity |
| **WFS** | Walton Functional Scale |

### Executive Summary

Facioscapulohumeral muscular dystrophy (FSHD) is a rare genetic muscle disorder that manifests itself as a progressive weakening and loss of skeletal muscles. FSHD is caused by an aberrant expression of DUX4 in skeletal muscle, resulting in the inappropriate presence of DUX4 protein.^1^ At disease onset, muscles of the face (facio), shoulder girdle (scapulo), and upper arms (humeral) are primarily affected. As the disease progresses, muscle weakness and loss spread to the arms, trunk and lower body.^2^ FSHD leads to significant physical limitations, including the facial weakness and difficulty using arms. While predominantly affecting upper extremities, approximately 20% of affected individuals eventually require a wheelchair.^3^

This systematic literature review (SLR) was conducted to review and synthesize the available evidence on the burden of and unmet need in FSHD. A comprehensive search was conducted in Embase (via Embase.com), MEDLINE (via Pubmed.gov) and Cochrane (via cochranelibrary.com) databases on October 11, 2022. In addition, a manual search of the following three congresses’ databases was performed: Muscular Dystrophy Association (MDA), Muscle Study Group (MSG), and FSHD Society International Research Congress (FSHD IRC) for 2019, 2020 and 2021 (as available). A total of 2,211 and 754 non-duplicate full texts and conference abstracts were identified. Of these,78 conference abstracts and 152 full text articles were included. The included studies reported on seven topics: “Treatment Outcomes and Clinical Trials”, “Disease Outcome Measures and Validation”, “Humanistic Burden and Patient-Reported Outcomes (PROs)”, “Economic Burden”, “Disease Classification”, “Diagnosis”, and “Guidelines”.

The evidence base for the topic “Treatment Outcomes and Clinical Trials” (n=50 full texts and n=14 conference abstracts) was the most reported topic but data were very heterogenous. Only full text studies reporting data on priority outcomes (motor function, muscle strength and endurance or muscle-related) proceeded to data extraction, resulting in a total of 33 studies. Among these 33 studies, four intervention types were identified: pharmacological (n=11), physiological (n=7), surgical (n=13) and other (n=3). Among pharmacological intervention studies, two studies reported significantly different between the pharmacological and placebo groups in two clinical trials (significant changes from baseline among training group and a third clinical trial reported significant changes from baseline among FSHD patients receiving the pharmacological intervention. Physiological interventions resulted in significant changes from baseline in two trials, though, in one study the change was observed in the control group and not the intervention group. Two observational studies reported significant changes from baseline through follow-up for patients receiving neuromuscular electrical stimulation. Of the studies included for surgical interventions, all but one study (n=12) reported on outcomes following scapulothoracic arthrodesis; the remaining study reported on scapuloplexy. Other interventions included herbal medicine, acupuncture and vibratory proprioceptive assistance.

The topic “Disease Outcome Measures and Validation” was the second-most reported topic (n=28 full texts and n=27 conference abstracts). The measurement property most often reported was construct validity, as measured by correlation analysis (n=22 full text and n=21 conference abstracts). Other measures of reliability and/or validity included Rasch analysis (n=3), factor analysis (n=2), reliability (n=22), responsiveness (n=7) and internal consistency (n=2). Correlation analyses were reporting for both instruments and for the validation of outcome assessments and were performed on muscle strength, function, clinical severity, muscle composition, mobility, lung function, patient-reported outcomes (PROs), and duration of disease. The FSHD-COM was the most assessed PRO instrument.

A total 53 studies (n=38 full text articles and n=15 conference abstracts) were identified for the topic “Humanistic Burden and PROs”. Within this topic, the most reported domains using a PRO instrument were general quality of life (QoL; n=21), followed by fatigue (n=14) then pain (n=12), with the SF-36 being the most reported PRO instrument. Other domains using an instrument that were reported in the literature included disease-specific QoL (n=11), depression (n=7), anxiety (n=7), sleep (n=3), activities of daily living (n=2), disability / functional status (n=9), neurological/psychological (n=7) and symptoms/illness perception (n=3). In addition to the identified studies that reported data using a PRO instrument, eight studies were identified that qualitatively reported results on the humanistic burden among FSHD patients.

Limited evidence was identified for the topic “Economic Burden” (n=12 full texts and n=6 conference abstracts) among people living with FSHD. Of the identified articles, the evidence suggests that FSHD is associated with substantial economic burden for patients, with one study reporting per-patient annual direct costs of illness to be five times higher than mean per capital health expenditure in the Netherlands. Healthcare resource use was reported in 16 studies, with 12 studies reporting the proportion of patients using ankle braces, leg braces or wheelchairs (part- or full-time).

“Disease Classification” (n=28 full texts and n=15 conference abstracts) studies indicated that there currently is a lack of consensus in classifying FSHD. Similarly, studies reporting on “Diagnosis” (n=17 full text articles and n=7 conference abstracts) used a variety of diagnostic techniques focusing on clinical assessments and genetic testing. Only three older studies (published in or before 2015) were identified reporting on “Guidelines” on FSHD management. The findings of the “Diagnosis” and “Guidelines” studies suggested that there is no established diagnostic pathway for patients suspected to have FSHD, with no formal up-to-date clinical recommendations on FSHD management.

Overall, the findings of this SLR indicate that despite being the second-most prevalent form of muscular dystrophy, FSHD is not well-characterized in the literature. Multiple unique outcomes assessing disease progression were identified; consensus on both what and how to measure treatment benefit for FSHD patients is lacking. Efforts should be made to develop objective, feasible and accessible clinical assessment tools to reliably evaluate and monitor outcomes in FSHD patients, along with detecting small changes over time to determine progression. While multiple studies reported on humanistic burden, there was heterogeneity in the instruments used to assess the multidimensional detriments of patients with FSHD, including anxiety, depression and pain. Multiple studies reported multiple measurement properties, with various measures assessed. Few studies were identified on diagnosis and disease management guidelines, highlighting the variability in time to diagnosis and treatment patterns experienced by patients. Given the breadth of outcomes reported, it is difficult to quantify the true disease burden experienced by FSHD patients, their families, and the healthcare system. Efforts should be made to harmonize the assessment of the debilitating and progressive nature of FSHD, as research on treatment options continue.

### 1 Background

Facioscapulohumeral muscular dystrophy (FSHD) is a rare genetic muscle disorder that manifests itself as a progressive weakening and loss of skeletal muscles.^1^ FSHD is caused by aberrant expression of DUX4 in skeletal muscle, resulting in the inappropriate presence of *DUX4* protein. *DUX4*-driven gene expression is usually limited to the early embryonic development stage, after which the DUX4 gene is silenced. In patients with FSHD, the DUX4 gene is unsilenced as a result of a genetic mutation. The aberrant DUX4 expression leads to death of muscle and its replacement by fat, resulting in skeletal muscle weakness and progressive disability.^2^ At disease onset, muscles of the face (facio), shoulder girdle (scapulo), and upper arms (humeral) are primarily affected. As the disease progresses, muscle weakness and loss spread to the arms, trunk and lower body.^2^ FSHD leads to significant physical limitations, including facial weakness and difficulty using arms. While predominantly affecting upper extremities, approximately 20% of affected individuals eventually require a wheelchair.^3^

The prevalence of FSHD in the United States (US) is between 16,000 to 38,000 individuals.^4^ It is estimated that two-thirds of cases are inherited, and the remaining one-third are caused by sporadic mutations that are not due to family history of the disease. FSHD equally affects both genders and all ethnic groups.^2^ Currently, there are no approved therapies for FSHD. Disease management focuses on supportive treatments with physical therapy being a key part.^3^

Losmapimod, a suppressor of *DUX4* expression, is currently being investigated by FULCRUM Therapeutics for the treatment of FSHD. FULCRUM’s phase 2b study, ReDUX4, demonstrated clinically relevant and nominally statistically significant benefits versus placebo on multiple measures of structural and functional FSHD disease progression and patient reported outcomes at 48 weeks.^5^

The primary objective of this project was to systematically review and synthesize evidence to gain a better understanding of the available published evidence on the burden of and unmet need in FSHD. Specifically, the questions were:

- What is the clinical, humanistic, and economic burden of FSHD?
- What is the available evidence on the real-world effectiveness of currently available interventions?
- What measures of efficacy and safety are used to assess emerging therapies in FSHD?
- Which study endpoints in FSHD have been reliably validated as outcome measures?

### 2 Methodology

#### 2.1 Protocol and registration

The protocol for this systematic literature review (SLR) was designed by Maple Health Group and approved by FULCRUM and expert neurologic clinician familiar with FSHD. The protocol was not registered in a publicly available database.

#### 2.2 Identification and Selection of Evidence

##### 2.2.1 Search strategy

The SLR was performed in accordance with the methodological principles of conduct as detailed in the University of York Centre for Reviews and Dissemination’s “Guidance for Undertaking Reviews in Health Care”,^6^ and reported per the Preferred Reporting Items for Systematic Literature Reviews and Meta-Analyses (PRISMA) statement (2020).^7^

##### 2.2.2 Data sources

A comprehensive search was done in Embase (via Embase.com), MEDLINE (via Pubmed.gov) and Cochrane (via cochranelibrary.com) databases. The search strategy was assembled by researching appropriate terms (both subject headings, i.e., Medical Subject Headings [MeSH] and Emtree terms, and text words as appropriate) for inclusion in the literature search strategy. The strategy was validated by comparing the search results to a sample set of published studies and review papers that would be expected to be identified for the project.

The search strategy is presented in **Appendix A**. Given the rare disease status of FSHD and the relatively small number of expected hits, no time limits were applied to either the search of full text publications or conference abstracts indexed in Embase.

In addition to the search of full text publications, FULCRUM identified the following congresses of interest:

- World Muscle Society (WMS)
- Muscular Dystrophy Association (MDA)
- Muscle Study Group (MSG)
- FSHD Society International Research Congress (FSHD IRC)
- International Congress on Neuromuscular Diseases (ICNMD)
- American Academy of Neurology (AAN)
- American Managed Care Pharmacy (AMCP)
- International Society for Pharmacoeconomic Research (ISPOR)

As WMS, ICNMD, AAN, and AMCP are indexed in Embase they did not need to be hand-searched. A manual search of the remaining conference proceedings was conducted using free-text terms (**Table 1**), for the years available as of the search date.

Table 1: Conference proceedings hand search

| Conference | Journal | Year(s) |
| --- | --- | --- |
| Muscular Dystrophy Association (MDA) | 2022: <https://www.mdaconference.org/abstracts/2022-abstract-library/>  2021: <https://www.mdaconference.org/abstracts/2021-abstract-library/> | 2021, 2022^§^ |
| Muscle Study Group (MSG) | RRNMF Neuromuscular journal (2020 onwards)  Muscle and Nerve (2019) | 2019, 2020, 2021  2022 abstracts not yet available* |
| FSHD Society International Research Congress (FSHD IRC) | available at: https://www.fshdsociety.org/intl-research-congress/ | 2019, 2020, 2021  2022 abstracts not yet available* |

FSHD, facioscapulohumeral muscular dystrophy

^§^Please note that abstracts published before 2021 are not indexed online.

*As of September 15, 2022.

##### 2.2.3 Study selection

Citations identified in the database searches were first downloaded to an EndNote library. Duplicate citations were then identified and removed. Citations were then be exported to DistillerSR®, a literature review screening platform. Each citation was evaluated against the predetermined Population, Intervention, Comparator, Outcome, Study design (PICOS) criteria (**Table 2**) at both the title/abstract and full text screening phases.

Table 2: PICOS Criteria

| Criteria | Inclusion | Exclusion |
| --- | --- | --- |
| Population | - Adults, children/adolescents with FSHD, type 1 and 2 | - Patients with conditions/muscular dystrophies other than FSHD - Mixed populations not presenting stratified FSHD data |
| Intervention | - No criteria | - No criteria |
| Comparator | - No criteria | - No criteria |
| Outcomes | - Studies containing efficacy, safety, humanistic, or economic outcomes assessments - Studies describing the validity of outcomes assessments in FSHD - Studies describing humanistic burden - Studies describing economic burden - Studies describing disease diagnosis - Studies describing disease classification (i.e., by severity) | - Outcomes other than those described in the inclusion criteria (e.g., pathoetiology, epidemiology only) |
| Study Design | Primary studies including:   - Clinical trials of investigational therapies or interventions (i.e., physical therapy) - Observational studies/real-world evidence studies and/or outcomes in FSHD (e.g., prospective cohort and cross-sectional studies, retrospective analyses of real-world databases, disease registries) - Endpoint validation studies - Systematic literature reviews of the burden of illness studies/treatment outcomes - Case reports and case studies - Trial registers | - Pre-clinical/non-human studies - Reviews, commentaries, letters, notes, editorials |
| Limits | - Language: English | - Conference abstracts published before 01 January 2019 for hand searched conferences only |

FSHD, facioscapulohumeral muscular dystrophy; PICOS = Population, Intervention, Comparator, Outcome, Study design

###### 2.2.3.1 Title/Abstract Review

Each title and abstract was independently reviewed against the inclusion/exclusion criteria (**Table 2**) by two reviewers. In cases where it was not clear if the title/abstract met the inclusion criteria or if the abstract was not available for review, the citation was included for full text review to ensure that all potentially relevant studies were captured. Any disagreement regarding the eligibility of a given study between the two reviewers was discussed until a consensus was met; unresolved conflicts were decided by a third reviewer.

At the end of the title/abstract review phase, all identified studies were classified as either included or excluded.

###### 2.2.3.2 Full-Text Review

All studies included in the title/abstract phase of screening proceeded to full-text review. Each full publication was independently reviewed against the eligibility criteria (**Table 2**) by two reviewers. Studies not providing sufficient information to enable evaluation against the eligibility criteria were excluded in order to ensure that only relevant publications were captured. As with title/abstract screening, any disagreement regarding the eligibility of a given study between the two reviewers was discussed until a consensus was met, with any unresolved conflicts decided by a third reviewer.

At the end of the full text review stage, all studies were classified as included or excluded and had their primary reason for exclusion captured and recorded as follows:

- Population did not include adults, children or adolescents with Type 1 or Type 2 FSHD
- Reported outcomes not relevant
- Study design not relevant (editorial, letter, commentary, notes, non-systematic review)

##### 2.2.4 Data extraction

Peer-reviewed full-text publications were used as the primary source of information for extraction over conference abstracts or trial registers identified for the same study for the same data reported (i.e., conference abstracts were listed as companions to the primary publication). In instances where multiple full text publications of the same study were identified, the publication presenting the most recent data for a given outcome was prioritized for data extraction (e.g., five-year follow-up to clinical trial, a follow-up to interim trial data). However, a conference abstract was prioritized over a full-text publication for extraction when the conference abstract presented follow-up or additional data to a previously peer-reviewed published study/trial. For cases in which more than one conference abstract was identified without a full-text publication, the most recently presented/published abstract was prioritized for extraction if both abstracts were presenting the same data.

Data extraction was performed in accordance with the guidelines from the University of York.^6^ Data extraction was performed by a single individual for each included study and was independently verified by a second individual once completed. Any discrepancies identified by the second individual was discussed by both individuals with unresolved conflicts decided by a third individual.

Data was extracted and presented by topic. For all articles, across all topics, the following elements were extracted: author, year, population (Type 1, Type 2, not specified or neuromuscular disease in general), age, proportion pediatric, proportion female, body mass index (BMI), age at onset, disease duration, clinical severity score (CSS; or other measure of severity of disease), and mean D4Z4 base pairs.

Relevant outcomes to be extracted, by topic, were discussed with both FULCRUM and the clinical expert. Data was extracted as available for each included study. Data were presented as mean ± standard deviation (SD) or mean (range / 95% confidence interval [CI]), unless otherwise specified. Where available, up to two decimal points were presented. For the topics “Disease Classification”, “Diagnosis” and “Guidelines”, given the heterogeneous nature of the studies, a narrative synthesis of key points was conducted.

##### 2.2.5 Risk of bias

Quality appraisal of included studies was conducted in accordance with the guidelines from the University of York.^6^ As no restriction was made on study design, multiple tools were used, as applicable. These tools included: the COSMIN tool for studies on reliability or measurement error of outcome measure instruments^8^, a checklist for cost-of-illness studies^9^; the SIGN checklist for cohort studies^10^; the Joanna Briggs Institute (JBI) checklist for cross-sectional studies^11^; the Newcastle Ottawa Scale (NOS) for case-control studies^12^; the Downs & Black checklist for randomized and non-randomized trials^13^; the JBI checklist for case series^14^; the JBI checklist for case reports^11^; and the JBI checklist for qualitative research^15^.

Quality assessment was performed by a single individual for each included full text study.

### 3 Results

#### 3.1 Study selection

The electronic database searches, in addition to the hand searching of the conference websites, yielded 2,211 full text articles and 754 conference abstracts after de-duplication (**Figure 1**). A total of 1,990 full texts and 653 conference abstracts were excluded at the title and abstract review stage. Following study mapping, 23 conference abstracts were excluded as a full text publication was identified for the same study. Of the full text articles, 68 were excluded during the full text review. Thus, 78 conference abstracts and 152 full text articles (of which 13 were registers of clinical trials) were included.

Figure 1. PRISMA flow diagram

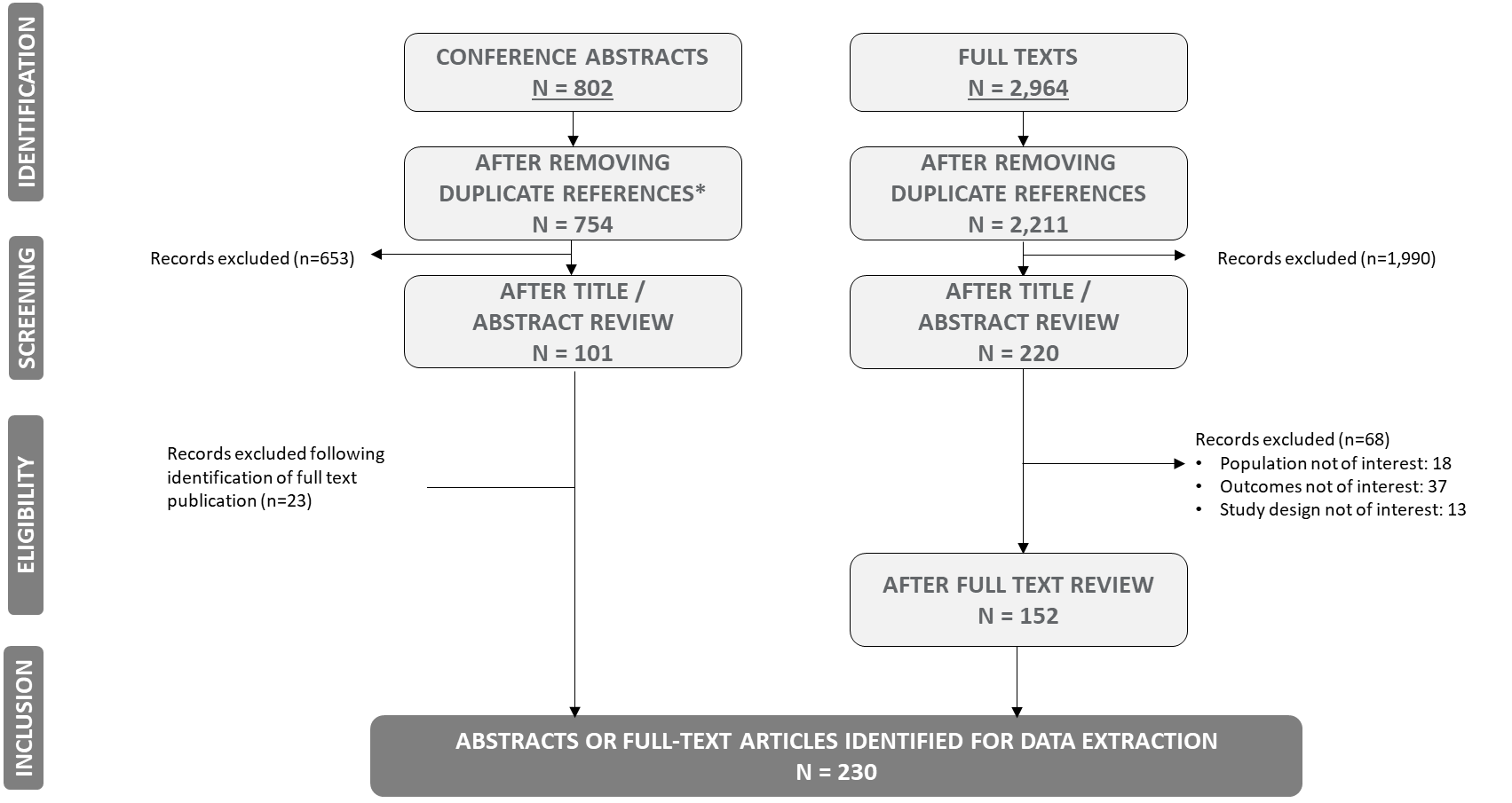

Abstracts or full text articles that were included for data summary and/or extraction (n=230) were categorized across seven broad topics (**Figure 2**). When considering all identified studies, the most commonly-reported topic was “Treatment Outcomes and Clinical Trials” (total n=64), followed by “Disease Outcome Measures and Validation” (total n=55). When considering conference abstracts only, the most reported topic was “Disease Outcome Measures and Validation” (n=27); for full text articles, the most commonly-reported topic was “Treatment Outcomes and Clinical Trials” (n=50). Of note, while the SLR identified 230 unique conference abstracts or full texts, a total of 30 studies reported on two topics.

Figure 2. Included studies by topic and publication type

PRO, patient-reported outcome

Number of studies

|  | Full texts | Conference abstracts |
| --- | --- | --- |
| Treatment Outcomes and Clinical Trials | 50 | 14 |
| Disease Outcome Measures and Validation | 28 | 27 |
| Humanistic Burden and PROs | 38 | 15 |
| Economic Burden | 12 | 6 |
| Disease Classification | 28 | 15 |
| Diagnosis | 17 | 7 |
| Guidelines | 3 | 0 |

#### 3.2 Treatment Outcomes and Clinical Trials

The topic “Treatment Outcomes and Clinical Trials” was the most reported topic in the SLR and included 37 full text articles, 14 conference abstracts, and 13 registers of clinical trials. An overview of the studies included for the topic “Treatment Outcomes and Clinical Trials” is provided in **Table 3,** with additional detail provided in **Table 9** in **Appendix 7.2**, noting that not all information was available for all studies. The most frequently identified study design among conference abstracts was clinical trials (n=10), followed by observational studies (n=3) and a single case study. Among full text articles, the most common study design was observational studies (n=16), followed by clinical trials reporting results (n=13), case studies (n=8); of note, there was one qualitative study.

Among all full text and conference abstracts, nine studies exclusively assessed patients with FSHD Type 1^16-24^ with sample sizes ranging from 1^23^ to 80^17^; while the remaining did not specify the FSHD type. Only 6 studies included pediatric patients^23-28^, of which 3 studies exclusively evaluated pediatric patients^23-25^; of note, both Li *et al.* and Xiao *et al.* were case studies with a total of three pediatric patients included^23,24^. Of the registers of clinical trials , two were noted to being conducted in Type 1 FSHD patients^29,30^; none were reported to include pediatric patients. The efficacy and effectiveness of different interventions were heterogeneous across the included studies.

Table 3. Overview studies proceeding to data extraction for “Treatment Outcomes and Clinical Trials”

|  | **Conference abstract** | **Full text** | **Randomized or non-randomized trial** | **Observational** | **Other^b^** |
| --- | --- | --- | --- | --- | --- |
| **Pharmacological** | 5 | 12^a^ | 14^a^ | 1 | 2 |
| **Physiological** | 7 | 9^a^ | 9^a^ | 6 | 1 |
| **Surgical** |  | 15 |  | 10 | 5 |
| **Other** | 2 | 3 | 1 | 2 | 2 |

^a^ One study assessed both a pharmacological and physiological intervention

^b^ Includes qualitative and case studies

Given the significant heterogeneity in studies, following a decision made by FULCRUM, only full text articles (either observational studies or clinical trials) reporting data on outcomes determined to be a priority (motor function, muscle strength and endurance or muscle-related outcomes) proceeded to data extraction. Though meeting the inclusion criteria of the SLR for the topic “Treatment Outcomes and Clinical Trials”, four studies did not report efficacy or effectiveness outcomes of interest, thus were not included for data extractions (Abel *et al.*^31^, Mellion *et al.*^32^, Andersen *et al.*^33^, and Eren *et al.*^34^).The full text articles proceeding to data extraction were categorized into one of four intervention types: pharmacological, physiological, surgical, or other. The study population, along with their baseline characteristics for each of these 33 full text studies that were extracted is presented in **Table 10** in **Appendix 7.2.** Of note, the clinical severity score, and the number D4Z4 pairs were not well-reported.

Of the 33 studies, a total of 11 studies reported on pharmacological interventions, which included both disease-modifying therapies as well as nutritional supplements; a description the interventions can be found in **Table 11** in **Appendix 7.2**. The majority of the studies were undertaken in an FSHD population with type unspecified, with the exception of Wagner *et al.*^35^, which was conducted in a general neuromuscular disease population (results were noted for the FSHD subgroup). One study explored both pharmacological and physiological training using a four group randomized controlled trial (RCT) study design and was included amongst the pharmacological interventions. Among the 7 studies included as physiological (exercise), interventions included strength training, high-intensity training, or aerobic conditioning. Exercise interventions were predominantly undertaken in Type 1 FSHD patients. A total of 13 studies were identified exploring surgical interventions (either scapulothoracic arthrodesis or scapulopexy) in FSHD patients (type not specified). Finally, 3 studies met the inclusion criteria of the SLR but did not fit into the previous three categories: one explored the impact of a Chinese herbal medicine in a single Type 1 FSHD patient, the second explored the impact of acupuncture in an FSHD patient, and the third assessed the intervention vibratory proprioceptive assistance among 8 FSHD patients. Efficacy and effectiveness outcomes are reported by intervention type and study design. A summary of the number of unique outcomes identified by publication type and intervention is presented in **Figure 3.**

Figure 3. Number of unique outcomes identified for “Treatment Outcomes and Clinical Trials”

Number of Outcomes

|  | Full Text Articles | Conference Abstracts | Registers of Trials |
| --- | --- | --- | --- |
| Pharmacological (n=11) | 44 | 23 | 23 |
| Physiological (n=7) | 36 | 19 | 12 |
| Surgical (n=13) | 33 | 0 | 0 |
| Other (n=3) | 16 | 3 | 0 |

##### 3.2.1 Pharmacological interventions

A total of 11 studies were included for data extraction for pharmacological studies. For these studies, 8 were either non-randomized or randomized trials, with the remaining either observational (n=1) or case studies (n=2). Few significantly different results (bolded in **Table 12** in **Appendix 7.2**) were observed across the studies. For FSHD patients on both low and high dose albuterol, significant changes from baseline were observed in grip strength after 52 weeks of treatment; no changes were observed in the placebo group.^36^ Among FSHD patients taking supplements, significant changes from baseline in muscle endurance and maximum voluntary isometric contraction (MVIC) were observed after 17 weeks of treatment; no changes were observed in the placebo group.^37^ Significant changes from baseline were also observed after 190 days of treatment with ACE-083 in the biceps brachii and tibialis anterior for MVIC, contractile muscle volume (CMV), total muscle volume (TMV) and muscle fat fraction (tibialis anterior only).^38^ One observational study explored the effect of 12 weeks of albuterol treatment and found that MVIC significantly increased among the 15 patients included.^39^ Two case studies were identified. The first reported treatment with denosumab in one FSHD patient and found that grip strength increased after treatment (follow-up not reported).^40^ The second case study found no improvement in muscle weakness following treatment with a supplement cocktail.^24^

##### 3.2.2 Physiological interventions

A total of 7 studies were included for physiological interventions and had data extracted (**Table 13** in **Appendix 7.2**). Of the four trials evaluating physiological interventions, significant differences were only observed in two. One trial found that the 6-minute walking test (6MWT) increased significantly compared to baseline among the control group after six weeks, while no change was observed among participants engaging in high-intensity training.^41^ The second trial observed significant changes from baseline in the 6MWT, muscle endurance, MVIC and enzyme activity after 24 weeks of training.^21^ Two of the three observational studies reported significant differences compared to baseline following exercise; both studies evaluated changes in FSHD patients following neuromuscular electro stimulation (NMES). Colson *et al.* found that the 6MWT, along with MVIC and manual muscle testing (MMT) increased significantly after 5 months of NMES.^42^ Similarly, Doix *et al.* found that both the 6MWT and the Motor Function Measure (MFM) increased significantly from baseline after 8 weeks of NMES training; these changes were only observed among patients with FSHD and not in the healthy volunteer group receiving NMES.^22^

##### 3.2.3 Surgical interventions

A total of 13 studies were included for surgical interventions: 10 were observational and 3 were case studies **(Table 14** in **Appendix 7.2**). All but one study reported on outcomes following scapulothoracic arthrodesis; the remaining study reported on scapuloplexy^26^. The majority of the studies reported on shoulder mobility, followed by descriptive reporting for activities of daily living. Of the 10 observational studies, 5 reported significant increases in either shoulder abduction, elevation or flexion following surgery^43-47^; no significant differences were reported in the Constant or Medical Research Council (MRC) score across the observational studies. The three case studies found no significant differences following shoulder surgery; however, two studies noted improvements in activities of daily living^27,48^.

##### 3.2.4 Other interventions

Three interventions were categorized as “other interventions” (**Table 15** in **Appendix 7.2**). One case study assessed shoulder abduction and muscle strength following herbal medicine in one patient over 20 days^23^. A second case study explored acupuncture over 1.5 years in 1 patient; no changes were observed in muscle strength in extremities though an improvement was noted in facial muscles^49^. An observational study explored vibratory proprioceptive assistance in eight patients over 36 days^50^; shoulder flexion, maximum isometric abduction force and the Constant score were noted to have significantly increased at follow-up.

#### 3.3 Disease Outcome Measures and Validation

The topic “Disease Outcome Measures and Validation” was the second-most reported topic in the SLR and included 28 full text articles and 27 conference abstracts. Among all studies, 12 studies (four conference abstracts^51-54^ and eight full text articles ^55-62^) exclusively assessed patients with FSHD Type 1; a mix of FSHD types was included in six studies^63-68^ and the remaining studies did not specify the type of FSHD. Sample sizes ranged from 7^51^ to 714^69^. Few pediatric patients (including children and adolescents) were included, with only one study exclusively conducted in pediatric patients^64^; many studies did not specify how many patients were pediatric, though the age range of the participants indicated that some pediatric patients were included^56,58,70-74^. Additional population characteristics are detailed in **Table 16** in **Appendix 7.3**.

Measurement properties reported included reliability and validity included correlation, Rasch analysis, factor analysis, reliability, responsiveness, and internal consistency; an overview of measurement properties reported in all identified studies is provided in **Table 4.**

Correlation is the degree to which a pair of variables are related and can indicate a predictive relationship. Correlation is reported via either the Pearson correlation coefficient or Spearman’s rank correlation. Among all studies identified in the topic “Disease Outcome Measures and Validation”, correlation was the most often reported measure. A total of 21 conference abstracts and 22 full text studies reported a correlation coefficient (**Table 4**).

A Rasch analysis is a model for analyzing categorical data as a trade-off between the respondent’s characteristics and the item difficulty. It is used often used to guide the development an analysis of tests. Rasch analyses were reported in three publications.^69,75,76^

Factor analysis describes the variability among observed, correlated variables. It also allows for the simplification of a set of items. As with Rasch analysis, it is commonly used when developing and refining instruments to produce evidence for the structural validity of a given measure. Only one conference abstract^77^ and one full text publication^72^ reported factor analyses.

Reliability refers to the consistency of a given measure and may be reported as test-retest reliability (over time), internal consistency (across items) or inter-rater reliability (across different researchers). Reliability was the second most reported in the literature and was reported in a total of 7 conference abstracts and 15 full text publications (**Table 4**).

Responsiveness is a measurement property that reflects the longitudinal validity of an instrument; that is, the degree to which a given score changes within a person is valid. Responsiveness can be reported as the minimal detectable change or as a standardized response mean. Responsiveness was reported in three conference abstracts^74,77,78^ and four full text articles ^61,64,65,79^.

Internal consistency is a measure of reliability, and measures whether the measure yields the same score each time it is administered, if all other things are equal. Two full text articles reported internal consistency using either Cronbach’s alpha^65^ or person separation^69^.

Table 4. Overview of measurement properties reported in “Disease Outcome Measures and Validation” studies

| **Author, year** | **Country** | **Measurement** | **Correlation** | **Rasch analysis** | **Factor analysis** | **Reliability** | **Responsiveness** | **Internal consistency** |
| --- | --- | --- | --- | --- | --- | --- | --- | --- |
| CONFERENCE ABSTRACTS | | | | | | | | |
| Berggren 2022^80^ | Multinational | IOPI | **✓** |  |  | **✓** |  |  |
|  |  | Lip strength | **✓** |  |  |  |  |  |
| Chan 2021^81^ | --- | TUG  CSS  PROMIS-PF | **✓** |  |  |  |  |  |
| Chan 2020^82^ | --- | TUG  oTUG |  |  |  | **✓** |  |  |
| De Lattre 2016^74^ | --- | MFM: D1 |  |  |  |  | **✓** |  |
| Eichinger 2022^83^ | Multinational | MRC scale | **✓** |  |  |  |  |  |
| Eichinger 2021^84^ | Multinational | MMT  QMT  Lower extremity muscles | **✓** |  |  |  |  |  |
| Eichinger 2017^85^ | --- | FSHD-COM |  |  |  | **✓** |  |  |
| Glenn 2016^51^ | United States | EIM | **✓** |  |  |  |  |  |
| Goselink 2017^86^ | --- | Muscle ultrasonography | **✓** |  |  |  |  |  |
| Haley 2020^77^ | --- | FCS | **✓** |  | **✓** |  | **✓** |  |
| Hatch 2022^87^ | Multinational | RWS |  |  |  | **✓** |  |  |
| Hristov 2021^78^ | --- | eTUG  In-home gait speed | **✓** |  |  |  | **✓** |  |
|  |  | Sleep schedule variability | **✓** |  |  |  |  |  |
| Liu 209^52^ | China | Fat infiltration by MRI | **✓** |  |  |  |  |  |
| Olivier 2014^88^ | --- | Peak torque | **✓** |  |  |  |  |  |
| Paoletti 2019^89^ | --- | MFF  waterT2 times | **✓** |  |  |  |  |  |
| Pirola 2018^90^ | Italy | FVC  SpO2  ODI | **✓** |  |  |  |  |  |
| Salort-Campana 2017^53^ | --- | MPI total | **✓** |  |  |  |  |  |
| Statland 2022^91^ | Multinational | MFM: D1  FSHD-COM |  |  |  | **✓** |  |  |
| Statland 2019^92^ | United States | Don/doff coat  Supine to sit  6MWT  Gait | **✓** |  |  |  |  |  |
| Statland 2016^93^ | United States | Wireless gait | **✓** |  |  | **✓** |  |  |
| Statland 2014^63^ | --- | Creatine kinase  Plasminogen pathway markers | **✓** |  |  |  |  |  |
| Vincenten 2021^94^ | --- | MRI total leg fat fraction | **✓** |  |  |  |  |  |
| Wang 2022^95^ | --- | LE lean muscle mass via DEXA scan | **✓** |  |  |  |  |  |
| Wang 2019^96^ | Multinational | MFF burden index  MFF tibilias anterior | **✓** |  |  |  |  |  |
| Wildholm 2022^54^ | --- | LMV via MRI  MFI via MRI  MFF via MRI | **✓** |  |  |  |  |  |
| Woodcock 2020^97^ | Australia | Mercuri score  MRI | **✓** |  |  |  |  |  |
| Zhuparris 2021^98^ | --- | Remote sensor data to estimate clinical severity | **✓** |  |  |  |  |  |
| FULL TEXTS | | | | | | | | |
| Andersen 2017^62^ | Denmark | MFF | **✓** |  |  |  |  |  |
| Bachasson 2014^99^ | France | MVC | **✓** |  |  | **✓** |  |  |
| de Valle 2021^64^ | Australia & New Zealand | FSHD-COM  FSHD-Peds | **✓** |  |  | **✓** | **✓** |  |
| Dos Santos 2021^100^ | Spain | MMSE  MoCA  Trail-Making Test A  Long-term memory (A7) | **✓** |  |  |  |  |  |
| Eichinger 2018^65^ | United States | FSHD-COM | **✓** |  |  | **✓** | **✓** | **✓** |
| Eichinger 2017^61^ | United States | 6MWT | **✓** |  |  | **✓** | **✓** |  |
| Eren 2020^73^ | Turkey | Novel shoulder staging system |  |  |  | **✓** |  |  |
| Esnault 2018^60^ | France | IPT  CSS  Berg Balance Scale  Functional Reach Test  TUG  6MWT  Functional Independence Measure | **✓** |  |  |  |  |  |
| Fatehi 2017^59^ | France | MPI  Duration of disease | **✓** |  |  |  |  |  |
| Goselink 2020^58^ | Netherlands | MUS | **✓** |  |  |  |  |  |
| Guillot 2018^72^ | France | MFM: D3 | **✓** |  | **✓** |  |  |  |
|  |  | MFM: D1  MFM: D2 |  |  | **✓** |  |  |  |
| Han 2015^101^ | United States | RSA | **✓** |  |  |  |  |  |
| Hatch 2021^79^ | United States | RSA | **✓** |  |  |  | **✓** |  |
| Huisinga 2018^102^ | United States | iTUG | **✓** |  |  | **✓** |  |  |
| Lamperti 2010^71^ | Italy | FCS |  |  |  | **✓** |  |  |
| Lareau-Trudel 2015^57^ | France | Visual fat infiltration  IFF with qMRI | **✓** |  |  | **✓** |  |  |
| Mul 2022^76^ | Netherlands | FFS | **✓** | **✓** |  | **✓** |  |  |
| Mul 2021^a69^ | Multinational | FSHD-RODS |  | **✓** |  | **✓** |  | **✓** |
| Mul 2021^b75^ | Netherlands | FSHD-RODS | **✓** |  |  |  |  |  |
|  |  | MFM |  | **✓** |  |  |  |  |
| Mul 2017^66^ | Netherlands | MFF | **✓** |  |  |  |  |  |
| Personius 1994^70^ | United States | QMT  MMT  Grip strength  Pinch strength  FVC  Facial muscle testing |  |  |  | **✓** |  |  |
| Regula 2016^56^ | Germany | MRC score | **✓** |  |  |  |  |  |
| Rijken 2015^103^ | Netherlands | 6MWT  10MWT  TUG  Berg Balance Scale  Step Up Test  Step Down Test  Sit-to-Stand Test | **✓** |  |  | **✓** |  |  |
| Sanson 2022^104^ | France | Brooke scale  Vignos Scale  Visual analogue scale: pain |  |  |  | **✓** |  |  |
| Statland 2016^67^ | United States | EIM | **✓** |  |  | **✓** |  |  |
| Tawil 1997^105^ | United States | MMT score  MVICT score | **✓** |  |  |  |  |  |
| Teeselink 2022^68^ | Netherlands | FVC | **✓** |  |  |  |  |  |
| Widholm 2022^55^ | Multinational | LMV  MFF |  |  |  | **✓** |  |  |

6MWT, 6-minute walk test; 10MWT, 10-minute walk test; 14SST, 14 step stair test; BMI, body mass index; CSS, clinical severity score; D, dimension; EIM, electrical impedance myography; eTUG, electronic timed up and go; FCS, FSHD clinical score; FFS, Facial Function Scale; FSHD; facioscapulohumeral muscular dystrophy; FSHD-COM, FSHD composite outcome measures; FSHD-HI, FSHD health index; FSHD-RODS, FSHD Rasch-built disability scale; FVC, forced vital capacity; IFF, infiltrated fat fraction; IOPI, Iowa Oral Performance Instrument; IPT, isokinetic peak torque; LMV, lower muscle volume; MFF, muscle fat fraction; MFI, muscle fat infiltration; MFM; Motor Function Measures; MMSE, Mini-Mental State Examination; MMT, manual muscle testing; MoCA, Montreal Cognitive Assessment; MPI, mean pixel intensity; MRC, Medical Research Council; MVC, maximum voluntary contractions; oTUG, optimized TUG; QMT, quantitative muscle testing; RSA, relative surface area; RWS, reachable work space; TUG, timed up and go.

Construct validity as measured by correlation was the most reported validation measure (**Table 17** in **Appendix 7.3**). Correlation was assessed for multiple instruments and outcomes including strength, functional assessments, clinical severity, fat fraction, lean muscle mass, shoulder mobility, lung function, muscle composition, patient-reported outcomes (PROs), and duration of disease. Multiple studies noted strong to excellent correlations between measures. Upper extremity strength was found to be significantly correlated (p<0.0001) with the don/doff coat assessment.^83,84^ Correlations, as noted by authors in the studies, ranging from good to very strong were found between measures of gait (expanded Timed Up-and-Go [eTUG] and gait speed) along with sleep and various clinical, functional and PRO measures.^78^ Chan *et al.* validated the use of the FSHD-TUG as a clinical outcome assessment of mobility and reported a strong correlation between the TUG and FSHD-TUG.^81^ Significant correlations were found between fat fraction and clinical severity and muscle strength^94^ and fat infiltration and strength^52,62^. Wildholm *et al*. found that magnetic resonance imaging (MRI) measurements can potentially provide important information about disease severity and progression.^54^ de Valle *et al*. found that the FSHD Composite Outcome Measure (FSHD-COM) and the FSHD-COM pediatrics (FSHD-COM peds) correlated well with other measures of disease severity.^64^

Three studies used a Rasch analysis (**Table 18** in **Appendix 7.3**). One analysis assessed the MFM in FSHD patients and found that it should only be used with caution in this group as it is not optimally suited to measure functional abilities.^75^ The second analysis assessed the FSHD-Rasch-built overall disability scale (FSHD-RODS) and found good item/person reliability and validity scores.^69^ The third study evaluated the developed Facial Functional scale, and determined that the scale was a clinimetrically sound patient-reported outcome measure for functional disabilities relating to facial weakness in FSHD patients.^76^

Factor analysis found that the FSHD clinical score was not informative in those patients with extremely mild or severe symptoms (**Table 19** in **Appendix 7.3**).^77^ Guillot *et al*. found that factor analysis was able to establish the construct validity of the MFM.^72^

Inter-rater, intra-rater and test-retest was used to assess reliability (**Table 20** in **Appendix 7.3**). Excellent test-retest reliability was noted for the FSHD-COM and domain 1 of the MFM^91^ along with RWS^87^. Statland *et al* found good to excellent reliability via test-retest using wireless gait metrics.^93^ The FSHD-RODS^69^, the FSHD-COM^64^, the instrumented TUG (iTUG^102^) and the 6MWT^61^ were found to have excellent reliability. A newly developed FSHD score was found to be reliable and easily used for monitoring the functional status of patients.^71^ Eren *et al*.^73^ found that their developed system for shoulder staging was reliable in guiding surgical decisions, with excellent inter- and intra-observer agreement. Antigravity tests (sit to stance, stance to sit, step up and step down) were found to be reliable when assessing functional capacity in FSHD patients.^103^ Other studies assessed the reliability of various muscle strength or muscle composition measures.^55,57,67,70,99^

Responsiveness was reported for the eTUG^78^, in-home gait speed^78^, the FSHD clinical score^77^, the MFM^74^, the FSHD-COM^64,65^ and the FSHD-COM Peds^64^, and the 6MWT^61^ (**Table 21** in **Appendix 7.3**). Most responsiveness was reported as a minimal detectable change using either 90% or 95% confidence intervals; one study reported standardized response means^74^.

Internal consistency was reported in two studies for two different instruments: the FSHD-RODS^69^ and the FSHD-COM^65^, reporting the person separation index and Cronbach’s alpha, respectively (**Table 22** in **Appendix 7.3**). Both measures were declared by the authors to be suitable to detect activity and participation restrictions (FSHD-RODS) as well as cross-sectional associations (FSHD-COM).

#### 3.4 Humanistic Burden and PROs

A total of 38 full text articles and 15 conference abstracts were identified for the topic “Humanistic Burden and PROs”. The most frequently represented study design across all identified studies was observational studies (n = 36) followed by clinical trials (both randomized and non-randomized; n=10), qualitative studies (n = 5) and case studies/series (n=2). Of the observational studies, the majority were cross-sectional in nature, assessing humanistic burden at a single point in time.

An overview of the studies by quality of life (QoL) domain is provided in **Table 5**. Some studies did not report data but noted the use of an instrument in the Methods; thus, they are included in order to provide an overview of the domains noted to be measured. Among all studies, general QoL, as measured by a PRO instrument, was the most reported domain (n=21), followed by fatigue (n=14) then pain (n=12). A total of eight studies reported humanistic burden results qualitatively without the use of a PRO instrument.

Table 5. Overview of studies reporting “Humanistic Burden and PROs”

| **Author, year** | **Country** | **Study design** | **General QoL** | **Disease-specific QoL** | **Depression** | **Anxiety** | **Pain** | **Sleep** | **Activities of daily living** | **Disability / function** | **Fatigue** | **Neurological / psychological** | **Symptoms / illness perception** | **Qualitative assessment** |
| --- | --- | --- | --- | --- | --- | --- | --- | --- | --- | --- | --- | --- | --- | --- |
| CONFERENCE ABSTRACTS | | | | | | | | | | | | | | |
| Bergsma 2012^106^ | Netherlands | Cross-sectional |  |  |  |  |  |  |  | **✓** |  |  |  |  |
| Dahlqvist 2014^107^ | NR | Cross-sectional |  |  |  |  | **✓** |  |  |  |  |  |  |  |
| Evangelista 2015^108^ | UK | Retrospective cohort |  | **✓** |  |  | **✓** |  |  |  |  |  |  |  |
| Gazzotti 2011^109^ | NR | Case series | **✓** |  |  |  |  |  |  |  |  |  |  |  |
| Gergev 2013^110^ | NR | Qualitative |  |  |  |  |  |  |  |  |  |  |  | Cognitive |
| Heatwole 2012^111^ | US | Cross-sectional |  |  |  |  |  |  |  |  |  |  |  | Activities of daily living |
| Hodge 2016^112^ | US | Prospective cohort |  |  |  |  |  |  |  |  |  |  |  | Disability |
| Kools 2022^16^ | NR | Non-randomized trial |  | **✓^b^** |  |  | **✓** |  |  | **✓** |  |  |  |  |
| Leclair-Visonneau 2013^113^ | NR | Cross-sectional |  |  | **✓^b^** | **✓** |  | **✓^b^** |  |  | **✓^b^** |  |  |  |
| Patel 2017^114^ | New Zealand | Cross-sectional |  |  |  |  | **✓** |  |  |  |  |  |  |  |
| Shirilla 2019^115^ | US | Cross-sectional |  |  |  |  |  |  |  |  |  |  |  | General QoL |
| Statland 2012^116^ | US | Retrospective cohort |  |  |  |  |  |  |  |  |  |  |  | Disability |
| Tawil 2022^17^ | NR | Randomized controlled trial |  | **✓** |  |  | **✓** |  |  |  |  |  |  |  |
| Vissing 2017^117^ | NR | Non-randomized trial |  | **✓** |  |  |  |  |  |  |  |  |  |  |
| Walker 2017^19^ | NR | Non-randomized trial |  | **✓** |  |  |  |  |  |  |  |  |  |  |
| FULL TEXTS | | | | | | | | | | | | | | |
| Andersen 2017^41^ | Denmark | Randomized controlled trial | **✓** |  |  |  | **✓** |  |  |  | **✓** |  |  |  |
| Aprile 2013^118^ | Italy | Case-control | **✓^a^** |  |  |  | **✓^a^** |  |  |  |  |  | **✓^a^** |  |
| Bachasson 2014^99^ | NR | Cross-sectional | **✓** |  |  |  |  |  |  |  | **✓** |  |  |  |
| Bakker 2017^119^ | Netherlands | Qualitative |  |  |  |  |  |  |  |  |  |  |  | Illness perception |
| Bankolé 2016^21^ | France | Randomized controlled trial | **✓** |  |  |  |  |  |  |  | **✓** |  |  |  |
| Bayram 2020^120^ | Turkey | Prospective cohort | **✓** |  |  |  |  |  |  |  |  |  |  |  |
| Bungener 1998^121^ | France | Cross-sectional |  |  | **✓** | **✓** |  |  |  |  |  | **✓** |  |  |
| de Valle 2021^64^ | Australia & New Zealand | Cross-sectional |  | **✓** |  |  |  |  |  |  |  |  |  |  |
| Dijkstra 2021^122^ | Netherlands | Prospective cohort | **✓** |  |  |  |  |  |  |  | **✓** |  |  | Pain and fatigue |
| Doix  2017^22^ | France | Prospective cohort | **✓** |  |  |  |  |  |  |  |  |  |  |  |
| Dos Santos^100^ | Brazil | Cross-sectional | **✓** |  |  |  |  |  |  |  |  | **✓** |  |  |
| Eren 2020b^34^ | Turkey | Case study |  | **✓** |  |  |  |  |  |  |  |  |  |  |
| Gallais 2015^123^ | France | Cross-sectional |  |  | **✓** | **✓** |  |  |  | **✓** | **✓** | **✓** |  |  |
| Goselink 2018^124^ | Netherlands | Cross-sectional |  |  |  |  |  |  |  |  | **✓** |  |  | Pain |
| Guy-Coichard 2008^125^ | France | Cross-sectional |  |  |  |  | **✓** |  |  | **✓^b^** |  |  |  | Pain |
| Hamel 2019^126^ | US | Cross-sectional |  |  |  |  |  |  |  |  |  |  |  | Symptoms |
| Hatch 2021^79^ | US | Cross-sectional |  | **✓^b^** |  |  |  |  |  |  |  |  |  |  |
| Hoffmann 2022^127^ | US | Prospective cohort |  |  |  |  |  | **✓** |  |  |  |  |  |  |
| Jacques 2019^128^ | UK | Cross-sectional | **✓** |  |  |  | **✓** |  | **✓** |  | **✓** | **✓** |  |  |
| Jensen 2008^129^ | US | Cross-sectional |  |  |  |  | **✓** |  |  |  |  |  |  |  |
| Johnson 2012^130^ | NR | Qualitative |  |  |  |  |  |  |  |  |  |  |  | Symptoms |
| Kalkman 2005^131^ | Netherlands | Cross-sectional | **✓** |  |  |  |  |  |  |  | **✓** |  |  |  |
| Koopman 2017^132^ | Netherlands | Cohort analysis |  |  |  |  |  |  |  |  | **✓** | **✓** | **✓** |  |
| Loock 2022^47^ | NR | Retrospective cohort |  |  |  |  | **✓** |  |  | **✓** |  |  |  |  |
| Lue 2009^133^ | Taiwan | Cross-sectional |  |  |  |  |  |  |  | **✓** |  |  |  |  |
| Minis 2010^134^ | Netherlands | Cross-sectional | **✓^a^** |  |  |  |  |  |  |  | **✓^a^** |  |  |  |
| Moris 2018^135^ | UK | Cross-sectional |  | **✓** |  |  | **✓** |  |  |  |  |  |  |  |
| O’Dowd 2021^136^ | UK | Cross-sectional | **✓** |  | **✓** | **✓** |  |  |  |  |  | **✓** |  |  |
| Padua 2009^137^ | Italy | Prospective cohort | **✓** |  | **✓^b^** |  | **✓** |  |  |  |  |  |  |  |
| Payan 2009^138^ | France | Randomized controlled trial | **✓** |  |  |  |  |  |  | **✓** |  |  |  |  |
| Ribot-Ciscar 2015^50^ | France | Prospective cohort | **✓** |  |  |  | **✓** |  |  |  |  |  |  |  |
| Rose 2012^139^ | US | Cross-sectional | **✓^b^** | **✓** | **✓^b^** | **✓^b^** |  |  |  | **✓** |  |  | **✓** |  |
| Schipper 2017^140^ | Netherlands | Qualitative |  |  |  |  |  |  |  |  |  |  |  | Fatigue |
| Sezer 2022^141^ | Netherlands | Qualitative |  |  |  |  |  |  |  |  |  |  |  | Disability |
| Statland 2022^38^ | US  Canada  Spain | Randomized controlled trial |  | **✓** |  |  |  |  |  |  |  |  |  |  |
| van de Geest-Buit 2023^142^ | Netherlands | Cross-sectional | **✓** | **✓** | **✓** | **✓** |  |  |  | **✓** | **✓** |  |  |  |
| van der Kooi 2007^143^ | Netherlands | Randomized controlled trial | **✓** |  | **✓** |  | **✓** |  | **✓** |  | **✓** | **✓** |  |  |
| Voet  2014^144^ | Netherlands | Randomized controlled trial | **✓** |  |  |  | **✓** | **✓** |  |  | **✓** |  |  |  |
| Winter 2010^145^ | Germany | Cross-sectional | **✓** |  |  |  |  |  |  |  |  |  |  |  |

NR, not reported; QoL, quality of life; UK, United Kingdom; US, United States

^a^ Data reported by subgroup only

^b^ Data not reported but use of an instrument mentioned in Methods and / or Results

The study population included for the topic “Humanistic Burden and PROs”, along with baseline characteristics for all included studies is presented in **Table 23** in **Appendix 7.4**. Population characteristics are reported for the overall study group, where available. Sample size ranged from 1 (case study^34^) to 690 (prospective cohort^127^). While most studies did not specify the type of FSHD included in the study population, 11 studies were conducted in a Type 1 FSHD population exclusively^16,17,19,21,22,79,108,122,132,135,146^ and 3 studies in a mixed Type 1 / Type 2 FSHD population^38,64,142^. The age of participants was not always explicitly noted: six conference abstracts and one of full text did not note a mean, median or age range, thus it was not always possible to determine whether pediatric participants were included. Of the studies that did report age, six conference abstracts and five full text manuscripts included pediatric participants; a total of three studies (in four publications)^64,122,124^ were conducted solely in pediatric patients. Studies included an FSHD population with a disease duration range from 6.7^100^ to 26^135^ years. As with the topic “Treatment Outcomes and Clinical Trials”, the clinical severity score and the number of D4Z4 pairs were not well reported across studies.

##### 3.4.1 General QoL

Multiple instruments were used to assess general QoL among participants**; Section 7.3** provides a brief description of each used.

The most used instrument to report general QoL was the Short-Form 36 (SF-36; **Section 7.5.57**), with data reported in 16 studies, with sample sizes ranging from 3 to 1,349; Rose *et al.* did not report data from the SF-36, but provided a narrative summary^139^. The majority of studies reported SF-36 scores for a single point in time, with the exception of Bankolé *et al*.^21^, Doix *et al*.^22^, and Ribot-Ciscar *et al*.^50^ For studies reporting the mental component score, scores ranged from 47.4^137^ to 60.8^120^; for studies reporting the physical component score, scores ranged from 30.8^100,128^ to 38.7^120^. As higher scores indicate higher QoL, these findings suggest, based on the SF-36, that patients with FSHD have higher mental versus physical well-being. Other studies reported SF-36 results for the individual scales only^50,99,131,142,145^. While Bankolé *et al*.^21^ reported results over time, they did not detail which score of the SF-36 was being reported. Andersen *et al.^20^,* Gazzotti *et al*.^109^ and Payan *et al.^138^* only reported narrative SF-36 results. Minis *et al*.^134^ and Aprile *et al*.^118^ only reported data by subgroup (employed/not employed and shoes with/without orthotics, respectively); as subgroup data extraction was beyond the scope of this project, it is not reported.

Other general QoL instruments, each reported by one study, were the Kidscreen (**Section** **7.5.33**), the Sickness Impact Profile (SIP; **Section 7.5.60**), the EQ-5D (**Section 7.5.12**) and the EQ Visual Analogue Scale (VAS; **Section 7.5.67**). Both the Kidscreen and the SIP were reported in studies at both baseline and follow-up. Dijkstra *et al*., using the Kidscreen, reported the mean SD at baseline and 2-year follow-up for children with Type 1 FSHD (age range 2 to 17).^122^ After a two-year observational follow-up, total score improved from -0.9 SD to -0.7 SD, however, this improvement was not significant; the only subscale that was found to be significantly different was “parent relation/home life”. Two studies reported results using the SIP: van der Kooi *et al.*^143^ and Voet *et al*.^144^, who both reported both baseline and follow-up results. While no significant results were observed between training versus no training and albuterol versus placebo at follow-up in van der Kooi *et al.*^143^, Voet *et al*.^144^ did observe a significant difference between both aerobic exercise training cognitive behavioral therapy versus usual care at both 16- and 28-weeks. Other measures of general QoL included the EQ-5D and EQ VAS reported by Winter *et al*.^145^ In the EQ-5D, a total of 82.4% (n=14) respondents noted moderate or severe problems in at least one of the 5 dimensions.

##### 3.4.2 Disease-specific QoL

Disease-specific QoL instruments are summarized in **Table 25** in **Appendix 7.4** and included FSHD-specific instruments as well as instruments for neuromuscular diseases. Instruments included the FSHD-Health Index (HI) for adults (**Section 7.5.16**), the FSHD-HI for pediatric patients (**Section 7.5.16**), the pediatric quality of life neuromuscular module (Peds QL NMM; **Section 7.5.47**), the Individualized Neuromuscular Quality of Life (INQoL; **Section 7.5.29**); the Quality of Life in Neurological Disorders (NeuroQoL; **Section 7.5.44**) were reported to be used, but data was not reported.

The INQoL was used in six studies.^19,44,108,117,135,139^ Scores were presented as a percentage of the maximum detrimental impact, with higher scores indicating a greater impact on QoL. While Evangelista *et al.^108^* and Rose *et al.^139^* reported mean INQoL values, Moris *et al.^135^* reported median values. Of the four studies reporting data, the domain with the greatest impact on QoL across all studies was muscle weakness (mean range of 59.9^139^ – 94.74^34^. Vissing *et al.^117^* and Walker *et al.^19^* only reported the results of the INQoL qualitatively within the text, as both these studies were conference abstracts.

de Valle *et al.* ^64^ used disease-specific pediatric instruments and reported data using the FSHD-HI Peds and the Peds QL NMM. The study was conducted among 18 pediatric patients (Type 1: n=16; Type 2: n=1; missing: n=1), with an age range of 7 to 18. The mean (SD; range) score of the FSHD-HI Peds was 21.9 (22.5; 0.2 – 67.5) and the mean (SD; range) of the child assessment of the PedsQL NMM was 73.9 (15.7; 47 – 95); the parent proxy of the PedsQL NMM was similar to the child assessment with a mean (SD; range) or 73.5 (17.1; 42 – 97).

Statland *et al.^38^* and Tawil *et al. ^17^* both reported the use of the FSHD-HI. Statland *et al.* did not report any significant differences in absolute scores between ACE-083 versus placebo for either the biceps brachii or tibilias anterior. As Tawil *et al*. was a conference abstract, detailed results were not provided, however, they did note that “no differences were observed” between losmapimod and placebo.

##### 3.4.3 Depression and anxiety

Depression and anxiety are presented together as some scales, such as the Hospital Anxiety Depression Scale (HADS), consider these two outcomes together. Multiple studies used multiple measures to assess depression and/or anxiety (**Table 26** in **Appendix 7.4**).

Anxiety was assessed using the Covi brief anxiety scale^121^ (**Section 7.5.8**), the HADS^113,139,142^ (**Section 7.5.24**), the State-Trait Anxiety Inventory^123,136^ (STAI; **Section 7.5.61**), the Social Appearance Anxiety Scale^142^ (SAAS; **Section 7.5.54**), the Social Interaction Anxiety Scale^142^ (SIAS; **Section 7.5.59**), and the Tyrer anxiety scale^121^ (**Section 7.5.65**). All instruments were used once with the exception of the HADS and STAI. While the HADS was reported in three studies, only van de Geest-Buit *et al.* ^142^ reported data; data was not reported in either Leclair-Visonneau *et al*. ^113^ nor Rose *et al*. ^139^. In addition to reporting the total score, van de Geest-Buit *et al.* ^142^ reported that 21.7% of FSHD patients scored above 8, with a score of 0 to 7 represents normal anxiety. The study also reported that few patients (0.7%) scored in the most negative 10% for fear of negative evaluation and no patients scored in the most negative 10% for social interaction anxiety.^142^ The STAI was reported by both Gallaiis *et al*. ^123^ (T score for A and B of mean [SD] 44.8 [11.0] and 48.7 [12.9], respectively) and O’Dowd *et al.* ^136^ (mean [SD] of 38.6 [11.6]).

Depression was assessed using the Beck Depression Inventory^136,137,143^ (BDI; **Section 7.5.3**), the Depressive Mood Scale^121^ (EHD; **Section 7.5.11**), the HADS^113,139,142^ (**Section 7.5.24**), the Hamilton Depression Rating Scale^121^ (HDRS; **Section 7.5.26**), the Mini-International Neuropsychiatric Interview (MINI; **Section 7.5.39**), and the Montgomery-Asberg Depression Rating Scale^121,123^ (MADRS; **Section 7.5.38**). Gallais *et al.,* using the MINI, found that 19 patients (5.3%) met the criteria for a current major depressive disorder.^123^ For studies using the BDI, O’Dowd *et al.* reported mean scores of 10.9^136^, representing mild depression, and van der Kooi *et al.* reported that 16% of patients were depressed (scoring 4 or above) at baseline in their RCT^143^; Padua *et al.* did not report data using the BDI^137^. While no significant differences were observed between groups in the RCT by van der Kooi *et al.* depression scores were generally lower at follow-up at week 52 compared to baseline.

##### 3.4.4 Pain

Pain was the second-most reported domain within the topic “Humanistic Burden and PROs” (**Table 27** in **Appendix 7.4**). Similar to other domains, multiple instruments were used to assess pain among FSHD patients, with some studies using more than one instrument.

Dahlqvist *et al.* assessed the prevalence of low back pain using the Low Back Pain Rating scale (LBPR; **Section 7.5.36**); the prevalence of lower back pain was significantly higher in FSHD patients than in controls (p<0.0001).^107^

The McGill Pain Questionnaire (MPQ; **Section 7.5.42**) was reported using the full version^108,114^, the short-form^135^ and the VAS within the MPQ^143^. Evangelista *et al.* found that 50% of FSHD patients reported persistent pain and 43% of FSHD patients reported recurrent localized pain; the shoulder was the most affected location for both.^108^ Patel *et al.* stated in their abstract that 69% of FSHD patients reported pain, with no further details of the MPQ provided.^114^ Moris *et al.*, using the short form of the MPQ, reported that 339 patients (85.2%) had presence of pain, with a present pain intensity index median of 1 (representing mild pain, on a scale of 1 to 5).^135^ van der Kooi assessed pain in their RCT using the MPQ VAS; no significant differences were observed between albuterol and placebo with or without strength training, from baseline to week 52 follow-up.^143^

Pain was assessed using a VAS (**Section 7.5.67**) in other studies as well. Guy-Coichard *et al.* reported that 30% of patients reported severe usual pain and 21% of patients reported moderate usual pain.^125^ Jacques *et al.* reported a mean pain score of 3.9 on a scale of 1 to 10.^128^ Padua *et al.* reported that 56% of FSHD patients reported mild/moderate pain, with a mean VAS of 31.6 on a scale of 0 to 100.^137^ Loock *et al*. reported that pre- and post-operative pain were significantly different.^47^

Both the universal pain assessment tool (**Section 7.5.66**) and the grading of chronic pain scale (**Section 7.5.22**), are similar to a VAS. The universal pain assessment tool was used by Moris *et al.*, and found that 55.6% (n=203) and 30.4% (n=69) of FSHD patients had total chronic and severe chronic pain, respectively.^135^ The grading of chronic pain scale was used by Jensen *et al.* with 92% (n=104) and 23% (n=24) of FSHD patients reporting any and severe pain, respectively.^129^ While not explicitly collected by this scale, the duration of pain was noted to be a mean (SD) of 13.4 (10.4) years.

Other tools to assess pain included the Patient Global Impression of Change scale (PGIC, **Section 7.5.49**), used by Kools *et al.* and Tawil *et al.* Kools *et al.* reported that over 80% of FSHD patients found improvement or no change in their pain over a 56-week period of receiving losmapimod^16^; Tawil *et al*. reported significant improvement in pain after treatment with losmapimod after 48 weeks compared to placebo (data not reported).^17^ Jensen *et al.*, in addition to the grading of chronic pain scale, used the Brief Pain Inventory (BPI; **Section 7.5.5**).^129^ Based on a scale of 0 to 10, the total mean pain score reported by FSHD patients was 2.74; the highest pain scores were reported for mobility (mean: 3.65) and lowest pain scores were reported for learning new skills (mean: 1.14). ID pain (**Section 7.5.27**) is a questionnaire focused on neuropathic and joint pain in patients; Padua *et al.* found that 6.7% of FSHD patients had very probable neuropathic pain with 15.6% of patients reporting joint pain exclusively.^137^ The Daily Observed Pain instrument (DOP; **Section 7.5.9**) was used in the RCT by van der Kooi *et al.*^143^ While no significant differences were observed in the DOP score between albuterol and placebo groups, with or without strength training, mean DOP scores may be considered on the lower end of the scale (range of 0 to 16).

Finally, Guy-Coichard *et al.* reported the impact of pain on various QoL domains, however, it was not clear which instrument was used to assess the impact of pain.^125^

##### 3.4.5 Sleep

Sleep was assessed using three different instruments in three studies (**Table 28** in **Appendix 7.4**). For the Pittsburgh Sleep Quality Index (PSQI; **Section 7.5.51**), Hoffmann *et al.* reported that 66.2% of participants (n=392) scored over 5, indicating “poor sleepers”.^127^ While Leclair-Visonneau *et al.* did not report the scores, they indicated that the FSHD population in the study had significantly worse sleep quality than those in the control group (p<0.01).^113^ For the Epworth Sleepiness Scale (ESS; **Section 7.5.13**), Hoffmann *et al.* reported that 15.0% scored over 10, indicating abnormal sleep (a score of 0 to 10 is considered “normal”). ^127^ Leclair-Visonneau *et al.* did not report data for the ESS. The sleep subscale on the Nottingham Health Profile (NHP; **Section 7.5.45**) was used by Voet *et al*. in their RCT. Significant differences were observed from baseline to 16-weeks between aerobic exercise or cognitive behavioral therapy versus usual care; however, significant differences were not observed at 28-weeks.^144^

##### 3.4.6 Activities of daily living

Activities of daily living refer to essential and routine tasks; these can be performed without assistance for healthy individuals. In the FSHD population, performing some activities of daily living may be challenging. Both Jacques *et al.*^128^ and van der Kooi *et al.*^143^ reported activities of daily living in FSHD patients (**Table 29** in **Appendix 7.4**). Jacques *et al.* used the Nottingham Extended Activities of Daily Living instrument (NEADL; **Section 7.5.42**) and reported a mean score of 39.4, where scores of 43 or less indicate the need for assistance of.^128^ van der Kooi *et al.* reported the use of the Daily Observed Activity instrument (DOA); a higher DOA score indicates more self-reported activity, however, no further information was available on this instrument, including a range or cut-off scores indicating lower capacity for activities, therefore interpretation of results is limited.^143^

##### 3.4.7 Disability and / or functional status

The disability and/or functional status of FSHD patients was assessed with multiple instruments; some of these instruments assess functional status separately for upper and lower extremity (**Table 30** in **Appendix 7.4**).

Upper extremity disability/function was assessed with the Abiland^106^ (focused only on the hand; **Section 7.5.1**), the Brooke Upper Extremity scale^47,106,125,133,138^ (BUE; **Section 7.5.6**) and the Capabilities of Upper Extremity scale^106^ (CUE; **Section 7.5.9**); lower extremity disability/function was assessed using the Vignos Lower Extremity Scale^47,125,133,138^ (VLE; **Section 7.5.68**). Of studies reporting the BUE, none reported mean scores of less than 2; a score of 2 represents “raising arms above head only by flexing the elbow” and 3 represents “cannot raise hands above head”. Similarly, of studies reporting data for the VLE, mean scores ranged from 1.7^138^ to 2.2^133^. A score of 2 on the VLE represents “walks and climbs stairs with aid of railing” and 3 represents “walks and climbs stairs slowly with aid of railing”.

General disability/functional status PRO instruments included the Barthel index (**Section 7.5.3**), the Health Assessment Questionnaire (HAQ; **Section 7.5.25**), the Walton Functional Scale (WFS; **Section 7.5.69**). A face-specific questionnaire was also used by van de Geest-Buit *et al.*^142^ to assess both impairment and disability associated with facial dysfunction (Section **7.5.15**). The FSHD-RODS (**Section 7.5.20**) was used in two studies^16,142^, however, data was not reported for either of these studies.

##### 3.4.8 Fatigue

Fatigue in the FSHD population was measured by three different validated questionnaires: the Checklist Individual Strength (CIS; **Section 7.5.7**), the Fatigue Severity Scale (FSS; **Section 7.5.21**), and the Quality of Life in Neurological Disease (NeuroQoL; **Section 7.5.44**); additionally, a VAS was used to assess fatigue (**Section 7.5.67**). van der Kooi *et al.* reported the use of the Daily Observed Fatigue scale (DOF), however, no additional detail was provided on its development or interpretation. A summary of the outcomes for the domain fatigue is presented in **Table 31** in **Appendix 7.4**.

Scores for the CIS were presented for multiple domains of the CIS, including fatigue severity, in one study^131^, and for the fatigue severity domain only in the remaining studies.^128,132,142-144^ Only one of the studies noted the proportion of patients scoring above the cutoff indicating severe fatigue: van de Geest-Buit *et al.* noted that 5.1% of patients scored 35 or over.^142^ While the proportion scoring 35 or over was not explicitly noted in Jacques *et al.*, the mean (SD) score was 43.0 (5.3).^128^

A score of 5 or greater indicates severe fatigue on the FSS; Gallais *et al.*^123^ reported a mean score below this. Bachasson *et al.* and Bankolé *et al.* transformed their scores and did not indicate a cutoff for interpreting severe fatigue.^21,99^ Bachasson *et al.*, however, did report that 11 (57.9%) of patients experienced severe fatigue as measured by the FSS.^99^

The fatigue scale of the NeuroQoL was reported in one study with two publications*.*^122,124^; compared to healthy subjects, the FSHD population had mean SD scores of 1.1 above average for fatigue. After 2-year follow-up, the mean SD score decreased to 1.0.^122^

##### 3.4.9 Neurological / psychological

This SLR also included all instruments that assess neurological or psychological well-being, including measurements of cognitive function. Detailed results are presented in **Table 32** in **Appendix 7.4**.

Neurological or psychological instruments included the Abrams scale for emotional blunting (**Section 7.5.2**), scales to measure anhedonia, which is the inability to feel pleasure, for both physical (**Section 7.5.45**) and social (**Section 7.5.55**) anhedonia, the General Self-Efficacy Scale (GSE; **Section 7.5.23**), the Jacobsen-Fatigue Catastrophizing Scale (J-FCS, **Section 7.5.33**), the Lille Apathy Rating Scale (LARS; **Section 7.5.35**), the Mini Mental State Evaluation (MMSE; **Section 7.5.41**), the Pain Catastrophizing Scale (PCS; **Section 7.5.47**), the Physical Self-Perception Profile (PSPP; **Section 7.5.50**), the Rosenberg Self-Esteem Scale (RSES; **Section 7.5.53**), the Self-Efficacy Scale (SES; **Section 7.5.57**), the Symptom Checklist (SCL; **Section 7.5.56**) and the Social Support Inventory (SSI; **Section 7.5.61**).

Gallais *et al.* noted that 21.1% of patients were apathetic, as measured by the LARS.^123^. Only one study explored psychological outcomes over time: van der Kooi *et al.* explored the impact of strength training with or without albuterol over a 52-week period in an RCT. While no significant differences were observed between groups over time, SCL scores after 52 weeks were lower in all groups.^143^ Jacques *et al*. reported that while the self-efficacy of FSHD patients were lower than a healthy control group, differences were not significantly different (as measured by the GSE).^128^ O’Dowd *et al*. also compared FSHD patients to a healthy control group, with significantly lower scores observed for PSPP (p<0.05) but not for RSES.^136^

Multiple studies aimed to assess the cognitive function of FSHD patients. These scales included the Frontal Assessment Battery (FAB; **Section 7.5.14**), the F-A-S test (**Section 7.5.16**) and the F-A-S categorical test (**Section 7.5.17**), the Montreal Cognitive Assessment (MoCA; **Section 7.5.39**), the Rey-Auditory Verbal Learning Test (RAVLT; **Section 7.5.52**), the Stroop test (**Section 7.5.63**), and the Trail Making Test (TMT; **Section 7.5.64**). Dos Santos *et al.^100^* reported the proportion assessed to be cognitively impaired using a variety of instruments: depending on the instrument used, the proportion cognitively impaired ranged from 0% with the F-A-S test to 100% using the TMT test A. They concluded that FSHD individuals have mild impairment in tasks involving attention, planning, and long-term memory functions.

##### 3.4.10 Symptoms

The impact of FSHD symptoms on one’s life was reported in three studies (**Table 33** in **Appendix 7.4**).

Koopman *et al.* reported results using three questionnaires: the Illness Cognitions Questionnaire (ICQ; **Section 7.5.27**); the Impact of Event Scale (IES; **Section 7.5.29**), and the Illness Management Questionnaire (IMQ; **Section 7.5.30**).^132^ Out of a possible score of 24, the mean score of the ICQ acceptance subscale was 17.4. The reported mean score of 6.0 indicates that FSHD patients in this study were not significantly impacted by a diagnosis of FSHD. The mean score of 3.1 from the IMQ indicates a lower focus on fatigue symptoms.

Rose et al. assessed the impact of symptoms on FSHD using the Illness Perception Questionnaire – Revised (IPQ-R; **Section 7.5.32**). For the IPQ-R, while an overall score was not calculated, mean scores on the individual subscales were reported. Higher scores on the consequences, timeline acute/chronic and cyclical subscales represent strongly held beliefs about the negative consequences and the chronicity and cyclical nature of the illness. Higher scores on the personal and treatment control and illness coherence subscales represent positive beliefs about controllability and a personal understanding of the illness.

Aprile *et al.* noted the use of the Lumbar Spine Questionnaire (LSQ; **Section 7.5.37**), however, data was only reported by subgroup of shoes with/without orthoses.^118^

##### 3.4.11 Qualitative

Some studies reported humanistic burden without the use of a formal PRO instrument, with either qualitative interviews with FSHD patients, or asking them specific questions regarding their quality of life. These are summarized in **Table 34** in **Appendix 7.4**.

Common themes reported by these studies include problems or weakness with shoulders or arms^111,115,126^, prevalence of pain^122,124,125^ and fatigue^122,124,140^. Gergev *et al*. reported that the cognitive profile of FSHD patients varied between normal abilities and mild intellectual disability, with all patients showing deficits in verbal and working memory and a reduced learning capacity.^110^ Bakker *et al*. via qualitative interviews explored illness perception themes among FSHD patients and reported both intra- (i.e. feelings evoked when hearing the diagnosis) and extra- (i.e. responses of the social environment to the diagnosis) individual themes.^119^ Sezer *et al*. explored facial weakness disability among FSHD patients.^141^ Among the 16 participants included (7 with moderate facial weakness and 9 with severe facial weakness), the resulting themes that emerged included how facial weakness impacts multiple aspects of life, the coping of facial weakness by the patients, and avoidance of discussing facial weakness. Johnson *et al*. analyzed 1,375 direct quotes from 20 participants; the most frequently identified symptoms were “impaired walking” (n=16), “difficult with stairs” (n=15) and “difficulty lifting objects” (n=14).^130^

#### 3.5 Economic Burden

The SLR identified 18 studies (12 full text articles and 6 conference abstracts) that reported at least one economic burden outcome and were included in the SLR (**Table 35** in **Appendix 7.6**). Of those that reported a geographic location, 9 studies were in the US, 6 in Europe (2 in Germany and 4 in the Netherlands), and 2 in the United Kingdom; 1 study did not report the geographic location. Two major study designs were used to assess economic burden: cohort (either retrospective or prospective) or survey, with most surveys being cross-sectional. Most studies did not detail the FSHD population included; of those which did (n=6), four were exclusively among those with Type 1 FSHD^135,147-149^, and two were in a mixed FSHD population^127,150^. Of the studies that reported the age of participants, all were among adults with mean/median age between 40 – 60 years of age; one study appeared to include pediatric patients according to the age range, but this was not specified^147^. Few studies reported the age at onset of FSHD, which was in the early 20s for all studies with the exception of Katz *et al.* which reported a median age of onset of 17 years^148^. None of the economic burden studies reported severity of disease and only three included information regarding D4Z4 pairs, repeats or contraction size.

Detailed outcomes of the topic “Economic Burden” are reported in **Table 36** in **Appendix 7.6**. Sample sizes across the studies ranged from 16 to 690. Outcomes reported were heterogenous. Per patient annual direct costs identified in the literature ranged from €9,270 (2010)^151^ to €21,256 (2018)^150^. Per patient annual societal costs ranged from €26,240 (2009)^152^ to €40,850 (2018).^153^ Konersman *et al.* reported that annual medical costs increased post-FSHD diagnosis; this increase post-diagnosis was much higher than for other diseases post-diagnosis, including connective tissues diseases.^154^ In a second conference abstract by Konersman *et al.*, compared to matched controls, individuals with FSHD had higher costs and total number of prescriptions. Blokhuis *et al.* reported that total mean annual costs increased by mobility level ranging from €14,008 (2018) without any mobility aids to €63,765 (2018) among those who were unable to walk; further, the per-patient direct costs of illness were reported to be approximately five times higher than the mean per-capita health expenditure in the Netherlands.^150^

Healthcare resource use was reported in multiple studies. Up to 41% of patients^116^ reported use of ankle braces, leg braces, canes or walkers and up to 58.2%^155^ respondents noted that they used a wheelchair at some point during the day; full-time wheelchair was reported in up to 25.5%^127^ of FSHD individuals. Statland *et al.* found that among 343 individuals with FSHD who were device-free at baseline, after a mean follow-up of 6 years (range 1 – 9 years), 11% used an ankle brace, leg brace, cane or walker and 22% used a wheelchair at some point during the day.^116^ A second study by Statland *et al.* found that the use of wheelchairs, ankle braces, canes, walkers, breathing machines and hearing aids was higher for patients with large D4Z4 contractions (≤18 kb) versus medium or small contractions.^149^ Blokhius *et al.* reported that in the preceding 3 months, 58%, 53% and 26% of FSHD patients had a physical therapy, general practitioner or rehabilitation physician visit, respectively.^150^

Employment among individuals with FSHD and the impact of FSHD on employment was reported in 4 studies. Among 574 individuals with FSHD, Hilbert *et al.* found that 50% of all adult members reported reduced employment opportunities.^155^ Fowler *et al.* reported that among the 16 individuals with FSHD included in their study, 9 were employed; of the 7 unemployed, 6 had been employed in the past.^156^ In a study in the Netherlands, among employed individuals with FSHD, 12% reported absenteeism in the past 4 weeks, with a median absence duration of 152 days.^150^ One study reported that respondents taking 6 or more medications per day were significantly more likely to be unemployed (75.4%) than those taking 0 – 5 medications (41.9%) (p<0.001).^157^

#### 3.6 Disease Classification

A total of 28 full text articles and 15 conference abstracts were identified for the topic “Disease Classification”. An overview of the studies included for the topic “Disease Classification” is provided in **Table 37** in **Appendix 7.7,** noting that not all information was available for all studies. Among all identified studies, the majority were observational in design (n=36).

Among the 43 studies included, 10 studies exclusively assessed patients with FSHD Type 1, one study exclusively assessed patients with FSHD Type 2, seven studies assessed a mix of FSHD Type 1 and 2, and the remaining did not specify the FSHD type. A total of 5 studies exclusively assessed pediatric patients, while 11 did not report explicitly whether pediatric patients were included; the remaining studies were conducted exclusively in adults.

Definition and phenotype characteristics varied across the studies and included classification by the Comprehensive Clinical Evaluation Form (CCEF), onset (early, typical, atypical), type of FSHD (1, 2, 1 and 2), subtype (classical, facial sparing, early upper limb weakness), number of D4Z4 repeats, among others (**Table 6**).

Multiple studies used the CCEF to define the various clinical categories by a combination of different features to define categories A through D. The CCEF was proposed by the Italian Clinical Network for FSHD and defines nine clinical categories, designed to capture the clinical diversity of FSHD. In addition to detailed descriptions of the CCEF, CCEF categories were linked to disease classification by onset (typical versus atypical) by Alobaidan *et al.^158^* Vercelli *et al.* explored change in score over time among patients and carrier relatives by CCEF, and found that the highest change in FSHD clinical scores was in subjects categorized as CCEF A.^159^

D4Z4 repeats were also used to describe classification of disease. Statland *et al.* observed that patients with 1 – 6 repeats were more severely affected than those with 7 – 10 repeats.^160^ Banerji *et al.* noted that shorter D4Z4 repeat length was associated with earlier onset^161^; Alavi *et al.* noted that patients with lower number of D4Z4 repeats showed earlier age at onset.^162^ Ruggiero *et al.* found that among patients with 7 – 8 D4Z4 repeats, only half displayed a classical FSHD phenotype, whereas the other half presented incomplete or atypical phenotypes.^163^ Both Rieken *et al.* and Sacconi *et al.* found that the overlap between Type 1 and Type 2 FSHD patients for D4Z4 repeat units suggests a disease continuum and that the previously established repeat size threshold for Type 1 FSHD (1 – 10 repeat units) and Type 2 FSHD (11 – 20 repeat units) needs to be reconsidered.^164,165^ Mul *et al.* reported that D4Z4 repeat array size explained approximately 29% and 15% of the variability in the involvement of facial muscles and upper extremity muscles, respectively.^69^

Other disease classifications and/or description of phenotype characteristics included distinguishing patients with Type 1 and/or Type 2 FSHD^166-169^; by EcoRI fragments^170^; FSHD by facial-sparing versus facial and shoulder weakness^171,172^; defining early onset^122,124,173-183^ and early versus classic onset^184,185^; CSS less than or greater or equal then 3.5^185^; sporadic FSHD^186^; the focal phenotype^187^; and mosaic FSHD^188^.

Table 6. Classification of disease types and phenotypes in FSHD

| **Author, year** | **Disease class** | **Prevalence, n (%)** | **Definition and phenotype characteristics** |
| --- | --- | --- | --- |
| CONFERENCE ABSTRACTS | | | |
| Alobaidan 2021^158^ | Typical onset | 82 (66%) | CCEF category A and B: facial weakness and/or impairment of upper arm abduction with winged scapula in addition to the absence of uncommon features. |
|  | Atypical onset | 43 (34%) | CCEF category D: presence of uncommon features.  Atypical cases had an older age of onset compared to typical FSHD (31 versus 20 years, p<0.001) and were more frequently sporadic. On average, typical cases were diagnosed 2.2 months (1–6) and atypical ones 9.5 months (1–108) after first assessment in neuromuscular clinic. 11 out of 43 atypical cases (25.6%) were initially misdiagnosed, the majority as inflammatory myopathy. |
| Banerji 2021^161^ | bFacial-sparing subtype | --- | Short D4Z4 repeat length was associated with earlier onset of all muscle weakness symptoms.  Smoking was associated with earlier onset of both facial and lower body muscle weakness symptoms.  Hypertension was associated with a slower onset of facial weakness.  Clustering on integrated data describing 213 UK and 171 US FSHD1 patients and adjusting for registry membership revealed 7 distinct clinical presentations of FSHD1 (not further described). |
| Bettio 2021^189^ | 1–3 D4Z4 repeats | NR (15%) | A more homogenous FSHD phenotype was observed among cases with 1–3 D4Z4 reduced allele, although a significant percentage of them (21.8%) showed atypical features. |
|  | 4–8 D4Z4 repeats | NR (71%) | Among index cases with larger D4Z4 repeats allele, 56.4% of them displayed a classical FSHD phenotype with facial and scapular muscle weakness, whereas 13% presented incomplete phenotype with facial weakness or scapular girdle involvement. 24.3% of index cases with 4–10 D4Z4 allele showed more complex phenotypes with atypical clinical features.  Carriers of D4Z4 reduced alleles and their families show distinct nosological entities with different disease courses and lower penetrance. Introduction of a new term, D4Z4-pathies, would better define the clinical variability of disease. |
|  | 9–10 D4Z4 repeats | NR (14%) |  |
| Chen 2010^173^ | Early onset | --- | iFSHD with very short 4q35 fragments (≤14 kb) and high prevalence of auditory, ophthalmologic, and cerebral involvements. Additional rare diverse features were present, such as arrhythmogenic abnormalities, dysphagia, kyphoscoliosis, and asymmetric muscular atrophies. |
| Duong 2015^174^ | Early onset | --- | iFSHD characterized by severe phenotype, with prominent proximal weakness and respiratory insufficiency. |
| Liu 2019^190^ | CCEF A | A1: 16 (30%)  A2: 22 (41%)  A3: 4 (7%) | CCEF category A (subcategories A1-A3): patients presenting facial and scapular girdle muscle weakness typical of FSHD (Ricci *et al.*).  Patients in the A1 group were younger, with a lower mean age at onset, shorter disease duration, smaller D4Z4 repeat number, and more severe clinical disability in FSHD clinical score, MMT, and MFM. |
|  | CCEF B | B1: 1 (2%)  B2: 1 (2%) | --- |
|  | CCEF D1 | 10 (18%) | CCEF category D (subcategories D1, D2): patients with myopathic phenotype presenting clinical features not consistent with FSHD canonical phenotype (Ricci *et al.*).  Six D1 patients displayed pes cavus (pathological foot condition). |
| Mah 2013^175^ | Early onset | <2% | iFSHD characterized severe phenotype. |
| Mah 2014^176^ | Early onset | --- | Onset of facial and shoulder weakness <10 years of age.  Smaller EcoRI fragment size was associated with great disease severity; the EcoRI fragment size had a strong negative correlation with Brooke scale (r=-0.80), Vignos scale (r=-0.67), and CSS (r=-0.72 for facial, -0.64 for shoulder, and -0.60 for pelvic muscle weakness). |
| Mele 2019^191^ | Facial-sparing subtype | 10% | Definition of facial-sparing subtype not provided. The group included patients with this phenotype regardless the size of the D4Z4 contracted allele. |
| Mongini 2015^192^ | CCEF A | --- | Subjects with typical FSHD phenotype. |
|  | CCEF B | --- | Subjects with incomplete FSHD phenotype. |
|  | CCEF C | --- | Asymptomatic subjects. |
|  | CCEF D | --- | Myopathic subjects without typic FSHD features. |
| Mul^69^ | D4Z4 repeat array size | 40% | The variability in disease severity by familial factors was approximately 40%. Asymptomatic gene carriers had longer repeat sizes compared to symptomatic individuals (7.3 vs 6.2 units, p=0.001). D4Z4 repeat array size explained approximately 29% and 15% of the variability in the involvement of facial muscles and upper extremity muscles, respectively. |
| Ricci 2021^193^ | CCEF B1 | 125 (27%) | CCEF category B (subcategories B1, B2): subjects with muscle weakness limited to scapular girdle or facial muscles (Ricci *et al.*).  Patients with facial-sparing phenotype (B1 clinical category) have a significantly milder phenoType 1n comparison with those presenting classic FSHD phenotype (p<0.001). |
| Ruggiero 2017^194^ | CCEF A | A1: 2% patients, 0% relatives  A2: 29% patients, 7% relatives  A3: 24% patients, 8% relatives | Definition of different categories not provided.  The majority of probands carrying 33-35 D4Z4 reduced alleles (53%) had moderate phenotype with partial or mild facial involvement in them (A2-A3).  40% showed a prevalent shoulder girdle impairment (B1) or atypical phenotype (D1) and 3% displayed clinical features inconsistent with the clinical diagnosis of FSHD (D2).  The majority of relatives were non penetrant carriers (55%) and only few of them showed typical phenotype (17%) while the remaining had atypical phenotype.  Overall, among 377 carriers of 33-35 D4Z4 reduced allele, 119 (31.6%) presented a classical FSHD phenotype (categories A), 124 (32.9%) were asymptomatic (categories C), 78 (20.6%) had incomplete phenotypes (categories B), 56 (14.9%) showed atypical phenotypes (categories D). |
|  | CCEF B | B1: 20% patients, 9% relatives  B2: 1% patients, 12% relatives |  |
|  | CCEF C | C1: 1% patients, 15% relatives  C2: 0% patients, 40% relatives |  |
|  | CCEF D | D1: 20% patients, 6% relatives  D2: 3% patients 3% relatives |  |
| Ruggiero 2019^163^ | 7–8 D4Z4 repeats | NR | Only half of patients displayed a classical FSHD phenotype, whereas the other half present incomplete or atypical phenotype. Carries of 7 – 8 D4Z4 repeats constitute a genetic subgroup different from classic FSHD. |
| Ruggiero 2021^195^ | CCEF D1 | 61.3% of relatives | 61.3% of relatives of D1 patients (on the basis of CCEF) were asymptomatic (category C). The probability for relatives of D1 probands to develop a myopathic phenotype 1s low. |
| Tabata 2021^184^ | Early onset | 12 (38%) | Onset of facial muscle weakness before age of 5 as well as onset of scapular muscle weakness before age of 10.  Major changes occur from second decade of age in patients with early-onset FSHD, indicating the importance of intensifying the medical follow-up before they become teenagers. |
|  | Classic onset | 20 (62%) | Not meeting criteria for early onset. |
| FULL TEXT ARTICLES | | | |
| Alavi 2018^162^ | Sporadic onset | 22 (47%) | Absence of familial history of FSHD.  Sporadic cases had a greater disease severity than familial cases (CSS: 9.3 ± 5.7 versus 6.5 ± 4.2, p=0.04). |
|  | Familial onset | 23 (54%) | Presence of familial history of FSHD. |
|  | Number of D4Z4 repeats | --- | Patients with lower number of D4Z4 repeats showed earlier age at onset than patients with larger repeat numbers (r=-0.56; p=0.003). Patients with 1-3 repeats presented with severe or moderate disease; patients with 4-7 repeats tended to have mild or moderate disease and variable manifestations. |
| Banerji 2020^172^ | Facial-sparing subtype | 24% | Included three facial-sparing phenotypes: mild presentation (5%) with later facial and periscapular involvement; an early shoulder presentation (10%) with accelerated periscapular weakness; and an early foot presentation (9%) with accelerated foot dorsiflexor weakness.  Rate of disease progression was inversely proportional to D4Z4 repeat length. |
|  | Classical presentation | 74% | Weakness of the facial muscles as the first manifestation. |
| Brouwer 1994^177^ | Early onset | 6 (4%) | Signs or symptoms of facial weakness before the age of 5 years and signs or symptoms of shoulder girdle weakness before the age of 10 years. |
| Dijkstra 2021^122^ | Early onset | 8 (44%) | Compared to classic-onset patients, those with early-onset FSHD had a shorter number of units within the pathogenic D4Z4 repeat, were significantly younger, had higher age-adjusted CSS (p=0.02), and showed a bigger mean echogenicity change (2.0 ± 1.4 versus 0.1 ± 0.5, p=0.02).  Motor functioning after 2 years as measured by MRC sum score, FSHD clinical score, CSS, and 6MWT did not differ significantly between these subgroups. No significant difference in disease progression was found between the 2 onset types. The presence of pain, fatigue, and systemic features did not vary significantly between the 2 groups, although hearing abnormalities were observed only in the children with early-onset FSHD who had a D4Z4 repeat size of 2 units. |
| Dorobek 2015^178^ | Early onset | 21% | Facial weakness before the age of 5 and signs of shoulder weakness by the age of 10 (Brouwer's criteria).  The frequency of early-onset FSHD was 22/103 cases (21.4%) (the disease was present in 18/69 of the examined families). |
| Eren 2021^73^ | --- | --- | Six stages of the disease were defined to create a system evaluating the shoulder disability consisting of shoulder elevation, deltoid function, and scapular winging, with an excellent inter-observer agreement on FSHD patients’ shoulder disability. This would provide surgeons a beneficial tool to define patient groups that would have negatively or positively been affected by scapulothoracic arthrodesis. |
| Evangelista 2016^169^ | FSHD1 | 92% | Contraction of the D4Z4 macrosatellite on chromosome 4qA of less than 38 kb. |
|  | FSHD2 | 2% | Presence of *SMCHD1* mutation with a permissive 4qA allele or hypomethylation. |
| Felice 2005^170^ | Borderline EcoRI fragments | 11 (27%) | DNA-confirmed FSHD patients harboring borderline EcoRI fragments ≥35 kb.  The group with borderline EcoRI fragments showed statistically significant less severe disability and weakness (CSS and total MRC score) than the group with EcoRI fragments <35 kb. |
|  | EcoRI fragments <35 kb | 30 (73%) | DNA-confirmed FSHD patients harboring EcoRI fragments <35 kb. |
| Giacomucci 2020^168^ | FSHD2 | --- | The most affected and spared muscles were similar to those detected in FSHD1, although differences regarding the frequencies and severity of involvement were present, with significant differences between FSHD1 and FSHD2 patients when the involvement of at least one trapezius, one abdominal muscle, one hamstring muscle and bilaterial iliopsoas and subscapularis sparing are prevalent via MRI (p=0.002). |
| Goselink 2018a^124^ | Early onset | 9 (30%) | Facial weakness with otherwise normal muscle strength and performance, limited functional exercise capacity, lumbar hyperlordosis, decreased quality of physical well-being, and presence of fatigue and pain, with rarely reported systemic features.  Compared to the classic-onset group, early-onset FSHD patients had a shorter repeat length (3.9 versus 5.8 number of units within the pathogenic D4Z4 repeat, p=0.03), a higher FSHD clinical score (3.7 versus 1.8, p=0.03), and a higher quantitative muscle ultrasound score (3.2 versus 0.8, p<0.005). The 6MWT, MFM, and MRC sum score did not differ significantly between early and classic-onset groups. |
|  | Classic onset | 17 (57%) | --- |
| Goselink 2018b^179^ | Early onset | --- | Facial weakness before the age of 5 and shoulder gridle weakness before the age of 10.  Early-onset is more severe than classical FSHD with a high proportion of patients being wheelchair-bound at a young age and more systemic features. Correlation between the D4Z4 repeat length and CSS at follow-up (p=0.009). |
| Goselink 2019^179^ | Early onset | 28 (34%) | Signs and symptoms of facial weakness before the age of 5 and of shoulder gridle weakness before the age of 10.  The patients with early-onset FSHD had more severe muscle weakness defined by the FSHD clinical score, the MRC sum score, and the percentage of wheelchair dependency.  The mean number of D4Z4 repeat units of the disease allele in the early-onset patients was significantly lower than in the classic-onset patients (3.6 versus 6.2 units). |
|  | Classic onset | 55 (66%) | --- |
| Guien 2018^167^ | FSHD1 | 95% | Contraction to 10 repeats or less of the 3.3 kb D4Z4 repeat units on chromosome 4. |
|  | FSHD2 | 5% | Presence of mutations in the *SMCHD1* gene or, in rare cases, pathogenic mutations in the *DNMT3B* gene. |
| Hassan 2012^187^ | Focal phenotype | 7 (5%) | Focal or monomelic presentations of FSHD. |
| Korf 1985^181^ | Early onset | --- | Early onset added to progressive and severe FSHD, with facial diplegia and sensorineural hearing loss.  Each patient was noted to have bilateral facial weakness early in life, with sparing of extraocular movements and most trigeminal muscles but involvement of the tongue and palate. Through early childhood, weakness developed in the muscles of the neck, shoulder, and hip girdles, with winging of scapulae and extensive lumbar lordosis. Most patients had become non-ambulatory by adolescence. In each a bilateral sensorineural hearing loss developed in the first decade, but none of the patients had any visual problems. |
| Mah 2018^182^ | Early onset | --- | Facial weakness before 5 years of age and/or shoulder girdle weakness before 10 years of age.  When adjusted for enrollment age, sex, and D4Z4 repeats, younger age at onset of facial weakness was associated with greater CSS, slower velocities in timed function tests, and lower MMT scores (p<0.05). There were no associations between age at onset of shoulder weakness and either disease severity or motor performance. |
| Mair 2017^166^ | FSHD1 | 45% | Definition not provided. Involvement of leg muscles appeared to be independent of FSHD type (FSHD1 and FSHD2). |
|  | FSHD2 | 55% | Mutation in the *SMCHD1* gene and severe hypomethylation of the D4Z4 fragment |
| Moreira 2017^185^ | Early onset | 6 (6%) | Early onset was defined according to Brouwer’s criteria.  The early-onset group had 7.7 times higher probability of becoming wheelchair dependent (odds ratio 95% CI [1.3, 45.3], p=0.025). Distribution of severity scores in the early-onset and classic-onset groups was significantly different (p=0.021). |
|  | Classic onset | 82 (82%) | Classic onset was defined according to Brouwer’s criteria.  In patients with D4Z4 fragments <18 kb, a significant linear correlation was found between the fragment size and both age of onset (r=0.737, p=0.01) and severity scores (r=-0.838, p<0.01). |
|  | Mildly affected | 46 (46%) | CSS <3.5.  There was a positive correlation between the duration of symptoms and the CSS (r=0.603, p=0.001). |
|  | Moderately / severely affected | 54 (54%) | CSS ≥3.5. |
| Qiu 2020^188^ | Mosaic FSHD | 17 (50%) | The mosaic FSHD cohort exhibited significant clinical heterogeneity and relatively slight disease severity compared to the non-mosaic FSHD group. Both genotypic severity scale and D4Z4 hypomethylation status served as modifiers of clinical phenotypes. |
| Ricci 2016^196^ | CCEF A | --- | Includes subcategories A1-A3: subjects presenting facial and scapular girdle muscle weakness typical of FSHD. |
|  | CCEF B | --- | Includes subcategories B1 and B2: subjects with muscle weakness limited to scapular girdle or facial muscles. |
|  | CCEF C | --- | Includes subcategories C1 and C2: symptomatic/healthy subjects. |
|  | CCEF D | --- | Subjects with myopathic phenotype presenting clinical features not consistent with FSHD canonical phenotype. |
| Ricci 2019^171^ | Facial-sparing subtype | 155 (24%) | Shoulder muscle impairment with facial sparing.  The facial-sparing group had a significantly higher mean age at onset (33.1 ± 16.1 years, p<0.0001, t-test versus classical group); only 6 of 155 (3.9%) patients were full-time wheelchair users (mean age of loss of ambulation, 60.2 ± 14.9 years); a high percentage (45%) of patients reported a negative family history for muscle disease. |
|  | Classical presentation | 437 (68%) | Facial and shoulder weakness.  Mean age at onset was 18.0 ± 13.4 years, and loss of ambulation was reported in 83 patients (19%; mean age of loss of ambulation, 38.5 ± 18.1 years). 144 (33%) patients recorded a negative family history for functional muscle impairment. |
| Rieken 2021^165^ | FSHD1 | 664 (94%) | 4q35A allele with EcoRI fragment ≤40 kb (1–10 D4Z4 repeats) or 4q35 allele with EcoRI fragment ≤40 kb (1–10 D4Z4 repeats) but haplotyping not requested. Median (IQR) D4Z4 size 6 (4 – 7). Majority (48.2%) had 4 – 6 repeats. FSHD1 accounts for 94.5% of genetically confirmed cases of FSHD. Data show a continuum of D4Z4 repeat numbers with FSHD1 samples having the fewest, FSHD2 an intermediate number and non-FSHD1/2, the most. |
|  | FSHD2 | 39 (6%) | Permissive 4q35A allele plus hypomethylation and presence of a pathogenic *SMCHD1* variant. Median (IQR) D4Z4 size 15 (12 -22). Majority (51.3%) had 11 – 15 repeats. |
|  | Borderline FSHD1 | --- | 4q35 allele with EcoRI fragment 41–43 kb (11 D4Z4 repeats) and presence of permissive 4q35A allele or haplotyping not requested.  A total of 20 cases with borderline FSHD1 were detected out of 1,594 tests performed. These cases were not part of the 703 cases with confirmed FSHD diagnosis. These 20 cases had an average EcoRI fragment size of 42.3 ± 0.5 kb. The average methylation value of the cases tested was 37.4 (n=15). Five of the borderline cases had a methylation level ≤28% resulting in *SMCHD1* sequencing, but none of these 5 cases harbored a pathogenic *SMCHD1* variant. |
|  | FSHD1 + FSHD2 | --- | 4q35A allele with EcoRI fragment 33–43 kb (8–11 D4Z4 repeats) plus hypomethylation and presence of *SMCHD1* variant.  A total of 4 cases were detected out of 1594 tests performed. These cases are not part of the 703 cases with confirmed FSHD diagnosis. In these 4 cases, the average EcoRI fragment size for the smallest 4q35A alleles was 40.8 kb (38–42) and the average methylation value was 13.3% (10–15). |
| Sacconi 2019^164^ | FSHD1 | 57 (55%) | 4–10 D4Z4 repeat units on 4qA.  Within this group, size of the D4Z4 repeat inversely correlated with disease severity: patients with 8–10 repeat units had a milder disease than those with 4–7 repeat units. |
|  | FSHD2 | 20 (19%) | ≥11 D4Z4 repeat units on a 4qA haplotype and presence of a pathogenic *SMCHD1* variant.  This group of patients are significantly more severely affected by all clinical scales when compared to patients with FSHD1 with upper-sized FSHD1 alleles (8–10 repeat units). |
|  | FSHD1 + FSHD2 | 7 (7%) | 9–10 D4Z4 repeat units on 4qA and presence of a pathogenic *SMCHD1* variant.  The overlap between FSHD1 and FSHD2 patients in the 9–10 D4Z4 repeat unit range suggests that FSHD1 and FSHD2 form a disease continuum. The previously established repeat size threshold for FSHD1 (1–10 repeat units) and FSHD2 (11–20 repeat units) needs to be reconsidered. |
| Statland 2015^160^ | 1–6 D4Z4 repeats | 46 (62%) | Patients with 1–6 repeats were more severely affected than those with 7–10 repeats. |
|  | 7–10 D4Z4 repeats | 28 (38%) | Compared to participants with 1–6 repeats, patients with 7–10 repeats were diagnosed 6.6 years older (p=0.17); had lower CpG methylation than would be predicted by D4Z4 repeat size (p=0.04); had age-adjusted clinical severity 39.8 points lower (p=0.004); had muscle pathology grades that were 2.4 points less severe (p<0.0001); had quantitative myometry 28.3% predicted of normal higher (p=0.002); had global manual muscle testing scores 0.6 higher (p=0.005); and did not require wheelchairs. In conclusion, participants with 7–10 residual D4Z4 repeats were more mildly affected than those with 1–6 repeats. |
| Steel 2019^183^ | Early onset | --- | Age at onset correlated with length of the Bln1-resistant fragment (r=0.81). |
| Tawil 1996^186^ | --- | --- | In the 7 sporadic cases, identified disease severity was highly correlated with fragment size (r=0.92, p=0.0036).  The mean rate of progression of FSHD patients with maternally and paternally inherited FSHD was -0.134 ± 0.111 and -0.172 ± 0.174, respectively; it was not significantly different (p=0.44).  When the offsprings’ QMT values were corrected, in 15 of 23, the offspring are more severely affected, in 7 of 23, the parent is more severely affected, and in 1 of 23 both are equally affected. The difference between the mean QMT score of the parents (4.71 ± 4.71) and corrected QMT score of the offspring (7.30 ± 4.96) is significant (p=0.011).  The mean number of offspring for FSHD males (2.17 ± 1.19) was significantly smaller than that for FSHD females (3.28 ± 1.82, p=0.01). The relative fitness of FSHD males (2.17/3.28) is therefore 0.66 when compared with FSHD females. |
| Vercelli 2021^159^ | Minimally affected | 13% patients, 30% carrier relatives | FSHD clinical score 1-2.  The increase in the FSHD clinical score observed at the end of follow-up period in the cohort was 1.3 (1.1, 1.4) (mean FSHD clinical score at baseline versus follow-up: 4.4 ± 3.8 versus 5.7 ± 4.3). The overall median ΔFSHD clinical score was 1 (0–7). ΔFSHD clinical score of all patients did not significantly vary on the basis of the D4Z4 reduced allele size. The highest ΔFSHD score (1.7) was found in a category A CCEF subject, whereas subjects with incomplete phenotypes (category B) had lower ΔFSHD score (0.6). |
|  | Moderately affected | 53% patients, 24% carrier relatives | FSHD clinical score 3-7. |
|  | Severely affected | 33% patients, 2% carrier relatives | FSHD clinical score 8-15. |
| Yasukohchi 1988^197^ | Nosological | 1 (50%) | Sensorineural hearing loss and tortuosity of retinal arterioles.  The patient also exhibited early onset and rapid aggravation of severe restrictive-type respiratory failure and cor pulmonale, leading to death. |
|  | Mild | 1 (50%) | Mild course FSHD: No involvement of any other organ than muscles at the time of diagnosis |

6MWT, 6-minute walking test; CCEF, Comprehensive Clinical Evaluation Form; CI, confidence interval ; CSS, clinical severity score; DNA, deoxyribonucleic acid; FSHD, facioscapulohumeral muscular dystrophy; iFSHD, infantile FSHD; MFM, motor function measure; MMT, manual muscle testing; MRC, Medical Research Council ; NR, not reported; QMT, quantitative muscle testing ; SMCHD1, structural maintenance of chromosomes flexible hinge domain containing 1; UK, United Kingdom ; US, United States.

#### 3.7 Diagnosis

The topic “Diagnosis” included 17 full text articles and 7 conference abstracts (**Table 32** in **Appendix 7.8**). Of note, some studies included in the topic “Disease Classification” also reported on “Diagnosis” and thus are included here.

Of those reporting the study design, most were cohort studies (prospective: n=7 and retrospective: n=7). Not all included studies reported the FSHD population type: most studies were conducted in FSHD populations not specified (n=15), followed by Type 1 FSHD (n=3) or a mix of Type 1 and Type 2 (n=2). Sample sizes ranged from 1 to 1,074. Two studies were conducted exclusively in pediatric patients (though one was a case study)^183,198^, with three others that included pediatric patients^188,199,200^. Few studies reported additional population characteristics.

Multiple diagnostic techniques for FSHD were reported in the literature (**Table 7**).

The Southern blotting technique was the most reported technique, though some publications reporting on this diagnostic technique tended to be older (1997 – 2012)^201-204^. The concordance of Southern blotting with polymerase chain reaction (PCR) was reported in three studies. Goto *et al.* found that 83% of those identified with Southern blotting were also identified using a long-range PCR.^204^ Skoblov *et al.* found concordance with Southern blotting in 4 (57%) of FSHD patients.^205^ Zernov *et al.* using quantitative PCR found that in 96% of the cases, qPCR-based results were concordant with Southern blotting^206^.

Nicolau *et al.* set out definitions for different diagnostic techniques for Type 1 and Type 2 FSHD, including the Southern blotting, haplotyping and methylation assay plus sequencing; however, they did not compare diagnostic techniques between each other.^207^ Similarly, Rieken *et al.* described Southern blotting, methylation assay and sequencing for diagnosing FSHD patients.^165^

Pulsed field gel electrophoresis (PFGE) was used to identify FSHD in two studies. In Orrell *et al.*, paired digests of genomic DNA followed by PGFE was described, though the type of FSHD was not specified.^200^ In Qiu *et al.*, PFGE-based Southern blotting was used to describe a mosaic FSHD patient.^188^

Next generations sequencing (NGS) was used to identify *SMCHD1* mutations by Larsen *et al.*^208^; two additional publications by Larsen *et al.*^209,210^ confirmed the use of NGS for diagnosis.

Muscle MRI was described in three studies to diagnose FSHD among NMD patients. Storch *et al.* described the use of muscle MRI to help direct genetic testing in undiagnosed patients.^211^ Fabry *et al.* used muscle MRI plus a deep learning tool (versus a radiologist) to differentiate Type 1 FSHD from myositis.^212^ Monforte *et al.* also used machine learning in combination with muscle MRI to improve the accuracy of diagnosis among atypical FSHD patients.^213^ Quantitative motor unit potential was validated as a diagnostic technique for FSHD patients by Podnar *et al.* in the biceps brachii muscles; the technique was also carried out in controls for comparison.^214^

Other techniques described in single studies were identified. These included genome optical sequencing, as described by Guruji *et al.,* to identify FSHD haplotype and D4Z4 repeat number.^199^ Bakker *et al.* described the use of DNA testing to identify FSHD in families and among sporadic cases.^215^ Genetic testing, as described by Parisien *et al.,* was used to identify a patient with FSHD and describe the location of the expression of the D4Z4 gene in the muscle causing FSHD.^198^ DNA digestion using the bacterial restriction enzyme Bln1 was used by Steel *et al.* to describe the symptoms and signs of Type 1 FSHD.^183^

Finally, two studies provided insights on establishing a diagnosis. Padberg *et al.* outlined the four criteria which define FSHD; it should be noted that this study was the oldest of the studies included for the topic “Diagnosis” and thus may be less relevant in light of more recent diagnostic techniques.^216^ While not a description of a technique, Rodrigues *et al.* used data from a registry and noted that among patients with FSHD, only 50% had a molecular test result confirming diagnosis; the remaining patients were diagnosed on clinical basis only.^217^

Table 7. Diagnostic techniques for FSHD

| **Author, year** | **Technique** | **Diagnosis: n (%)** | **Description** |
| --- | --- | --- | --- |
| CONFERENCE ABSTRACTS | | | |
| Guruju 2022^199^ | Genome optical mapping | FSHD1: 96 (48%) | Genome optical mapping was performed using the Bionano Genomics Saphyr with subsequent analysis by Bionano Enfocus FSHD analysis software (Bionano, San Diego, CA) to identify FSHD haplotype and D4Z4 repeat number. Molecules aligning the D4Z4 repeat regions on chromosome 4 using human genome reference build GRCh38 are distinguished from regions of high homology on chromosome 10. The permissive (4qA) and non-permissive alleles (4qB) were assigned using the dynamic-programming algorithm included in the Enfocus FSHD analysis pipeline. The D4Z4 repeat size is determined based upon the measurement of the interval distance between labels flanking the D4Z4 arrays.  Among 196 patients tested, a D4Z4 contraction repeat (1-10) on the permissive haplotype (4qA) was observed in 96 individuals, 62 males (64.5%) and 32 females (35.5%), which is consistent with a clinical diagnosis of FSHD1. |
| Larsen 2014a^208^ | NGS | *SMCHD1* mutations:  9 (9%) | *SMCHD1* mutations (deletions, missense, splice site and nonsense) were identified in 9 patients without D4Z4 contractions.  *SMCHD1* mutations were also found in 2 patients with moderately contracted D4Z4 alleles thus suffering from FSHD1 and 2. |
|  | Pyrosequencing | *---* | Assay to determine methylation status of the D4Z4 repeat array. Significantly lower methylation levels for FSHD2 patients were found compared to healthy controls and FSHD1 patients. |
| Larsen 2014b^209^ | NGS | *SMCHD1* mutations:  NR (9%) | Seven novel and one reported mutation in *SMCHD1* were identified, ranging from missense mutations (p.Gly478Glu, p.Arg1449Lys, p.Val615Asp, p.Gln1463Pro, p.Pro1485Leu) and deletions of the 3' splice site (p.Val1093Leufs∗26, p.Lys1092del) to the first nonsense mutation (p.Leu1663∗).  *SMCHD1* mutations were found in 9.4% of the patients analyzed.  In addition to the mutation in *SMCHD1*, two of the patients showed a contracted D4Z4 allele and therefore have a double trouble situation for FSHD1 and 2.  When comparing the phenotype of the patients, all FSHD2 patients were mildly affected while patients with FSHD1 plus 2 were much more severely affected than expected from the number of repeat units (10 units) on the contracted allele. |
|  | Sanger sequencing | *SMCHD1* mutations:  NR (100%) | All mutations were confirmed by Sanger sequencing. |
|  | Bioinformatics tools of Alamut | *SMCHD1* mutations:  NR (100%) | All mutations were predicted as pathogenic by the bioinformatics tools of Alamut (interactive bio-software). |
|  | Pyrosequencing | D4Z4 methylation: FSHD1: 14-55%  FSHD2: 5-19%  Healthy controls: 30-65% | The methylation status of the D4Z4 repeat array for FSHD2 patients (5-19%) was found to be significantly lower than for healthy controls (30-65%) and for FSHD1 patients (14-55%). |
| Skoblov 2019^205^ | PCR | Concordance with Southern blotting: 4(57%) | The PCR-based diagnostic was applied to 7 FSHD patients with one 4qA repeat below 10 units. |
| Storch 2017^211^ | Muscle MRI | FSHD NS: 1 | In 61 NMD patients, a muscle MRI was performed in undiagnosed patients to help direct genetic testing. In 24 patients, the suspected diagnosis based on muscle MRI was confirmed genetically, including 1 patient with FSHD. |
| Walker 2022^218^ | Genetic confirmation | FSHD1: NR (46%) | --- |
| Zernov 2020^206^ | qPCR | 36 (56%) | In 96% of the cases, qPCR-based results were concordant with the Southern blotting and molecular combing references. In 69% of the confirmed cases, pathogenic alleles had between 3-6 D4Z4 repeats.  We defined 13 carriers in a group of 39 healthy relatives (33.3%). Among them, 76.9% of cases had the permissive allele with 6-8 D4Z4 repeats. We observed enrichment of the permissive alleles in the range of 3-6 D4Z4 units among affected individuals, whereas among carriers permissive alleles were enriched in the range 6-8 D4Z4 units. |
| FULL TEXT ARTICLES | | | |
| Bakker 1996^215^ | DNA testing | FSHD NS:  27 (81%) families  25 (44%) sporadic cases | In 27 of the 33 families, a p13E-11 EcoRI fragment smaller than 35 kb was observed in the DNA of the index patient; the size of the fragments ranged from 11 to 34 kb.  In 25 out of the 57 sporadic cases, p13E-11 fragments were detected between 11 kb and 35 kb. |
| Fabry 2022^212^ | Muscle MRI + deep learning | --- | The deep learning tool was able to differentiate FSHD1 from myositis with a correct classification percentage respectively of 69% (39, 99), 75% (48, 100) and 77% (60, 94) when thigh only, thigh and calf or the thigh, calf, and shoulder MR images were analyzed.  The percentages of correct classification of the two radiologists for these later MRI images were 38/40 (95%) and 35/40 (87.5%), respectively; with no differences with deep learning tool correct classification (p=0.41 and p>0.99, respectively).  Among the 7 patients who were misclassified by the radiologists, the deep learning tool correctly classified 6 of them. |
| Goto 2006^204^ | Southern blotting | FSHD NS: 105 (100%) | Southern blotting with p13E-11 probe was used as standard. |
|  | Long range PCR | FSHD NS: 87 (83%) | PCR products were amplified in DNA samples from the 87 patients with an EcoRI fragment of ≤25 kb, while DNA from healthy individuals or the 18 patients with fragments ≥26 kb were not amplified/detected. |
| Larsen 2015^210^ | Haplotyping | Permissive haplotype 4qA161: 79 (83%) | 79 individuals were positive for the FSHD-permissive haplotype 4qA161 as determined by the SNP assay. |
|  | NGS | *SMCHD1* mutations:  11 (14%) | 11 patients had heterozygous variants in the *SMCHD1* gene by NGS. All were confirmed by Sanger sequencing and pathogenicity was assessed using the software package Alamut. |
|  | Pyrosequencing | D4Z4 methylation: FSHD1: 14-64%  FSHD2: 5-27%  Healthy controls: 30-65% | The methylation-sensitive Southern blot showed methylation levels of ≤25% for all tested patients with potentially pathogenic variants in *SMCHD1*.  For our pyrosequencing assay, patients were divided into four groups according to their diagnostic results: FSHD-like patients negative for FSHD1 and FSHD2, FSHD1 patients, FSHD2 patients and FSHD1+2 patients. In addition, we analyzed 48 healthy controls. The methylation levels in healthy individuals ranged from 30 to 65%, while FSHD2 patients had the lowest levels with 5–27% methylation. FSHD1 patients show methylation levels of 14–64%. The few patients with mutations in both FSHD loci (FSHD1+2) were ranging between 13 and 33% methylation. For FSHD-like patients (FSHD1 and 2 negative) a methylation of 31–74% was found, which is in the range of healthy controls. |
| Lemmers 2012^203^ | Southern blotting | --- | Genomic DNA is digested with restriction enzymes and sized by gel electrophoresis, followed by visualization using p13E-11 probe. Allow detection of 95% of FSHD1 cases. |
|  | Long range PCR | --- | PCR with specific primers and visualization by gel electrophoresis. Allow detection of 95% of Japanese cases, but not recommended for European cases. |
|  | Molecular combing | --- | Fluorescent-based sizing and visualization of D4Z4 fragments. |
|  | Marker analysis | --- | Detailed chromosomal analysis but does not allow to determine size of D4Z4 repeats. Mainly for rapid screening to determine the chromosome 4 haplotype, for prenatal diagnosis, and for preimplantation genetic diagnosis |
| Monforte 2022^213^ | Muscle MRI + random forest machine learning | Atypical FSHD: NR (91%) | The combination of trapezius involvement and bilateral subscapularis muscle sparing achieved the best diagnostic accuracy (0.89 [0.85, 0.92]) with 0.90 [0.85, 0.94] sensitivity and 0.88 [0.80, 0.93] specificity. This pattern correctly identified 91% atypical FSHD patients of our cohort.  The combination of trapezius involvement, bilateral subscapularis and iliopsoas sparing and asymmetric involvement of upper and lower-limb muscles was pathognomonic for FSHD, yielding a specificity of 0.99 [0.95, 1.00]. |
| Nicolau 2021^207^ | Southern blotting | --- | The diagnosis of patients with typical clinical phenotype and 1-10 D4Z4 repeats is FSHD1. |
|  | Haplotyping | --- | 4q35 haplotyping should be performed when the patient present atypical phenotype and 1-10 D4Z4 repeats. Presence of 4q35A confirms FSHD1. |
|  | Methylation assay + sequencing | --- | D4Z4 methylation and *SMCHD1* sequencing should be performed in addition to 4q35 haplotyping when the patient present typical phenotype and ≥11 D4Z4 repeats. Hypomethylation and presence of *SMCHD1* mutation confirms FSHD2.  In patients with reduced methylation, but lacking an *SMCHD1* mutation, sequencing of *DNMT3B* and *LRIF1* could be considered. |
| Orrell 1999^200^ | PFGE-based DNA digestion | FSHD NS: 82 (100%) | The technique consisted in paired digests of genomic DNA followed by pulsed field gel electrophoresis (PFGE). All patients with a clinical diagnosis of FSHD had a distinct reduction in the 3.3-kb D4Z4 repeat number, with a 4q35 EcoRI/p13E-11 allele ≤38 kb. |
| Padberg 1991^216^ | --- | --- | Four main criteria which define FSHD: (1) onset of the disease in facial or shoulder girdle muscles; sparing of the extra-ocular, paryngeal and lingual muscles and the myocardium; (2) facial weakness in more than 50% of the affected family members; (3) autosomal dominant inheritance in familial cases; (4) evidence of myopathic disease in muscle biopsy in at least one affected member without biopsy features specific to alternative diagnoses. |
| Parisien 2020^198^ | Genetic testing | FSHD NS: 1 (100%) | The patient possessed 3 copies of the 4,335 D4Z4 region located on chromosome 4. 3/4 ends of chromosomes 4 and 10 were found to possess the 4qA variant allowing for expression of the D4Z4 gene in muscle, causing FSHD. |
| Podnar 2009^214^ | Quantitative motor unit potential analysis | --- | For the biceps brachii muscles, sensitivity of the analysis was 59%, specificity 91%, the positive predictive value 85%, and negative predictive value 72% with at least two criteria (mean values or outliers for MUP thickness, amplitude and duration) below the reference intervals. In addition, all subjects with three abnormal EMG criteria were FSHD patients, and 90% of subjects with normal EMG were controls. |
| Qiu 2020^188^ | PFGE-based southern blotting | Mosaic FSHD: NR (100%) | The criterion for inclusion as a mosaic patient with FSHD was genetic diagnosis that presented ≥5 p13E-11 hybridizing fragments with at least one contracted 4qA-type D4Z4 array. |
| Rieken 2021^165^ | Southern blotting | FSHD1 and FSHD2:  703 (44%) | Of the 1,594 genetic tests performed via DNA digestion and southern blotting, 703 cases (44%) met criteria for a genetic diagnosis of FSHD. Of the total FSHD cases, 664 (94.5%) met criteria for a genetic diagnosis of FSHD1 and 39 cases (5.5%) met criteria for a genetic diagnosis of FSHD2. |
|  | Methylation assay | Hypomethylation:  98 (18%) | Of the 550 cases with methylation assays, 98 cases (18%) had hypomethylation, defined as ≤28%. The average methylation values for FSHD1, FSHD2, and non-FSHD1,2 cases were 34.7% (n=21), 13.5% (n=39), and 45.5% (n=490). |
|  | Sequencing | FSHD2: 39 (28%) | Of the 138 cases, 39 cases met criteria for FSHD2 based on identifying a pathogenic *SMCHD1* variant.  An additional 56 cases had D4Z4 hypomethylation, but no *SMCHD1* sequence variant was identified. The average methylation value for the D4Z4 hypomethylation cases without pathogenic *SMCHD1* variants was 22.6%. Of the remaining 43 cases that were sequenced, 24 had borderline low methylation values (>28%, but ≤35%), 14 had methylation values >35%, and 5 did not undergo methylation testing; in these cases, no pathogenic variants were identified. |
| Rodrigues 2015^217^ | Molecular diagnosis | FSHD NS: 12 (50%) | Only 50% of patients with FSHD had molecular test results confirming clinical diagnosis, while the other 50% had clinical diagnosis only. |
| Steel 2019^183^ | DNA digestion | FSHD NS: NR (100%) | Digestion of DNA using the bacterial restriction enzyme Bln1, to which the repeat fragment is resistant, was performed followed by analysis of the fragments’ length.  Larger fragments correspond to a preserved number of repeats. A Bln1-resistant fragment size of over 43 kb is considered normal and most individuals with FSHD1 have fragments of 10 kb to 35 kb. A small number of individuals have symptoms and signs of FSHD1 with fragments of between 35 kb and 43 kb and this range is considered borderline. |
| Upadhyaya 1997^202^ | Southern blotting | ≤34 kb: 123 (95%) | After performing southern blotting with p13E-11 probe, 123 out of the 130 FSHD patients (95%) had fragment sizes that were ≤34 kb.  Among the seven cases with larger fragments, the fragment sizes varied from 38 ± 2 to >48 kb.  Among 27 sporadic FSHD cases, the size distribution ranged from 10 to 48 kb (17.8 ± 8.8), while the size distribution among 103 familial cases ranged from 10 to >48 (22.0 ± 0.1) (p=0.01).  Among the control population, the smallest fragment size detected was 38 ± 2 kb, while the majority had fragment sizes that were within the limiting mobility of the DNA on the agarose gel (>48 kb). Nineteen subjects had smaller fragment sizes that ranged between 38 and 48 kb. |
| Yanoov-Sharav 2012^201^ | Southern blotting | FSHD NS: 32 (48%) | Of the 66 individuals tested for FSHD (41 males and 25 females), 59 patients had clinical findings suggestive of FSHD while 7 asymptomatic individuals were tested because of having a close relative with FSHD or suggestive laboratory results.  After performing southern blotting with p13E-11 probe, 32 cases were diagnosed with FSHD (D4Z4 repeat size <15-35 kb), 15 cases were in the grey zone (D4Z4 30-<40), and 19 had a normal allele conclusive of non-FSHD (D4Z4 ≥40). |

DNA, deoxyribonucleic acid; DNMT3B, DNA methyltransferase 3 beta; EMG, electromyography; FSHD, facioscapulohumeral muscular dystrophy; LRIF1, ligand dependent nuclear receptor interacting factor 1; MRI, magnetic resonance imaging; NGS, next-generation sequencing; NMD, neuromuscular disorder; NR, not reported; NS, not specified; PCR, polymerase chain reaction; PFGE, pulsed field gel electrophoresis; qPCR, quantitative PCR; SMCHD1, structural maintenance of chromosomes flexible hinge domain containing 1.

#### 3.8 Guidelines

Three guidelines were identified in the SLR (**Table 8**). Of the three guidelines, one was multinational and issued in 1993 following the 171^st^ International Workshop on FSHD^219^, one was issued from the French Muscular Dystrophy Association in France in 2011^220^ and the most recent one was issued from the American Academy of Neurology in 2015 ^221^. The multinational guidelines published in 1993 contained fewer information, as expected, and provided no recommendations on how to assess or evaluate FSHD nor on possible drug therapies, given the lack of available pharmacological treatments at the time.^219^ Genetic testing appeared as a recommendation only in the latest guidelines published in 2015, where it is noted that genetic confirmation of FSHD1 should be obtained in patients with atypical presentations and no first-degree relatives with genetic confirmation.^221^ Further, these guidelines also note that in addition to managing pain with pharmacological treatments, albuterol, corticosteroids or dilitiazem for improving strength should *not* be prescribed. The importance of aerobic/strength training was noted in two of the three guidelines.^219,221^ Recommendations on ophthalmic care and surgical intervention were included in all three guidelines.

Table 8. Guidelines for the management and treatment of FSHD

| **Guidelines** | **Country** | **Recommendations** |
| --- | --- | --- |
| **171^st^ ENMC International Workshop on FSHD ^219^** | Multinational | **Ophthalmic care:**   - Ocular lubricants may be offered. - Exudative retinopathy may be prevented by early intervention with laser treatment.   **Orthopedic care:**   - Identify assistive devices that may increase mobility and decrease risk of falls at home, such as ankle/foot orthoses to improve mobility/prevent falls in those with foot drop.   **Drug therapy:**   - NSAIDs for acute pain. - Antidepressants or anti-seizure medication for chronic pain.   **Aerobic / strength training:**   - Establish appropriate exercise regimens (e.g., moderate weight training, aerobic training).   **Surgical intervention:**   - Surgical fixation of scapula to chest wall should be offered cautiously with careful consideration of risk and benefit in the context of the patient’s symptoms.   **Other recommendations:**   - Ventilatory support (e.g., BiPAP) is recommended as necessary. - Standard therapies for hearing loss are recommended. - OT/speech therapy is recommended in patients with infantile onset. |
| **Recommendations for the management of FSHD in 2011 ^220^** | France | **Assessment and evaluations:**   - A functional workup should be carried out by a physical rehabilitation specialist physician, if possible, with the aid of a physiotherapist. - Respiratory functional tests with simple spirometry should be performed (measurement of VC, the VC/theoretical VC ratio). - Measurement of maximum inspiratory and expiratory pressures (Pi_Max_, Pe_Max_) would give further useful information; however, it is not systematically performed. - Cardiac evaluation is performed to screen for functional signs during the interview. It should include measurement of cardiac parameters (heart rate and BP), and a systematic ECG. Initially, echocardiography and 24-hour ECG monitoring are usually proposed. - A sensory assessment is performed when visual or auditory presenting symptoms are observed: an audiogram and/or fundoscopy at the slightest doubt (this test should be systematically performed in infantile forms of FSHD). Fluorescence angiography can be proposed if fundoscopy demonstrates retinal damage. - Assessment of psychological condition is desirable during the multidisciplinary assessment.   **Ophthalmic care:**   - A gel eye-drop can be prescribed upon request.   **Multidisciplinary care:**   - Children with early-onset and serious forms require specialized multidisciplinary monitoring coordinated by a neuro-pediatrician. The recommended frequency of monitoring visits is at least twice a year.   **Drug therapy:**   - Evaluation of pain should be performed systematically and, where necessary, be followed by prescription of an analgesic or a non-steroidal anti-inflammatory agent in the absence of contraindications.   **Social management:**   - Adjustment of the patient’s workstation or decision to declare the patient disabled should be studied on a case-by-case basis depending on the impairment.   **Surgical intervention:**   - The usefulness of scapulopexia (or scapulothoracic arthrodesis) is highly debatable and currently is only rarely proposed. |
| **Guidelines for the evaluation, diagnosis, and management of FSHD from the AAN ^221^** | US | **Assessment and evaluations:**   - Genetic confirmation of FSHD1 should be obtained in patients with atypical presentations and no first-degree relatives with genetic confirmation of the disease. - Baseline pulmonary function tests should be performed on all patients with FSHD. - Patients with FSHD should be referred for cardiac evaluation if they develop overt signs or symptoms of cardiac disease (e.g., shortness of breath, chest pain, palpitations). However, routine cardiac screening is not essential in the absence of cardiac signs or symptoms. - Clinicians should screen all young children with FSHD at diagnosis and yearly thereafter until these children start school, as hearing loss may not be present at diagnosis and can be progressive.   **Ophthalmic care:**   - Patients with FSHD and large deletions (contracted D4Z4 allele of 10–20 kb) should be referred to an experienced ophthalmologist (e.g., retina specialist) for dilated indirect ophthalmoscopy.   **Drug therapy:**   - In patients with persistent pain and no contraindications, a trial of nonsteroidal anti-inflammatory medications is appropriate for acute pain and antidepressants or antiepileptics for chronic pain. - In patients with FSHD, albuterol, corticosteroid, or diltiazem for improving strength should not be prescribed.   **Aerobic / strength training:**   - Patients with FSHD might be encouraged to engage in low-intensity aerobic exercise. An experienced physical therapist can help guide development of individualized exercise programs. The practical physical activities guidelines for individuals with disabilities, provided by the US Department of Health and Human Services, might also be used when counseling patients about aerobic exercise. - In patients interested in strength training, patients may be referred to physical therapists to establish a safe exercise program using appropriate low/medium weights/resistance that takes into consideration the patients’ physical limitations.   **Surgical intervention:**   - Surgical scapular fixation might be offered cautiously to select patients after careful consideration of the overall muscle impairment in the involved arm, assessment of potential gain in range of motion by manual fixation of the scapula, the patient’s rate of disease progression, and the potential adverse consequences of surgery and prolonged postsurgical bracing. |

AAN, American Academy of Neurology; BiPAP, bilevel positive airway pressure; BP, blood pressure; ECG, electrocardiogram; ENMC, European Neuromuscular Center; FSHD, facioscapulohumeral muscular dystrophy; NSAID, non-steroidal anti-inflammatory drugs; OT, occupational therapy; US, United States; VC, vital capacity.

### 4 Quality Assessment

A brief summary of the quality assessment is provided here, with details of the study quality assessment available in the accompanying Excel worksheet. Of note, not all tools provide an overall quality assessment. Thus, no summary of quality assessment is provided for Larg and Moss (n=3^150,151,156^), the JBI checklist for case series or case studies (case series, n=15^26-28,43,46-48,177,183,222-225^; case studies, n=7^23,24,34,40,49,197,198^) or the COSMIN risk of bias tool for studies reporting measures of reliability or validity (see the Excel sheet for the complete list of studies assessed with this tool).

Among the 20 clinical trials assessed with the Downs and Black checklist for both randomized and non-randomized studies, the mean methodological quality score was 17.6 (range 12 to 24) out of a possible 28.

Observational studies were assessed with multiple tools. All 31 cross-sectional studies were deemed to have sufficient quality to include, as assessed by the JBI checklist for cross-sectional studies (see the Excel sheet for complete list of studies assessed with this tool). A total of five case-control studies were identified in the SLR. Using the NOS case control quality assessment. Studies were assessed to have varying quality with a total number of “stars” awarded of 4^118^, 5^202^, 6^200^ and 7^188,214^ (out of a possible 8, indicating the highest quality). A total of 22 studies were assessed using the SIGN cohort checklist; while the majority of the studies were acceptable, overall quality was deemed low in eight studies due to either relatively small sample sizes, lack of calculation of confidence intervals or lack of controlling for risk of bias (see the Excel sheet for complete list of studies and assessment).

Qualitative studies were assessed using the JBI checklist for qualitative studies. While all five studies were assessed as “Include” in the overall appraisal^31,119,130,140,141^, none of the five studies included a statement addressing the role of the researcher and whether the beliefs and values have a potential influence on the study.

### 5 Discussion

FSHD is a genetic disorder that leads to the weakening of the skeletal muscles, profoundly impacting patients throughout the course of their life. Though 1 in 8,000 individuals worldwide are affected by FSHD, to date, there is no effective treatment or cure.

This SLR assessed the level of available published evidence to date on the burden of and unmet need in FSHD. A total of 230 studies, of which 78 were conference abstracts and 152 were full test articles, were identified. These publications were categorized into one of seven topics: “Treatment Outcomes and Clinical Trials”, “Disease Outcome Measures and Validation”, “Humanistic Burden and PROs”, “Economic Burden”, “Disease Classification”, “Diagnosis” and “Guidelines”. Globally, fewer identified studies focused solely on Type 1 FSHD patients and pediatric patients.

The most reported topic identified in the SLR was “Treatment Outcome and Clinical Trials”, where 50 full text articles (including 13 registers of clinical trials) and 14 conference abstracts, were identified. Given this breadth of evidence, the SLR focused on studies reporting outcomes on motor function, muscle strength and endurance or other muscle-related outcomes, from full text articles only. A total of 33 full text articles proceeded to data extraction, efficacy and effectiveness outcomes were categorized as: pharmacological (n=11), physiological (n=7), surgical (n=13) or other (n=3). Studies that reported on pharmacological interventions included both randomized and non-randomized trials along with observational studies and reported mainly on motor function and muscle strength and endurance outcomes. Studies reporting on physiological interventions also included a mix of study design and in addition to muscle strength and endurance outcomes, these studies included outcomes around timed function tests. Surgical interventions were the most homogenous of the four categories and included observational and case studies. All surgical intervention studies reported on shoulder mobility post-surgery. The three studies not fitting into the previous categories assessed herbal medicine, acupuncture and vibratory proprioceptive assistance and reported outcomes on shoulder mobility and muscle strength. Despite the ongoing research exploring multiple interventions, and given the impact of FSHD on the musculoskeletal system, there remains a substantial unmet need for effective treatments for patients living with FSHD.

The topic “Disease Outcome Measures and Validation” was the second-most reported topic identified in the SLR, with 28 full text and 28 conference abstracts identified. Measures of reliability and validity reported in these studies included correlation, Rasch analysis, factor analysis, reliability, responsiveness, and internal consistency. Multiple measures were assessed for reliability and/or validity, including measures of muscle strength, functional assessments, clinical severity (including disease severity), muscle composition (including fat fraction, lean muscle mass and fat infiltration), mobility, lung function, and PROs. Given this, there were few measures that were assessed by more than one study, with the exception of the MFM, FSHD-COM, FSHD-RODS, 6MWT and TUG. In addition to the assessment of these measures, studies focused on either using proxy measurements (such as MRI) to provide information about disease severity or progression, use of instruments in a home setting for clinical trials or the development of new instruments (such as the Facial Functional Scale).

The ”Humanistic Burden” of patients with FSHD was reported in 38 full text articles and 15 conference abstracts. While the majority of the studies in “Humanistic Burden” were observation in nature with a single point in time of measurement, some clinical trials included in the topic “Treatment Outcomes and Clinical Trials” did include an assessment of one or more PROs. Multiple instruments across the domains of general QoL, disease-specific QoL, depression, anxiety, pain, sleep, activities of daily living, disability/function, fatigue, neurological/psychological and symptom/illness perception were identified. Further, some studies reported a qualitative assessment of QoL without the use of an instrument. The most reported domains included general QoL, followed by fatigue then pain. QoL was assessed in both adult and pediatric FSHD patients, with some instruments developed exclusively for use among children and adolescents (for example, Kidscreen for general QoL and the FSHD-HI Peds for disease-specific QoL). Based on the available evidence, it is evident that FSHD impacts QoL, with pain and/or fatigue impacting QoL.

Limited evidence was found for the topic “Economic Burden”. A total of 18 studies (12 full text articles and 6 conference abstracts) reported on the economic burden among FSHD patients. Economic outcomes reported across the studies were heterogenous including medical and societal costs, number of prescriptions and healthcare resource use, such as wheelchair and hearing aid use and specialist visits. While al range of per patient annual direct and societal costs were identified in the literature, which was reported to be approximately five times higher than the mean per-capita health expenditure in the Netherlands.^150^ Nearly half of FSHD patients reported use of either ankle braces, leg braces, canes or walkers and over half noting that they used a wheelchair at some point during the day. In addition, a large proportion of patients with FSHD have reduced employment opportunities. While limited, the available evidence suggests that FSHD is associated with a substantial economic burden for patients, their caregivers and society.

A high number of studies reporting on the topic of “Disease classification” was identified in the SLR, including 28 full text articles and 15 conference abstracts. Across the studies, multiple criteria were employed to classify FSHD, including classification by the CCEF, onset (early, typical, atypical), type of FSHD (1, 2, 1 and 2), subtype (classical, facial sparing, early upper limb weakness), number of D4Z4 repeats, among others. The multiple classification criteria currently in use highlight the diversity of FSHD and the lack of consensus in this space.

Of the 17 full text articles and 7 conference for the topic “Diagnosis”, ranging from clinical assessments to genetic testing. The most reported technique was Southern blotting, though publications tended to be older (pre-2012). Other techniques reported in the identified literature included muscle MRI, haplotyping, methylation assay plus sequencing, PFGE, PCR, and NGS. While the results of this SLR suggest that genetic testing is a crucial step to confirming a clinical FSHD diagnosis, there are currently no guidelines or consensus on the preferred technique/diagnostic algorithm.

Only three guidelines were identified in the SLR, with the latest guideline published in 2015; thus, they may not reflect recent medical advances (e.g., in pain management) that translate into better management of patients with FSHD. Overall, the identified guidelines recommend a holistic approach to FSHD management including strength training, pain medications, orthopedic and ophthalmic care as well as social management. This SLR found no national or international recent (<5 years) clinical recommendations on FSHD management, highlighting the need for establishing an effective management plan for patients with FSHD.

Overall, the findings of this SLR indicate that despite being the second most prevalent form of muscular dystrophy, FSHD is not well characterized in the literature. Multiple unique outcomes assessing disease progression were identified; consensus on both what and how to measure treatment benefit for FSHD patients is lacking. While multiple studies reported on the humanistic burden, there was heterogeneity in the instruments used to assess the multidimensional detriments of patients with FSHD, including depression, anxiety, depression and pain. Multiple studies reported on the reliability and validity of instruments in FSHD, however, there the measures assessed were heterogeneous. Few studies were identified on diagnosis and disease management guidelines, highlighting the variability in time to diagnosis and treatment patterns experienced by patients. Given the breadth of outcomes reported, it is difficult to quantify the true disease burden experienced by patients, their families, and the healthcare system. Efforts should be made to harmonize the assessment of the debilitating and progressive nature of FSHD, as research on treatment options continue.

5.	FULCRUM Therapeutics. Fulcrum Therapeutics Announces Results from ReDUX4 Trial with Losmapimod in Facioscapulohumeral Muscular Dystrophy (FSHD) Demonstrating Slowed Disease Progression and Improved Function. 2021.

6.	University of York Centre for Reviews and Dissemination. Systematic Reviews: CRD's guidance for undertaking reviews in health care, 2008.

7.	Page MJ, McKenzie JE, Bossuyt PM, et al. The PRISMA 2020 statement: an updated guideline for reporting systematic reviews. *BMJ* 2021; **372**: n71.

8.	Mokkink LB, Boers M, van der Vleuten CPM, et al. COSMIN Risk of Bias tool to assess the quality of studies on reliability or measurement error of outcome measurement instruments: a Delphi study. *BMC Medical Research Methodology* 2020; **20**(1): 293.

9.	Larg A, Moss JR. Cost-of-illness studies: a guide to critical evaluation. *Pharmacoeconomics* 2011; **29**(8): 653-71.

10.	SIGN. Methodology checklist 3: cohort studies. <https://www.sign.ac.uk/what-we-do/methodology/checklists/>.

11.	JBI Manual for Evidence Synthesis: JBI; 2020.

25.	Ian Woodcock KDV, Eppie Yiu,Kate Carroll,Rachel Kennedy,Monique Ryan. "Effect of creatine monohydrate on functional muscle strength and muscle mass in children with FSHD: a multi-centre, randomised, double-blind placebo-controlled crossover trial.". *FSHD IRC 2019 P12* 2019.

77.	Haley K, Perera R, Wagner K, Tawil R, Statland J, Johnson N. Analysis of the facioscapulohumeral muscular dystrophy clinical score (FSHD CS) using item response theory. *FSHD IRC 2020 P503* 2020.

78.	Hristov RR, Hariharan; Kabelac, Zachary; Chan, Vicky; et al,. In-Home Passive Measurements of Mobility and Sleep in FSHD Patients and their Relationship with Clinical Disease Severity Metrics. *MNDA 2021* 2021.

79.	Hatch MN, Kurillo G, Chan V, Han JJ. Motion sensor-acquired reachable workspace correlates with patient-reported upper extremity activities of daily living (ADL) function in facioscapulohumeral dystrophy. *Muscle Nerve* 2021; **63**(2): 250-7.

80.	Berggren KE, Kate; McDermott, Michael; Higgs, Kiley; et al,. Orofacial strength assessment and correlates with participant and clinician report of function in FSHD. *MNDA 2022* 2022.

81.	Chan VH, Maya; Mellion, Michelle; Cadavid, Diego; Han, Jay. An optimized Timed Up and Go (TUG) for the Facioscapulohumeral Dystrophy (FSHD) community. *MNDA 2021* 2021.

82.	Chan V, Hatch M, Kurillo G, Han J, Cadavid D. Development of an optimized timed up and go (OTUG) for measurement of changes in mobility impairment in facioscapulohumeral muscular dystrophy (FSHD) clinical trials. *Neurology* 2020; **94**(15).

83.	Eichinger K, McDermott M, Higgs K, et al. Strength is Associated with Functional Abilities and Changes over a 12 Month Period in Individuals with Facioscapulohumeral Dystrophy (FSHD). *Neurology* 2022; **98**(18 SUPPL).

84.	Eichinger K, Higgs K, Lewis L, Tawil R, Statland J. Relationship of strength to functional assessments for individuals participating in the ReSolve FSHD study. *FSHD IRC 2021 P715* 2021.

97.	Woodcock I, Valle Kd, Ryan M. Baseline MRI data and correlations with functional measures from the creatine in FSHD clinical trial. *FSHD IRC 2020 P501* 2020.

109.	Gazzotti M, Cordoba Lanza, F.,Custódio Marchetto, A.,Bandoszeweski Castilho, A.,Luque, A. Exercise training in facioscapulohumeral muscular dystrophy (FSHD): Serial cases with 1 year of follow-up. *Physiotherapy (United Kingdom)*.

117.	Vissing J, Attarian, S.,Gidaro, T.,Mozaffar, T.,Iyadurai, S.,Walker, G.,Shukla, S.,Servais, L.,Wagner, K. Results of a Phase 1b/2 Study of ATYR1940 in adult patients with limb girdle muscular dystrophy Type 2B (LGMD2B) and facioscapulohumeral muscular dystrophy (FSHD) (ATYR1940-C-004). *Neurology* 2017.

153.	Zielman-Blokhuis A, Deenen J, Voermans N, Kievit W, Groothuis J. The socioeconomic burden of facioscapulohumeral muscular dystrophy. *FSHD IRC 2020 P538* 2020.

219.	Preston MK, Tawil R, Wang LH. Facioscapulohumeral Muscular Dystrophy. In: Adam MP, Mirzaa GM, Pagon RA, et al., eds. GeneReviews(®). Seattle (WA): University of Washington, Seattle

Copyright © 1993-2023, University of Washington, Seattle. GeneReviews is a registered trademark of the University of Washington, Seattle. All rights reserved.; 1993.

227.	Jeffrey Statland EB, Chafic Karam,Lauren Elman,Nicholas Johnson,Nanette Joyce,John T Kissel,Perry B Shieh,Lawrence Korngut,Chris Weihl,Rabi Tawil,Anthony Amato,Craig Campbell,Angela Genge,Georgios Manousakis,Ashley Leneus,Barry Miller,Chad E Glasser,Robert K Zeldin,Kenneth M Attie. "Dose escalation results from a phase 2 study of ACE-08, a local muscle therapeutic, in patients with facioscapulohumeral muscular dystrophy (FSHD).". *FSHD IRC 2019 S91* 2019.

228.	van Engelen BGM, Janssen B, Voet N, et al. S.P.43 Physical activity decreases fatty infiltration in FSHD, a quantitative muscle MR study. *Neuromuscular Disorders* 2012; **22**(9): 896.

229.	Di Marco J, Frageta, C.,Lansaman, T.,Missaoui, B.,Thoumie, P. Gait and balance parameters in patients with fascioscapulohumeral muscular dystrophy: A short term evaluation of a rehabilitation program. *Annals of Physical and Rehabilitation Medicine*.

230.	Kulshrestha R, Emery, N.,Arkesteijn, M.,Willis, T. Arm cycling in Facioscapulohumeral dystrophy (FSHD) patients: Results of pilot study. *Journal of Neuromuscular Diseases*.

267.	Schwarzer R, Jerusalem M, Weinman J, Wright S, Johnston M. Generalized Self-Efficacy Scale. *Measures in Health Psychology: A User's Portfolio Causal and control beliefs Windsor* 1995.

296.	Rey A. L'examen clinique en psychologie. [The clinical examination in psychology.]. Oxford, England: Presses Universitaries De France; 1958.

297.	Rosenberg M. Society and the Adolescent Self-Image: Princeton University Press; 1965.

305.	Spielberger C, Gorsuch R, Lushene R, Vagg P, Jacobs G. Manual for the State-Trait Anxiety Inventory; Palo Alto, CA, Ed. *Palo Alto: Spielberger* 1983.

311.	Karpati G. Disorders of Voluntary Muscle. 8 ed. Cambridge: Cambridge University Press; 2010.

312.	Konersman C, Munoz K, Brook R, et al. Prescription medications during patients' journey to the diagnosis of facioscapulohumeral muscular dystrophy: a real-world retrospective data analysis. *Journal of Managed Care and Specialty Pharmacy* 2022; **28**(3-A): S52-S3.

### 7 Appendix

#### 7.1 Electronic Database Search Strategy

Preliminary results suggest a broader search strategy would be preferred to avoid missing potentially relevant studies of interest. Keywords and search terms related to inclusion/exclusion criteria and/or topic (e.g., clinical, humanistic, economic burden) are not recommended.

Search strategy presented below will be used to search MEDLINE (via PubMed.gov), EMBASE (via Embase.com) and Cochrane (via Cochranelibrary.com) databases for full-text publications. Conference abstracts will only be searched in Embase.com.

Table A-1. Search Strategy: PICOS Criteria

|  | **SEARCH SYNTAX: PubMed / MEDLINE** |
| --- | --- |
| **POPULATION/DISEASE** | "Muscular Dystrophy, Facioscapulohumeral"[MESH] OR FSHD OR "Facioscapulohumeral muscular dystrophy" OR “Facioscapulohumeral dystrophy” OR “facio-scapulo-humeral dystrophy” OR “Landouzy-Dejerine” |
| **INTERVENTION** | Terms not included given manageable number of hits retrieved with population search terms |
| **COMPARATOR** |  |
| **OUTCOMES** |  |
| **STUDY TYPE** |  |

Table A- 2. Search Strategy: PubMed / MEDLINE

|  | **Key word** | **Hits** |
| --- | --- | --- |
| 1 | "Muscular Dystrophy, Facioscapulohumeral"[MESH] OR FSHD OR "Facioscapulohumeral muscular dystrophy" OR “Facioscapulohumeral dystrophy” OR “facio-scapulo-humeral dystrophy” OR “Landouzy-Dejerine” | 1,670 |
| 2 | #1 AND English[la] | 1,546 |

Search conducted on October 11, 2022.

Table A- 3. Search Strategy: EMBASE

|  | **Key word** | **Hits** |
| --- | --- | --- |
| 1 | FSHD OR "Facioscapulohumeral muscular dystrophy" OR “Facioscapulohumeral dystrophy” OR “facio-scapulo-humeral dystrophy” OR “Landouzy-Dejerine” | 2,924 |
| 2 | #1 AND ([conference abstract]/lim OR [conference paper]/lim OR [conference review]/lim) | 699 |
| 3 | #1 NOT #2 | 2,225 |
| 4 | #3 AND [english]/lim | 2,026 |
| 5 | #2 AND [english]/lim | 694 |

Search conducted on October 11, 2022.

Table A- 4. Search Strategy: Cochrane CENTRAL

| **N** | **Key word** | **Hits** |
| --- | --- | --- |
| 1 | FSHD OR "Facioscapulohumeral muscular dystrophy" OR “Facioscapulohumeral dystrophy” OR “facio-scapulo-humeral dystrophy” OR “Landouzy-Dejerine” | 91 |

Search conducted on October 11, 2022.

#### 7.2 Detailed tables for “Treatment Outcomes and Clinical Trials”

Table 9. Study characteristics for “Treatment Outcomes and Clinical Trials”

| **Author, year** | **Country** | **Intervention type** | **Study design** | **FSHD population type: n (%)** | **Proportion pediatric** | **Efficacy / effectiveness outcomes included** | **Other outcomes included** |
| --- | --- | --- | --- | --- | --- | --- | --- |
| CONFERENCE ABSTRACTS – TRIALS (RANDOMIZED AND NON-RANDOMIZED) | | | | | | | |
| Andersen 2013a^146^ | NR | Physiological: Cycle training with protein supplementation | Non-randomized trial | FSHD NS: 23 (100%) | 0% | 6MWT  Maximal load  Maximal oxygen uptake  Muscle strength | --- |
| Buch  2015^226^ | NR | Physiological: High intensity training versus control | Randomized controlled trial | FSHD NS: 13 | 0% | VO_2_ max | --- |
| Kools 2022^16^ | NR | Pharmacological: Losmapimod | Non-randomized trial | Type 1: 14 (100%) | 0% | 6MWT  Gene expression in muscle  MFM  Muscle health  RWS  FSHD TUG & TUG | FSHD-HI  FSHD RODS  PGI-C  TEAEs |
| Statland 2019^227^ | NR | Pharmacological: ACE-083 | Non-randomized trial  NCT02927080 | FSHD NS: 36 (100%) | 0% | Muscle fat fraction | AEs |
| Tawil 2022^17^ | NR | Pharmacological: Losmapimod | Randomized controlled trial | Type 1: 80 (100%) | 0% | Dynamometry  Gene expression in muscle  MFM  Muscle health  RWS  FSHD TUG & TUG | FSHD-HI  PGI-C  SAEs |
| Van Engelen 2012^228^ | NR | Physiological: Aerobic exercise training versus cognitive behavioral therapy versus control | Randomized controlled trial | FSHD NS: 24 (100%) | 0% | Muscle fat infiltration  Muscle fat fraction | --- |
| Van Engelen 2014^18^ | NR | Physiological: Aerobic exercise training versus cognitive behavioral therapy versus usual care | Randomized controlled trial | Type 1: 57 (100%) | 0% | Muscle fat fraction  Registered physical activity | Fatigue  Sleep quality  Social participation |
| Vissing 2017^117^ | NR | Pharmacological: ATYR1940 | Non-randomized trial | FSHD NS: 8 (100%) | 0% | Biomarkers  MMT  Muscle health  Pulmonary function | AEs  INQoL |
| Walker 2017^19^ | NR | Pharmacological: ATYR1940 | Non-randomized trial | Type 1: 8 (100%) | 0% | Biomarkers  Hearing  MMT  Muscle health  Ophthalmology  Pulmonary function | AEs  INQoL |
| Woodcock 2019^25^ | NR | Other: Creatinine monohydrate | Randomized controlled trial | FSHD NS: NR (100%) | 100% | MFM | --- |
| CONFERENCE ABSTRACTS – OBSERVATIONAL | | | | | | | |
| Di Marco 2013^229^ | France | Physiological: Physiotherapy, hydrotherapy and occupation therapy | Retrospective cohort | FSHD NS: 24 (100%) | 0% | Berg Balance Scale  Functional reach test  Gait speed  Muscle strength  TUG | --- |
| Kulshrestha 2019^230^ | NR | Physiological: Upper limb cycling | Prospective cohort | FSHD NS: 15 | 0% | Oxford shoulder score  Rating of perceived exertion | --- |
| Torrente 2014^231^ | NR | Other: Autologous stem cells | Prospective cohort | FSHD NS: 13 | 0% | 6MWT  QMT | --- |
| CONFERENCE ABSTRACTS – OTHER | | | | | | | |
| Gazzotti 2011^109^ | NR | Physiological: Exercise training | Case series | FSHD NS: 3 (100%) | 0% | 6MWT  Maximal respiratory pressures | SF-36 |
| FULL TEXT ARTICLES – TRIALS (NON-RANDOMIZED AND RANDOMIZED) | | | | | | | |
| Andersen 2015^20^ | Denmark | Physiological:  Training ± protein supplement versus control | Randomized controlled trial | Type 1: 41 (100%) | 0% | 5-times-sit-to-stand-test  6MWT  14-step-stair-test  Dynamometry  Standing balance test  VO_2_ max  Wmax | AEs  Daily activity  Fatigue  Muscle damage  Pain  Plasma CK levels  SF-36 |
| Andersen 2017^41^ | Denmark | Physiological: High intensity training versus usual care | Randomized controlled trial | FSHD NS: 12 (100%) | 0% | 5-times-sit-to-stand-test  6MWT  Maximal workload  Dynamometry  VO_2_ max | Plasma CK levels |
| Bankolé 2016^21^ | France | Physiological: Training versus control | Randomized controlled trial | Type 1: 16 (100%) | 0% | 6MWT  Creatine kinase  MAP  Muscle cross-sectional area  Muscle endurance  MVIC  Voluntary activation at rest  VO_2_ max | Fatigue Severity Scale  Serum CK levels  SF-36  Soreness and tiredness |
| Elsheikh 2007^232^ | NR | Pharmacological: Diltiazem hydrochloride | Non-randomized trial | FSHD NS: 20 (100%) | 0% | Lean body mass  MMT  MVIC  Walk 30 ft | --- |
| Kissel 2001^36^ | US | Pharmacological: Albuterol high dose versus albuterol low dose versus placebo | Randomized controlled trial | FSHD NS: 90 (100%) | 0% | 4-stair climb  Body composition  Functional grading  Grip strength  Lean body mass  MMT  MVIC  Walk 30 ft | AEs |
| Mellion 2021^32^ | Netherlands | Pharmacological:  Losmapimod 7.5 mg or 15 mg versus placebo | Randomized controlled trial  NL8000 | FSHD NS: 15 (100%) | 0% | --- | AEs  SAEs |
| Passerieux 2015^37^ | France | Pharmacological: Supplements versus placebo | Randomized controlled trial  NCT01596803 | FSHD NS: 53 (100%) | 0% | 2MWT  Muscle endurance  MVC | --- |
| Payan 2009^138^ | France | Pharmacological: Salbutamol versus placebo | Randomized controlled trial | FSHD NS: 112 (100%) | 0% | 4-stair climb  Barré test  Brooke score  Mingazzini test  MMT  MVIC QMT  Vignos score  Walk 10 m | AEs  Plasma CK levels  SF-36 |
| Statland 2022^38^ | US  Canada  Spain | Pharmacological: ACD-083 versus placebo | Randomized controlled trial  NCT03943290 | FSHD NS: 55 | 0% | 4-stair climb  6MWT  Walk/run 10 m  MMT MRC  Muscle fat fraction  MVIC  PUL  Total and contractile muscle volume | FSHD-HI  TEAEs |
| Tawil  1997^233^ | US | Pharmacological: Prednisone | Non-randomized trial | FSHD NS: 8 (100%) | 0% | Lean body mass  MMT  MVIC  Serum CK levels | AEs |
| Van der Kooi  2004^234^ | Netherlands | Pharmacological+ physiological:  Placebo ± training versus albuterol ± training | Randomized controlled trial | FSHD NS: 65 (100%) | 0% | 3-stair climb  Functional grading  Muscle endurance  Muscle strength  MVIC  Pulmonary function  Standing from lying  Standing from sitting  Total muscle volume  Walk 30 ft | AEs  Tolerability of training |
| Voet  2014^144^ | Netherlands | Physiological: Aerobic exercise training versus cognitive behavior therapy versus control | Randomized controlled trial | FSHD NS: 57 (100%) | 0% | 6MWT  Body acceleration per 5-minute  MVIC  VO_2_ max | AEs  CIS-activity  CIS-fatigue  NHP-sleep  SIP68-behavior  VAS-pain |
| Wagner 2008^35^ | US  UK | Pharmacological: MYO-029 1 mg/kg, 3 mg/kg, 10 mg/kg, 30 mg/kg versus placebo | Randomized controlled trial | FSHD NS: 42 (100%) | 0% | 4-stair climb  FVC  Lean body mass  MMT  Muscle histology  Muscle volume  Myostatin levels  QMT  Standing from sitting  Walk 9 m | AEs  SAEs |
| FULL TEXT ARTICLES – OBSERVATIONAL | | | | | | | |
| Andersen 2013b^33^ | NR | Physiological: High intensity training | Prospective cohort | FSHD NS: 5 (100%) | 0% | Creatine kinase  VO_2_ max | Pain |
| Andrews 1998^28^ | NR | Surgical: Scapulothoracic arthrodesis | Retrospective cohort | FSHD NS: 6 (100%) | 17% | Activities of daily living  Forced expiratory volume  FVC  Shoulder abduction  Shoulder flexion  Vital capacity | Complications  Cosmetic satisfaction  Pain  Patient satisfaction |
| Bunch 1993^222^ | US | Surgical: Scapulothoracic arthrodesis | Retrospective cohort | FSHD NS: 12 (100%) | NR | Activities of daily living  Shoulder abduction  Shoulder flexion | Complications |
| Colson 2010^42^ | France | Physiological: Neuromuscular electrical stimulation training | Prospective before-after trial | FSHD NS: 9 (100%) | 0% | 6MWT  MMT  MVIC | Satisfaction  Plasma CK levels |
| Cooney 2014^43^ | NR | Surgical: Scapulothoracic arthrodesis | Prospective cohort | FSHD NS: 11 (100%) | 0% | FVC  DASH  Shoulder abduction  Shoulder flexion | Complications |
| Diab  2005^223^ | NR | Surgical: Scapulothoracic arthrodesis | Retrospective cohort | FSHD NS: 8 (100%) | NR | Clinical grading  Shoulder abduction  Shoulder flexion | Complications |
| Doix  2017^22^ | NR | Physiological: Neuromuscular electrical stimulation training | Prospective cohort | Type 1: 10 (100%) | 0% | 6MWT  MFM  MMT  MVIC | Fatigue  Plasma CK levels  SF-36 |
| Eren 2020a^44^ | Turkey | Surgical: Scapulothoracic arthrodesis | Retrospective cohort | FSHD NS: 40 (100%) | NR | Pulmonary function  qDASH  Shoulder abduction  Shoulder elevation | Complications |
| Erşen 2018^45^ | Turkey | Surgical: Scapulothoracic arthrodesis | Retrospective cohort | FSHD NS: 13 (100%) | 0% | qDASH  Shoulder abduction  Shoulder flexion | Complications |
| Giannini 2007^26^ | NR | Surgical: Scapulopexy | Retrospective cohort | FSHD NS: 13 (100%) | 15% | Activities of daily living  FVC  MRC  Shoulder abduction  Shoulder flexion | Complications  Cosmetic satisfaction  Functional satisfaction |
| Kissel 1998^39^ | US | Pharmacological: Albuterol | Prospective cohort | FSHD NS: 15 (100%) | 0% | Body fat  Body mass  Lean body mass  MMT  MVIC | AEs |
| Le Hanneur 2017^46^ | NR | Surgical: Scapulothoracic arthrodesis | Retrospective cohort | FSHD NS: 4 (100%) | NR | Activities of daily living  Constant score  Shoulder abduction strength  Shoulder abduction  Shoulder extension  Shoulder flexion  Shoulder rotation | Complications  Cosmetic satisfaction |
| Loock 2022^47^ | NR | Surgical: Scapulothoracic arthrodesis | Retrospective cohort | FSHD NS: 7 (100%) | 0% | Brooke score  Constant score  FVC  MRC  Shoulder abduction  Shoulder flexion  Shoulder rotation  Vignos score | Complications  VAS-pain |
| Olsen 2005^235^ | NR | Physiological: Aerobic exercise training | Prospective cohort | FSHD NS: 8 (100%) | 0% | Capillary density  Muscle fiber area  Muscle histology  VO_2_ max  Workload | Plasma CK levels |
| Ribot-Ciscar 2015^50^ | France | Other: Vibratory proprioceptive assistance | Prospective cohort | FSHD NS: 9 (100%) | 0% | Constant score  Maximum isometric abduction force  Shoulder abduction  Shoulder flexion | Pain  SF-36 |
| Van Tongel 2013^225^ | NR | Surgical:  Thoracoscapular arthrodesis | Retrospective cohort | FSHD NS: 24 | NR | Activities of daily living  Constant score  Shoulder elevation  Shoulder motion range  Strength | Complications  Cosmetic satisfaction  Pain  Subjective Shoulder Value |
| FULL TEXT ARTICLES – OTHER | | | | | | | |
| Abel  2018^31^ | Netherlands  Belgium | Surgical:  Bariatric surgery | Qualitative | FSHD NS: 3 (100%) | 0% | Activities of daily living  Body mass & BMI  Impact on co-morbidity | --- |
| Cakmak 2018^236^ | Turkey | Surgical: Scapulothoracic arthrodesis | Narrative | FSHD NS: 32 (100%) | NR | Shoulder elevation | Complications  QoL |
| Eren 2020b^34^ | Turkey | Surgical: Spinal fusion | Case study | FSHD NS: 1 (100%) | 0% | --- | INQoL |
| Jakab 1993^27^ | NR | Surgical: Scapulothoracic arthrodesis | Case study | FSHD NS: 3 (100%) | 66% | Activities of daily living  Forced expiratory volume  Respiratory volume  Shoulder abduction  Shoulder flexion  Vital capacity | Complications  Cosmetic satisfaction  Pain |
| Lefkowitz 2012^40^ | NR | Pharmacological: Denosumab | Case study | FSHD NS: 1 (100%) | 0% | Grip strength  Standing from sitting  TUG | --- |
| Li  2021^23^ | China | Other: Herbal medicine | Case study | Type 1: 1 (100%) | 100% | Muscle strength  Shoulder abduction | --- |
| Liu  2019^49^ | NR | Other: Acupuncture | Case study | FSHD NS: 1 (100%) | 0% | Muscle strength  Pulmonary function | --- |
| Xiao 2021^24^ | NR | Pharmacological:  Supplement cocktail | Case study | Type 1: 2 (100%) | 100% | Muscle weakness | --- |
| Ziaee 2006^48^ | Iran | Surgical: scapulothoracic arthrodeses | Case study | FSHD NS: 6 (100%) | NR | Activities of daily living  FVC  Shoulder abduction  Shoulder flexion | Complications  Cosmetic satisfaction |
| REGISTERS OF CLINICAL TRIAL | | | | | | | |
| 2022^237^ | Multinational | Pharmacological | Randomized controlled trial  NCT05397470 | FSHD NS: NR (100%) | 0% | Muscle fat infiltration  RWS | Neuro-QoL UE  PGI-C  TEAEs |
| 2020^29^ | Europe  US  Canada | Pharmacological | Non-randomized trial  NCT04264442 | Type 1: NR (100%) | 0% | --- | AEs  SAEs |
| 2022^238^ | US  Canada  Germany  France  Italy  Spain  UK  Netherlands  Denmark | Pharmacological | Randomized controlled trial  EUCTR2022-000389-16-ES | FSHD NS: NR (100%) | 0% | Muscle fat infiltration  RWS | AEs  Neuro-QoL UE  PGI-C  SAEs |
| 2000^239^ | US | Pharmacological | Randomized controlled trial  NCT00004685 | FSHD NS: NR (100%) | 0% | Lean body mass  MVIC | AEs |
| 2019^30^ | US  Germany  France  Spain | Pharmacological | Randomized controlled trial  EUCTR2019-01181-15-ES | Type 1: NR (100%) | 0% | Gene expression in muscle | AEs  SAEs |
| 2014^240^ | US  France  Italy  Netherlands | Pharmacological | Randomized controlled trial  EUCTR2014-001753-17-IT | FSHD NS: NR (100%) | 0% | MMT  Pulmonary function | AEs  INQoL |
| 2015^241^ | US  Italy  Netherlands | Pharmacological | Non-randomized trial  EUCTR2015-001912-36-NL | FSHD NS: NR (100%) | 0% | MMT  Pulmonary function  QMT  Vignos score | AEs  FSHD-HI  INQoL  SAEs  Sleep quality |
| 2005^242^ | NR | Pharmacological | Randomized controlled trial  EUCTR2004-000622-67-GB | FSHD NS: NR (100%) | 0% | Lean body mass  Muscle histology  Muscle strength  Total muscle volume | AEs |
| 2016^243^ | Multinational | Pharmacological | Non-randomized trial  EUCTR2016-000624-25-DK | FSHD NS: NR (100%) | 0% | Biomarkers  Brooke score  MMT  Muscle disease burden  Pulmonary function  Vignos score | AEs  FSHD-HI  INQoL  SAEs  TEAEs |
| 2014^244^ | Multinational | Pharmacological | Randomized controlled trial  NCT02239224 | FSHD NS: NR (100%) | 0% | MMT | INQoL  TEAEs |
| 2010^245^ | France | Physiological | Randomized controlled trial  NCT01116570 | FSHD NS: NR (100%) | 0% | Muscle histology  VO_2_ max | Fatigue Severity Scale  SF-36 |
| 2016^246^ | France | Physiological | Randomized controlled trial  NCT02861911 | FSHD NS: NR (100%) | 0% | Body composition  Bone mineral density  Muscle endurance  MVIC | SF-36 |
| 2012^247^ | Denmark | Physiological | Randomized controlled trial  NCT01618331 | FSHD NS: NR (100%) | 0% | 6MWT  Creatine kinase  Risk of falls  VO_2_ max | Daily activity level questionnaire |

2MWT, 2-minute walking test; 6MWT, 6-minute walking test; AE , adverse event; BMI, body mass index; CIS, Checklist Individual Strength; CK, creatine kinase; DASH, Disabilities of the Arm, Shoulder and Hand; FSHD, facioscapulohumeral muscular dystrophy; FSHD-HI, FSHD health index; FSHD-RODS, FSHD Rasch-built overall disability scale; ft, feet; FVC, forced vital capacity; INQoL, Individualized Neuromuscular Quality of Life; Neuro-QoL UE, Quality of Life in Neurology – Upper Extremity; MAP, maximal aerobic power; MFM, motor function measurement; MMT, manual muscle testing; MRC, Medical Research Council; MVC, maximal voluntary contraction; MVIC, maximal voluntary isometric contraction; NHP, Nottingham Health Profile; NR, not reported; NS, not specified; PGI-C, Patient Global Impression – Change; PUL, performance of the upper limb; qDASH, Quick Disabilities of the Arm, Shoulder and Hand; QMT, quantitative muscle testing; QoL, quality of life; RWS, reachable work space; SAE, serious adverse event; SES, Self-efficacy Scale; SF-36, Short Form 36; SIP68, Sickness Impact Profile 68; TEAE, treatment emergent adverse event; TUG, timed up and go; UK, United Kingdom; US, United States; VAS, Visual Analogue Scale; VO_2_, oxygen consumption; W_max_, maximal workload.

Table 10. Population characteristics for extracted “Treatment Outcomes and Clinical Trials” studies

| **Author, year** | **Treatment arm** | **FSHD population type: n (%)** | **Age (years)** | **Female, n (%)** | **BMI (kg/m^2^)** | **Age onset (years)** | **Disease duration (years)** | **CSS** | **D4Z4 pairs (kb)** |
| --- | --- | --- | --- | --- | --- | --- | --- | --- | --- |
| PHARMACOLOGICAL | | | | | | | | | |
| Elsheikh  2007^232^ | Diltiazem hydrochloride | FSHD NS: 19 (100%) | NR (21–60) | --- | --- | --- | --- | --- | --- |
| Kissel  1998^39^ | Albuterol | FSHD NS: 15 (100%) | 36.2 ± 9.3 (19–50) | 7 | --- | --- | --- | --- | --- |
| Kissel  2001^36^ | Placebo | FSHD NS: 30 (33%) | 34.7 ± 10.9 | NR (33%) | --- | 18.5 ± 8.3 | --- | --- | --- |
|  | Albuterol  low dose | FSHD NS: 30 (33%) | 35.9 ± 10.8 | NR (47%) | --- | 17.6 ± 8.0 | --- | --- | --- |
|  | Albuterol  high dose | FSHD NS: 30 (33%) | 35.5 ± 9.7 | NR (57%) | --- | 17.7 ± 8.4 | --- | --- | --- |
| Lefkowitz 2012^40^ | Denosumab | FSHD NS: 1 (100%) | 66.0 | 1 | --- | --- | --- | --- | --- |
| Passerieux 2015^37^ | Supplements | FSHD NS: 26 (49%) | 42.4 ± 10.2 | 11 (42%) | 24.7 ± 4.2 | --- | --- | --- | --- |
|  | Placebo | FSHD NS: 27 (51%) | 37.6 ± 9.3 | 14 (52%) | 22.6 ± 3.5 | --- | --- | --- | --- |
| Payan  2009^138^ | Placebo | FSHD NS: 56 (50%) | 37.7 ± 11.1 (18–60) | NR (39%) | --- | --- | 10.6 ± 7.5 (1–28) | --- | --- |
|  | Salbutamol | FSHD NS: 56 (50%) | 42.3 ± 10.7 (22–60) | NR (25%) | --- | --- | 8.9 ± 7.7  (1–42) | --- | --- |
| Statland  2022^38^ | Biceps brachii Placebo | Type 1 & II: 14 (25%) | 42.5 (21–65) | 3 (21%) | 23.5 (12.0–27.9) | --- | 20.5 (5–50) | --- | --- |
|  | Biceps brachii  ACE-083 | Type 1 & II: 14 (25%) | 47.5 (28–68) | 4 (29%) | 24.0 (19.1–35.0) | --- | 21.5 (4–42) | --- | --- |
|  | Tibialis anterior placebo | Type 1 & II: 14 (25%) | 43.5 (18–62) | 7 (50%) | 25.7 (15.6–51.2) | --- | 19.5 (2–44) | --- | --- |
|  | Tibialis anterior ACE-083 | Type 1 & II: 13 (24%) | 54.0 (31–70) | 7 (54%) | 23.8 (19.7–28.9) | --- | 24.0 (4–62) | --- | --- |
| Tawil  1997^233^ | Prednisone | FSHD NS: 8 (100%) | 32.5 | 2 | --- | --- | --- | --- | --- |
| Van der Kooi 2004^234*^ | Placebo | FSHD NS: 16 (25%) | 39.0 ± 9.0 | 7 | --- | --- | --- | --- | --- |
|  | Placebo + training | FSHD NS: 19 (29%) | 36.0 ± 9.0 | 8 | --- | --- | --- | --- | --- |
|  | Albuterol | FSHD NS: 15 (23%) | 41.0 ± 12.0 | 6 | --- | --- | --- | --- | --- |
|  | Albuterol + training | FSHD NS: 15 (23%) | 36.0 ± 11.0 | 4 | --- | --- | --- | --- | --- |
| Wagner  2008^35^ | Placebo | --- | --- | --- | --- | --- | --- | --- | --- |
|  | MYO-029  1 mg/kg | --- | --- | --- | --- | --- | --- | --- | --- |
|  | MYO-029  3 mg/kg | --- | --- | --- | --- | --- | --- | --- | --- |
|  | MYO-029  10 mg/kg | --- | --- | --- | --- | --- | --- | --- | --- |
|  | MYO-029  30 mg/kg | --- | --- | --- | --- | --- | --- | --- | --- |
| Xiao  2021^24^ | Supplement cocktail | Type 1: 2 (100%) | --- | 1 (50%) | --- | --- | --- | --- | --- |
| PHYSIOLOGICAL | | | | | | | | | |
| Andersen 2015^20^ | Control (no intervention) | Type 1: 9 (22%) | 51.3 (24–65) | 4 | 26.9 (19.0–36.0) | --- | --- | 5.9 (2.0–9.0)^†^ | 27 (15–36) |
|  | Training + placebo supplement | Type 1: 13 (32%) | 45.7 (22–63) | 6 | 24.2 (19.0–33.0) | --- | --- | 6.2 (1.0–11.0)^†^ | 22 (10–31) |
|  | Training + protein supplement | Type 1: 13 (32%) | 42.6 (24–55) | 5 | 25.5 (20.0–37.0) | --- | --- | 5.2 (3.0–7.0)^†^ | 24 (15–36) |
| Andersen 2017^41^ | Control  (usual care) | Type 1: 6 (50%) | 46.0 ± 9.0 (32–59) | 1 | 27.0 ± 6.0 (20.0–36.0) | --- | --- | 7.5 ± 2.1  (5.0–11.0)^†^ | --- |
|  | Supervised HIT | Type 1: 6 (50%) | 53.0 ± 15.0 (26–67) | 2 | 26.0 ± 4.0 (22.0–31.0) | --- | --- | 6.3 ± 4.3  (1.0–11.0)^†^ | --- |
|  | Supervised HIT | HV: 6 (100%) | 52.0 ± 14.0 (27–64) | 2 | 26.0 ± 3.0 (24.0–32.0) | --- | --- | --- | --- |
| Bankolé  2016^21^ | Control (no  intervention) | Type 1: 8 (50%) | 41.0 ± 9.0 | 1 | --- | --- | --- | --- | --- |
|  | Training | Type 1: 8 (50%) | 40.0 ± 13.0 | 3 | --- | --- | --- | --- | --- |
|  | Control-training | Type 1: 8 (50%) | 42.0 ± 9.0 | 1 | --- | --- | --- | --- | --- |
| Colson  2010^42^ | NMES training | FSHD NS: 9 (100%) | 55.2 ± 10.4 (39–69) | 3 | --- | --- | --- | --- | 5–9 repeats |
| Doix  2017^22^ | NMES training | Type 1: 10 (100%) | 62.3 ± 10.2 | 5 (50%) | --- | --- | --- | --- | --- |
|  | NMES training | HV: 10 (100%) | 56.0 ± 4.8 | 7 (70%) | --- | --- | --- | --- | --- |
| Olsen  2005^235^ | AET | FSHD NS: 8 (100%) | 36.0 (18–55) | 4 | --- | --- | --- | --- | --- |
|  | AET | HV: 7 (100%) | 49.0 (35–60) | 3 | --- | --- | --- | --- | --- |
| Voet  2014^144^ | Control  (usual care) | Type 1: 24 (42%) | 52.0^‡^ (20–79) | NR (29%) | --- | --- | 16.7^‡^ (1–49) | 3.0^‡^ (1.5–4.0) | --- |
|  | AET | Type 1: 20 (35%) | 59.0^‡^ (21–68) | NR (40%) | --- | --- | 13.0^‡^ (1–42) | 3.0^‡^ (0.5–4.0) | --- |
|  | CBT^§^ | Type 1: 13 (23%) | 49.0^‡^ (24–69) | NR (38%) | --- | --- | 7.0^‡^ (0–40) | 2.0^‡^ (1.0–4.0) | --- |
| SURGICAL | | | | | | | | | |
| Andrews  1998^28^ | Scapulothoracic arthrodesis | FSHD NS: 6 (100%) | NR (17–33) | 3 | --- | NR (6–25) | --- | --- | --- |
| Bunch  1993^222^ | Scapulothoracic arthrodesis | FSHD NS: 12 (100%) | NR (17–48) | 7 | --- | --- | --- | --- | --- |
| Cakmak  2018^236^ | Scapulothoracic arthrodesis / spinal fusion | FSHD NS: 32 (100%) | 34.4 (11–73) | 16 (50%) | --- | 14.9 (2–50) | 19.5 (1–50) | 9.8 (1–15)^†^ | --- |
| Cooney  2014^43^ | Scapulothoracic arthrodesis | FSHD NS: 11 (100%) | 33.0 (18–50) | 5 | --- | --- | --- | --- | --- |
| Diab  2005^223^ | Scapulothoracic arthrodesis | FSHD NS: 8 (100%) | 17.0 (11–21) | 4 | --- | --- | --- | --- | --- |
| Eren  2020a^44^ | Scapulothoracic arthrodesis | FSHD NS: 40 (100%) | 25.4 (15–60) | --- | --- | --- | --- | --- | --- |
| Ersen  2018^45^ | Scapulothoracic arthrodesis | FSHD NS: 13 (100%) | 29.1 ± 8.1 (20–52) | 5 | --- | --- | --- | --- | --- |
| Giannini  2007^26^ | Scapulopexy | FSHD NS: 13 (100%) | 27.0 (15–43) | 7 | --- | --- | --- | --- | --- |
| Jakab  1993^27^ | Scapulothoracic arthrodesis | FSHD NS: 3 (100%) | NR (14–19) | 0 | --- | --- | --- | --- | --- |
| Le Hanneur 2017^46^ | Scapulothoracic arthrodesis | FSHD NS: 4 (100%) | NR (17–58) | 1 | --- | NR (15–35) | --- | --- | --- |
| Loock  2022^47^ | Scapulothoracic arthrodesis | FSHD NS: 7 (100%) | 31.6 (26–44) | 4 | --- | --- | --- | --- | --- |
| Van Tongel 2013^225^ | Scapulothoracic arthrodesis | FSHD NS: 21 (100%) | 26.0 (16–62) | 7 | --- | --- | --- | --- | --- |
| Ziaee  2006^48^ | Scapulothoracic arthrodesis | FSHD NS: 6 (100%) | 25.4 | 1 | --- | --- | 11.0 | --- | --- |
| OTHER | | | | | | | | | |
| Li  2021^23^ | Herbal medicine | Type 1: 1 (100%) | 15.0 | 1 | 16.2 | 9.0 | --- | 9.0 (2.0–9.0)^†^ | 2 repeats |
| Liu  2019^49^ | Acupuncture | FSHD NS: 1 (100%) | 62.0 | 1 | --- | 19.0 | --- | --- | --- |
| Ribot-Ciscar 2015^50^ | Vibratory proprioceptive assistance | FSHD NS: 8 (100%) | 58.0 ± 11.0 | 6 | --- | --- | --- | --- | --- |

Data are presented as mean ± SD or mean (range), unless otherwise specified.

AET, aerobic exercise training; BMI, body mass index; CBT, cognitive-behavioral therapy; CSS, Clinical Severity Score; FSHD, facioscapulohumeral muscular dystrophy; HIT, high intensity training; HV, healthy volunteer; NMES, neuromuscular electrical stimulation; NR, not reported; NS, not specified; SD, standard deviation.

^*^This study evaluated both pharmacological and physiological interventions.

^†^FSHD clinical score.

^‡^Median values.

^§^CBT was classified as other interventions but was presented under physiological interventions to centralize data from Voet 2014^144^.

Table 11. Description of interventions in “Treatment Outcomes and Clinical Trials”

| **Author, year** | **Treatment arm** | **Treatment detail** |
| --- | --- | --- |
| PHARMACOLOGICAL | | |
| Elsheikh 2007^232^ | Diltiazem hydrochloride | --- |
| Kissel 1998^39^ | Albuterol | Sustained-release albuterol 16 mg/day for 3 months |
| Kissel 2001^36^ | Placebo | --- |
|  | Albuterol low dose | Albuterol 8mg twice/day |
|  | Albuterol high dose | Albuterol 16mg twice/day |
| Lefkowitz 2012^40^ | Denosumab | Denosumab 3 injections |
| Passerieux 2015^37^ | Supplements | 500 mg vitamin C, 25 mg zinc gluconate, 200 μg selenomethionine, and 400 mg dl-alpha tocopheryl-acetate per day |
|  | Placebo | Microcrystalline cellulose |
| Payan 2009^138^ | Placebo | --- |
|  | Salbutamol | Oral salbutamol 16mg/day in periodic dosage regimen of 3 week on/1 week off |
| Statland 2022^38^ | Biceps brachii placebo | Placebo injected bilaterally in the biceps brachii muscle every 3 weeks for 2 weeks, total 9 doses |
|  | Biceps brachii ACE-083 | ACE-083 240mg per muscle injected bilaterally in the biceps brachii muscle every 3 weeks for 2 weeks, total 9 doses |
|  | Tibialis anterior placebo | Placebo injected bilaterally in the tibialis anterior muscle every 3 weeks for 2 weeks, total 9 doses |
|  | Tibialis anterior ACE-083 | ACE-083 240mg per muscle injected bilaterally in the tibialis anterior muscle every 3 weeks for 2 weeks, total 9 doses |
| Tawil 1997^233^ | Prednisone | 1.5 mg/kg/day, max 80 mg/day, for 12 weeks |
| Wagner 2008^35^ | Placebo | Formulation containing excipients |
|  | MYO-029 1 mg/kg | Delivered necessary volume of MYO-029 70 mg/ml |
|  | MYO-029 3 mg/kg |  |
|  | MYO-029 10 mg/kg |  |
|  | MYO-029 30 mg/kg |  |
| Xiao 2021^24^ | Supplement cocktail | Vitamin B1 tablets, vitamin B2 tablets, vitamin B6 tablets, vitamin C tablets, vitamin E tablets, idebenone tablets, etc. |
| PHARMACOLOGICAL & PHYSIOLOGICAL | | |
| Van der Kooi 2004^234^ | Placebo | Non-training + placebo |
|  | Placebo + training | Training (program below) + placebo |
|  | Albuterol | Non-training + albuterol (dose below) |
|  | Albuterol + training | Training: 3 non-consecutive days per week for 52 weeks, focused on elbow flexors and ankle dorsiflexors  Medication: albuterol sustained-release 8 mg twice daily |
| PHYSIOLOGICAL | | |
| Andersen 2015^20^ | Control | No intervention |
|  | Training + placebo supplement | Training (program below) + placebo supplement: Sukrin (0 kcal, 33 g carbohydrate, 0 g fat, 0 g protein) |
|  | Training + protein supplement | Training: 3 times weekly (36 sessions) on an upright stationary bike. Training sessions lasted 15 minutes the first week, 20 minutes the second week, and 30 minutes thereafter  Protein supplement: 1 protein-carbohydrate beverage after each training session, containing 163 kcal (23 g whey protein, 17 g carbohydrate, 0 g fat) |
| Andersen 2017^41^ | Control | Usual care |
|  | Supervised high intensity training | Each session lasted 21 min including an 8-min standardized warm-up and two sets of 5-min HIT separated by a 3-min break at very low intensity |
|  | Healthy volunteers + supervised high intensity training | Training program as above |
| Bankolé 2016^21^ | Control | No intervention |
|  | Training | Cycling 3 times weekly for 35 minutes (combination of strength, high-intensity interval, and low-intensity aerobic) at home for 24 weeks |
|  | Control-training | Patients from Control group followed training (program above) from week 24 to 48 |
| Colson 2010^42^ | Neuromuscular electrical stimulation training | 20-minute sessions, 5 sessions/week for 5 months |
| Doix 2017^22^ | Neuromuscular electrical stimulation training | 3 sessions/week for 8 weeks. Each session included 2-min warm-up, 20-min work out, and 3-min relaxation |
|  | Healthy volunteers + neuromuscular electrical stimulation training | Training program as above |
| Olsen 2005^235^ | Aerobic exercise training | 35-min sessions, up to 5 sessions/week for 12 weeks, total 44 sessions |
|  | Healthy volunteers + aerobic exercise training | Training program as above |
| Voet 2014^144^ | Control | Usual care |
|  | Aerobic exercise training | 3 weekly sessions of aerobic cycling exercises on a Monark 827E ergometer |
|  | Cognitive-behavioral therapy | 50-minute sessions focused on 6 possible modules based on the known fatigue-perpetuating factors |
| SURGICAL | | |
| Andrews 1998^28^ | Scapulothoracic arthrodesis | Surgical technique by Bunch and Siegel |
| Bunch 1993^222^ | Scapulothoracic arthrodesis | --- |
| Cakmak 2018^236^ | Scapulothoracic arthrodesis / spinal fusion | Scapulothoracic arthrodesis or spinal stenosis |
| Cooney 2014^43^ | Scapulothoracic arthrodesis | Scapulothoracic arthrodesis with cerclage wires, plates, and allograft |
| Diab 2005^223^ | Scapulothoracic arthrodesis | --- |
| Eren 2020a^44^ | Scapulothoracic arthrodesis | --- |
| Ersen 2018^45^ | Scapulothoracic arthrodesis | Scapulothoracic fusion using multifilament cables |
| Giannini 2007^26^ | Scapulopexy | Scapulothoracic fixation without arthrodesis |
| Jakab 1993^27^ | Scapulothoracic arthrodesis | Scapulothoracic arthrodesis using 16-gauge Luque wires for fixation |
| Le Hanneur 2017^46^ | Scapulothoracic arthrodesis | Letournel surgical technique |
| Loock 2022^47^ | Scapulothoracic arthrodesis | --- |
| Van Tongel 2013^225^ | Scapulothoracic arthrodesis | Scapulothoracic arthrodesis with screw ﬁxation and bone-grafting |
| Ziaee 2006^48^ | Scapulothoracic arthrodesis | Plate and wire fixation of the scapula to the ribs with corticocancellous iliac bone graft |
| OTHER | | |
| Li 2021^23^ | Herbal medicine | Buzhong Yiqi once/day for 20 days |
| Liu 2019^49^ | Acupuncture | 3 times/week during 5 months in 2 consecutive years, total 10 months treatment |
| Ribot-Ciscar 2015^50^ | Vibratory proprioceptive assistance | 8 sessions of 40-min stimulation on the more affected side of the body |

Table 12. Effectiveness / efficacy of pharmacological interventions for prioritized outcomes of extracted “Treatment Outcomes and Clinical Trials” studies

| **Author, year** | **Treatment arm** | **Follow-up** | **Motor function** | **Muscle strength & endurance** | **Other** |
| --- | --- | --- | --- | --- | --- |
| TRIALS (NON-RANDOMIZED AND RANDOMIZED) | | | | | |
| Elsheikh 2007^232^ | Diltiazem hydrochloride (n=19) | 24 weeks | Walk 30 ft (time)^*^: -0.36 ± 1.06 | MMT AMS: -0.0025 ± 0.1320  MVIC^*^: +0.11 ± 2.08 | --- |
| Kissel  2001^36^ | Placebo  (n=30) | 52 weeks | 4-stair climb: no changes  Functional grading: no changes  Walk 30 ft: no changes | Grip strength: -0.53 ± 4.13  MMT: -0.04 ± 0.16  MVIC: +0.20 ± 0.91 | --- |
|  | Albuterol low dose (n=30) | 52 weeks | 4-stair climb: no changes  Functional grading: no changes  Walk 30 ft: no changes | Grip strength: +1.90 ± 3.34  **ES: +2.49 (-0.05, 5.02)**^†^  MMT: -0.03 ± 0.13  MVIC: -0.04 ± 0.84 | --- |
|  | Albuterol high dose (n=30) | 52 weeks | 4-stair climb: no changes  Functional grading: no changes  Walk 30 ft: no changes | Grip strength: +1.70 ± 4.13  **ES: +2.31 (-0.29, 4.92)**^†^  MMT: +0.00 ± 0.15  MVIC: +0.08 ± 0.98 | --- |
| Passerieux 2015^37^ | Supplements  (n=26) | 17 weeks | --- | Endurance T_lim_ (MM:SS)^*^:  Dominant leg: **+02:52 ± 03:47**  Non-dominant: **+02:48 ± 04:05**  MVC (kg)^*^:  Dominant leg: **+1.15 ± 1.69**  Non-dominant: **+1.30 ± 2.39** | 2MWT (m)^*^: +8.36 ± 14.14 |
|  | Placebo  (n=27) | 17 weeks | --- | Endurance T_lim_ (MM:SS)^*^:  Dominant leg: -00:18 ± 04:28  Non-dominant: -00:20 ± 02:16  MVC (kg):  Dominant leg: +0.02 ± 1.35  Non-dominant: -0.30 ± 1.30 | 2MWT (m): +4.27 ± 8.16 |
| Payan  2009^138^ | Placebo  (n=56) | 6 months | 4-stair climb (s): -0.3 ± 1.9  Barré test (s): -3.8 ± 48.5  Brooke score: +0.0 ± 0.4  Mingazzini test (s): +1.9 ± 43.0  Vignos score: +0.1 ± 0.6  Walk 10 m (s): -0.20 ± 4.00 | MMT^*^:  Upper limb: +0.03 ± 0.18  Lower limb: +0.04 ± 0.19  Total: +0.03 ± 0.16  QMT Z-score^*^:  Upper limb: +0.02 ± 0.31  Lower limb: -0.04 ± 0.36  Total: +0.01 ± 0.27 | --- |
|  | Salbutamol  (n=56) | 6 months | 4-stair climb (s): -0.1 ± 1.3  Barré test (s): -0.1 ± 38.1  Brooke score: +0.1 ± 0.4  Mingazzini test (s): +2.8 ± 34.1  Vignos score: +0.1 ± 0.5  Walk 10 m (s): +0.03 ± 2.40 | MMT^*^:  Upper limb: +0.04 ± 0.21  Lower limb: +0.05 ± 0.20  Total: +0.04 ± 0.18  QMT Z-score^*^:  Upper limb: +0.08 ± 0.26  Lower limb: -0.01 ± 0.49  Total: +0.05 ± 0.28 | --- |
| Statland  2022^38^ | Biceps brachii placebo  (n=14) | 190 days | PUL (%)^‡^: -1.2 ± 1.2 | MMT MRC (%)^‡^: +0.20 ± 1.62  MVIC (%)^‡^: -3.54 ± 10.00 | CMV (%)^‡^: +2.60 ± 5.16  Muscle fat fraction (%)^‡^: +1.00 ± 0.96  TMV (%)^‡^: +2.70 ± 2.81 |
|  | Biceps brachii  ACE-083  (n=14) | 190 days | PUL (%)^‡^: +1.7 ± 1.2  ES: +2.9 ± 1.7 | MMT MRC (%)^‡^: -2.48 ± 1.75  ES: -2.68 ± 2.39  MVIC (%)^‡^: +32.58 ± 10.44  **ES: +36.12 ± 14.18** | CMV (%)^‡^: +25.80 ± 5.45  **ES: +23.30 ± 7.59**  Muscle fat fraction (%)^‡^: -0.22 ± 0.98  ES: -1.30 ± 1.36  TMV (%)^‡^: +19.10 ± 2.82  **ES:** **+16.40 ± 4.03** |
|  | Tibialis anterior placebo  (n=14) | 190 days | 4-stair climb (%)^‡^: -5.20 ± 4.07  Walk/run 10 m (%)^‡^: -8.60 ± 3.35 | MMT MRC (%)^‡^: +1.97 ± 2.15  MVIC (%)^‡^: +8.72 ± 7.76 | 6MWT (%)^‡^: +8.60 ± 2.76  CMV (%)^‡^: +5.60 ± 4.89  Muscle fat fraction (%)^‡^: -0.32 ± 0.89  TMV (%)^‡^: +4.30 ± 2.72 |
|  | Tibialis anterior  ACE-083  (n=13) | 190 days | 4-stair climb (%)^‡^: -4.80 ± 4.32  ES: +0.45 ± 6.03  Walk/run 10 m (%)^‡^: -3.90 ± 3.59  ES: +4.70 ± 4.97 | MMT MRC (%)^‡^: +2.84 ± 2.32  ES: +0.88 ± 3.17  MVIC (%)^‡^: -5.82 ± 8.37  ES: -14.54 ± 11.50 | 6MWT (%)^‡^: +3.30 ± 2.94  ES: -5.30 ± 4.07  CMV (%)^‡^: +24.00 ± 5.24  **ES: +18.40 ± 7.01**  Muscle fat fraction (%)^‡^: -3.10 ± 0.95  **ES: -2.70 ± 1.30**  TMV (%)^‡^: +13.80 ± 2.85  **ES: +9.50 ± 3.88** |
| Tawil  1997^233^ | Prednisone  (n=8) | 12 weeks | --- | MMT: -0.01 (-0.24, 0.23)  MVIC: -0.24 (-0.90, 0.43) | --- |
| Van der Kooi  2004^234^ | Placebo  (n=16) | 52 weeks | 3-stair climb: no changes  Functional grading: no changes  Standing from lying: no changes  Standing from sitting: no changes  Walk 30 ft: no changes | 1RM (kg):  Elbow:  BL: 6.5 (4.8, 8.2)  FU: 7.5 (5.8, 9.2)  Ankle:  BL: 5.9 (4.4, 7.5)  FU: 4.7 (3.2, 6.2)  FTI30 (kgF·s):  Elbow:  BL: 281 (211, 352)  FU: 278 (207, 349)  Ankle:  BL: 189 (133, 245)  FU: 160 (104, 216)  MVIC (kgF):  Elbow:  BL: 17.3 (13.7, 21.0)  FU: 16.0 (12.4, 19.7)  Ankle:  BL: 10.8 (7.8, 13.9)  FU: 9.6 (6.5, 12.6) | --- |
|  | Placebo + training (n=19) | 52 weeks | 3-stair climb: no changes  Functional grading: no changes  Standing from lying: no changes  Standing from sitting: no changes  Walk 30 ft: no changes | 1RM (kg):  Elbow:  BL: 5.7 (4.0, 7.4)  FU: 7.8 (6.1, 9.5)  Ankle:  BL: 5.4 (4.1, 6.7)  FU: 3.9 (2.6, 5.2)  FTI30 (kgF·s):  Elbow:  BL: 221 (156, 286)  FU: 210 (145, 275)  Ankle:  BL: 205 (155, 256)  FU: 159 (109, 210)  MVIC (kgF):  Elbow:  BL: 12.6 (9.3, 16.0)  FU: 12.2 (8.8, 15.5)  Ankle:  BL: 10.4 (7.6, 13.2)  FU: 8.7 (5.9, 11.5) | --- |
|  | Albuterol  (n=15) | 52 weeks | 3-stair climb: no changes  Functional grading: no changes  Standing from lying: no changes  Standing from sitting: no changes  Walk 30 ft: no changes | 1RM (kg):  Elbow:  BL: 6.0 (4.1, 7.9)  FU: 7.8 (5.9, 9.7)  Ankle:  BL: 5.9 (4.5, 7.4)  FU: 5.1 (3.7, 6.5)  FTI30 (kgF·s):  Elbow:  BL: 259 (185, 332)  FU: 264 (191, 338)  Ankle:  BL: 195 (139, 252)  FU: 144 (87, 202)  MVIC (kgF):  Elbow:  BL: 14.0 (10.2, 17.8)  FU: 14.1 (10.3, 17.9)  Ankle:  BL: 9.9 (6.7, 13.1)  FU: 8.0 (4.8, 11.2) | --- |
|  | Albuterol + training (n=15) | 52 weeks | 3-stair climb: no changes  Functional grading: no changes  Standing from lying: no changes  Standing from sitting: no changes  Walk 30 ft: no changes | 1RM (kg):  Elbow:  BL: 5.8 (3.9, 7,6)  FU: 8.9 (7.0, 10.7)  Ankle:  BL: 6.2 (4.7, 7.6)  FU: 4.6 (3.2, 6.1)  FTI30 (kgF·s):  Elbow:  BL: 241 (166, 316)  FU: 259 (185, 334)  Ankle:  BL: 181 (123, 239)  FU: 149 (90, 207)  MVIC (kgF):  Elbow:  BL: 15.1 (11.3, 19.0)  FU: 15.4 (11.6, 19.3)  Ankle:  BL: 9.7 (6.4, 12.9)  FU: 9.0 (5.8, 12.3) | --- |
| Wagner  2008^35^ | Placebo  (n=29) | 26 weeks | 4-stair climb: no changes  Standing from sitting: no changes  Walk 9 m: no changes | MMT (%)^‖^: +2.87 ± 1.33  QMT: no changes | Muscle fiber diameter (%)^*¶^: +2.7  Muscle histology: no changes in central nucleated fibers, fiber necrosis, inflammation, fibrosis, regeneration  Muscle volume (%): +0.7 ± 0.8  Myostatin levels: no changes |
|  | MYO-029 1 mg/kg (n=27) | 26 weeks | 4-stair climb: no changes  Standing from sitting: no changes  Walk 9 m: no changes | MMT (%)^‖^: +1.99 ± 2.71  QMT: no changes | Muscle fiber diameter (%)^*¶^: -0.9  Muscle histology: no changes in central nucleated fibers, fiber necrosis, inflammation, fibrosis, regeneration  Muscle volume (%): -0.6 ± 0.9  Myostatin levels: no changes |
|  | MYO-029 3 mg/kg (n=27) | 26 weeks | 4-stair climb: no changes  Standing from sitting: no changes  Walk 9 m: no changes | MMT (%)^‖^: +2.22 ± 1.67  QMT: no changes | Muscle fiber diameter (%)^*¶^: +14.4  Muscle histology: no changes in central nucleated fibers, fiber necrosis, inflammation, fibrosis, regeneration  Muscle volume (%): +2.1 ± 1.0  Myostatin levels: no changes |
|  | MYO-029 10 mg/kg (n=27) | 26 weeks | 4-stair climb: no changes  Standing from sitting: no changes  Walk 9 m: no changes | MMT (%)^‖^: +5.28 ± 5.31  QMT: no changes | Muscle fiber diameter (%)^*¶^: +15.2  Muscle histology: no changes in central nucleated fibers, fiber necrosis, inflammation, fibrosis, regeneration  Muscle volume (%): +1.2 ± 1.1  Myostatin levels: no changes |
|  | MYO-029 30 mg/kg (n=6) | Discontinued | | | |
| OBSERVATIONAL | | | | | |
| Kissel  1998^39^ | Albuterol  (n=15) | 12 weeks | --- | MMT: +0.06 ± 0.13  MVIC: **+0.33 ± 0.60**^§^ | --- |
| CASE STUDY | | | | | |
| Lefkowitz  2012^40^ | Denosumab  (n=1) | --- | Standing from sitting (n times in 30 s):  2^nd^ injection:  Prior: 6  24h: 15  3^rd^ injection:  Prior: 9.5  24h: 18 | Grip strength (lbs. sq. in.):  Right hand:  2^nd^ injection:  Prior: 45.0 ± 3.7  **24h: 54.8 ± 3.9**^§^  3^rd^ injection:  Prior: 39.8 ± 5.1  **24h: 53.2 ± 2.1**^§^  Left hand:  2^nd^ injection:  Prior: 45.0 ± 3.5  **24h: 49.3 ± 1.0**^§^  3^rd^ injection:  Prior: 35.3 ± 3.7  **24h: 56.3 ± 1.5**^§^ | TUG (s):  2^nd^ injection:  Prior: 11  24h: 6  3^rd^ injection:  Prior: 27  24h: 5 |
| Xiao  2021^24^ | Supplement cocktail  (n=2) | --- | --- | Muscle weakness: no improvement | --- |

Data are presented as mean ± SD or mean (95% CI), unless otherwise specified. Statistical significance between treatment and control arms (unless otherwise specified) are presented in bold.

1RM, 1 repetition maximum; 2MWT, 2-minute walking test; 6MWT, 6-minute walking test; AMS, average muscle score; BL, baseline; CI, confidence interval; CMV, contractile muscle volume; ES, effect size; ft, feet; FTI30, force-time integral 30; FU, follow-up; MMT, manual muscle testing; MVC, maximal voluntary contraction; MVIC, maximal voluntary isometric contraction; PUL, performance of the upper limb; QMT, quantitative muscle testing; SD, standard deviation; SE, standard error; SEM, standard error of the mean; T_lim_, limit time; TMV, total muscle volume; TUG, timed up and go.

^*^Not full sample size.

^†^Data presented as mean (98.3% CI).

^‡^Data presented as mean ± SEM.

^§^Statistical significance of follow-up versus baseline of the same treatment arm.

^‖^Only FSHD patients. Data presented as mean ± SE.

^¶^Median values.

Table 13. Effectiveness / efficacy of physiological interventions for prioritized outcomes of extracted “Treatment Outcomes and Clinical Trials” studies

| **Author, year** | **Treatment arm** | **Follow-up** | **Timed function tests** | **Muscle strength & endurance** | **Other** |
| --- | --- | --- | --- | --- | --- |
| TRIALS (NON-RANDOMIZED AND RANDOMIZED) | | | | | |
| Andersen 2015^20^ | Control (no intervention)  (n=9) | 12 weeks | Walking speed via 6MWT (%)^*^: +0 | Dynamometry (n):  Elbow flexion:  BL: 93 (1–192)  FU: 92 (1–185)  Elbow extension:  BL: 77 (1–173)  FU: 83 (1–207)  Knee flexion:  BL: 129 (13–229)  FU: 136 (22–337)  Knee extension:  BL: 128 (42–238)  FU: 160 (18–306) | 5-times-sit-to-stand-test (s):  BL: 21 (11–34)  FU: 19 (11–27)  14-step-stair-test (s):  Usual pace:  BL: 52 (22–102)  FU: 48 (19–89)  Fast pace:  BL: 40 (13–81)  FU: 40 (13–76)  Standing balance test (s, max 30 s):  BL: 30.0 (29.0–30.0)  FU: 30.0 |
|  | Training + placebo supplement  (n=13) | 12 weeks | Walking speed via 6MWT (%): +4 | Dynamometry (n)^†^:  Elbow flexion:  BL: 130 (1–293)  FU: 146 (14–293)  Elbow extension:  BL: 83 (1–273)  FU: 98 (2–273)  Knee flexion:  BL: 140 (9–369)  FU: 140 (1–377)  Knee extension:  BL: 205 (5–582)  FU: 19 (10–513) | 5-times-sit-to-stand-test (s)^†^:  BL: 13 (7–27)  FU: 14 (7–23)  14-step-stair-test (s)^†^:  Usual pace:  BL: 41 (14–154)  FU: 41 (15–120)  Fast pace:  BL: 28 (7–89)  FU: 25 (7–70)  Standing balance test (s, max 30 s)^†^:  BL: 28.6 (25.0–30.0)  FU: 28.9 (20.0–30.0) |
|  | Training + protein supplement  (n=13) | 12 weeks | Walking speed via 6MWT (%)^*^: +9 | Dynamometry (n)^†^:  Elbow flexion:  BL: 96 (9–210)  FU: 95 (0–197)  Elbow extension:  BL: 87 (8–224)  FU: 83 (3–224)  Knee flexion:  BL: 147 (18–278)  FU: 156 (18–299)  Knee extension:  BL: 186 (60–301)  FU: 209 (78–358) | 5-times-sit-to-stand-test (s)^†^:  BL: 15 (9–18)  FU: 13 (8–21)  14-step-stair-test (s)^†^:  Usual pace:  BL: 27 (16–66)  FU: 26 (17–66)  Fast pace:  BL: 21 (11–56)  FU: 20 (12–56)  Standing balance test (s, max 30 s)^†^:  BL: 30.0  FU: 29.6 (25.0–30.0) |
| Andersen  2017^41^ | Control (usual care) (n=6) | 8 weeks | 6MWT (m): **+21 ± 16**^§^ | --- | --- |
|  | Supervised HIT  (n=6) | 8 weeks | 6MWT: no changes | Dynamometry: no changes | 5-times-sit-to-stand-test: no changes |
|  | HV + Supervised HIT (n=6) | 8 weeks | 6MWT: no changes | Dynamometry: no changes | 5-times-sit-to-stand-test: no changes |
| Bankolé  2016^21^ | Control (no intervention)  (n=8) | 24 weeks  (week 0 to 24) | 6MWT (%): -0.5 (-4.3, 3.3) | Muscle endurance (%): -3.2 (-11.0, 4.6)  MVIC (%): -1.5 (-5.8, 2.8) | Enzyme activity in muscle biopsy (%):  CK: -7.2 (-26.6, 12.2)  CS: +3.8 (-28.7, 36.4)  LDH: +12.9 (-10.0, 35.8)  Muscle cross-sectional area (%):  Type 1 fibers: -4.0 (-20.6, 12.5)  Type 2 fibers: -4.3 (-13.7, 5.2)  Whole muscle: +0.1 (-11.1, 11.2) |
|  | Training  (n=8) | 24 weeks  (week 0 to 24) | 6MWT (%): +13.3 (4.6, 21.9)  **ES: +13.7 (4.1, 23.3)** | Muscle endurance (%): +19.4 (4.8, 34.0)  **ES: +22.6 (5.7, 39.6)**  MVIC (%): +13.6 (3.9, 23.2)  **ES: +15.0 (4.3. 25.9)** | Enzyme activity in muscle biopsy (%):  CK: +15.2 (4.7, 25.6)  ES: +22.4 (-0.1, 44.9)  CS: +46.1 (25.8, 66.4)  **ES: +42.3 (3.1, 81.4)**  LDH: -6.3 (-20.1, 7.3)  ES: -19.3 (-46.5, 7.9)  Muscle cross-sectional area (%):  Type 1 fibers: +28.4 (12.6, 44.2)  **ES: +32.4 (9.1, 55.8)**  Type 2 fibers: +39.8 (19.7, 59.8)  **ES: +44.0 (21.4, 66.7)**  Whole muscle: +33.7 (15.4, 52.0)  **ES: +33.6 (11.8, 55.5)** |
|  | Control-training (n=8) | 24 weeks  (week 24 to 48) | 6MWT (%): **+13.0**^**^ | Muscle endurance (%): **+11.0**^**^  MVIC: no changes | --- |
| Voet  2014^144^ | Control (usual care)  (n=24) | 28 weeks | 6MWT (m)^†‖^:  BL: 436 (80–708)  FU: 461 (86–832) | MVIC^†‖^:  BL: 19.7 (1.9–40.3)  FU: 17.7 (0.9–58.0) | --- |
|  | AET  (n=20) | 28 weeks | 6MWT (m)^†‖^:  BL: 388 (136–630)  FU: 450 (157–622) | MVIC^†‖^:  BL: 21.0 (10.0–42.7)  FU: 24.8 (5.6–47.4) | --- |
|  | CBT^¶^  (n=13) | 28 weeks | 6MWT (m)^‖^:  BL: 457 (178–650)  FU: 459 (120–733) | MVIC^‖^:  BL: 21.6 (9.2–40.7)  FU: 20.7 (9.6–39.1) | --- |
| OBSERVATIONAL | | | | | |
| Colson  2010^42^ | NMES training  (n=9) | 5 months | 6MWT (m)^‡^:  BL: 305.7 ± 40.1  **FU: 333.8 ± 44.9**^§^ | MMT (%)^‡^:  Shoulder flexion:  Right: **+11.2 ± 5.6**^§^  Left: **+15.8 ± 7.4**^§^  Shoulder extension:  Right: **+13.8** **± 5.7**^§^  Left: **+16.3** **± 4.4**^§^  Knee extension:  Right: **+14.3 ± 5.5**^§^  Left: **+14.0** **± 6.7**^§^  MVIC (%)^‡^:  Shoulder abduction:  Right: **+45.5 ± 20.0**^§^  Left: **+42.5 ± 13.2**^§^  Shoulder flexion:  Left: **+83.7 ± 35.9**^§^ | --- |
| Doix  2017^22^ | NMES training (n=10) | 8 weeks | 6MWT (m):  BL: **309.67 ± 132.14**  FU: **311.11 ± 126.88** | MMT: slight increase  MVC: no significant changes | MFM:  BL: **68.86 ± 19.35**  FU: **66.97 ± 17.42** |
|  | HV + NMES training  (n=10) | 8 weeks | 6MWT (m):  BL: 462.22 ± 113.66  FU: 475.25 ± 131.07 | MVC: no significant changes | MFM:  BL: 100 ± 0  FU: 100 ± 0 |
| Olsen  2005^235^ | AET  (n=8) | 12 weeks | --- | --- | Muscle fiber area (um^2^)^‡^:  BL: 6648 ± 1026  FU: 6802 ± 399  Muscle fiber distribution (%)^‡^:  Type 1:  BL: 51 ± 7  FU: 46 ± 9  Type 2:  BL: 49 ± 7  FU: 54 ± 9 |
|  | HV + AET  (n=7) | 12 weeks | --- | --- | Muscle fiber area (um^2^)^‡^:  BL: 4175 ± 432  FU: 4750 ± 289  Muscle fiber distribution (%)^‡^:  Type 1:  BL: 62 ± 5  FU: 64 ± 13  Type 2:  BL: 38 ± 5  FU: 36 ± 13 |

Data are presented as mean ± SD or mean (range / 95% CI), unless otherwise specified; lower and upper limits of the ranges and 95% CI are separated by a dash or a comma, respectively. Statistical significance between treatment and control arms (unless otherwise specified) are presented in bold.

6MWT, 6-minute walking test; AET, aerobic exercise training; BL, baseline; CBT, cognitive-behavioral therapy; CI, confidence interval; CIS, Checklist Individual Strength; CK, creatine kinase; CS, citrate synthase; ES, effect size; FU, follow-up; HIT, high intensity training; HV, healthy volunteer; LDH, lactate dehydrogenase; MFM, motor function measurement; MMT. manual muscle testing; MVC, maximal voluntary contraction; MVIC, maximal voluntary isometric contraction; NMES, neuromuscular electrical stimulation; SD, standard deviation; SE, standard error; SEM, standard error of the mean.

^*^Full sample size + dropout patients.

^†^Not full sample size.

^‡^Data presented as mean ± SE.

^§^Statistical significance of follow-up versus baseline of the same treatment arm.

^‖^Data presented as median (range).

^¶^CBT was classified as other interventions but was presented here to centralize data from Voet 2014^144^.

^**^Statistical significance compared to control (no intervention) arm at week 24.

Table 14. Effectiveness / efficacy of surgical interventions for prioritized outcomes of extracted “Treatment Outcomes and Clinical Trials” studies

| **Author, year** | **Treatment arm** | **Follow-up** | **Shoulder mobility** | **Shoulder strength** | **Other** |
| --- | --- | --- | --- | --- | --- |
| OBSERVATIONAL | | | | | |
| Andrews 1998^28^ | Scapulothoracic arthrodesis  (n=6) | 28–120 months | Shoulder abduction (°): +46.5 (5.0–75.0)  Shoulder flexion (°): +60.5 (20.0–75.0) | --- | Activities of daily living: data per patient (only available per patient) |
| Bunch 1993^222^ | Scapulothoracic arthrodesis  (n=12) | 3–21 years | Shoulder abduction (°):  BL: 65 ± 10  FU: 125 ± 25 | --- | Activities of daily living: improved (descriptive outcomes) |
| Cooney 2014^43^ | Scapulothoracic arthrodesis  (n=11) | 29 months  (6–50) | Shoulder abduction (°):  BL: 68 (30–90)  **FU: 109 (90–120)**  Shoulder flexion (°):  BL: 72 (30–90)  **FU: 117 (90–130)** | --- | Preoperative DASH score:  BL: 48.0 (27.0–74.0)  **FU: 34.0** (0.8–70.0) |
| Diab  2005^223^ | Scapulothoracic arthrodesis  (n=8) | 6.3 years  (2–10) | Shoulder abduction (°):  BL: 75 (70–90)  1 year: 145 (110–160)  Last FU:  7 shoulders: 139 (110–150)   7 shoulders: 48 (20–70)   Shoulder flexion (°):  BL: 75 (70–90)  1 year: 144 (130–160)  Last FU:  7 shoulders: 134 (100–150)  4 shoulders: 48 (20–70) | --- | Clinical grading (n patients [n shoulders])^†^:  Score 2: 1 (2)  Score 3: 3 (3)  Score 6: 5 (6) |
| Eren 2020a^44^ | Scapulothoracic arthrodesis  (n=40) | 71.2 months (12–185) | Shoulder abduction (°):  BL: 52.7 ± 15.8  **FU: 98.8 ± 20.3**  Shoulder elevation (°):  BL: 60.6 ± 17.2  **FU: 123.7 ± 26.7** | --- | qDASH score:  BL: 34.7 ± 11.4  **FU: 13.3 ± 13.1** |
| Ersen 2018^45^ | Scapulothoracic arthrodesis  (n=13) | 128 months (94–185) | Shoulder abduction (°):  BL: 47.2 ± 16.1  **FU: 101.9 ± 8.0**  Shoulder flexion (°):  BL: 55.6 ± 16.1  **FU: 124.1 ± 12.0** | --- | qDASH score:  BL: 33.8 ± 8.8  **FU: 9.1 ± 5.0** |
| Giannini 2007^26^ | Scapulopexy  (n=13)^*^ | 10 years  (3–18) | Shoulder abduction (°):  BL: 70 ± 20  1 year: 98 ± 18  Last FU: 90 ± 17  Shoulder flexion (°):  BL: 55 ± 16  1 year: 120 ± 33  Last FU: 105 ± 22 | MRC score:  Flexion:  BL: 2.4  1 year: 4.5  Last FU: 4.1  Abduction:  BL: 2.5  1 year: 4.5  Last FU: 4.0 | Activities of daily living: limited at BL |
| Le Hanneur 2017^46^ | Scapulothoracic arthrodesis  (n=4) | 14 years ± 73 months | Shoulder abduction (°): **+36 ± 32**  Shoulder extension (°):  FU: 46 ± 28  Shoulder external rotation (°):  FU: 23 ± 9  Shoulder flexion (°): **+40 ± 44**  Shoulder internal rotation (°):  FU: 103 ± 12  Shoulder abduction strength (kg): 4.6 | Constant score (%): 57 ± 25 | Activities of daily living: improved (descriptive outcomes) |
| Loock 2022^47^ | Scapulothoracic arthrodesis  (n=7) | 7.3 years  (3.5–13.0) | Shoulder abduction (°):  BL: 63 ± 9  **FU: 92 ± 13**  Shoulder external rotation (°):  BL: 49 ± 19  FU: 43 ± 10  Shoulder flexion (°):  BL: 64 ± 11  **FU: 113 ± 20**  Shoulder internal rotation:  BL: T9 (S1–T2)  FU: T9 (S1–T2) | Constant score (max 100):  FU: 71.8 ± 16.0  MRC score (max 5):  BL: 2–4  FU: 3–4 | Brooke score (n patients):  Score 2: 2  Score 3: 5  Vignos score (n patients):  Score 1: 2  Score 2: 3  Score 3: 1  Score 7: 1 |
| Van Tongel 2013^225^ | Scapulothoracic arthrodesis  (n=21) | 88 months  (24–174) | Shoulder elevation (°):  BL: 65  FU: 119  Shoulder motion range (°, max 40):  BL: 14.6 (8.0–22.0)  FU: 25.6 (12.0–36.0) | Strength (max 25):  BL: 0  FU: 8 (0–20)  Constant score:  BL: 30 (17–41)  FU: 61 (30–90) | Activities of daily living score (max 20):  BL: 5.4 (2.0–13.0)  FU: 14.6 (6.0–20.0) |
| CASE STUDY | | | | | |
| Cakmak 2018^236^ | Scapulothoracic arthrodesis  (n=4) | --- | Shoulder elevation (°):  BL: 70 (45–120)  FU: 132 (90–150) | --- | --- |
| Jakab 1993^27^ | Scapulothoracic arthrodesis  (n=3) | 2.9 years  (2.5–3.3) | Shoulder abduction (°): +27 (20–35)  Shoulder flexion (°): +28 (20–40) | --- | Activities of daily living: improved |
| Ziaee 2006^48^ | Scapulothoracic arthrodesis  (n=6) | 32.5 months (14–55) | Shoulder abduction (°):  BL: 67.5  FU: 125  Shoulder flexion (°):  BL: 64  FU: 104 | --- | Activities of daily living: improved (descriptive outcomes) |

Data are presented as mean ± SD or mean (range), unless otherwise specified. Statistically significant changes at follow-up versus baseline of the same treatment arm are presented in bold.

---, data not reported; BL, baseline; DASH, Disabilities of the Arm, Shoulder, and Hand; FU, follow-up; MRC, Medical Research Council; SD, standard deviation.

^*^Expansion study that includes the nine patients from Giannini 2006^224^.

^†^Clinical grading included scoring of three criteria: shoulder range of motion; appearance; fatigue and pain.

Table 15. Effectiveness / efficacy of other interventions for prioritized outcomes of extracted “Treatment Outcomes and Clinical Trials” studies

| **Author, year** | **Treatment arm** | **Follow-up** | **Shoulder mobility** | **Muscle strength** | **Other** |
| --- | --- | --- | --- | --- | --- |
| Li  2021^23^ | Herbal medicine  (n=1) | 20 days | Shoulder abduction (°):  FU: >90 | Muscle strength (Oxford Scale grade, max 5):  BL: 3–4  FU: 4–5 | **---** |
| Liu  2019^49^ | Acupuncture  (n=1) | 1.5 year | --- | Muscle strength:  Facial: improved  Extremities: no changes | **---** |
| Ribot-Ciscar 2015^50^ | Vibratory proprioceptive assistance  (n=8) | 36 days | Shoulder abduction (°): increased^*^  Shoulder flexion (°): **increased**^*^ | Maximum isometric abduction force (kg):  BL: 1.7 ± 0.3  **FU: 2.4 ± 0.3** | Constant score:  BL: 44.8 ± 3.8  **FU: 54.4 ± 3.9** |

Data are presented as mean ± SD or ranges, unless otherwise specified. Statistically significant changes at follow-up versus baseline of the same treatment arm are presented in bold.

BL, baseline; FU, follow-up; SD, standard deviation.

^*^A figure illustrating the effect magnitude is available in the publication.

#### 7.3 Detailed tables for “Disease Outcome Measures and Validation”

Table 16. Population characteristics for the topic “Disease Outcome Measures and Validation”

| **Author, year** | **FSHD population type: n (%)** | **Age (years)** | **% pediatric** | **Female, n (%)** | **BMI (kg/m^2^)** | **Age at onset (Years)** | **Disease duration (years)** | **CSS** | **D4Z4 pairs (kb)** |
| --- | --- | --- | --- | --- | --- | --- | --- | --- | --- |
| CONFERENCE ABSTRACTS | | | | | | | | | |
| Berggren 2022^80^ | FSHD NS: 227 (100%) | 50.3 ± 14.1 | --- | --- | --- | --- | --- | --- | --- |
| Chan 2021^81^ | FSHD NS: 22 (100%) | --- | --- | --- | --- | --- | --- | --- | --- |
| Chan 2020^82^ | FSHD NS: 22 (100%) | --- | --- | --- | --- | --- | --- | NR (0 – 5)^a^ | --- |
| De Lattre 2016^74^ | FSHD NS: 269 (100%) | 6.1 – 82.8 | NS | 46.1% | --- | 24.5 ± 15.9 | --- | --- | --- |
| Eichinger 2022^83^ | FSHD NS: 240 (100%) | 50.5 (19-75) | --- | 45% | --- | --- | --- | --- | --- |
| Eichinger 2021^84^ | FSHD NS: 133 (100%) | 49.9 (19-75) | 0% | 44% | --- | --- | --- | FCS: 7.32 (1 – 15) | --- |
| Eichinger 2017^85^ | FSHD NS: 36 (100%) | 52.7 ±11.6 (22.2-67.7) | 0% | 33% | --- | --- | 13.3 ±9.5 (1 – 29) | FCS: 7.0±2.8 (2 – 12) | --- |
| Glenn 2016^51^ | Type 1: 7 (100%) | 55 (35-67) | 0% | 28.6% | --- | --- | --- | --- | --- |
| Goselink 2017^86^ | FSHD NS: 25 (100%) | 40 (10 – 67) | --- | --- | --- | --- | --- | FCS: 5 (2-15) | --- |
| Haley 2020^77^ | FSHD NS: 130 (100%) | --- | --- | --- | --- | --- | --- | FCS: 7.12 ±3.57 | --- |
| Hatch 2022^87^ | FSHD NS: 167 (100%) | --- | --- | --- | --- | --- | --- | --- | --- |
| Hristov 2021^78^ | FSHDH NS: 10 (100%) | 31 – 52 | --- | 70% | --- | --- | --- | 1.5 – 4.0 | --- |
| Liu 2019^52^ | Type 1: 30 (100%) | --- | --- | --- | --- | --- | --- | --- | --- |
| Olivier 2014^88^ | FSHD NS: 8 (100%) | --- | --- | --- | --- | --- | --- | --- | --- |
| Paoletti 2019^89^ | FSHD NS: 14 (100%) | --- | --- | --- | --- | --- | --- | --- | --- |
| Pirola 2018^90^ | FSHD NS: 44 (100%) | --- | --- | 43.2% | --- | --- | --- | --- | --- |
| Salort-Campana 2017^53^ | Type 1: 35 (100%) | --- | --- | 48.6% | --- | --- | --- | --- | --- |
| Statland 2022^91^ | FSHD NS: 240 (100%) | 53 (19-75)^c^ | 0% | 45% | --- | 20^c^ | --- | FCS: 6 (1 – 9)^c^ | 6 (2-10)^c^ |
| Statland 2019^92^ | FSHD NS: 75 (100%) | --- | --- | --- | --- | --- | --- | --- | --- |
| Statland 2016^93^ | FSHD NS: 8 (100%) | 56.5 | --- | 62.5% | --- | --- | --- | n=4 mild-moderate; n=4 severe | --- |
| Statland 2014^63^ | Type 1: 19 (86.4%); Type 2: 3 (13.6%) | --- | --- | --- | --- | --- | --- | --- | --- |
| Vincenten 2021^94^ | FSHD NS: 105 (100%) | 54 ± 14 | --- | 59% | --- | --- | --- | --- | Repeat size: 6 ± 2 |
| Wang 2022^95^ | FSHD NS: 160 (100%) | --- | --- | --- | --- | --- | --- | --- | --- |
| Wang 2019^96^ | FSHD NS: 36 (100%) | --- | --- | --- | --- | --- | --- | --- | --- |
| Wildholm 2022^54^ | Type 1: 17 (100%) | NR (18 – 65) | 0% | --- | --- | --- | --- | --- | --- |
| Woodcock 2020^97^ | FSHD NS: 11 (100%) | --- | --- | --- | --- | --- | --- | --- | --- |
| Zhuparris 2021^98^ | FSHD NS: 38 (100%) | --- | --- | --- | --- | --- | --- | --- | --- |
| FULL TEXTS | | | | | | | | | |
| Andersen 2017^62^ | Type 1: 45 (100%) | 48 ± 15 (20 – 75) | 0% | 44.4% | 25.2 ±3.8 | --- | --- | --- | Residual allele size: 24.9 ±6.8 (10- 36) |
| Bachasson 2014^99^ | FSHD NS: 19 (100%0 | 41 ±13 | --- | 26.3% | 23.8 ±4.0 | --- | --- | --- | --- |
| de Valle 2021^64^ | Type 1: 16 (88.9%); Type 2: 1 (5.6%); missing: 1 (5.6%) | 13.3 ± 3 (7 – 18) | 100% | 44.4% | 20.3 ± 3.9 (15.1 -30.2) | --- | --- | Score 0 – 2: 2.0%±1.2 | Fragment size: 16.9 (10-30) |
| Dos Santos 2021^100^ | FSHD NS: 13 (100%) | 49.5±13.2 | 0% | 69.2% | --- | 42.7 ±15.9 | 6.7±5.9 | GMWS^a^ | --- |
| Eichinger 2018^65^ | Type 1: 35 (85.4%); FSHD NS: 6 (14.6%) | 52.8±11.9 (22 – 70) | 0% | 36.6% | --- | 37.9±11.9  (12 – 60) | 15.7±11  (1 – 42) | FCS: 7.1±2.9 (2 – 13) | Residual fragment size: 25.3±6.6 |
| Eichinger 2017^61^ | Type 1: 86 (100%) | 49.1 ± 15.2 (18-84) | 0% | 42% | --- | --- | --- | FCS: 6.4 ±3  (1 – 12) | Residual fragment size: 23.7 ± 6 (13 -41) |
| Eren 2020^73^ | FSHD NS: 29 (100%) | 34.5 (13 – 73) | NS | --- | --- | --- | --- | --- | --- |
| Esnault 2018^60^ | Type 1: 48 (100%) | 49.7 ± 15.9 (19 – 77) | 0% | 33.3% | --- | --- | --- | 3.5 (1)^b^ | Repeats: 6 ±1.7 (2.3 – 9.4) |
| Fatehi 2017^59^ | Type 1: 35 (100%) | 45 (35 – 56) | 0% | 48.6% | 23.9 (21.6-26.1) | 27.5 (16 – 41) | --- | 3.0 (2.5-3.0)^c^ | --- |
| Goselink 2020^58^ | Type 1: 22 (100%) | 38 (8 – 67) | NS | 45.5% | --- | --- | --- | 4.8 | Repeat arrays: 5.2 ±1.8 (2 – 8) |
| Guillot 2018^72^ | FSHD NS: 193 (100%) | ---^a^ | NS | 45.6% | --- | --- | --- | --- | --- |
| Han 2015^101^ | FSHD NS: 26 (100%) | ---^a^ | --- | 46.2% | --- | --- | --- | --- | --- |
| Hatch 2021^79^ | FSHD NS: 21 (100%) | 41.6 (19 -55) | 0% | 42.9% | 26.5 (17.9- 33.3) | --- | --- | 2.7 (1 – 4) | --- |
| Huisinga 2018^102^ | FSHD NS: 17 (100%) | 53.7 (32 – 67) | 0% | 59% | --- | --- | --- | FCS: 6.7 (2 – 10) | Fragment kb: 24.3 (15-35) |
| Lamperti 2010^71^ | FSHD NS: 69 (100%) | 16-89 | NS | 56.5% | --- | --- | --- | --- | --- |
| Lareau-Trudel 2015^57^ | Type 1: 35 (100%) | 45.9 ±14.3 | --- | 54.2% | 24.6 ±4.2 | 29.7±14.4 | 16.0±10.6 | 2.8 ±1.0 | Repeat units: 6.4±1.4 |
| Mul 2022^76^ | FSHD NS: 119 (100%) | 56.0 ±14.4 (23 – 82) | 0% | 50.4% | --- | --- | --- | FCS: 7.8±4.4 | --- |
| Mul 2021^a69^ | FSHD NS: 714 (100%) | 54.4 (18 – 87) | 0% | 49% | --- | --- | --- | --- | --- |
| Mul 2021^b75^ | FSHD NS: 194 (100%) | 51.6 ± 15.7 | 0% | 51.0% | --- | --- | 26.8 ±17.5 | FCS: 7.2 ±4.4 (0 – 15) | --- |
| Mul 2017^66^ | Type 1: 130 (92.9%); Type 2: 8 (5.7%); Type 1 & 2: 2 (1.4%) | 50.2 ±14.9 (18-81) | 0% | 43.6% | 25.3±4.1 (17.0 – 37.6) | --- | 25.0 ±15.0 (1 – 59) | 0: n=5; 1-3: n=37; 4-6: n=43; 7-10: n=54 | 1-3: n=5; 4 – 7: n=88; 8 – 10: n=30 |
| Personius 1994^70^ | FSHD NS: 32 (100%) | 36.1±9.6 (17 - 49 | NS | --- | --- | --- | --- | --- | --- |
| Regula 2016^56^ | Type 1: 20 (100%) | 46 (7 – 74) | NS | 35.0% | --- | 29.0 ±15.6 | 17.3 ±11.9 | --- | Allele size:27.3 ±6.2 |
| Rijken 2015^103^ | FSHD NS: 9 (100%) | 52 ± 8 | 0% | 22.2% | --- | --- | --- | 3: n=4; 3.5: n=1; 4: n=3; 4.5: n=1 | --- |
| Sanson 2022^104^ | FSHD NS: 281 (100%) | 54.8 ±16.0 | 0% | 46.6% | --- | --- | --- | --- | 2 – 10 |
| Statland 2016^67^ | Type 1: 33 (94.3%); Type 1 2 (5.7)% | 53.0 (22-68) | 0% | 22.9% | --- | 26.0 (10 – 55) | 14.1 (1 – 41) | FCS: 7.3 (2 – 12) | Residual fragment: 25.2 (13 – 41) |
| Tawil 1997^105^ | FSHD NS: 81 (100%) | 39.0±13.4 | --- | 56.8% | --- | ---^a^ | ---^a^ | --- | --- |
| Teeselink 2022^68^ | Type 1: 88 (95.7%); Type 2: 4 (4.3%) | 48.4±15.3 (18 – 75) | 0% | 43% | --- | --- | 21.3 ± 16.8 (0 – 59) | ---^a^ | Type 1 6 (2 – 9)^c^ |
| Widholm 2022^55^ | Type 1: 17 (100%) | 49.4 (23 – 65) | 0% | 29.4% | --- | --- | --- | 3.0±0.71 | --- |

Data are presented as mean ± SD or ranges, unless otherwise specified.

^a^Reported by subgroup or by category only.

^b^ Presented as median (inter-quartile range).

^c^ Presented as median with or without (range)

BMI, body mass index; CSS, clinical severity score; FCS, FSHD clinical score; FSHD, facioscapulohumeral dystrophy; GMWS, Gardner-Medwin-Walton scale; NR, not reported; NS, not specified.

Table 17. Correlation outcomes for the topic “Disease Outcome Measures and Validation”

| **Author, year** | **Assessment Measures** | **Correlated with** | **n** | **Timepoint** | **Measure of correlation** | **Value** | **p-value** | **95% CI** | **Notes** |
| --- | --- | --- | --- | --- | --- | --- | --- | --- | --- |
| CONFERENCE ABSTRACTS | | | | | | | | | |
| Berggren 2022^80^ | Left cheek strength (IOPI) | D4Z4 repeats | 227 | NR | Spearman | 0.38 | <.01 | --- |  |
|  | Right cheek strength (IOPI) | D4Z4 repeats | 227 | NR | Spearman | 0.49 | <.01 | --- |  |
|  | Lip strength | D4Z4 repeats | 227 | NR | Spearman | 0.32 | <.01 | --- |  |
|  | Left cheek strength (IOPI) | MMT Total score | 227 | NR | Spearman | 0.47 | <.01 | --- |  |
|  | Right cheek strength (IOPI) | MMT Total score | 227 | NR | Spearman | 0.50 | <.01 | --- |  |
|  | Lip strength | MMT Total score | 227 | NR | Spearman | 0.37 | <.01 | --- |  |
|  | Left cheek strength (IOPI) | FSHD-HI impaired facial expression rating | 227 | NR | Spearman | -0.34 | <.01 | --- |  |
|  | Right cheek strength (IOPI) | FSHD-HI impaired facial expression rating | 227 | NR | Spearman | -0.32 | <.01 | --- |  |
|  | Lip strength | FSHD-HI impaired facial expression rating | 227 | NR | Spearman | -0.37 | <.01 | --- |  |
|  | Left cheek strength (IOPI) | FCS (Overall) | 227 | NR | Spearman | -0.35 | <.01 | --- |  |
|  | Right cheek strength (IOPI) | FCS (Overall) | 227 | NR | Spearman | -0.45 | <.01 | --- |  |
|  | Lip strength | FCS (Overall) | 227 | NR | Spearman | -0.33 | <.01 | --- |  |
| Chan 2021^81^ | TUG | FSHD-TUG | 22 | All visits | Pearson | ≤0.95 | ≤.01 | --- | Strong |
|  | CSS | TUG | 22 | 2 initial visits, separated by 14 days;  3 additional visits, every 3 months, up to 12 months | Pearson | 0.57-0.59 | ≤.01 | --- | Moderate |
|  | CSS | FSHD-TUG | 22 | 3 initial visits, separated by 14 days;  3 additional visits, every 3 months, up to 12 months | Pearson | 0.58-0.67 | ≤.01 | --- | Moderate |
|  | PROMIS-PF | TUG | 22 | 4 initial visits, separated by 14 days;  3 additional visits, every 3 months, up to 12 months | Pearson | -0.56 to -0.69 | ≤.01 | --- | Moderate |
|  | PROMIS-PF | FSHD-TUG | 22 | 5 initial visits, separated by 14 days;  3 additional visits, every 3 months, up to 12 months | Pearson | -0.59 to -0.72 | ≤.01 | --- | Moderate |
| Eichinger 2021^84^ | Overall lower extremity strength (MMT) | "functional assessments" | 133 | --- | Spearman | 0.66-0.84 | --- | --- |  |
|  | Overall lower extremity strength (QMT) | "functional assessments" | 133 | --- | Spearman | 0.49-0.64 | --- | --- |  |
|  | Individual lower extremity muscle groups | 10MWT | 133 | --- | Spearman | -0.41 to -0.74 | --- | --- |  |
|  | Overall upper extremity strength (MMT) | Don/doff coat | 133 | --- | Spearman | -0.63 | 0.01 | --- |  |
| Eichinger 2022^83^ | Lower extremity strength (MRC Scale) | MMT | Baseline: 240 Follow-up: 157 | Baseline; 12-months | Spearman | −0.68 to −0.83 | --- | --- |  |
|  | Lower extremity strength (MRC Scale) | 10MWT | Baseline: 240 Follow-up: 158 | Baseline; 12-months | Spearman | −0.43 to −0.72 | --- | --- | Hip and knee extensors had the strongest correlation |
|  | Upper extremity strength (MRC scale) | Don/doff coat | Baseline: 240 Follow-up: 159 | Baseline; 12-months | Spearman | −0.60 | <0.0001 | --- |  |
| Glenn 2016^51^ | EIM leg score | CSS | 7 | --- | Pearson | -0.58 | --- | --- | Moderate |
|  | EIM 50Hz reactance | Fatty replacement on MRI T1 sequences | 7 | --- | Pearson | -0.64 to -0.89 | --- | --- | EIM 50 kHz reactance decreased with increasing fatty replacement on MRI T1 sequences |
| Goselink 2017^86^ | Muscle ultrasonography of 5 muscles (trapezius, biceps brachii, rectus abdominis, rectus femoris, tibialis anterior) | FCS | 25 | Baseline | --- | "reasonable to good correlation" | --- | --- |  |
|  | Muscle ultrasonography of 5 muscles (trapezius, biceps brachii, rectus abdominis, rectus femoris, tibialis anterior) | CSS | 25 | Baseline | --- | "reasonable to good correlation" | --- | --- |  |
|  | Muscle ultrasonography of 5 muscles (trapezius, biceps brachii, rectus abdominis, rectus femoris, tibialis anterior) | MMT | 25 | Baseline | --- | "reasonable to good correlation" | --- | --- |  |
| Haley 2020^77^ | FCS | FVC | 42 | --- | --- | "correlated in exploratory evaluations" | | |  |
|  | FCS | IOPI | 42 | --- | --- | "correlated in exploratory evaluations" | | |  |
| Hristov 2021^78^ | eTUG | CSS | 10 | Over 3 months | Spearman | 0.88 | 0.0001 | --- | Very strong |
|  | eTUG | TUG | 10 | Over 3 months | Spearman | 0.73 | 0.017 | --- | Good |
|  | eTUG | FSHD TUG | 10 | Over 3 months | Spearman | 0.73 | 0.73 | --- | Good |
|  | eTUG | NeuroLE | 10 | Over 3 months | Spearman | -0.73 | 0.017 | --- | Good |
|  | eTUG | NeuroUE | 10 | Over 3 months | Spearman | 0.69 | 0.027 | --- | Good |
|  | eTUG | PROMIS | 10 | Over 3 months | Spearman | -0.66 | 0.038 | --- | Good |
|  | In-home Gait Speed | TUG | 10 | Over 3 months | Spearman | -0.73 | 0.018 | --- | Good |
|  | In-home Gait Speed | FSHD TUG | 10 | Over 3 months | Spearman | -0.68 | 0.032 | --- | Good |
|  | Sleep Schedule Variability | CSS | 10 | Over 3 months | Spearman | 0.8 | 0.005 | --- | Very strong |
|  | Sleep Schedule Variability | NeuroUE | 10 | Over 3 months | Spearman | 0.72 | 0.018 | --- | Good |
|  | Sleep Schedule Variability | PROMIS | 10 | Over 3 months | Spearman | -0.68 | 0.03 | --- | Good |
|  | Sleep Schedule Variability | RWS | 10 | Over 3 months | Spearman | -0.68 | 0.032 | --- | Good |
| Liu 2019^52^ | Fat infiltration score of total skeletal muscles PATCHS MRI | MFM | 30 | --- | "correlation coefficient" | -0.8767 | --- | --- | Correlated highly |
|  | Fat infiltration score of total skeletal muscles PATCHS MRI | FCS | 30 | --- | "correlation coefficient" | 0.7768 | --- | --- |  |
|  | Fat infiltration score of total skeletal muscles PATCHS MRI | CSS | 30 | --- | "correlation coefficient" | 0.7705 | --- | --- |  |
|  | Fat infiltration score of total skeletal muscles PATCHS MRI | 6MWT | 30 | --- | "correlation coefficient" | -0.7077 | --- | --- |  |
|  | Fat infiltration score of total skeletal muscles PATCHS MRI | MMT | 30 | --- | "correlation coefficient" | 0.6973 | --- | --- |  |
| Olivier 2014^88^ | Peak torque | Initial muscle deoxygenation | 8 FSHD; 15 age-matched controls | --- | --- | "correlated" | --- | --- |  |
|  | Peak torque | Functional impairment (walking endurance) | 9 FSHD; 15 age-matched controls | --- | --- | "correlated" | --- | --- |  |
| Paoletti 2019^89^ | MFF (tibialis anterior, hamstring, adductor magnus) | CSS | Baseline: 14;  Follow-up: 11 | Baseline; 6-month follow-up | --- | "positive correlation" | --- | --- |  |
|  | MFF (tibialis anterior, hamstring, adductor magnus) | 6MWT | Baseline: 14;  Follow-up: 11 | Baseline; 6-month follow-up | --- | "negative correlation" | --- | --- |  |
|  | MFF (tibialis anterior, hamstring, adductor magnus) | Dynamometric measures | Baseline: 14;  Follow-up: 11 | Baseline; 6-month follow-up | --- | "negative correlation" | --- | --- |  |
|  | waterT2 times ((tibialis anterior, hamstring, adductor magnus) | CSS | Baseline: 14;  Follow-up: 12 | Baseline; 6-month follow-up | --- | "positive correlation" | --- | --- |  |
|  | waterT2 times ((tibialis anterior, hamstring, adductor magnus) | 6MWT | Baseline: 14;  Follow-up: 13 | Baseline; 6-month follow-up | --- | "negative correlation" | --- | --- | --- |
|  | waterT2 times ((tibialis anterior, hamstring, adductor magnus) | Dynamometric measures | Baseline: 14;  Follow-up: 13 | Baseline; 6-month follow-up | --- | "negative correlation" | --- | --- |  |
| Pirola 2018^90^ | FVC | 10MWT | 44 | --- | Pearson | -0.42 | <0.05 | --- |  |
|  | FVC | 4 step walking test | 44 | --- | Pearson | -0.53 | <0.05 | --- |  |
|  | FVC | 6MWT | 44 | --- | Pearson | 0.60 | <0.05 | --- |  |
|  | FVCsupine | 6MWT | 44 | --- | Pearson | 0.50 | <0.05 | --- |  |
|  | SpO2 | 4 step walking test | 44 | --- | Pearson | -0.60 | <0.05 | --- |  |
|  | SpO2 | 6MWT | 44 | --- | Pearson | 0.60 | <0.05 | --- |  |
|  | ODI | 4 step walking test | 44 | --- | Pearson | 0.50 | <0.05 | --- | ODI not defined in conference abstract |
|  | ODI | 6MWT | 44 | --- | Pearson | -0.60 | <0.05 | --- |  |
| Salort-Campana 2017^53^ | MPI Total | "clinical scores" (MMT using MRC, MFM) | 35 | --- | --- | "significantly related" | --- | --- |  |
|  | MPI Total | MRC sum score changes | 35 | --- | --- | "no correlation" | --- | --- |  |
|  | MPI Total | MFM total | 35 | --- | --- | "no correlation" | --- | --- |  |
|  | MPI Total | MFM subscores | 35 | --- | --- | "no correlation" | --- | --- |  |
| Statland 2014^63^ | Creatine kinase MB fraction, tissue-type plasminogen activator, epidermal growth factor | FSHD disease severity | 22 FSHD patients;  23 age-matched health controls | --- | --- | "moderate correlation" | --- | --- |  |
|  | Markers in the plasminogen pathway (tPA, plasminogen activator inhibitor, and vitronectin) | Each other (in FSHD patients only) | 23 FSHD patients;  23 age-matched health controls | --- | --- | "correlated with each other" | --- | --- |  |
| Statland 2016^93^ | Spatial and temporal gait metrics using wireless system | 6MWT | 5 | --- | Pearson | 0.76-0.96 | --- | --- | Moderate to strong |
| Statland 2019^92^ | Don/doff coat | MMT: UE (shoulder extension, elbow flexion) | 75 | --- | Pearson | 0.35-0.46 | --- | --- |  |
|  | Don/doff coat | MMT: LE (ankle dorsiflexion, knee flexion) | 75 | --- | Pearson | 0.32-0.41 | --- | --- |  |
|  | Supine to sit | MMT LE | 74 | --- | --- | --- | --- | --- | "Conversely depended more on lower extremity strength" |
|  | 6MWT | MMT: UE & LE muscle groups | 75 | --- | Pearson | >0.4 in 9/13 muscle groups | --- | --- |  |
|  | Gait | MMT: knee flexion | 75 | --- | Pearson | 0.58-0.72 | --- | --- |  |
| Vincenten 2021^94^ | MRI total leg fat fraction | CSS & MFM | 105 | Basline & 5-yr followup | "correlation coefficient" | 0.864 | <0.05 | --- | Strong |
|  | Change in MRI total leg fat fraction | CSS & MFM | 105 | Basline & 5-yr followup | "correlation coefficient" | 0.22-0.387 | --- | --- | Significantly |
| Wang 2019^96^ | MFF Burden Index (sum of 16 LE muscles) | 6MWT | 36 | "over one year" | --- | "correlated" | --- | --- |  |
|  | MFF Burden Index (sum of 16 LE muscles) | Go-30-feet | 36 | "over one year" | --- | "correlated" | --- | --- |  |
|  | MFF of tibilias anterior | Strength assessment of tibialis anterior | 36 | "over one year" | --- | "correlated" | --- | --- |  |
| Wang 2022^95^ | LE lean muscle mass via DEXA scan | 6MWT | 160 | Baseline | Pearson | 0.34 | --- | --- | Moderate |
| Wildholm 2022^54^ | Total LMV at baseline via MRI | LMV at follow-up | 17 | 4-12 weeks later | Pearson | 0.9984 | --- | --- | Strong |
|  | Total MFI at baseline via MRI | MFI at follow-up | 17 | 4-12 weeks later | Pearson | 0.9845 | --- | --- | Strong |
|  | Total MFF at baseline via MRI | MFF at follow-up | 17 | 4-12 weeks later | Pearson | 0.9918 | --- | --- | Strong |
|  | Composite MFF total for LE muscles via MRI | TUG | 17 | Baseline; 4-12 weeks later | Pearson | 0.72 | --- | --- |  |
|  | Composite MFI total for LE muscles via MRI | TUG | 17 | Baseline; 4-12 weeks later | Pearson | 0.83 | --- | --- |  |
|  | Composite MFF total for UE/LE via MRI | FSHD-TUG | 17 | Baseline; 4-12 weeks later | Pearson | 0.73 | --- | --- |  |
|  | Composite MFI total for trunk via MRI | FSHD-TUG | 17 | Baseline; 4-12 weeks later | Pearson | 0.73 | --- | --- |  |
|  | Composite total MFF UE via MRI | RWS: left arm | 17 | Baseline; 4-12 weeks later | Pearson | -0.71 | --- | --- |  |
|  | Composite total MFF trunk via MRI | RWS: right arm | 17 | Baseline; 4-12 weeks later | Pearson | -0.61 | --- | --- |  |
|  | Composite total MFF UE via MRI | RWS: left arm | 17 | Baseline; 4-12 weeks later | Pearson | -0.53 | --- | --- |  |
|  | Composite total MFF trunk via MRI | RWS: right arm | 17 | Baseline; 4-12 weeks later | Pearson | -0.65 | --- | --- |  |
| Woodcock 2020^97^ | Mercuri score | CSS | 11 | --- | Pearson | 0.95 | --- | --- | Excellent |
|  | Mercuri score | FSHD-COM | 11 | --- | Pearson | 0.92 | --- | --- | Excellent |
|  | Mercuri score | FCS | 11 | --- | Pearson | 0.93 | --- | --- | Excellent |
|  | Mercuri score | 6MWT | 11 | --- | Pearson | -0.72 | --- | --- | Good |
|  | Mercuri score | PUL2.0 | 11 | --- | Pearson | -0.85 | --- | --- | Good |
|  | MRI | MFM | 11 | --- | Pearson | -0.83 | --- | --- | Trend suggest may not be sensitive |
| Zhuparris 2021^98^ | Remote sensor data to estimate clinical severity | FCS | 38 | Day 1; day 42 | R^2^ | 0.57 | --- | --- |  |
|  | Remote sensor data to estimate clinical severity | TUG | 38 | Day 1; day 42 | R^2^ | 0.59 | --- | --- |  |
|  | Remote sensor data to estimate clinical severity | FCS and TUG | 38 | Day 1; day 42 | R^2^ | 0.74 | --- | --- | Outperformed single task model |
| FULL TEXTS | | | | | | | | | |
| Andersen 2017^62^ | MFF | FCS | 45 | Baseline | Pearson | 0.76 | <0.0001 | --- |  |
|  | MFF | Neck Flexion | 38 | Baseline | Pearson | -0.67 | <0.0001 | --- |  |
|  | MFF | Back flexion | 36 | Baseline | Pearson | -0.74 | <0.0001 | --- |  |
|  | MFF | Hip Flexion | 82 | Baseline | Pearson | -0.33 | <0.05 | --- |  |
|  | MFF | Knee Flexion | 80 | Baseline | Pearson | -0.63 | <0.0001 | --- |  |
|  | MFF | Knee Extension | 81 | Baseline | Pearson | -0.65 | <0.0001 | --- |  |
|  | MFF | Plantar Flexion | 40 | Baseline | Pearson | -0.50 | <0.01 | --- |  |
|  | MFF | Dorsal Flexion | 39 | Baseline | Pearson | -0.54 | <0.001 | --- |  |
|  | MFF | 5TSTST | 40 | Baseline | Pearson | 0.62 | <0.0001 | --- |  |
|  | MFF | 14SST | 41 | Baseline | Pearson | 0.65 | <0.0001 | --- |  |
|  | MFF | 6MWT | 42 | Baseline | Pearson | -0.79 | <0.0001 | --- |  |
| Bachasson 2014^99^ | MVC | Voluntary Activation | 19 | --- | Pearson | -0.41 | <0.05 | --- |  |
|  | MVC per kg of body weight | 6MWT | 19 | --- | Pearson | 0.77 | < 0.001 | --- |  |
|  | MVC per kg of body weight | Peak oxygen consumption per kg of body weight | 19 | --- | Pearson | 0.74 | <0.001 | --- |  |
|  | MVC per kg of body weight | Maximal workload during cycling test | 19 | --- | Pearson | 0.82 | <0.001 | --- |  |
|  | MVC per kg of body weight | Fatigue severity scale score | 19 | --- | Pearson | -0.65 | <0.05 | --- |  |
|  | MVC per kg of body weight | SF-36 physical functioning subscore | 19 | --- | Pearson | 0.58 | <0.01 | --- |  |
| de Valle 2021^64^ | FSHD-COM | FCS | 18 | Baseline | Spearman | 0.82 | 0.0001 | --- |  |
|  | FSHD-COM | CSS | 18 | Baseline | Spearman | 0.70 | 0.001 | --- |  |
|  | FSHD-COM | Age-corrected CSS | 18 | Baseline | Spearman | 0.73 | 0.0005 | --- |  |
|  | FSHD-COM | MFM-32 | 18 | Baseline | Spearman | -0.80 | 0.0001 | --- | Correlations with MFM component scores available |
|  | FSHD-COM | PUL 2.0 | 13 | Baseline | Spearman | -0.94 | 0.0001 | --- | Correlations with PUL component scores available |
|  | FSHD-COM | PedsQL NMM participant | 17 | Baseline | Spearman | -0.70 | 0.002 |  | Correlations with PedsQL participant component scores available |
|  | FSHD-COM | PedsQL NMM proxy | 18 | Baseline | Spearman | -0.63 | 0.006 | --- | Correlations with PedsQL proxy component scores available |
|  | FSHD-COM | FSHD-HI | 18 | Baseline | Spearman | 0.60 | 0.008 | --- | Correlations with FSHD-HI component scores available |
|  | FSHD-COM Peds | FCS | 18 | Baseline | Spearman | 0.88 | 0.0001 | --- |  |
|  | FSHD-COM Peds | CSS | 18 | Baseline | Spearman | 0.80 | 0.0001 | --- |  |
|  | FSHD-COM Peds | Age-corrected CSS | 18 | Baseline | Spearman | 0.81 | 0.0001 | --- |  |
|  | FSHD-COM Peds | MFM-32 | 18 | Baseline | Spearman | -0.75 | 0.0003 | --- | Correlations with MFM component score available |
|  | FSHD-COM Peds | PUL 2.0 | 13 | Baseline | Spearman | -0.96 | 0.0001 | --- | Correlations with PUL component scores available |
|  | FSHD-COM Peds | PedsQL NMM participant | 17 | Baseline | Spearman | -0.66 | 0.004 | --- | Correlations with PedsQL participant component scores available |
|  | FSHD-COM Peds | PedsQL NMM proxy | 18 | Baseline | Spearman | -0.67 | 0.003 | --- | Correlations with PedsQL proxy component scores available |
|  | FSHD-COM Peds | FSHD-HI | 18 | Baseline | Spearman | 0.69 | 0.002 | --- | Correlations with FSHD-HI component scores available |
| Dos Santos 2021^100^ | MMSE | Initial symptoms | 13 | Baseline | Spearman | --- | 0.056 | --- |  |
|  | MMSE | Length of illness | 13 | Baseline | Spearman | --- | 0.292 | --- |  |
|  | MMSE | Gardner-Medwin-Walton Scale | 13 | Baseline | Spearman | --- | 0.876 | --- |  |
|  | MMSE | Education | 13 | Baseline | Spearman | 0.677 | 0.011 | --- |  |
|  | MMSE | SF36: domain of emotional limitation | 13 | Baseline | Spearman | 0.657 | 0.015 | --- |  |
|  | MoCA | Initial symptoms | 13 | Baseline | Spearman | --- | 0.173 | --- |  |
|  | MoCA | Length of illness | 13 | Baseline | Spearman | --- | 0.422 | --- |  |
|  | MoCA | Gardner-Medwin-Walton Scale | 13 | Baseline | Spearman | --- | 0.578 | --- |  |
|  | MoCA | Education | 13 | Baseline | Spearman | 0.556 | 0.048 | --- |  |
|  | Trail-Making Test A | Initial symptoms | 13 | Baseline | Spearman | --- | 0.119 | --- |  |
|  | Trail-Making Test A | Length of illness | 13 | Baseline | Spearman | --- | 0.402 | --- |  |
|  | Trail-Making Test A | Gardner-Medwin-Walton Scale | 13 | Baseline | Spearman | --- | 0.807 | --- |  |
|  | Trail-Making Test A | Education | 13 | Baseline | Spearman | -0.704 | 0.0007 | --- |  |
|  | Trail-Making Test A | SF36: domain of emotional limitation | 13 | Baseline | Spearman | -0.601 | 0.03 | --- |  |
|  | Trail-Making Test A | Age | 13 | Baseline | Spearman | 0.578 | 0.039 | --- |  |
|  | Long-term memory (A7) | Initial symptoms | 13 | Baseline | Spearman | -0.593 | 0.033 | --- |  |
|  | Long-term memory (A7) | Length of illness | 13 | Baseline | Spearman | 0.131 | 0.339 | --- |  |
|  | Long-term memory (A7) | Gardner-Medwin-Walton Scale | 13 | Baseline | Spearman | --- | 0.131 | --- |  |
|  | Long-term memory (A7) | Education | 13 | Baseline | Spearman | 0.644 | 0.013 | --- |  |
|  | Long-term memory (A7) | Age | 13 | Baseline | Spearman | -0.628 | 0.022 | --- |  |
|  | Long-term memory (A7) | SF36: domain of emotional limitation | 13 | Baseline | Spearman | 0.617 | 0.025 | --- |  |
| Eichinger 2017^61^ | 6MWT | MMT Lext | 86 | --- | Spearman | 0.79 | <0.0001 | --- | Strong |
|  | 6MWT | MMT Lext | 86 | --- | R^2^ | 0.47 | <0.0001 | --- |  |
|  | 6MWT | TUG | 48 | --- | Spearman | -0.81 | <0.0001 | --- |  |
|  | 6MWT | 30 foot go | 50 | --- | Spearman | -0.83 | <0.0001 | --- |  |
|  | 6MWT | 10MWR | --- | --- | Spearman | -0.78 | <0.0001 | --- |  |
|  | 6MWT | FCS | 38 | --- | Spearman | -0.57 | <0.0001 | --- |  |
| Eichinger 2018^65^ | FSHD-COM total score | FCS | 40 | --- | Pearson | 0.89 | <0.0001 | --- |  |
|  | FSHD-COM total score | Years since diagnosis | 41 | --- | Pearson | 0.63 | 0.001 | --- |  |
|  | FSHD-COM total score | MMT (34 muscles) | 41 | --- | Pearson | -0.91 | <0.0001 | --- |  |
|  | FSHD-COM total score | MMT UE | 31 | --- | Pearson | -0.76 | <0.0001 | --- |  |
|  | FSHD-COM total score | MMT LE | 31 | --- | Pearson | -0.79 | <0.0001 | --- |  |
|  | FSHD-COM total score | Quantitative Myometry standard score | 31 | --- | Pearson | -0.77 | <0.0001 | --- |  |
|  | FSHD-COM total score | Upper Extremities Quantitative Myometry standard score | 31 | --- | Pearson | -0.71 | <0.0001 | --- |  |
|  | FSHD-COM total score | Lower Extremities Quantitative Myometry standard score | 31 | --- | Pearson | -0.71 | <0.0001 | --- |  |
|  | FSHD-COM total score | SF36 Physical | 41 | --- | Pearson | -0.63 | <0.0001 | --- |  |
|  | FSHD-COM total score | SF36 Mental | 41 | --- | Pearson | 0.02 | 0.89 | --- |  |
|  | FSHD-COM total score | Facial Disability Index | 41 | --- | Pearson | -0.27 | 0.09 | --- | Component Scores available |
|  | FSHD-COM total score | PROMIS57 Physical | 31 | --- | Pearson | 0.75 | <0.001 | --- |  |
|  | FSHD-COM total score | PROMIS57 Anxiety | 31 | --- | Pearson | 0.28 | 0.12 | --- |  |
|  | FSHD-COM total score | PROMIS57 Depression | 31 | --- | Pearson | 0.30 | 0.10 | --- |  |
|  | FSHD-COM total score | PROMIS57 Fatigue | 31 | --- | Pearson | 0.100 | 0.58 | --- |  |
|  | FSHD-COM total score | PROMIS57 Sleep | 31 | --- | Pearson | 0.26 | 0.15 | --- |  |
|  | FSHD-COM total score | PROMIS57 Pain | 31 | --- | Pearson | 0.14 | 0.60 | --- |  |
|  | FSHD-COM total score | PROMIS57 Social Satisfaction | 31 | --- | Pearson | -0.48 | 0.006 | --- |  |
| Esnault 2018^60^ | IPT at 60°/s | Number of D4Z4 repeats | 48 | --- | Spearman | 0.440 | 0.002 | --- | Significant |
|  | Trunk flexors IPT at 60°/s | Number of D4Z4 repeats | 48 | --- | Spearman | 0.347 | 0.018 | --- | Significant |
|  | CSS | Number of D4Z4 repeats | 48 | --- | Spearman | -0.386 | 0.007 |  | Significant |
|  | Berg Balance Scale | Number of D4Z4 repeats | 48 | --- | Spearman | 0.194 | 0.190 | --- | Weren't significantly correlated |
|  | Functional Reach Test | Number of D4Z4 repeats | 48 | --- | Spearman | 0.220 | 0.137 | --- | Weren't significantly correlated |
|  | TUG | Number of D4Z4 repeats | 48 | --- | Spearman | 0.115 | 0.460 | --- | Weren't significantly correlated |
|  | 6MWT | Number of D4Z4 repeats | 48 | --- | Spearman | 0.187 | 0.218 | --- | Weren't significantly correlated |
|  | Functional Independence Measure | Number of D4Z4 repeats | 48 | --- | Spearman | 0.081 | 0.584 | --- | Weren't significantly correlated |
|  | Trunk extensors IPT at 60°/s | Number of D4Z4 repeats | 48 | --- | Spearman | 0.391 | 0.006 | 0.121-0.662 |  |
|  | Trunk flexors IPT at 60°/s | Number of D4Z4 repeats | 48 | --- | Spearman | 0.334 | 0.033 | 0.28-0.641 |  |
|  | Trunk extensors IPT | CSS | 48 | --- | Spearman | -0.743 | 0.0001 | -0.938 to -0.548 |  |
|  | Trunk flexors IPT at 60°/s | CSS | 48 | --- | Spearman | -0.404 | 0.01 | -0.708 to -0.100 |  |
|  | CSS | Number of D4Z4 repeats | 48 | --- | Spearman | -0.439 | 0.02 | -0.709 to -0.170 |  |
| Fatehi 2017^59^ | MPI Total | Right Thigh MMT (MRC Score) | 35 | Timepoint 1 | Spearman | -0.63 | 0.000 | --- |  |
|  | MPI Total | Left Thigh MMT (MRC Score) | 35 | Timepoint 1 | Spearman | -0.68 | 0.000 | --- |  |
|  | MPI Total | MMT MRC Sumscore | 35 | Timepoint 1 | Spearman | -0.66 | 0.000 | --- |  |
|  | MPI Total | CSS | 35 | Timepoint 1 | Spearman | 0.65 | 0.000 | --- |  |
|  | MPI Total | MFM D1% | 35 | Timepoint 1 | Spearman | -0.64 | 0.000 | --- |  |
|  | MPI Total | MFM D2% | 35 | Timepoint 1 | Spearman | -0.37 | 0.037 | --- |  |
|  | MPI Total | MFM D3% | 35 | Timepoint 1 | Spearman | -0.22 | 0.228 | --- |  |
|  | MPI Total | MFM Total | 35 | Timepoint 1 | Spearman | -0.62 | 0.000 | --- |  |
|  | Duration of disease | MPI Total | 35 | Timepoint 1 | Spearman | 0.37 | 0.030 | --- |  |
|  | Duration of disease | MMT MRC Sumscore | 35 | Timepoint 1 | Spearman | -0.14 | 0.420 | --- |  |
|  | Duration of disease | CSS | 35 | Timepoint 1 | Spearman | 0.19 | 0.260 | --- |  |
|  | Duration of disease | MFM D1% | 35 | Timepoint 1 | Spearman | -0.07 | 0.700 | --- |  |
|  | Duration of disease | MFM D2% | 35 | Timepoint 1 | Spearman | -0.32 | 0.700 | --- |  |
|  | Duration of disease | MFM D3% | 35 | Timepoint 1 | Spearman | 0.04 | 0.820 | --- |  |
|  | Duration of disease | MFM Total | 35 | Timepoint 1 | Spearman | -0.06 | 0.760 | --- |  |
|  | MPI Total | Repeat Units | 35 | Timepoint 1 | Spearman | -0.22 | 0.220 | --- |  |
|  | MPI Total | BMI | 35 | Timepoint 1 | Spearman | 0.15 | 0.410 | --- |  |
|  | MPI Total | BMI | 35 | Timepoint 2 | Spearman | 0.22 | 0.220 | --- |  |
|  | MPI Total | BMI | 35 | Timepoint 3 | Spearman | 0.06 | 0.850 | --- |  |
|  | Rate of Change MPI Total | Duration of disease | 35 | --- | Spearman | -0.21 | 0.230 | --- |  |
|  | Rate of Change MPI Total | Age of patients at T1 | 35 | --- | Spearman | 0.12 | 0.480 | --- |  |
|  | Rate of Change MPI Total | MMT MRC Sumscore | 35 | --- | Spearman | 0.04 | 0.81 | --- |  |
|  | Rate of Change MPI Total | MFM D1% | 35 | --- | Spearman | -0.17 | 0.34 | --- |  |
|  | Rate of Change MPI Total | MFM D2% | 35 | --- | Spearman | 0.22 | 0.23 | --- |  |
|  | Rate of Change MPI Total | MFM D3% | 35 | --- | Spearman | -0.07 | 0.70 | --- |  |
|  | Rate of Change MPI Total | MFM Total | 35 | --- | Spearman | -0.06 | 0.76 | --- |  |
| Goselink 2020^58^ | MUS (Qualitative: Heckmatt Score) | MUS (Quantitative: echo-intensity) | 22 | --- | Spearman | 0.79 | <0.001 | --- | Strong |
|  | MUS (Qualitative: Heckmatt Score) | FCS | 22 | --- | Spearman | 0.81 | <0.001 | --- | Positive |
|  | MUS (Quantitative: echo-intensity) | FCS | 22 | Baseline | Spearman | 0.61 | 0.002 | --- | Positive |
| Guillot 2018^72^ | MFM: D3 | MFM: D2 | 193 | "most recent" | --- | 0.82 | --- | --- |  |
|  | MFM: D3 | MFM: D1 | 193 | "most recent" | --- | 0.74 | --- | --- |  |
| Han 2015^101^ | Total RSA | Elbox flexion + shoulder abduction % - predicted MVIC | 26 FHSD | Baseline | Pearson | 0.477 | 0.0003 | --- |  |
|  | Total RSA | Shoulder abduction % - predicted MVIC | 26 FHSD | Baseline | Pearson | 0.094 | 0.508 | --- |  |
|  | Total RSA | Elbow flexion % - predicted MVIC | 26 FHSD | Baseline | Pearson | 0.528 | <0.0001 | --- |  |
| Hatch 2021^79^ | Total RSA | NeuroQoL raw | 21 | --- | Spearman | 0.7609 | 0.0001 | --- |  |
|  | Total RSA | Sub-NeuroQol | 21 | --- | Spearman | 0.8397 | <0.000 | --- |  |
|  | Total RSA | NeuroQoL raw | 21 | --- | Spearman | 0.6969 | 0.0004 | --- |  |
|  | RSA above shoulder | Sub-NeuroQol | 21 | --- | Spearman | 0.8082 | <0.000 | --- |  |
| Huisinga 2018^102^ | iTUG: Stride velocity | FCS | 17 | Baseline | Pearson | -0.65 | 0.005 | --- | Not correlated |
|  | iTUG: Stride velocity | MMT LEXT | 17 | Baseline | Pearson | 0.52 | 0.03 | --- | Not correlated |
|  | iTUG: Stride velocity | PROMIS PF2A | 17 | Baseline | Pearson | 0.44 | 0.07 | --- | Not correlated |
|  | iTUG: Sagittal Trunk ROM | FCS | 17 | Baseline | Pearson | 0.70 | 0.002 | --- | Not correlated |
|  | iTUG: Sagittal Trunk ROM | MMT LEXT | 17 | Baseline | Pearson | -0.68 | 0.003 | --- | Not correlated |
|  | iTUG: Sagittal Trunk ROM | PROMIS PF2A | 17 | Baseline | Pearson | -0.53 | 0.03 | --- | Not correlated |
|  | iTUG: Z-score | FCS | 17 | Baseline | Pearson | 0.79 | <0.01 | --- |  |
|  | iTUG: Z-score | MMT LEXT | 17 | Baseline | Pearson | -0.72 | <0.01 | --- |  |
|  | iTUG: Z-score | PROMIS PF2A | 17 | Baseline | Pearson | -0.58 | 0.01 | --- |  |
| Lareau-Trudel 2015^57^ | Mean visual score of fatty infiltration in thighs | MFF with qMRI | 35 | --- | R^2^ | 0.64 | <0.001 | --- | Non-linear relationship assumed |
|  | Mean visual score of fatty infiltration in thighs | MFF with qMRI | 35 | --- | R^2^ | 0.8 | --- | --- | Linear relationship assumed |
|  | Intramuscular fat fraction with qMRI | CSS | 35 | --- | --- | "significanlty correlated" | <0.01 | --- |  |
|  | Intramuscular fat fraction with qMRI | MMT score | 35 | --- | --- | "inversely correlated" | <0.01 | --- |  |
|  | Intramuscular fat fraction with qMRI | MFM D1 | 35 | --- | --- | "inversely correlated" | <0.01 | --- |  |
|  | Intramuscular fat fraction with qMRI | disease duration | 35 | --- | --- | "no correlation" | 0.10 | --- |  |
|  | Intramuscular fat fraction with qMRI | size of contracted allele | 35 | --- | --- | "no correlation" | 0.40 | --- |  |
| Mul 2017^66^ | MFF, legs | MFM | 140 | --- | Spearman | -0.845 | <0.001 | --- |  |
|  | MFF, legs | 6-MWT | 111 | --- | Spearman | -0.701 | <0.001 | --- |  |
|  | MFF, legs | FCS | 140 | --- | Spearman | 0.835 | <0.001 | --- |  |
|  | MFF, legs | CSS | 140 | --- | Spearman | 0.791 | <0.001 | --- |  |
|  | MFF, right leg | MRC score: knee flexion | --- | --- | Spearman | -0.74 | <0.001 | --- |  |
|  | MFF, right leg | MRC score: knee extension | --- | --- | Spearman | -0.77 | <0.001 | --- |  |
|  | MFF, right leg | MRC score: foot dorsal flexion | --- | --- | Spearman | -0.69 | <0.001 | --- |  |
|  | MFF, right leg | MRC score: plantar flexors | --- | --- | Spearman | -0.38 | <0.001 | --- | Weak |
|  | MFF, right leg | Quantitative isometric knee extensor strength | --- | --- | Spearman | -0.42 | <0.001 | --- |  |
|  | MFF, right leg | MMT: knee extension strength of vastus muscles | --- | --- | Spearman | -0.82 | <0.001 | --- |  |
|  | MFF, right leg | QMT: knee extension strength of vastus muscles | --- | --- | --- | -0.51 | <0.001 | --- |  |
|  | MFF, left leg | MRC score: knee flexion | --- | --- | --- | -0.74 | <0.001 | --- |  |
|  | MFF, left leg | MRC score: knee extension | --- | --- | --- | -0.58 | <0.001 | --- |  |
|  | MFF, left leg | MRC score: foot dorsal flexion | --- | --- | --- | -0.76 | <0.001 | --- |  |
|  | MFF, left leg | MRC score: plantar flexors | --- | --- | --- | -0.43 | <0.001 | --- | Weak |
|  | MFF, left leg | Quantitative isometric knee extensor strength | --- | --- | --- | -0.37 | <0.001 | --- |  |
|  | MFF, left leg | MMT: knee extension strength of vastus muscles | --- | --- | --- | -0.79 | <0.001 | --- |  |
|  | MFF, left leg | QMT: knee extension strength of vastus muscles | --- | --- | --- | -0.46 | <0.001 | --- |  |
| Mul 2021^b75^ | FSHD-RODS | MFM | 153 | --- | Spearman | 0.86 | --- | --- |  |
| Mul 2022^76^ | FFS | Communicative Participation Item Bank (CPIB) | 155 | Baseline | Pearson | 0.733 | <0.001 | --- | Strong |
|  | FFS | Facial weakness subscale | 155 | Baseline | Pearson | -0.566 | <0.001 | --- | Moderate |
|  | FFS | FSHD evaluation scale facial weakness subscore | 155 | Baseline | Pearson | 0.441 | <0.001 | --- | Moderate |
| Regula 2016^56^ | MRC score: sternonceidomastoideus | MRI score: sternonceidomastoideus | 20 muscles | Baseline | Spearman | 0.12 | 0.3 | --- |  |
|  | MRC score: trapezius | MRI score: trapezius | 38 muscles | Baseline | Spearman | -0.57 | <0.001 | --- |  |
|  | MRC score: deltoideus | MRI score: deltoideus | 37 muscles | Baseline | Spearman | -0.04 | 0.41 | --- |  |
|  | MRC score: pectoralis | MRI score: pectoralis | 37 muscles | Baseline | Spearman | -0.73 | <0.001 | --- |  |
|  | MRC score: rhomboideus | MRI score: rhomboideus | 12 muscles | Baseline | Spearman | -0.71 | 0.005 | --- |  |
|  | MRC score: supraspinatus | MRI score: supraspinatus | 8 muscles | Baseline | Spearman | --- | --- | --- |  |
|  | MRC score: infraspinatus | MRI score: infraspinatus | 38 muscles | Baseline | Spearman | -0.33 | 0.02 | --- |  |
|  | MRC score: serratus anterior | MRI score: serratus anterior | 24 muscles | Baseline | Spearman | -0.32 | 0.067 | --- |  |
|  | MRC score: biceps brachii | MRI score: biceps brachii | 36 muscles | Baseline | Spearman | -0.42 | 0.006 | --- |  |
|  | MRC score: triceps brachii | MRI score: triceps brachii | 35 muscles | Baseline | Spearman | -0.38 | 0.012 | --- |  |
|  | MRC scoe: gluteus maximus | MRI scoe: gluteus maximus | 40 muscles | Baseline | Spearman | -0.20 | 0.111 | --- |  |
|  | MRC score: iliopsoas | MRI score: iliopsoas | 40 muscles | Baseline | Spearman | -0.34 | 0.017 | --- |  |
|  | MRC score: gluteus medius | MRI score: gluteus medius | 40 muscles | Baseline | Spearman | -0.43 | 0.003 | --- |  |
|  | MRC score: adductors | MRI score: adductors | 40 muscles | Baseline | Spearman | -0.43 | 0.003 | --- |  |
|  | MRC score: quadriceps | MRI score: quadriceps | 40 muscles | Baseline | Spearman | -0.47 | 0.001 | --- |  |
|  | MRC score: semimembranosus | MRI score: semimembranosus | 40 muscles | Baseline | Spearman | -0.86 | <0.001 | --- |  |
|  | MRC score: tibialis anterior | MRI score: tibialis anterior | 8 muscles | Baseline | Spearman | -0.81 | 0.008 | --- |  |
|  | MRC score: triceps surae | MRI score: triceps surae | 8 muscles | Baseline | Spearman | -0.76 | 0.007 | --- |  |
|  | MRC score peroneus longus | MRI score peroneus longus | 8 muscles | Baseline | Spearman | NR | NR | --- |  |
| Rijken 2015^103^ | 6MWT | 10MWT | 9 | Baseline | Spearman | 0.95 | <0.01 | --- |  |
|  | 6MWT | Berg Balance Scale | 9 | Baseline | Spearman | 0.88 | <0.01 | --- |  |
|  | 6MWT | TUG | 9 | Baseline | Spearman | -0.95 | <0.01 | --- |  |
|  | 10MWT | Berg Balance Scale | 9 | Baseline | Spearman | 0.85 | <0.01 | --- |  |
|  | 10MWT | TUG | 9 | Baseline | Spearman | -0.87 | <0.01 | --- |  |
|  | TUG | Berg Balance Scale | 9 | Baseline | Spearman | -0.88 | <0.01 | --- |  |
|  | 6MWT | Step Up test | 9 | Baseline | Spearman | 0.59 | --- | --- |  |
|  | 6MWT | Step Down test | 9 | Baseline | Spearman | 0.53 | <0.05 | --- |  |
|  | 6MWT | Sit-to-Stand test | 9 | Baseline | Spearman | 0.54 | --- | --- |  |
|  | 6MWT | Stance-to-Sit test | 9 | Baseline | Spearman | 0.81 | <0.01 | --- |  |
|  | 10MWT | Step Up test | 9 | Baseline | Spearman | 0.52 | --- | --- |  |
|  | 10MWT | Step Down test | 9 | Baseline | Spearman | 0.47 | --- | --- |  |
|  | 10MWT | Sit-to-Stand test | 9 | Baseline | Spearman | 0.54 | --- | --- |  |
|  | 10MWT | Stance-to-Sit test | 9 | Baseline | Spearman | 0.72 | <0.05 | --- |  |
|  | Berg Balance Scale | Step Up test | 9 | Baseline | Spearman | 0.61 | --- | --- |  |
|  | Berg Balance Scale | Step Down test | 9 | Baseline | Spearman | 0.70 | <0.05 | --- |  |
|  | Berg Balance Scale | Sit-to-Stand test | 9 | Baseline | Spearman | 0.67 | --- | --- |  |
|  | Berg Balance Scale | Stance-to-Sit test | 9 | Baseline | Spearman | 0.70 | <0.05 | --- |  |
|  | TUG | Step Up test | 9 | Baseline | Spearman | -0.62 | --- | --- |  |
|  | TUG | Step Down test | 9 | Baseline | Spearman | -0.61 | --- | --- |  |
|  | TUG | Sit-to-Stand test | 9 | Baseline | Spearman | -0.59 | --- | --- |  |
|  | TUG | Stand-to-Sit test | 9 | Baseline | Spearman | -0.83 | <0.01 | --- |  |
|  | Step Up test | Step Down test | 9 | Baseline | Spearman | 0.90 | <0.01 |  |  |
|  | Step Up test | Sit-to-Stand test | 9 | Baseline | Spearman | 0.87 | <0.01 |  |  |
|  | Step Up test | Stand-to-Sit test | 9 | Baseline | Spearman | 0.89 | <0.01 |  |  |
|  | Step Down test | Sit-to-Stand test | 9 | Baseline | Spearman | 0.89 | <0.01 |  |  |
|  | Step Down test | Stand-to-Sit test | 9 | Baseline | Spearman | 0.91 | <0.01 |  |  |
|  | Sit-to-Stand test | Stand-to-Sit test | 9 | Baseline | Spearman | 0.83 | <0.01 |  |  |
| Statland 2016^67^ | EIM 50 Hz: right vastus lateralis | Age | 35 | Baseline | Pearson | -0.22 | 0.22 | --- |  |
|  | EIM 50 Hz: right vastus lateralis | Disease duration | 35 | Baseline | Pearson | -0.39 | 0.02 | --- |  |
|  | EIM 50 Hz: right vastus lateralis | FCS | 35 | Baseline | Pearson | -0.60 | 0.0003 | --- |  |
|  | EIM 50 Hz: right vastus lateralis | 6MWT | 35 | Baseline | Pearson | 0.69 | <0.0001 | --- |  |
|  | EIM 50 Hz: right vastus lateralis | Ascend 4 stairs | 35 | Baseline | Pearson | -0.65 | <0.0001 | --- |  |
|  | EIM 50 Hz: right vastus lateralis | Don/doff coat | 35 | Baseline | Pearson | -0.58 | 0.0003 | --- |  |
|  | EIM 50 Hz: right vastus lateralis | Supine-to-Sit | 35 | Baseline | Pearson | -0.64 | <0.0001 | --- |  |
|  | EIM 50 Hz: right vastus lateralis | Combined MMT | 35 | Baseline | Pearson | 0.63 | <0.0001 | --- |  |
|  | EIM 50 Hz: right vastus lateralis | Combined LEXT MMT | 35 | Baseline | Pearson | 0.57 | 0.0004 | --- |  |
|  | EIM 50 Hz: right vastus lateralis | MMT Uext | 35 | Baseline | Pearson | 0.46 | 0.006 | --- |  |
|  | EIM 50 Hz: right vastus lateralis | MMT Lext | 35 | Baseline | Pearson | 0.57 | 0.0004 | --- |  |
|  | EIM 50 Hz: right vastus lateralis | RKE QMT Std | 35 | Baseline | Pearson | 0.49 | 0.003 | --- |  |
|  | EIM 50 Hz: right vastus lateralis | REF QMT PP | 35 | Baseline | Pearson | 0.50 | 0.005 | --- |  |
|  | EIM 50 Hz: right bicep | Age | 35 | Baseline | Pearson | -0.38 | 0.03 | --- |  |
|  | EIM 50 Hz: right bicep | Diseae Duration | 35 | Baseline | Pearson | -0.41 | 0.02 | --- |  |
|  | EIM 50 Hz: right bicep | FCS | 35 | Baseline | Pearson | -0.50 | 0.003 | --- |  |
|  | EIM 50 Hz: right bicep | 6MWT | 35 | Baseline | Pearson | 0.38 | 0.02 | --- |  |
|  | EIM 50 Hz: right bicep | Ascend 4 stairs | 35 | Baseline | Pearson | -0.39 | 0.02 | --- |  |
|  | EIM 50 Hz: right bicep | Don/doff coat | 35 | Baseline | Pearson | -0.60 | 0.0002 | --- |  |
|  | EIM 50 Hz: right bicep | Supine-to-Sit | 35 | Baseline | Pearson | -0.50 | 0.002 | --- |  |
|  | EIM 50 Hz: right bicep | Combined MMT | 35 | Baseline | Pearson | 0.54 | 0.0007 | --- |  |
|  | EIM 50 Hz: right bicep | MMT Uext | 35 | Baseline | Pearson | 0.62 | <0.0001 | --- |  |
|  | EIM 50 Hz: right bicep | MMT Lext | 35 | Baseline | Pearson | 0.33 | 0.05 | --- |  |
|  | EIM 50 Hz: right bicep | RKE QMT PP | 35 | Baseline | Pearson | 0.09 | 0.59 | --- |  |
|  | EIM 50 Hz: right bicep | REF QMT PP | 35 | Baseline | Pearson | 0.68 | <0.0001 | --- |  |
|  | EIM 50 Hz: right thoracic paraspinal | Age | 35 | Baseline | Pearson | -0.33 | 0.05 | --- |  |
|  | EIM 50 Hz: right thoracic paraspinal | Disease Duration | 35 | Baseline | Pearson | -0.44 | 0.008 | --- |  |
|  | EIM 50 Hz: right thoracic paraspinal | FCS | 35 | Baseline | Pearson | -0.51 | 0.002 | --- |  |
|  | EIM 50 Hz: right thoracic paraspinal | 6MWT | 35 | Baseline | Pearson | 0.37 | 0.03 | --- |  |
|  | EIM 50 Hz: right thoracic paraspinal | Ascend 4 stairs | 35 | Baseline | Pearson | -0.42 | 0.01 | --- |  |
|  | EIM 50 Hz: right thoracic paraspinal | Don/doff coat | 35 | Baseline | Pearson | -0.49 | 0.003 | --- |  |
|  | EIM 50 Hz: right thoracic paraspinal | Supine-to-Sit | 35 | Baseline | Pearson | -0.50 | 0.002 | --- |  |
|  | EIM 50 Hz: right thoracic paraspinal | Combined MMT | 35 | Baseline | Pearson | 0.54 | 0.0009 | --- |  |
|  | EIM 50 Hz: right thoracic paraspinal | MMT Uext | 35 | Baseline | Pearson | 0.51 | 0.002 | --- |  |
|  | EIM 50 Hz: right thoracic paraspinal | MMT Lext | 35 | Baseline | Pearson | 0.43 | 0.01 | --- |  |
|  | EIM 50 Hz: right thoracic paraspinal | RKE QMT PP | 35 | Baseline | Pearson | 0.05 | 0.77 | --- |  |
|  | EIM 50 Hz: right thoracic paraspinal | REF QMT PP | 35 | Baseline | Pearson | 0.37 | 0.05 | --- |  |
|  | EIM 50 Hz: leg | Age | 35 | Baseline | Pearson | -0.39 | 0.02 | --- |  |
|  | EIM 50 Hz: leg | Disease Duration | 35 | Baseline | Pearson | -0.52 | 0.001 | --- |  |
|  | EIM 50 Hz: leg | FCS | 35 | Baseline | Pearson | -0.72 | <0.0001 | --- |  |
|  | EIM 50 Hz: leg | 6MWT | 35 | Baseline | Pearson | 0.74 | <0.0001 | --- |  |
|  | EIM 50 Hz: leg | Ascend 4 stairs | 35 | Baseline | Pearson | -0.65 | <0.0001 | --- |  |
|  | EIM 50 Hz: leg | Don/doff coat | 35 | Baseline | Pearson | -0.65 | <0.0001 | --- |  |
|  | EIM 50 Hz: leg | Supine-to-Sit | 35 | Baseline | Pearson | -0.69 | <0.0001 | --- |  |
|  | EIM 50 Hz: leg | Combined MMT | 35 | Baseline | Pearson | 0.76 | <0.0001 | --- |  |
|  | EIM 50 Hz: leg | MMT Uext | 35 | Baseline | Pearson | 0.60 | 0.0002 | --- |  |
|  | EIM 50 Hz: leg | MMT Lext | 35 | Baseline | Pearson | 0.70 | <0.0001 | --- |  |
|  | EIM 50 Hz: leg | RKE QMT PP | 35 | Baseline | Pearson | 0.48 | 0.003 | --- |  |
|  | EIM 50 Hz: leg | REF QMT PP | 35 | Baseline | Pearson | 0.54 | 0.002 | --- |  |
|  | EIM 50 Hz: arm | Age | 35 | Baseline | Pearson | -0.33 | 0.06 | --- |  |
|  | EIM 50 Hz: arm | Disease Duration | 35 | Baseline | Pearson | -0.55 | 0.0008 | --- |  |
|  | EIM 50 Hz: arm | FCS | 35 | Baseline | Pearson | -0.53 | 0.001 | --- |  |
|  | EIM 50 Hz: arm | 6MWT | 35 | Baseline | Pearson | 0.45 | 0.007 | --- |  |
|  | EIM 50 Hz: arm | Ascend 4 stairs | 35 | Baseline | Pearson | -0.53 | 0.002 | --- |  |
|  | EIM 50 Hz: arm | Don/doff coat | 35 | Baseline | Pearson | -0.70 | <0.0001 | --- |  |
|  | EIM 50 Hz: arm | Supine-to-Sit | 35 | Baseline | Pearson | -0.58 | 0.0002 | --- |  |
|  | EIM 50 Hz: arm | Combined MMT | 35 | Baseline | Pearson | 0.63 | <0.0001 | --- |  |
|  | EIM 50 Hz: arm | MMT Uext | 35 | Baseline | Pearson | 0.71 | <0.0001 | --- |  |
|  | EIM 50 Hz: arm | MMT Lext | 35 | Baseline | Pearson | 0.38 | 0.03 | --- |  |
|  | EIM 50 Hz: arm | RKE QMT PP | 35 | Baseline | Pearson | 0.09 | 0.62 | --- |  |
|  | EIM 50 Hz: arm | REF QMT PP | 35 | Baseline | Pearson | 0.57 | 0.001 | --- |  |
|  | EIM 50 Hz: trunk | Age | 35 | Baseline | Pearson | -0.41 | 0.01 | --- |  |
|  | EIM 50 Hz: trunk | Disease Duration | 35 | Baseline | Pearson | -0.41 | 0.02 | --- |  |
|  | EIM 50 Hz: trunk | FCS | 35 | Baseline | Pearson | -0.54 | 0.001 | --- |  |
|  | EIM 50 Hz: trunk | 6MWT | 35 | Baseline | Pearson | 0.48 | 0.004 | --- |  |
|  | EIM 50 Hz: trunk | Ascend 4 stairs | 35 | Baseline | Pearson | -0.47 | 0.006 | --- |  |
|  | EIM 50 Hz: trunk | Don/doff coat | 35 | Baseline | Pearson | -0.54 | 0.0007 | --- |  |
|  | EIM 50 Hz: trunk | Supine-to-Sit | 35 | Baseline | Pearson | -0.53 | 0.001 | --- |  |
|  | EIM 50 Hz: trunk | Combined MMT | 35 | Baseline | Pearson | 0.56 | 0.0004 | --- |  |
|  | EIM 50 Hz: trunk | MMT Uext | 35 | Baseline | Pearson | 0.51 | 0.002 | --- |  |
|  | EIM 50 Hz: trunk | MMT Lext | 35 | Baseline | Pearson | 0.47 | 0.004 | --- |  |
|  | EIM 50 Hz: trunk | RKE QMT PP | 35 | Baseline | Pearson | 0.04 | 0.82 | --- |  |
|  | EIM 50 Hz: trunk | REF QMT PP | 35 | Baseline | Pearson | 0.40 | 0.03 | --- |  |
|  | EIM 50 Hz: face | Age | 35 | Baseline | Pearson | -0.37 | 0.04 | --- |  |
|  | EIM 50 Hz: face | Disease Duration | 35 | Baseline | Pearson | -0.34 | 0.05 | --- |  |
|  | EIM 50 Hz: face | FCS | 35 | Baseline | Pearson | -0.43 | 0.02 | --- |  |
|  | EIM 50 Hz: face | 6MWT | 35 | Baseline | Pearson | 0.30 | 0.1 | --- |  |
|  | EIM 50 Hz: face | Ascend 4 stairs | 35 | Baseline | Pearson | -0.25 | 0.19 | --- |  |
|  | EIM 50 Hz: face | Don/doff coat | 35 | Baseline | Pearson | -0.22 | 0.24 | --- |  |
|  | EIM 50 Hz: face | Supine-to-Sit | 35 | Baseline | Pearson | -0.26 | 0.15 | --- |  |
|  | EIM 50 Hz: face | Combined MMT | 35 | Baseline | Pearson | 0.40 | 0.02 | --- |  |
|  | EIM 50 Hz: face | MMT Uext | 35 | Baseline | Pearson | 0.30 | 0.09 | --- |  |
|  | EIM 50 Hz: face | MMT Lext | 35 | Baseline | Pearson | 0.40 | 0.02 | --- |  |
|  | EIM 50 Hz: face | RKE QMT PP | 35 | Baseline | Pearson | 0.32 | 0.08 | --- |  |
|  | EIM 50 Hz: face | REF QMT PP | 35 | Baseline | Pearson | 0.16 | 0.44 | --- |  |
| Tawil 1997^105^ | Composite MMT score | Composite MVICT score | 81 | Baseline | R^2^ | 0.878 | --- | --- | "strong" |
|  | Composite MMT score | 24hr-Creatinine (indicator of muscle mass) | 82 | Baseline | R^2^ | 0.813 | --- | --- |  |
|  | Composite MVICT score | 24hr-Creatinine (indicator of muscle mass) | 83 | Baseline | R^2^ | 0.808 | --- | --- |  |
| Teeselink 2022^68^ | FVC% predicted | Age at baseline | 92 | Baseline | Spearman | -0.035 | 0.74 | --- | Weak |
|  | FVC% predicted | BMI at baseline | 92 | Baseline | Spearman | -0.014 | 0.898 | --- | Weak |
|  | FVC% predicted | CSS at baseline | 92 | Baseline | Spearman | -0.297 | 0.004 | --- | Weak |
|  | FVC% predicted | FCS at baseline | 92 | Baseline | Spearman | -0.311 | 0.003 | --- | Weak |
|  | FVC% predicted | MMT sum score at baseline | 92 | Baseline | Spearman | 0.268 | 0.01 | --- | Weak |
|  | FVC% predicted | MFM trunk score % of total at baseline | 92 | Baseline | Spearman | 0.373 | <0.001 | --- | Weak |
|  | FVC% predicted | MFM score % of total at baseline | 92 | Baseline | Spearman | 0.36 | <0.001 | --- | Weak |
|  | Change in FVC% predicted | age at baseline | 92 | 5-yr follow-up | Spearman | 0.187 | 0.075 | --- | Did not correlate |
|  | Change in FVC% predicted | BMI at baseline | 92 | 5-yr follow-up | Spearman | 0.08 | 0.463 | --- | Did not correlate |
|  | Change in FVC% predicted | CSS at baseline | 92 | 5-yr follow-up | Spearman | 0.151 | 0.152 | --- | Did not correlate |
|  | Change in FVC% predicted | FCS at baseline | 92 | 5-yr follow-up | Spearman | 0.185 | 0.078 | --- | Did not correlate |
|  | Change in FVC% predicted | MMT sum score at baseline | 92 | 5-yr follow-up | Spearman | -0.204 | 0.051 | --- | Did not correlate |
|  | Change in FVC% predicted | MFM trunk score % of total at baseline | 92 | 5-yr follow-up | Spearman | -0.184 | 0.080 | --- | Did not correlate |
|  | Change in FVC% predicted | MFM score % of total at baseline | 92 | 5-yr follow-up | Spearman | -0.187 | 0.074 | --- | Did not correlate |
|  | Change in FVC% predicted | Change in CSS | 92 | 5-yr follow-up | Spearman | 0.018 | 0.862 | --- | Did not correlate |
|  | Change in FVC% predicted | Change in FCS | 92 | 5-yr follow-up | Spearman | -0.059 | 0.577 | --- | Did not correlate |
|  | Change in FVC% predicted | Change in MMT sum score | 92 | 5-yr follow-up | Spearman | 0.087 | 0.412 | --- | Did not correlate |
|  | Change in FVC% predicted | Change in MFM trunk score % of total | 92 | 5-yr follow-up | Spearman | 0.205 | 0.05 | --- | Did not correlate |
|  | Change in FVC% predicted | Change in MFM score % of total | 92 | 5-yr follow-up | Spearman | 0.245 | 0.019 | --- | Weak |

---, not reported; 5TSTST, 5times sit to stand test; 6MWT, 6-minute walk test; 10MWT, 10-minute walk test; 14SST, 14 step stair test; BMI, body mass index; CSS, clinical severity score; EIM, electrical impedance myography; eTUG, electronic timed up and go; FCS, FSHD clinical score; FSHD; facioscapulohumeral muscular dystrophy; FSHD-COM, FSHD composite outcome measures; FSHD-HI, FSHD health index; FVC, forced vital capacity; IOPI, Iowa Oral Performance Instrument; IPT, isokinetic peak torque; LE, lower extremity; LMV, lower muscle volume; MFF, muscle fat fraction; MFI, muscle fat infiltration; MFM; Motor Function Measures; MMSE, Mini-Mental State Examination; MMT, manual muscle testing; MoCA, Montreal Cognitive Assessment; MPI, mean pixel intensity; MRC, Medical Research Council; MVC, maximum voluntary contractions; RSA, relative surface area; RWS, reachable work space; TUG, timed up and go; UE, upper extremity

Table 18. Construct validity outcomes using Rasch Analysis for the topic “Disease Outcome Measures and Validation”

| **Author, year** | **Assessment measure** | **n** | **Analysis** | **Item location** | **Item fit residual** | **Person location** | **Person fit residual** | **Item trait interaction (df; p-value)** | **PSI** | **Floor or ceiling effect** | **Notes** |
| --- | --- | --- | --- | --- | --- | --- | --- | --- | --- | --- | --- |
| FULL TEXT ARTICLES | | | | | | | | | | | |
| Mul 2021^69^ | FSHD-RODS | 762 | Final | --- | -0.389 ± 0.940 | --- | -0.231 ± 0.788 | 342; 0.31 | 0.98 | --- | Unable to test unidimensionality |
| Mul 2021^b75^ | Total MFM | 194 | --- | 0 ± 2.96 | -0.46 ± 1.46 | 3.53 ± 2.52 | -0.26 ± 0.67 | 64; 0.0000 | 0.94 | Ceiling: 14% | Unidimensionality test: 18.1% (95%CI: 14.8 – 21.4) |
| Mul 2022^76^ | FFS | --- | Final | 0.00 ± 0.99 | -0.33 ± 1.16 | -1.84 ± 1.45 | -0.27 ± 0.87 | 50; 0.678 | 0.92 | Floor: 7% | Unidimensionality test: 6.3% (2.3 – 10.4) |

Note: all values presented as mean ± standard deviation

---, not reported; CI, confidence interval; df, degrees of freedom; FFS, Facial Function Scale; FHSD-RODS, facioscapulohumeral muscular dystrophy Rasch-built overall disability scale; MFM, Motor Functional Measure; PSI, Person Separation Index.

Table 19. Construct validity outcomes using factor loads for the topic “Disease Outcome Measures and Validation”

| **Author, year** | **Assessment measure** | **n** | **Value** | **Notes** |
| --- | --- | --- | --- | --- |
| CONFERENCE ABSTRACTS | | | | |
| Haley 2020^77^ | FCS | 42 | CFA confirmed that a one-factor model was appropriate for determining dimensionality of the items on the IOPI with FVC. | Facial weakness and abdominal muscle involvement contributed the least to total FSHD CS score. |
| FULL TEXT ARTICLES | | | | |
| Guillot 2018^72^ | MFM D1 | 193 | Range: 0.31-0.90 | Factor loads presented in figure. |
|  | MFM D2 |  | Range: 0.68-0.95 |  |
|  | MFM D3 |  | Range: 0.35-0.77 |  |

CFA, confirmatory factor analysis; CS, clinical score; D, dimension; FCS, FSHC clinical score; FSHD, facioscapulohumeral muscular dystrophy; FVC, forced vital capacity; IOPI, Iowa Oral Performance Instrument; MFM, Motor Function Measure

Table 20. Reliability outcomes for the topic “Disease Outcome Measures and Validation”

| **Author, year** | **Measure** | **n** | **Number reported measurements and interval** | **Number of raters** | **Reliability** | **Measure of reliability** | **Value  (95% CI)** | **p-value** | **Notes** |
| --- | --- | --- | --- | --- | --- | --- | --- | --- | --- |
| CONFERENCE ABSTRACTS | | | | | | | | | |
| Berggren 2021^80^ | IOPI | 227 | 12 study sites; --- | --- | Inter-rater | --- | 0.66 – 0.85^a^ | --- | Represents reliability scores across study sites for orofacial strength and endurance measures |
| Chan 2020^82^ | TUG | 22 | 2; one week | --- | Test-retest | --- | --- | --- | “Strong reliability” |
|  | oTUG | 22 | 2; one week | --- | Test-retest | --- | --- | --- | “Strong reliability” |
| Eichinger 2017^85^ | FSHD-COM | 36 | 4; baseline, <3 weeks, 6- and 12-months | --- | --- | ICC | 0.97 | --- | Subscale 0.9 – 0.98. |
| Hatch 2022^87^ | RWS Q1, Q3, Q4 and total | 167 | 2; one-day | --- | Test-retest | ICC | >0.90 | <0.001 |  |
|  | RWS Q2 |  | 2; one-day | --- | Test-retest | ICC | ~0.85 | <0.001 |  |
| Statland 2016^93^ | Wireless gait | 5 | 2; within 2 weeks | --- | Test-retest | ICC | 0.85-0.99 | --- | Good to excellent for spatial and temporal gait metrics. |
| Statland 2022^91^ | FSHD-COM | 240 | 5; baseline, 3- / 12- / 18- / 24-months | --- | Test-retest | ICC | 0.98 | 0.97^b^ | Excellent |
|  | MFM D1 |  |  | --- | Test-retest | ICC | 0.97 | 0.96^b^ | Excellent |
| FULL TEXT ARTICLES | | | | | | | | | |
| Bachasson 2014^99^ | MVC | 12 | 4; --- | --- | Test-retest | ICC | 0.99 (0.99 – 0.99) | --- | 50% volitional values and evoked/central parameter values available. |
|  |  | 12 |  | --- | Test-retest | CV | 2.3 (1.6 – 3.9) | --- |  |
| de Valle 2021^64^ | FSHD-COM | 15 | 2; mean 14.9 days (range: 4 – 26) | 1 | Test-retest | ICC | 0.995  (0.985-0.998) | --- |  |
|  |  |  |  | 1 | Test-retest | SEM | 1.25  (0.9-2.0) | --- |  |
|  | FSHD-COM Peds | 15 | 2; mean 14.9 days (range: 4 – 26) | 1 | Test-retest | ICC | 0.995 (0.987-0.998) | --- |  |
|  |  |  |  | 1 | Test-retest | SEM | 0.31 (0.96 – 2.1) | --- |  |
| Eichinger 2017^61^ | 6MWT | 25 | 2; baseline versus <3weeks | --- | Test-retest | ICC | 0.99 (0.98)^b^ | --- | Excellent |
| Eichinger 2018^84^ | FSHD-COM | 32 | 2; within 3 weeks | --- | Test-retest | ICC | 0.96 (0.92)^b^ | --- | Component scores available. |
| Eren 2020^73^ | Novel shoulder staging system | 37 | 2; within 2 weeks | 2 | Inter-rater | Kappa | 0.872 | --- | Excellent reliability; reliability available for subcomponents or staging system |
|  |  | 38 | 2; within 2 weeks | 2 | Intra-rater | Kappa | 0.822; 0.821 | --- |  |
| Huisinga 2018^102^ | iTUG: stride velocity | 14 | 2; within 2 weeks | --- | Test-retest | ICC | 0.99 | <0.01 |  |
|  | iTUG sagittal trunk ROM | 14 | 2; within 2 weeks | --- | Test-retest | ICC | 0.99 | <0.01 |  |
|  | iTUG total duration | 14 | 2; within 2 weeks | --- | Test-retest | ICC | 0.99 | 0.03 |  |
|  | iTUG stride length | 14 | 2; within 2 weeks | --- | Test-retest | ICC | 0.97 | 0.02 |  |
|  | iTUG cadence | 14 | 2; within 2 weeks | --- | Test-retest | ICC | 0.99 | 0.03 |  |
|  | iTUG double support | 14 | 2; within 2 weeks | --- | Test-retest | ICC | 0.99 | 0.07 |  |
|  | iTUG knee ROM | 14 | 2; within 2 weeks | --- | Test-retest | ICC | 0.96 | 0.02 |  |
|  | iTUG horizontal trunk ROM | 14 | 2; within 2 weeks | --- | Test-retest | ICC | 0.99 | 0.13 |  |
|  | iTUG arm ROM | 14 | 2; within 2 weeks | --- | Test-retest | ICC | 0.84 | 0.84 |  |
|  | iTUG turn duration | 14 | 2; within 2 weeks | --- | Test-retest | ICC | 0.95 | 0.17 |  |
| Lamperti 2010^71^ | FCS: total | 69 | ---; --- | 2 | Inter-rater | Kappa | 0.7744 | --- | Available for individual components. |
| Lareau-Trudel 2015^57^ | IFF with qMRI | 35 | 2; --- | 2 | Inter-rater | ICC | 0.97 (CV: 3.3±4.7) | --- |  |
|  |  | 35 |  | 2 | Intra-rater | ICC | 0.98 (CV: 5.6±4.6) | --- |  |
|  | Muscle fraction | 35 | 2; --- | 2 | Inter-rater | ICC | 0.97  (CV: 3.2±4) | --- |  |
|  |  | 35 |  | 2 | Intra-rater | ICC | 0.98 (CV: 4.8±2.9) | --- |  |
| Mul 2021^69^ | FSHD-RODS | 230 | 2; 2 – 4 weeks | --- | Test-retest | R^2^ | 0.98 | --- | Item hierarchy |
|  |  | 230 |  | --- | Test-retest | R2 | 0.97 | --- | Patient abilities |
| Mul 2022^76^ | FFS | 73 | 2; 2 weeks | --- | Test-retest | ICC | 0.978 | --- | Excellent |
| Personius 1994^70^ | QMT: dorsiflexor R | 32 | 2; first 2 days of testing | 2 | Intra-rater | ICC | 0.98 | --- | Available for consecutive days as well (inter-rater). |
|  | QMT: dorsiflexor L | 32 |  | 2 | Intra-rater | ICC | 0.97 | --- |  |
|  | QMT abductor R | 32 | 2; first 2 days of testing | 2 | Intra-rater | ICC | 0.91 | --- |  |
|  | QMT: abductor L | 32 |  | 2 | Intra-rater | ICC | 0.97 | --- |  |
|  | QMT: elbow extensor R | 32 | 2; first 2 days of testing | 2 | Intra-rater | ICC | 0.96 | --- |  |
|  | QMT: elbow extensor L | 32 |  | 2 | Intra-rater | ICC | 0.96 | --- |  |
|  | QMT: elbow flexor R | 32 | 2; first 2 days of testing | 2 | Intra-rater | ICC | 0.99 | --- |  |
|  | QMT: elbow flexor L | 32 |  | 2 | Intra-rater | ICC | 0.98 | --- |  |
|  | QMT: lateral rotator R | 32 | 2; first 2 days of testing | 2 | Intra-rater | ICC | 0.95 | --- |  |
|  | QMT: lateral rotator L | 32 |  | 2 | Intra-rater | ICC | 0.91 | --- |  |
|  | QMT: horizontal abductor R | 32 | 2; first 2 days of testing | 2 | Intra-rater | ICC | 0.93 | --- |  |
|  | QMT: horizontal abductor L | 32 |  | 2 | Intra-rater | ICC | 0.91 | --- |  |
|  | QMT: horizontal adductor R | 32 | 2; first 2 days of testing | 2 | Intra-rater | ICC | 0.95 | --- |  |
|  | QMT: horizontal adductor L | 32 |  | 2 | Intra-rater | ICC | 0.91 | --- |  |
|  | QMT: knee extensor R | 32 | 2; first 2 days of testing | 2 | Intra-rater | ICC | 0.96 | --- |  |
|  | QMT: knee extensor L | 32 |  | 2 | Intra-rater | ICC | 0.96 | --- |  |
|  | QMT: knee flexor R | 32 | 2; first 2 days of testing | 2 | Intra-rater | ICC | 0.94 | --- |  |
|  | QMT: knee flexor L | 32 |  | 2 | Intra-rater | ICC | 0.96 | --- |  |
| Personius 1994 | MMT: dorsiflexor R | 32 | 2; first 2 days of testing | 2 | Intra-rater | Kappa | 0.90 | --- | Available for inter-rater reliability as well. |
|  | MMT: dorsiflexor L | 32 |  | 2 | Intra-rater | Kappa | 0.92 | --- |  |
|  | MMT: shoulder abductor R | 32 | 2; first 2 days of testing | 2 | Intra-rater | Kappa | 0.94 | --- |  |
|  | MMT: shoulder abductor L | 32 |  | 2 | Intra-rater | Kappa | 0.89 | --- |  |
|  | MMT: elbow extensor R | 32 | 2; first 2 days of testing | 2 | Intra-rater | Kappa | 0.92 | --- |  |
|  | MMT: elbow extensor L | 32 |  | 2 | Intra-rater | Kappa | 0.93 | --- |  |
|  | MMT: elbow flexor R | 32 | 2; first 2 days of testing | 2 | Intra-rater | Kappa | 0.98 | --- |  |
|  | MMT: elbow flexor L | 32 |  | 2 | Intra-rater | Kappa | 0.97 | --- |  |
|  | MMT: lateral rotator R | 32 | 2; first 2 days of testing | 2 | Intra-rater | Kappa | 0.79 | --- |  |
|  | MMT: lateral rotator L | 32 |  | 2 | Intra-rater | Kappa | 0.82 | --- |  |
|  | MMT: horizontal abductor R | 32 | 2; first 2 days of testing | 2 | Intra-rater | Kappa | 0.92 | --- |  |
|  | MMT: horizontal abductor L | 32 |  | 2 | Intra-rater | Kappa | 0.81 | --- |  |
|  | MMT: horizontal adductor R | 32 | 2; first 2 days of testing | 2 | Intra-rater | Kappa | 0.87 | --- |  |
|  | MMT: horizontal adductor L | 32 |  | 2 | Intra-rater | Kappa | 0.90 | --- |  |
|  | MMT: knee extensor R | 32 | 2; first 2 days of testing | 2 | Intra-rater | Kappa | 0.92 | --- |  |
|  | MMT: knee extensor L | 32 |  | 2 | Intra-rater | Kappa | 0.83 | --- |  |
|  | MMT: knee flexor R | 32 | 2; first 2 days of testing | 2 | Intra-rater | Kappa | 0.85 | --- |  |
|  | MMT: knee flexor L | 32 |  | 2 | Intra-rater | Kappa | 0.89 | --- |  |
|  | Grip strength R | 31 | 2; first 2 days of testing | 2 | Intra-rater | ICC | 0.97 | --- |  |
|  | Grip strength L | 31 |  | 2 | Intra-rater | ICC | 0.94 | --- |  |
|  | Pinch strength R | 22 | 2; first 2 days of testing | 2 | Intra-rater | ICC | 0.97 | --- |  |
|  | Pinch strength L | 22 |  | 2 | Intra-rater | ICC | 0.95 | --- |  |
|  | FVC | 32 | 2; first 2 days of testing | 2 | Intra-rater | ICC | 0.95 | --- |  |
|  | Facial muscle testing | 32 | 2; first 2 days of testing | 2 | Intra-rater | ICC | 0.63 | --- |  |
| Rijken 2015^103^ | Step Up Test | 36 | 2; different day within 9 weeks | 3 | Inter-rater | Fleiss K | 0.65 | <0.001 |  |
|  | Step Down Test | 34 |  | 3 | Inter-rater | Fleiss K | 0.67 | <0.001 |  |
|  | Sit-to-Stand | 19 |  | 3 | Inter-rater | Fleiss K | 0.85 | <0.001 |  |
|  | Stand-to-Sit | 19 |  | 3 | Inter-rater | Fleiss K | 0.81 | <0.001 |  |
| Sanson 2022^104^ | Brooke scale | 254 | Comparing CEF & SEQ; within 3 months | 2 | Inter-rater | ICC | 0.66  (0.58 – 0.72) | --- | Other assessments available for correlation between CEF and SEQ |
|  | Vignos scale | 250 |  | 2 | Inter-rater | ICC | 0.86 (0.82 – 0.89) | --- |  |
|  | Visual analogue pain score | 191 |  | 2 | Inter-rater | ICC | 0.61 (0.51 – 0.69( | --- |  |
| Statland 2016^67^ | EIM 50 Hz: right vastus lateralis | 18 | 2; 3 weeks | --- | Test-retest | ICC | Phase:  0.95 (0.87)^b^  Reactance:  0.91 (0.77)^b^ | --- | Also reported for 100 Hz. |
|  | EIM 50 Hz: right bicep | 18 | 2; 3 weeks | --- | Test-retest | ICC | Phase:  0.99 (0.97)^b^ Reactance:  0.92 (0.81)^b^ | --- | Also reported for 100 Hz. |
|  | EIM 50 Hz: right thoracic paraspinal | 18 | 2; 3 weeks | --- | Test-retest | ICC | Phase: 0.91 (0.77)^b^ Reactance:  0.74 (0.43)^b^ | --- | Also reported for 100 Hz. |
|  | EIM 50 Hz: leg | 18 | 2; 3 weeks | --- | Test-retest | ICC | Phase: 0.91 (0.77)^b^ Reactance:  0.89 (0.74)^b^ | --- | Also reported for 100 Hz. |
|  | EIM 50 Hz: arm | 18 | 2; 3 weeks | --- | Test-retest | ICC | Phase: 0.98 (0.94)^b^ Reactance:  0.96 (0.90)^b^ | --- | Also reported for 100 Hz. |
|  | EIM 50 Hz: trunk | 18 | 2; 3 weeks | --- | Test-retest | ICC | Phase:  0.98 (0.95)^b^  Reactance:  0.95 (0.87) | --- | Also reported for 100 Hz. |
|  | EIM 50 Hz: face | 13 | 2; 3 weeks | --- | Test-retest | ICC | Phase: 0.36 (-0.21)^b^ Reactance: 0.70 (0.30)^b^ | --- | Also reported for 100 Hz. |
|  | EIM 50 Hz: right deltoid | 18^c^ | 2; 3 weeks | --- | Test-retest | ICC | Phase: 0.97 (0.92)^b^ Reactance:  0.88 (0.70)^b^ | --- | Also reported for 100 Hz. |
|  | EIM 50 Hz: left deltoid | 18 | 2; 3 weeks | --- | Test-retest | ICC | Phase:  0.93 (0.83)^b^  Reactance:  0.93 (0.81)^b^ | --- | Also reported for 100 Hz. |
|  | EIM 50 Hz: left triceps | 18 | 2; 3 weeks | --- | Test-retest | ICC | Phase: 0.84 (0.62)^b^ Reactance: 0.90 (0.74)^b^ | --- | Also reported for 100 Hz. |
|  | EIM 50 Hz: right triceps | 18 | 2; 3 weeks | --- | Test-retest | ICC | Phase: 0.91 (0.78)^b^ Reactance:  0.91 (0.79)^b^ | --- | Also reported for 100 Hz. |
|  | EIM 50 Hz: left biceps | 18 | 2; 3 weeks | --- | Test-retest | ICC | Phase: 0.92 (0.79)^b^ Reactance: 0.92 (0.80)^b^ | --- | Also reported for 100 Hz. |
|  | EIM 50 Hz: left vastus lateralis | 18 | 2; 3 weeks | --- | Test-retest | ICC | Phase: 0.92 (0.78)^b^ Reactance: 0.89 (0.74)^b^ | --- | Also reported for 100 Hz. |
|  | EIM 50 Hz: left tibialis anterior | 18 | 2; 3 weeks | --- | Test-retest | ICC | Phase:  0.91 (0.77)^b^ Reactance: 0.74 (0.44)^b^ | --- | Also reported for 100 Hz. |
|  | EIM 50 Hz: right tibialis anterior | 18 | 2; 3 weeks | --- | Test-retest | ICC | Phase:  0.76 (0.77)^b^ Reactance: 0.72 (0.39)^b^ | --- | Also reported for 100 Hz. |
|  | EIM 50 Hz: left thoracic paraspinal | 17 | 2; 3 weeks | --- | Test-retest | ICC | Phase:  0.97 (0.93)^b^ Reactance:  0.90 (0.75)^b^ | --- | Also reported for 100 Hz. |
|  | EIM 50 Hz: left abdominal | 18 | 2; 3 weeks | --- | Test-retest | ICC | Phase:  0.95 (0.87)^b^ Reactance:  0.76 (0.46)^b^ | --- | Also reported for 100 Hz. |
|  | EIM 50 Hz: right abdominal | 16 | 2; 3 weeks | --- | Test-retest | ICC | Phase: 0.98 (0.94)^b^ Reactance:  0.78 (0.45)^b^ | --- | Also reported for 100 Hz. |
|  | EIM 50 Hz: left orbicularis oris | 13 | 2; 3 weeks | --- | Test-retest | ICC | Phase: 0.87 (0.62)^b^ Reactance: 0.91 (0.74)^b^ | --- | Also reported for 100 Hz. |
|  | EIM 50 Hz: right orbicularis oris | 13 | 2; 3 weeks | --- | Test-retest | ICC | Phase: 0.15 (0.44)^b^ Reactance:  0.37 (-0.23)^b^ | --- | Also reported for 100 Hz. |
| Widholm 2022^55^ | LMV within-subject SD | 16 | 2; 4-12 weeks | --- | NS | Pearson | 0.97 | <0.001 | Long-term repeatability results available or all muscles and measures. |
|  | LMV within subject CV | 16 | 2; 4-12 weeks | --- | NS | Pearson | -0.39 | 0.11 |  |
|  | MFF within subject SD | 16 | 2; 4-12 weeks | --- | NS | Pearson | 0.14 | 0.59 |  |
|  | MFI within subject SD | 16 | 2; 4-12 weeks | --- | NS | Pearson | -0.09 | -0.72 |  |

^a^ Range

^b^ Lower confidence bound

^c^ Reactance had an n=17

---, not reported; 6MWT, 6 minute walk test; CI, confidence interval; CV, coefficient of variation; D, dimension; EIM, electrical impedance myography; FCS, FSHD clinical score; FFS, Facial Function Scale; FSHD-COM, FSHD composite outcome measure; FVC, forced vital capacity; IOPI, Iowa Oral Performance Instrument; ICC, intraclass correlation coefficient; IFF, infiltrated fat fraction; LMV, lean muscle volume; MFF, muscle fat fraction; MFI, muscle fat infiltration; MFM, Motor Function Measure; MMT, manual muscle testing; MVC, maximum voluntary contraction; NS, not specified; oTUG, optimized TUG; Q, frontal quadrant; QMT, quantitative muscle testing; ROM, range of motion; RWS, Reachable Workspace; SD, standard deviation; TUG, timed up and go

Table 21. Responsiveness outcomes for the topic “Disease Outcome Measures and Validation”

| **Author, year** | **Measure** | **n** | **Timepoint** | **Measure of responsiveness** | **Value (95% CI)** | **p-value** | **Notes** |
| --- | --- | --- | --- | --- | --- | --- | --- |
| CONFERENCE ABSTRACTS | | | | | | | |
| De Lattre 2016^74^ | MFM, D1 | 117 | Two evaluations, at least 6 months apart | Standardized response mean | 0.640 | --- |  |
|  |  |  |  | Sensitivity of score | -2.75 points + -4.3 points per year | --- |  |
| Haley 2020^77^ | FCS | 42 | --- | MDC95 | --- | --- | The FCS is capable of detecting change of as little as one point in 95% of participants. |
| Hristov 2021^78^ | eTUG | 10 | Over 3 months (after 1000 measurements) | MDC95 | 4.65% | --- |  |
|  | In-home gait speed | 10 |  | MDC95 | 1.17% | --- |  |
| FULL TEXT ARTICLES | | | | | | | |
| de Valle 2021^64^ | FSHD-COM | 15 | Average test interval: 14.9 days (range: 4 – 26) | MDC90 | 2.9 (2.1-4.6) | --- |  |
|  |  |  |  | MDC95 | 3.5 (2.1 – 4.6) | --- |  |
|  | FSHD-COM Peds | 15 | Average test interval: 14.9 days (range: 4 – 26) | MDC90 | 3.1 (2.2 – 4.8) | --- |  |
|  |  |  |  | MDC95 | 3.6 (2.2 – 4.8) | --- |  |
| Eichinger 2017^61^ | 6MWT | 86 | --- | MDC95 | 34.3m | --- |  |
| Eichinger 2018^65^ | FSHD-COM | 32 | --- | MDC90 | 6.67 (Δ9.3%) | --- |  |
| Hatch 2021^79^ | RSA total | 21 | --- | ROC cut-off | 0.70 | --- | To achieve 100% sensitivity and 94% specificity (AUC=0.975) |
|  | RSA above shoulder | 21 | --- | ROC cut-off | 0.40 | --- | To achieve 80% sensitivity and 100% specificity (AUC=0.963) |

6MWT, 6-minute walk test; AUC, area under the curve; CI, confidence interval; FCD, FSHD clinical score; MDC, minimal detectable change; ROC, receiver operator characteristic; RSA, relative surface area; TUG, timed up and go.

Table 22. Internal consistency outcomes for the topic “Disease Outcome Measures and Validation”

| **Author, year** | **Measure** | **n** | **Timepoint** | **Measure of internal consistency** | **Value** | **p-value** | **Notes** |
| --- | --- | --- | --- | --- | --- | --- | --- |
| FULL TEXT ARTICLES | | | | | | | |
| Eichinger 2018^65^ | FSHD-COM | 32 | --- | Cronbach’s alpha | 0.79 | --- |  |
| Mul 2021^69^ | FSHD-RODS | 230 | --- | Person separation index (PSI) | 0.97 | --- |  |

FSHD-COM, FSHD composite outcome measure; FSHD-RODS, FSHD Rasch-built disability scale

#### 7.4 Detailed tables for “Humanistic Burden and PROs”

Table 23. Population characteristics for the topic “Humanistic Burden and PROs”

| **Author, year** | **Measures reported** | **FSHD population type: n (%)** | **Age (years)** | **% pediatric** | **% female** | **BMI (kg/m^2^)** | **Age at onset (Years)** | **Disease duration (years)** | **CSS** | **D4Z4 pairs (kb)** |
| --- | --- | --- | --- | --- | --- | --- | --- | --- | --- | --- |
| CONFERENCE ABSTRACTS | | | | | | | | | | |
| Bergsma 2012^106^ | Abilhand  Brooke  CUE | FSHD NS: 76 (100%) | NR (13 – 74) | NS | --- | --- | --- | --- | --- | --- |
| Dahlqvist 2014^107^ | Low back pain rating scale | FSHD NS: 49 (100%) | --- | --- | --- | --- | --- | --- | --- | --- |
| Evangelista 2015^108^ | INQoL  MPQ | Type 1: 416 (100%) | --- | --- | --- | --- | --- | --- | --- | --- |
| Gazzotti 2011^109^ | SF-36 | FSHD NS: 3 (100%) | NR (16 – 58) | 33% | 100% | --- | --- | --- | --- | --- |
| Gergev 2013^110^ | Qualitative | FSHD NS: 4 (100%) | 15 (NR) | NS | 50% | --- | --- | --- | --- | --- |
| Heatwole 2012^111^ | Qualitative | FSHD NS: 328 (100%) | --- | 0% | --- | --- | --- | --- | --- | --- |
| Hodge 2016^112^ | Qualitative | FSHD NS: 53 (100%) | 23.1 ± 14.6 | NS | 59% | --- | --- | --- | --- | --- |
| Kools 2022^16^ | FSHD-HI^a^  FSHD-RODS^a^  PGIC | Type 1: 14 (100%) | 45.7 ±11.6 | 0% | --- | --- | --- | --- | 3.5 | --- |
| Leclair-Visonneau 2013^113^ | ESS^a^  FSS^a^  HADS^a^  PSQI | FSHD NS: 22 (100%) | --- | --- | --- | --- | --- | --- | --- | --- |
| Patel 2017^114^ | MPQ | FSHD NS: 77 (100%) | --- | --- | --- | --- | --- | --- | --- | --- |
| Shirilla 2019^115^ | Qualitative | FSHD NS: 390 (100%) | 52 (15 – 86) | NS | 55% | --- | 16^b^ | --- | --- | --- |
| Statland 2012^116^ | Qualitative | FSHD NS: 343 (100%) | 51 | --- | 51% | --- | NS: 22 | --- | --- | 25kb^b^ |
| Tawil 2022^17^ | FSHD-HI  PGIC | Type 1: 80 (100%) | 45.7 ±12.5 | 0% | --- | --- | --- | --- | 3.2 (2-4) | --- |
| Vissing 2017^117^ | INQoL | FSHD NS: 18 (100%) | --- | --- | --- | --- | --- | --- | --- | --- |
| Walker 2017^19^ | INQoL | Type 1: 8 (100%) | NR (16 – 20) | NS | --- | --- | --- | --- | --- | --- |
| FULL TEXTS | | | | | | | | | | |
| Andersen 2015^20^ | Fatigue VAS  Pain VAS  SF-36 | Type 1: 41 (100%) | ---^c^ | 0% | 48.8% | ---^c^ | --- | --- | ---^c^ | ---^c^ |
| Aprile 2013^118^ | LSQ^a^  Pain VAS^a^  SF-36^a^ | FSHD NS: 15 (100%) | 47.3 ±16.7 | 0% | 73.3% | --- | --- | --- | NR (2.5 – 4) | --- |
| Bachasson 2014^99^ | FSS  SF-36 | FSHD NS: 19 (100%) | 41 ±13 | 0 | 26.3% | 23.8 ±4.0 | --- | --- | --- | --- |
| Bakker 2017^119^ | Qualitative | FSHD NS: 25 (100%) | 56 (24-77) | 0% | 44% | --- | --- | --- | --- | --- |
| Bankolé 2016^21^ | FSS  SF-36 | Type 1: 19 (100%) | ---^c^ | 0% | ---^c^ | --- | --- | --- | --- | --- |
| Bayram 2020^120^ | SF-36 | FSHD NS: 37 (100%) | 39.1 ±11.5  (18 – 65) | 0% | 43.2% | 24.2 ±4.8 (16.8-37.3) | --- | 13.5 ±11.4  (1 – 40) | --- | --- |
| Bungener 1998^121^ | Abrams  EHD  Covi  HDRS  MADRS  PAS  SAS  Tyrer | FSHD NS: 11 (100%) | 33.2±8.3  (22-48) | 0% | 64% | --- | --- | --- | --- | --- |
| de Valle 2021^64^ | PedsQL NMM  FSHD-HI Peds | FSHD: 18 (100%); Type 1: 16 (95%); Type 2: 1 (6%); missing (1 (6%) | 13.3 ±3  (7 – 18) | 100% | 44% | 20.3 ±3.9 (15.1-30.2) | --- | --- | 2 ±1.2 (0.5-4.5) | Fragment size 16.9 (10 – 30) |
| Dijkstra 2021^122^ | Kidscreen  NeuroQoL Fatigue  Qualitative | Type 1: 28 (100%) | 10.8 ±4.6 (2 – 17) | 100% | 60% | --- | 6.9 ±5.1 (1 – 16) | --- | ---^a^ | Repeats 5.1 ±2.2 (2 -10) |
| Doix 2017^22^ | SF-36 | Type 1: 10 (100%) | 62.3±10.2 | 0% | 50% | --- | --- | --- | --- | Repeat units: 6.89 ±1.37 |
| Dos Santos 2021^100^ | FAS  FAS-cat  MMSE  MoCA  RAVLT  SF-36  TMT | FSHD NS: 13 (100%) | 49.5 ±13.2 | 0% | 69.2% | --- | 42.7 ±15.9 | 6.7 ±5.9 | --- | --- |
| Eren 2020b^34^ | INQoL | FSHD NS: 1 (100%) | 66 (NR) | 0% | 100% | --- | 21 (NR) | 45 | --- | --- |
| Gallais 2015^123^ | FAB  FSS  LARS  MADRS  MDE  MMSE  STAI  Stroop  TMT  Walton | FSHD NS: 19 (100%) | 46 ±13.5  (20 – 66) | 0% | --- | --- | --- | 22.4 ±12.5 | --- | --- |
| Goselink 2018^124^ | Kidscreen  NeuroQoL  Qualitative | Type 1: 31 (96.9%)  Type 2: 1 (3.1%) | 12 (4 – 17) | 100% | 56% | --- | 6.7 (NR) | --- | 2.5 ±1.5 (0 – 6) | Repeat arrays 5.2 ±2.0 (2- 10) |
| Guy-Coichard 2008^125^ | Brooke  Pain VAS  Qualitative  Vignos | FSHD NS: 121 (100%) | 45.7 ±14.8 | 0% | 47.9% | --- | --- | --- | --- | --- |
| Hamel 2019^126^ | Qualitative | FSHD NS: 328 (100%) | 54.5 ±13.4 (23-86) | 0% | 53.6% | --- | 23.1 ±15.4 | --- | --- | --- |
| Hatch 2021^79^ | NeuroQoL | Type 1: 21 (100%) | 41.6 (19 – 55) | 0% | 43% | 26.5  (17.9-33.3) | --- | --- | 2.7  (1 – 4) | --- |
| Hoffmann 2022^127^ | ESS  PSQI | FSHD: 690 (100%); Type 1: 352 (52.3%); Type 2: 50 (7.4%); unsure: 249 (37.0%) | --- | 1% | 50% | --- | --- | --- | --- | --- |
| Jacques 2019^128^ | CIS-severity  GSE  NEADL  SF-36  VAS pain | FSHD NS: 14 (100%) | 47.1 ±11.1 | 0% | 0% | 26.6 ±3.4 | --- | --- | --- | --- |
| Jensen 2008^129^ | BPI  Chronic pain scale | FSHD NS: 127 (100%) | 51.8 ±13.8 | 0% | 52% | --- | --- | --- | --- | --- |
| Johnson 2012^130^ | Qualitative | FSHD NS: 20 (100%) | 51 ±12.1 | 0% | 65% | --- | 30.5 ±15.5 | --- | --- | --- |
| Kalkman 2005^131^ | CIS  SF-36 | FSHD NS: 139 (100%) | 43.7 ±10.1  (22 – 61) | 0% | 51% | --- | --- | --- | --- | --- |
| Koopman 2017^132^ | CIS-Fatigue  ICQ-Acceptance  IES  IMQ  J-FCS  PCS  SES  SSI | Type 1: 24 (100%) | 51.3 ±13.4 (24-76) | 0% | 29% | --- | --- | 14.4 ±13.0 | 2.7 ±1.0 | --- |
| Loock 2022^47^ | Brooke  Pain VAS  Vignos | FSHD NS: 7 (100%) | 31.6 (26-44) | 0% | 57.1% | --- | --- | --- | --- | --- |
| Lue 2009^133^ | Barthel  Brooke  Vignose | FSHD NS: 20 (100%) | 35.5 ±16.7  (13 – 80) | NS | 40% | --- | --- | 17.3 ±14.5 | --- | --- |
| Minis 2010^134^ | CIS^e^  SF-36^e^ | FSHD NS: 139 (100%) | ---^c^ | 0% | ---^c^ | --- | --- | --- | --- | --- |
| Moris 2018^135^ | INQoL  MPQ  Universal pain assessment tool | Type 1: 398 (100%) | 47 (60.6)^b^ | --- | 50.4% | --- | 17 (21.2)^b^ | 26 (27)^b^ | --- | Total size: 25 kb (10 kb)^b^ |
| O’Dowd 2021^136^ | BDI  PSPP  RSS  SF-36  STAI | FSHD NS: 8 (100%) | 46.9 ±12.6 | 0% | --- | --- | --- | --- | --- | --- |
| Padua 2009^137^ | ID Pain  SF-36  VAS | FSHD NS: 65 (100%) | 40.9 ±13.5  (16 - 71) | NS | 52% | --- | --- | 17.2 ±11.3  (0 – 49) | 3 ±1.3  (0.5 -4.5) | Size: 21.1 kb (NR) |
| Payan 2009^138^ | Brooke  SF-36  Vignos | Placebo FSHD NS: 56 (100%) | 37.7 ±11.1  (18 – 60) | 0% | 39% | --- | --- | 10.6±7.5  (1 – 28) | --- | --- |
|  |  | Salbutamol FSHD NS: 56 (100%) | 42.3 ±10.7 (22-60) | 0% | 25% | --- | --- | 8.9±7.7 (1 – 42) | --- | --- |
| Ribot-Ciscar 2015^50^ | Pain VAS  SF-36 | FSHD NS: 8 (100%) | 58 ±11 | 0% | 75% | --- | --- | --- | --- | --- |
| Rose 2012^139^ | HAQ  HADS^a^  INQoL  IPQ-R  SF-36^a^ | FSHD NS: 49 (100%) | 52 ±17 | 0% | 59.2% | --- | --- | --- | --- | --- |
| Schipper 2017^140^ | Qualitative | FSHD NS 25 (100%) | NR (24 – 77) | 0% | 44% | --- | --- | NR (1-49) | --- | --- |
| Sezer 2022^141^ | Qualitative | FSHD NS: 16 (100%) | 47 (24 – 77) | 0% | 56% | --- | --- | --- | FCS: NR  (3 – 15) | --- |
| Statland  2022^38^ | FSHD-HI | Biceps brachii, placebo Type 1: 13 (92.9%); Type 2: 1 (7.1%) | 42.5 (21.65) | 0% | 21.4% | 23.5 (12.0 – 27.9) | --- | 20.5 (5 – 50) | --- | --- |
|  |  | Biceps brachii, ACE-083 Type 1: 14 (100%) | 47.5 (28 – 68) | 0% | 28.6% | 24.0 (19.1 – 35.0) | --- | 21.5 (4 – 42) | --- | --- |
|  |  | Tibialis anterior, placebo Type 1: 12 (85.7%); Type 2: 2 (14.3%) | 43.5 (18-62) | 0% | 50.0% | 23.7 (15.6 – 51.2) | --- | 19.5 (2 – 44) | --- | --- |
|  |  | Tibialis anterior, ACE-083 Type 1: 11 (84.6%); Type 2: 2 (15.4%) | 54.0 (31 – 70) | 0% | 53.8% | 23.8 (19.7 – 28.9) | --- | 24.0 (4 – 62) | --- | --- |
| van der Kooi 2007^143^ | BDI  CIS-Fatigue  DOA  DOF  DOP  MPQ  SCL  SIP | FSHD NS: 65 (100%) | 38 ±10 | 0% | 40% | --- | --- | --- | --- | --- |
| van de Geest-Buit 2023^142^ | CIS  FaCE  FSHD-RODS  HADS  SAAS  SF-36  SIAS | FSHD: 138 (100%); Type 1: 83 (60%); Type 2: 10 (7%); Type 1 & 2: 4 (3%) | 57 (20 – 85)^d^ | 0% | 52% | --- | --- | 16 (0 – 55)^d^ | --- | --- |
| Voet  2014^144^ | CIS-Fatigue  Nottingham sleep  Pain VAS  SIP | Usual care FSHD NS: 24 (100%) | 52 (20 – 79)^d^ | 0% | 29.2% | --- | --- | 16.7 (1 – 49)^d^ | 3  (1.5 – 4)^d^ | --- |
|  |  | Aerobic exercise training FSHD NS: 20 (100%) | 59 (21 – 68)^d^ | 0% | 40.0% | --- | --- | 13.0  (1.0 – 42.0)^d^ | 3  (0.5 – 4)^d^ | --- |
|  |  | Cognitive behavioral therapy FSHD NS: 13 (100%) | 49 (24 – 69)^d^ | 0% | 39.5% | --- | --- | 7.0  (0 – 40.0)^d^ | 2  (1 – 4)^d^ | --- |
| Winter 2010^145^ | EQ-5D  EQ-VAS  SF-36^f^ | FSHD NS: 17 (100%) | 43.8 ±9.8 | 0% | 64.7% | --- | 25 ±12 | 18.8 ±12.9 | FSHD stage I-III: 76.5% | --- |

Data are presented as mean ± SD or ranges, unless otherwise specified. ---, represents data not reported.

BDI, Beck’s Depression Inventory; BMI, body mass index; BPI, Brief Pain Inventory; CSS, clinical severity score; CUE, Capabilities of Upper Extremity; DOA, Daily Observed Activity; DOF, Daily Observed Fatigue; DOP, Daily Observed Pain; EHD, Depressive Mood Scale; ESS, Epworth Sleepiness Scale; EQ-5D, EuroQoL 5-dimension; EQ-VAS, EuroQoL-visual analogue scale; FAB, Frontal Assessment Battery; FaCE, Facial Clinimetric Evaluation Scale; FSHD, facioscapulohumeral dystrophy; FSHD-HI, FSHD Health Index; FSHD-HI Peds, FHSD-HI pediatric; FSHD-RODS, FSHD Rasch-built overall disability scale; FSS, Fatigue Severity Scale; GSE, General Self-efficacy Scale; HADS, Hospital Anxiety Depression Scale; HAQ, Health Assessment Questionnaires; HDRS, Hamilton Depression Rating Scale; ICQ, Illness Cognitions Questionnaire; IEQ, Impact of Event Scale; IMQ, Illness Management Questionnaire; INQoL, Individualized Neuromuscular Quality of Life; IPQ-R, Illness Perception Questionnaire-Revised; IQR, interquartile range; J-FCS, Jacobsen-fatigue catastrophizing scale; LARS, Lille Apathy Rating Scale; LSQ, Lumbar Spine Questionnaire; MADRS, Montgomery-Asberg Depression Rating Scale; MINI, Mini-International Neuropsychiatric Interview; MMSE, Mini Mental State Evaluation; MPQ, McGill Pain Questionnaire; NEADL, Nottingham Extended Activities of Daily Living; NeuroQoL, Quality of Life in Neurological Disorders; NHP, Nottingham Health Profile; NR, not reported; NS, not specified; PAS, Physical Anhedonia Scale; PCS, Pain Catastrophizing Scale; Peds QL NMM, Pediatric Quality of Life Neuromuscular Module; PGIC, Patient Global Impression of Change; PSPP, Physical Self-Perception Profile; PSQI, Pittsburgh Sleep Quality Index; RAVLT, Ray Auditory Verbal Learning Test; RSES, Rosenberg Self-Esteem Scale; SAAS, Social Appearance Anxiety Scale; SAS, Social Anhedonia Scale; SCL, Symptom Check List; SD, standard deviation; SES, Self-Efficacy Scale; SF-36, short-form 36; SIAS, Social Interaction Anxiety Scale; SIP, Sickness Impact Profile; SSI, Social Support Inventory; STAI, State-Trait Anxiety Inventory; TMT, Trail Making Test; VAS, visual analogue scale

^a^ Data not reported but use of an instrument mentioned in Methods and / or Results

^b^ Median with or without (IQR)

^c^Reported by subgroup (observational study) / treatment group (clinical trial) only.

^d^ Median (range)

^e^Results reported only by subgroup

^f^Results reported only in figure, not possible to extract

Table 24. Outcomes in studies using general QoL instruments, for the topic “Humanistic Burden and PROs”

| **Author, year** | **n** | **Instrument** | **Results** | **Comment** |
| --- | --- | --- | --- | --- |
| Andersen 2015^20^ | 41 | SF-36 | “Training improved role limitations due to physical health (p=0.04) and role limitations due to emotional problems.” |  |
| Aprile 2013^118^ | NR | SF-36 | Data reported only by subgroup of shoes with/without orthoses. |  |
| Bachasson 2014^99^ | 19 | SF-36 | - Role physical, mean (SD): 69 (37) - Physical functioning, mean (SD): 63 (28) - Bodily pain, mean (SD): 62 (26) - Role emotional, mean (SD): 80 (40) - Social functioning, mean (SD): 69 (31) - Mental health, mean (SD): 52 (23) - Vitality (energy/fatigue), mean (SD): 50 (23) - General health, mean (SD): 51 (18) |  |
| Bankolé 2016^21^ | 16 | SF-36 | Control group (n=8):   - T0, mean (SD): 65 (16) - T6, mean (SD): 70 (13) - T12, mean (SD): 67 (19) - T18, mean (SD): 65 (20) - T24, mean (SD): 60 (17)   Training group (n=8):   - T0, mean (SD): 60 (18) - T6, mean (SD): 60 (21) - T12, mean (SD): 65 (21) - T18, mean (SD): 64 (20) - T24, mean (SD): 69 (20) | SF-36 subscale/domain not detailed. |
| Bayram 2020^120^ | 37 | SF-36 | - Mental component score, mean (SD; range): 60.8 (20.1; 17 – 82) - Physical component score, mean (SD; range): 38.7 (15.3; 10 – 62) - Bodily pain, mean (SD; range): 77.6 (20.6; 32 – 100) - Mental health, mean (SD; range): 72 (19.7; 24 – 92) - Role emotional, mean (SD; range): 84.1 (26.3; 0 – 100) - Social functioning, mean (SD; range): 81.1 (18.2; 50 – 100) - Role physical, mean (SD; range): 18.4 (14.1; 0 – 50) - Physical functioning, mean (SD; range): 60.2 (15.2; 10 – 85) - Vitality, mean (SD; range): 60.8 (13.3; 20 – 80) - General health, mean (SD; range): 56.6 (12.3; 10 – 72) |  |
| Dijkstra 2021^122^ | 9 | Kidscreen | Total score:   - Baseline, mean (n=9): -0.9 - 2-year follow-up, mean (n=11): -0.7   Physical well-being:   - Baseline, mean (n=9): -1.5 - 2-year follow-up, mean (n=11): -2.0   Psychological well-being:   - Baseline, mean (n=9): -1.0 - 2-year follow-up, mean (n=11): -0.4   Autonomy:   - Baseline, mean (n=9): -0.7 - 2-year follow-up, mean (n=11): -0.4   Parent relation and home life:   - Baseline, mean (n=9): -0.8 - 2-year follow-up, mean (n=11): 0.2   Financial resources:   - Baseline, mean (n=9): -0.7 - 2-year follow-up, mean (n=11): -0.3   Social support and peers:   - Baseline, mean (n=9): -0.8 - 2-year follow-up, mean (n=11): -0.9   School environment:   - Baseline, mean (n=9): -0.7 - 2-year follow-up, mean (n=11): -0.1   Social acceptance:   - Baseline: insufficient number of replies - 2-year follow-up, mean (n=11): -1.6 | Children ≥8 completed the Kidscreen, otherwise, completed by parent proxy.  Parent relation/home life as well as school environment significantly different between baseline and follow-up.  Note that baseline scores are also reported in Goselink 2018^124^. |
| Doix  2017^22^ | 10 | SF-36 | Pre-training:   - Mental component score, mean (SD): 55.8 (3.7) - Physical component score, mean (SD): 34.9 (11.5) - Physical functioning, mean (SD): 43.0 (30.1) - Physical role, mean (SD): 70.0 (32.9) - Bodily pain, mean (SD): 57.5 (23.6) - Mental health, mean (SD): 75.6 (14.9) - Emotional role, mean (SD): 96.3 (11.1) - Social functioning, mean (SD): 75.0 (21.5) - Vitality, mean (SD): 48.9 (20.4) - General health, mean (SD): 59.9 (14.9) - Health change, mean (SD): 50.0 (28.9)   Post-training:   - Mental component score, mean (SD): 52.0 (10.4) - Physical component score, mean (SD): 34.8 (7.9) - Physical functioning, mean (SD): 35.0 (22.2) - Physical role, mean (SD): 71.9 (41.1) - Bodily pain, mean (SD): 52.6 (24.9) - Mental health, mean (SD): 73.0 (16.1) - Emotional role, mean (SD): 70.8 (45.2) - Social functioning, mean (SD): 75.0 (22.2) - Vitality, mean (SD): 47.3 (17.8) - General health, mean (SD): 50.4 (17.6) - Health change, mean (SD): 43.8 (17.7) |  |
| Dos Santos 2021^100^ | 13 | SF-36 | - Physical function, median (IQR; range): 30 (7.50 – 45; 0 - 85) - Role physical, median (IQR): 0 (0 – 62.50; 0 - 100) - Bodily pain, median (IQR): 21 (15 – 46; 0 - 90) - General health, median (IQR): 27 (26 – 48.5; 15 - 95) - Vitality, median (IQR): 60 (25 – 65; 0 - 85) - Social function, median (IQR): 50 (31.25-75; 25 - 100) - Role emotional, median (IQR): 33.5 (0 – 83.25; 0 - 100) - Mental health, median (IQR): 76 (42 – 89; 16 – 96) |  |
| Gazzotti 2011^109^ | 3 | SF-36 | “SF-36 score did not change during the evaluated (3-month) period.” |  |
| Jacques 2019^128^ | 14 | SF-36 | - Mental composite score, mean (SD): 49.8 (11.0) - Physical composite score, mean (SD): 30.8 (6.3) - Physical functioning, mean (SD): 18.2 (12.5) - Role physical, mean (SD): 41.5 (26.7) - Bodily pain, mean (SD): 48.1 (28.6) - General health, mean (SD): 44.1 (24.2) - Vitality, mean (SD): 37.9 (16.9) - Social functioning, mean (SD): 67.9 (29.7) - Role emotional, mean (SD): 69.0 (24.1) - Mental health, mean (SD): 65.0 (19.1) |  |
| Kalkman 2005^131^ | 139 | SF-36 | - Physical functioning, mean (SD): 45.2 (31.4) - Social functioning, mean (SD): 71.6 (24.2) - Role limitations, physical, mean (SD): 47.9 (42.0) - Role limitations, emotional, mean (SD): 69.5 (41.6) - Mental health, mean (SD): 72.6 (17.0) - Bodily pain, mean (SD): 66.6 (23.8) - General health, mean (SD): 51.7 (21.6) |  |
| Minis 2010^134^ | 1349 | SF-36 | Data only reported by subgroups for employed / not employed |  |
| O’Dowd 2021^136^ | NR | SF-36 | - Mental composite score, mean (SD): 51 (13) - Physical composite score, mean (SD): 37 (5) - Physical function, mean (SD): 26 (21) - Role physical, mean (SD): 61 (20) - Bodily pain, mean (SD): 52 (29) - General health, mean (SD): 56 (25) - Vitality, mean (SD): 51 (21) - Social function, mean (SD): 67 (30) - Role emotional, mean (SD): 73 (28) - Mental health, mean (SD): 71 (23) |  |
| Padua 2009^137^ | 65 | SF-36 | - Mental composite score: 47.4 - Physical composite score: 38.3 - Physical function, mean (Italian norm): 48.2 (84.5) - Role-physical, mean (Italian norm): 46.1 (78.2) - Bodily pain, mean (Italian norm): 63.2 (73.7) - General health, mean (Italian norm): 52.2 (65.2) - Vitality, mean (Italian norm): 55.9 (61.9) - Social function, mean (Italian norm): 69.0 (77.4) - Role-emotional, mean (Italian norm): 63.0 (76.2) - Mental health, mean (Italian norm): 64.1 (66.6) | Subgroup data available by sex, age, CSS, and fragment size |
| Payan  2009^138^ | NR | SF-36 | “Quality of life did not show any significant differences between the treated and non-treated groups. The 8 dimensions of the SF-36 were unchanged between baseline and month 6 for both groups, but with great variability.” |  |
| Ribot-Ciscar 2015^50^ | 8 | SF-36 | Pre-treatment:   - Global score, mean (SD): 43 (7.4) - Emotional well-being, mean (SD): 53 (10) - Self-rated health state, mean (SD): 53 (8.3)   Post-treatment:   - Global score, mean (SD): 54 (6.7) - Emotional well-being, mean (SD): 66 (6) - Self-rated health state, mean (SD): 72 (8) | Scales written as identified in manuscript. Emotional well-being and self-rated health state significantly different from pre- to post-treatment (p=0.05 and p=0.04, respectively). |
| Rose 2012^139^ | NR | SF-36 | “All SF-36 domains were negatively affected, but those of physical functioning, role physical and general health were significantly lower than what is seen in a normal population (data not shown).” |  |
| van der Kooi 2007^143^ | 65 | SIP | Total score in overall population, baseline:   - Mean (SD): 708 (612)   Placebo, no training group (n=16):   - Baseline, mean (range): 691 (380 – 1001) - Week 52, mean (range): 565 (227 – 903)   Placebo, training group (n=19):   - Baseline, mean (range): 613 (328 – 898) - Week 52, mean (range); 530 (220 – 840)   Albuterol, no training group (n=15)   - Baseline, mean (range): 723 (402 – 1043) - Week 52, mean (range): 599 (247 – 951)   Albuterol, training group (n=15):   - Baseline, mean (range): 834 (513 – 1154) - Week 52, mean (range): 790 (441 – 1139) | Subgroup data available at baseline by pain / no pain, not severely fatigued / fatigued.  Follow-up scores after intervention by albuterol and/or strength training.  No significant differences between training versus no training / albuterol versus placebo. |
| van de Geest-Buit 2023^142^ | 138 | SF-36 | Pain:   - Mean (SD; range): 67.8 (21.5; 20 – 100) - % scoring most negative 10% (0 – 10): 0% - % scoring most positive 10% (90 – 100): 23.2%   Emotional well-being:   - Mean (SD; range): 76.7 (15.3; 24 – 100) - % scoring most negative 10% (0 – 10): 0% - % scoring most positive 10% (90 – 100): 17.4%   Emotional role limitations:   - Mean (SD; range): 75.4 (37.8; 0 – 100 - % scoring most negative 10% (0 – 10): 13.8% - % scoring most positive 10% (90 – 100): 66.7% | Subgroup data available by facial weakness |
| Voet  2014^144^ | 57 | SIP | Period 1  Usual care (n=24):   - Pretreatment, median (range): 130 (0 – 387) - 16 week, median (range): 140 (0 -372) - 28 week, median (range): 169 (0-572)   Aerobic exercise training:   - Pretreatment (n=20), median (range): 187 (0 – 572) - 16 week (n=19), median (range): 144 (0-572) - 28 week (n=19), median (range): 152 (0 – 523)   Cognitive behavioral therapy (n=13):   - Pretreatment, median (range): 152 (0 – 476) - 16 week, median (range): 149 (0 – 387) - 28 week, median (range): 100 (0 – 387)   Period 2  Aerobic exercise training (n=8):   - Pretreatment, median (range): 149 (0 – 387) - 16 week, median (range):112 (0 - 344) - 28 week, median (range): 137 (43 - 386)   Cognitive behavioral therapy (n=12):   - Pretreatment, median (range): 215 (0 - 572) - 16 week, median (range): 144 (0 - 420) - 28 week, median (range): 109 (0 – 294) | Significant differences were observed in aerobic exercise training and cognitive behavioral therapy versus usual care. |
| Winter 2010^145^ | 17 | SF-36 | - Mental composite score, mean (SD; median): 55.5 (10.5; 55.1) - Physical composite score, mean (SD; median): 35.0 (9.5; 34.9) | Individual domains presented graphically |
|  | 17 | EQ-5D | Overall:   - Mean (SD; median): 0.75 (0.21; 0.81)   Proportion with moderate or severe problems across dimensions:   - Mobility: 70.6% - Self-care: 52.9% - Usual activities: 76.5% - Pain/discomfort: 70.6% - Anxiety/depression: 41.2% | Proportion of respondents noting moderate or severe problems in at least one dimension was 82.4% (n=14) |
|  | 17 | EQ VAS | Mean (SD; median): 57.1 (19.2; 50.0) |  |

EQ-5D, EuroQoL 5-dimension; EQ-VAS, EuroQoL-visual analogue scale; HAQ, Health Assessment Questionnaire; NR, not reported; PGIC, Patient Global Impression of Change; QOL, quality of life; SD, standard deviation; SF-36, short-form 36; SIP, Sickness Impact Profile; T, time in weeks from baseline

Table 25. Outcomes in studies using disease-specific QoL instruments, for the topic “Humanistic Burden and PROs”

| **Author, year** | **n** | **Instrument** | **Results** | **Comment** |
| --- | --- | --- | --- | --- |
| de Valle 2021^64^ | 18 | FSHD-HI Peds | - Mean (SD; range): 21.9 (22.5; 0.2 – 67.5) |  |
|  | 18 | PedsQL NMM | Child assessment:   - Mean (SD; range): 73.9 (15.7; 47 – 95)   Parent proxy:   - Mean (SD; range): 73.5 (17.1; 42 – 97) |  |
| Eren 2020b^34^ | 1 | INQoL | Pre-operative:   - Weakness: 100 - Pain: 94.74 - Fatigue: 94.74 - Muscle locking: 0 - Droopy eyelids: 47.37 - Double visions: 0 - Swallowing difficulties: 63.16 - Activities: 76.85 - Independence: 97.22 - Social relationship: 44.44 - Emotions: 19.44 - Body image: 88.88 - Treatment (expected): 83.33 - Quality of life: 55.55   Post-operative:   - Weakness: 94.74 - Pain: 0 - Fatigue: 52.63 - Muscle locking: 0 - Droopy eyelids: 0 - Double visions: 0 - Swallowing difficulties: 0 - Activities: 57.41 - Independence: 100 - Social relationship: 0 - Emotions: 0 - Body image: 61.11 - Treatment (perceived): 100 - Quality of life: 40 |  |
| Evangelista 2015 ^108^ | 354 | INQoL | - Overall, mean: 50.87 - Muscle weakness, mean: 66.32 - Fatigue, mean: 46.89 - Activities, mean: 57.07 - Body image, mean: 55.21 |  |
| Hatch 2021^79^ | NR | NeuroQoL | Instrument cited in Methods, but no data presented. |  |
| Kools 2022^16^ | NR | FSHD-HI | Instrument cited in Methods, but no data presented. |  |
| Moris 2018^135^ | 340 | INQoL | - QoL, median (IQR): 53.1 (34.3) - Muscle weakness, median (IQR): 63.2 (26.4) - Locking, median (IQR): 10.5 (22.4) - Pain, median (IQR): 44.7 (31.6) - Fatigue, median (IQR): 50.0 (34.2) - Activities, median (IQR): 57.4 (58.6) - Independence, median (IQR): 50.0 (64.6) - Relationship, median (IQR): 20.4 (16.7) - Feelings, median (IQR): 54.2 (56.2) - Body image, median (IQR): 59.7 (51.4) | Subgroup data available by gender and presence of chronic pain |
| Rose 2012^139^ | 49 | INQoL | - Weakness, mean (SD): 59.9 (28.1) - Locking, mean (SD): 39.0 (21.8) - Pain, mean (SD): 40.0 (23.0) - Fatigue, mean (SD): 52.4 (22.9) - Activities, mean (SD): 56.1 (25.3) - Independence, mean (SD): 42.3 (27.7) - Social, mean (SD): 28.8 (23.2) - Emotional, mean (SD): 41.9 (26.5) - Body image, mean (SD): 59.0 (28.4) |  |
| Statland  2022^38^ | 55 | FSHD-HI | Biceps brachii  Placebo (n=14):   - Absolute change in total score, LS mean (SEM): 2.3 (2.41) - Absolute change in arm/shoulder score, LS mean (SEM): 0.92 (3.83) - Absolute change in activity score, LS mean (SEM): -1.6 (3.52)   ACE-083 (n=14):   - Absolute change in total score, LS mean (SEM): 2.1 (2.61) - Absolute change in arm/shoulder score, LS mean (SEM): -3.8 (4.05) - Absolute change in activity score, LS mean (SEM): -2.0 (3.62)   Tibialis anterior  Placebo (n=14):   - Absolute change in total score, LS mean (SEM): 2.5 (2.35) - Absolute change in arm/shoulder score, LS mean (SEM): 0.06 (2.94) - Absolute change in activity score, LS mean (SEM): 1.2 (3.45)   ACE-083 (n=13):   - Absolute change in total score, LS mean (SEM): 0.53 (2.53) - Absolute change in arm/shoulder score, LS mean (SEM): -0.91 (3.20) - Absolute change in activity score, LS mean (SEM): 3.3 (3.70) | No statistically significant differences between treatment groups in LS mean change from baseline to day 190 in total score. |
| Tawil 2022^17^ | 80 | FSHD-HI | “No differences were observed.” |  |
| Vissing 2017^117^ | 18 | INQoL | “No consistent changes over time were observed for INQoL scores.” |  |
| Walker 2017^19^ | 8 | INQoL | “Mean overall INQoL score did not markedly change.” |  |

FSHD-HI, Facioscapulohumeral Dystrophy Health Index; FSHD-HI Peds, FSHD-HI pediatric; INQoL, Individualized Neuromuscular Quality of Life; IQR, interquartile range; LS, least squares; NeuroQoL, Quality of Life in Neurological Disorders; Peds QL NMM, Pediatric Quality of Life Neuromuscular Module; QoL, quality of life; SD, standard deviation; SEM, standard error of the mean

Table 26. Outcomes in studies using instruments assessing depression and/or anxiety, for the topic “Humanistic Burden and PROs”

| **Author, year** | **n** | **Instrument** | **Results** | **Comment** |
| --- | --- | --- | --- | --- |
| Bungener 1998^121^ | 11 | Covi anxiety | - Mean (SD): 2.5 (3.1) |  |
|  | 11 | EHD | - Emotional deficit, mean (SD): 2.5 (3.4) - Loss of control, mean (SD): 5.9 (6.5) - Irritability, mean (SD): 4.8 (5.0) - Anhedonia, mean (SD): 1.4 (2.3) - Expressiveness, mean (SD): 0.1 (2.5) - Sadness, mean (SD): 1.9 (2.2) - Anxious hyperaesthesia, mean (SD): 0.8 (1.7) |  |
|  | 11 | HDRS | - Mean (SD): 7.2 (7.1) |  |
|  | 11 | MADRS | - Mean (SD): 8.3 (8.4) |  |
|  | 11 | Tyrer anxiety | - Mean (SD): 9.4 (8.9) |  |
| Gallais 2015^123^ | 19 | MADRS | - Mean (SD): 7.0 (7.4) |  |
|  | 19 | STAI | - STAI-A T score, mean (SD): 44.8 (11.0) - STAI-B T score, mean (SD): 48.7 (12.9) |  |
|  | 19 | MINI | - Of the 19 patients, 1 (5.3%) met the criteria for a current major depressive order |  |
| Leclair-Visonneau 2013^113^ | NR | HADS | Instrument cited in Methods, but no data presented. |  |
| O’Dowd 2021^136^ | NR | BDI | - Mean (SD): 10.9 (7.9) |  |
|  | NR | STAI | - Mean (SD): 38.6 (11.6) |  |
| Padua 2009^137^ | NR | BDI | Instrument cited in Methods, but no data presented. |  |
| Rose 2012^139^ | NR | HADS | Instrument cited in Methods, but no data presented. |  |
| van de Geest-Buit 2023^142^ | 138 | HADS | - Total score, mean (SD; range): 9.5 (6.7; 0 – 32) - Total score, % scoring above cut-off score ≥11: 41.3% - Depression, mean (SD; range): 4.8 (3.6; 0 – 16) - Depression, % scoring above cut-off score ≥8: 22.5% - Anxiety, mean (SD; range): 4.8 (3.8; 0 – 20) - Anxiety, % scoring above cut-off score ≥8: 21.7% | Subgroup data available by facial weakness. |
|  | 138 | SAAS | - Fear of negative evaluation, mean (SD; range): 24.3 (9.2; 16-73) - Fear of negative evaluation, % scoring most negative 10% (73 – 80): 0.7% - Fear of negative evaluation, % scoring most positive 10% (16 – 22): 57.2% |  |
|  | 138 | SIAS | - Social interaction anxiety, mean (SD; range): 33.1 (9.1; 20 – 71( - Social interaction anxiety, % scoring most negative 10% (93 – 100): 0% - Social interaction anxiety, % scoring most positive 10% (20 – 27): 29.7% |  |
| van der Kooi 2007^143^ | 65 | BDI short form | Overall population, baseline:   - Mean (SD): 1.7 (3.0) - % scoring ≥4: 16%   Placebo, no training (n=16):   - Baseline, mean (range): 1.9 (0.4-3.4) - Week 52, mean (range): 1.5 (0.5 – 2.5)   Placebo, training (n=19):   - Baseline, mean (range): 0.7 (-0.7 – 2.1) - Week 52, mean (range): 0.7 (-0.2 – 1.6)   Albuterol, no training (n=15):   - Baseline, mean (range): 2.2 (0.7 – 3.7) - Week 52, mean (range): 1.1 (0.1 – 2.1)   Albuterol, training (n=15):   - Baseline, mean (range): 2.4 (0.8 – 3.9) - Week 52, mean (range): 1.9 (0.9 – 2.9) | No significant differences observed between groups.  Baseline subgroup data available by pain / no pain, not severely fatigued / fatigued. |

BDI, Beck’s Depression Inventory; EHD, Depressive Mood Scale; HADS, Hospital Anxiety Depression Scale; HDRS, Hamilton Depression Rating Scale; MADRS, Mongtomery-Asberg Depression Rating Scale; MINI, Mini-International Neuropsychiatric Interview; NR, not reported; SAAS, Social Appearance Anxiety Scale; SIAS, Social Interaction Anxiety Scale; STAI, State-Trait Anxiety Inventory

Table 27. Outcomes in studies using instruments assessing pain, for the topic “Humanistic Burden and PROs”

| **Author, year** | **n** | **Instrument** | **Results** | **Comment** |
| --- | --- | --- | --- | --- |
| Andersen 2015^20^ | 28 | VAS | Protein (n=9):   - Baseline, mean (range): 0.7 (0 – 3) - End, mean (range): 1.2 (0 – 5)   Placebo (n=10):   - Baseline, mean (range): 1.3 (0 – 5) - End, mean (range): 0.3 (0 – 1)   Control (n=9)   - Baseline, mean (range): 1.7 (0 – 5) - End, mean (range): 1.9 (0 – 5) |  |
| Aprile 2013^118^ | NR | VAS | Data reported only by subgroup of shoes with/without orthoses. |  |
| Dahlqvist 2014^107^ | 39 | LBPR | Prevalence of lower back pain was significantly higher in FSHD patients than in controls (p<0.0001). |  |
| Evangelista 2015 ^108^ | 379 | MPQ | Persistent pain was reported by 50% and recurrent localised pain by 43%. Patients with persistent pain described it as discomforting and distressing in 19.7% and 14.4% of cases, respectively. Localised pain was described as discomforting (16%). The shoulder was the most affected location for both persistent (40.4%) and localised pain (23.6%). The population with localised pain reported no change in muscle bulk or weakness after the painful episodes. |  |
| Guy-Coichard 2008^125^ | 91 | VAS | - Usual daytime pain intensity, mean (SD): 5.0 (2.5) - Usual nocturnal pain intensity, mean (SD): 3.3 (2.8) - Worst pain intensity, mean (SD): 6.5 (2.4) - % of patients reporting severe (7 or greater) usual pain: 30% - % of patients reporting moderate (5-6) usual pain: 21% | The impact of pain was assessed using a PRO, however, the questionnaire was not specified. |
|  | NR | NS | Impact of pain on:   - Global score, mean (SD): 4.8 (2.3) - Ability to move, mean: 4.6 - Mood, mean: 4.8 - Sleep, mean: 3.4 - Leisure activities, mean: 6.0 - Relationship with others, mean: 3.4 - Occupational home activities, mean: 6.0 - Activities in general, mean: 5.7 |  |
| Jacques 2019^128^ | 14 | VAS | - Mean (SD): 3.9 (2.2) |  |
| Jensen 2008^129^ | 127 | Grading of chronic pain scale | - Subjects reporting pain, n (%) 104 (82%) - Mean (SD) pain for subjects with pain: 4.4 (2.4) - Subjects reporting severe pain (7 or greater): 24 (23%) - Duration of pain, mean (SD) years: 13.4 (10.4) |  |
|  | 104 | BPI | - Total score, mean (SD): 2.74 (2.23) - General activity, mean (SD): 2.89 (2.67) - Mood, mean (SD): 3.17 (2.70) - Mobility, mean (SD): 3.65 (2.94) - Normal work (including housework), mean (SD): 3.44 (2.86) - Relations with other people, mean (SD): 2.02 (2.31) - Sleep, mean (SD): 3.50 (2.95) - Enjoyment of life, mean (SD): 3.53 (3.29) - Self-care, mean (SD): 2.28 (2.63) - Recreational activities, mean (SD): 3.57 (3.32) - Social activities, mean (SD): 2.76 (2.89) - Communication with others, mean (SD): 1.15 (2.00) - Learning new skills, mean (SD): 1.14 (2.02) |  |
| Kools 2022^16^ | NR | PGIC | Over 80% reported improvement or no change over a 56-week period of receiving losmapimod. |  |
| Loock  2022^47^ | 7 | VAS | Pre-operative:   - Mean (SD; range): 3 (2; 0 – 7)   Post-operative:   - Mean (SD; range): 1 (1; 0- 4) | Pre- and post-operative pain scores were significantly different (p=0.008). |
| Moris 2018^135^ | 383 | Short form MPQ | - Presence of pain total, n (%): 339 (85.2) - Total score, median (IQR): 6.0 (12.5) - Sensory score, median (IQR): 5.0 (9.0) - Affective score, median (IQR): 1.0 (3.0) - Present pain intensity index, median (IQR): 1.0 (2.0) | Subgroup data available by age, disease duration, fragment size and motor function. |
|  | 367 | Universal pain assessment tool | - Total chronic pain, n (%): 203 (55.6) - Severe chronic pain, n (%): 69 (30.4) |  |
| Padua 2009^137^ | 65 | VAS | - Mean (SD; range): 31.6 (25.2; 0 – 76) - % reporting at least mild/moderate pain: 56% |  |
|  | 65 | ID Pain | - Very probable neuropathic pain: 6.7% - Probable neuropathic pain: 20% - Possible neuropathic pain: 24.4% - Improbable pain: 33.3% - Joint pain exclusively: 15.6% |  |
| Patel 2017^114^ | NR | MPQ | - Pain was reported by 69% of patients |  |
| Ribot-Ciscar 2015^50^ | 2 | VAS | Subject 1:   - Pre-treatment: 9 (out of 15) - Post-treatment: 0   Subject 2:   - Pre-treatment: 14 (out of 15) - Post-treatment: “remained in pain” |  |
| Tawil 2022^17^ | 80 | PGIC | “There was significant improvement compared to placebo (difference -0.58 on 1-7 Likert scale, p=0.02).” |  |
| van der Kooi 2007^143^ | 65 | MPQ VAS | Overall population, baseline:   - Mean (SD) mm: 12.9 (17.1)   Placebo, no training (n=16):   - Baseline mean (range) mm: 13.4 (4.9 – 22.0) - Week 52 mean (range) mm: 9.0 (0.4 – 17.6)   Placebo, training (n=19)   - Baseline mean (range) mm: 9.7 (1.9 – 17.6) - Week 52 mean (range) mm: 12.1 (4.2 – 20.0)   Albuterol, no training (n=15)   - Baseline mean (range) mm: 10.5 (1.6 – 19.3) - Week 52 mean (range) mm: 12.0 (3.1 – 20.9)   Albuterol, training (n=15)   - Baseline mean (range) mm: 18.5 (9.7 – 27.4) - Week 52 mean (range) mm: 17.9 (9.0 – 26.8) | No significant differences observed between groups.  Baseline subgroup data available by pain / no pain, not severely fatigued / fatigued. |
|  | 65 | DOP | Overall population, baseline:   - Mean (SD): 2.4 (2.2)   Placebo, no training (n=16):   - Baseline mean (range): 1.9 (0.7 – 3.0) - Week 52 mean (range): 1.9 (0.8 – 3.0)   Placebo, training (n=19)   - Baseline mean (range): 2.1 (1.0 – 3.1) - Week 52 mean (range): 1.9 (0.9 – 2.9)   Albuterol, no training (n=15)   - Baseline mean (range): 2.6 (1.4 – 3.8) - Week 52 mean (range): 2.5 (1.4 – 3.6)   Albuterol, training (n=15)   - Baseline mean (range): 3.2 (2.0 – 4.4) - Week 52 mean (range): 1.9 (0.8 – 3.1) |  |
| Voet  2014^144^ | 57 | VAS | Period 1  Usual care (n=24):   - Pretreatment, median (range): 18.5 (0-59) - 16 week, median (range): 21 (0-65) - 28 week, median (range): 21 (0-69)   Aerobic exercise training:   - Pretreatment (n=20), median (range): 19.5 (4-65) - 16 week (n=19), median (range): 20 (2-65) - 28 week (n=19), median (range): 16 (0-71)   Cognitive behavioral therapy (n=13):   - Pretreatment, median (range): 15 (0-84) - 16 week, median (range): 10 (0-72) - 28 week, median (range): 10 (0-80)   Period 2  Aerobic exercise training (n=8):   - Pretreatment, median (range): 13 (0-47) - 16 week, median (range): 10 (0-72) - 28 week, median (range): 10 (0-80)   Cognitive behavioral therapy (n=12):   - Pretreatment, median (range): 20 (0-56) - 16 week, median (range): 20 (0-70) - 28 week, median (range): 20 (0-69) | No differences observed between aerobic exercise or cognitive behavioral therapy and usual care. |

BPI, Brief Pain Inventory; DOP, Daily Observed Pain; IQR, interquartile range; LBPR, Low Back Pain Rating scale; MPQ, McGill Pain Questionnaire; NR, not reported; NS, not specified; PGIC, Patient’s Global Impression of Change; VAS, visual analogue scale.

Table 28. Outcomes in studies using instruments assessing sleep, for the topic “Humanistic Burden and PRO”

| **Author, year** | **n** | **Instrument** | **Results** | **Comments** |
| --- | --- | --- | --- | --- |
| Hoffmann 2022^127^ | 592 | PSQI | - Mean (SD): 7.81 (4.12) - % scoring >5: 66.2% (n=392); 95% CI: 62.4 – 70.0 |  |
|  | 592 | ESS | - Mean (SD): 5.85 (4.07) - % scoring >10: 15.0% (n=89); 95% CI: 12.2 – 17.9 |  |
| Leclair-Visonneau 2013^113^ | 22 | PSQI | FSHD population had significantly worse sleep quality than the control group (p<0.01), with loss of subjective sleep quality, nocturnal pain, bad dreams and day dysfunction due to sleepiness. |  |
|  | NR | ESS | Instrument cited in Methods, but no data presented. |  |
| Voet  2014^144^ | 57 | NHP | Period 1  Usual care (n=24):   - Pretreatment, median (range): 0 (0 – 83.9) - 16 week, median (range): 12.57 (0 – 77.63) - 28 week, median (range): 12.57 (0 – 77.63)   Aerobic exercise training:   - Pretreatment (n=20), median (range): 6.29 (0 – 77.63) - 16 week (n=19), median (range): 0 (0 – 87.43) - 28 week (n=19), median (range): 12.57 (0 – 72.74)   Cognitive behavioral therapy (n=13):   - Pretreatment, median (range): 12.57 (0 – 61.53) - 16 week, median (range): 0 (0 – 61.53) - 28 week, median (range): 0 (0 – 61.53)   Period 2  Aerobic exercise training (n=8):   - Pretreatment, median (range): 0 (0 – 34.27) - 16 week, median (range): 0 (0 – 21.7) - 28 week, median (range): 12.57 (0 – 21.7)   Cognitive behavioral therapy (n=12):   - Pretreatment, median (range): 12.57 (0 – 77.63) - 16 week, median (range): 0 (0 – 34.27) - 28 week, median (range): 0 (0 – 21.7) | Significant differences were observed at 16-weeks only between aerobic exercise or cognitive behavioral therapy versus usual care. |

CI, confidence interval; ESS, Epworth Sleepiness Scale; NHP, Nottingham Health Profile; PSQI, Pittsburgh Sleep Quality Index; SD, standard deviation

Table 29. Outcomes in studies using instruments assessing activities of daily living, for the topic “Humanistic Burden and PROs”

| **Author, year** | **n** | **Instrument** | **Results** | **Comment** |
| --- | --- | --- | --- | --- |
| Jacques 2019^128^ | 14 | NEADL | - Mean (SD): 39.4 (14.6) |  |
| van der Kooi 2007^143^ | 65 | DOA* | - Mean (SD): 6.1 (2.0) | Subgroup data available by pain / no pain and by not severely fatigued / severely fatigued. |

*No further information available on how this was derived; a higher DOA score indicates more self-reported activity

DOA, Daily Observed Activity; NEADL, Nottingham Extended Activities of Daily Living; NS, not specified.

Table 30. Outcomes in studies using instruments assessing disability/functional status, for the topic “Humanistic Burden and PROs”

| **Author, year** | **n** | **Instrument** | **Results** | **Comment** |
| --- | --- | --- | --- | --- |
| Bergsma 2012^106^ | 76 | BUE | - % patients scoring 2 – 4: 88% |  |
|  | 76 | CUE | - Total score, median (range): 155 (53 – 210)   - Corresponds to 73.8% of maximum score |  |
|  |  | Abilhand | - Total score, median (range): 38 (13 – 44)   - Corresponds to 86.4% of maximum score |  |
| Gallais 2015^123^ | 19 | WFS | - Mean (SD): 3.16 (2.1) |  |
| Guy-Coichard 2008^125^ | NR | BUE | Instrument cited in Methods, but no data presented. |  |
|  | NR | VLE | Instrument cited in Methods, but no data presented. |  |
| Kools 2022^16^ | NR | FSHD-RODS | Instrument cited in Methods, but no data presented. |  |
| Loock  2022^47^ | 7 | BUE | Post-operative:   - Score (n): 3 (5), 2 (2) |  |
|  |  | VLE | Post-operative:   - Score (n): 1 (2), 2 (3), 3 (1), 7 (1) |  |
| Lue 2009^133^ | 20 | BUE | - Mean (SD; range): 2.3 (0.8; 1 – 3) |  |
|  | 20 | VLE | - Mean (SD; range): 2.2 (1.3; 1 – 5) |  |
|  | 20 | Barthel index | - Mean (SD; range): 97.8 (4.7; 85 – 100) |  |
| Payan  2009^138^ | 109 | BUE | Placebo (n=55):   - Baseline, mean (SD): 2.6 (0.5) - Month 6, mean change (SD): 0.0 (0.4)   Salbutamol (n=54):   - Baseline, mean (SD): 2.6 (0.7) - Month 6, mean change (SD): 0.1 (0.4) | No significant differences between placebo and salbutamol in mean change at Month 6. |
|  | 109 | VLE | Placebo (n=55):   - Baseline, mean (SD): 1.7 (1.0) - Month 6, mean change (SD): 0.1 (0.6)   Salbutamol (n=54):   - Baseline, mean (SD): 1.7 (1.1) - Month 6, mean change (SD): 0.1 (0.5) | No significant differences between placebo and salbutamol in mean change at Month 6. |
| Rose 2012^139^ | 49 | HAQ | - Dress, mean (SD): 1.1 (0.9) - Eat, mean (SD): 1.3 (0.9) - Arise, mean (SD): 0.7 (0.8) - Walk, mean (SD): 1.5 (1.1) - Hygiene, mean (SD): 1.4 (1.2) - Reach, mean (SD): 1.9 (1.0) - Grip, mean (SD): 0.5 (0.8) - Activity, mean (SD): 1.6 (1.0) - Total score, mean (SD): 1.2 (0.9) |  |
| van de Geest-Buit 2023^142^ | NA | FSHD-RODS | Instrument cited in Methods, but no data presented. |  |
|  | 138 | FaCE | - Social functioning, mean (SD; range): 94.3 (10.8; 50 – 100) - Social functioning, % scoring most negative 10% (0 – 10): 0% - Social functioning, %scoring most positive 10% (90 – 100): 78.2% | Subgroup data available by facial weakness |

BUE, Brooke Upper Extremity; CUE, Capabilities of Upper Extremity; FaCE, Facial Clinimetric Evaluation Scale; FSHD-RODS, Facioscapulohumeral Dystrophy Rasch-built overall disability scale; NA, not applicable; NR, not reported; VLE, Vignos Lower Extremity; WFS, Walton Functional Scale

Table 31. Outcomes in studies using instruments to assess fatigue, for the topic “Humanistic Burden and PROs”

| **Author, year** | **n** | **Instrument** | **Results** | **Comment** |
| --- | --- | --- | --- | --- |
| Andersen 2015^20^ | 28 | VAS | Protein (n=9):   - Baseline, mean (SD): 3.4 (1-6) - End, mean (SD): 3.4 (1-7)   Placebo (n=10):   - Baseline, mean (SD): 3.1 (0 – 7) - End, mean (SD): 1.6 (0 – 3)   Control (n=9):   - Baseline, mean (SD): 3.2 (0 – 6) - End, mean (SD): 3.2 (0 – 6) |  |
| Bachasson 2014^99^ | 19 | FSS | - Mean (SD): 38 (12) - Severe fatigue experienced by 11 (57.9%) patients |  |
| Bankolé 2016^21^ | 16 | FSS | Control group (n=8):   - T0, mean (SD): 35 (9) - T6, mean (SD): 34 (15) - T12, mean (SD): 32 (10) - T18, mean (SD): 37 (12) - T24, mean (SD): 40 (11)   Training group (n=8):   - T0, mean (SD): 44 (8) - T6, mean (SD): 37 (13) - T12, mean (SD): 36 (15) - T18, mean (SD): 35 (14) - T124, mean (SD): 34 (15) |  |
| Dijkstra 2021^122^ | 10 / 11 | NeuroQoL Fatigue | Baseline (n=10)   - Mean SD 1.1   2-year follow-up (n=11)   - Mean SD 1.0 |  |
| Gallais 2015^123^ | 19 | FSS | - Mean (SD): 4.8 (1.5) |  |
| Goselink 2018^124^ | 10 | NeuroQoL Fatigue | Mean SD was:   - Child: 1.1 SD above average (compared to healthy subjects) - Parent: 1.2 SD above average (compared to healthy subjects) |  |
| Jacques 2019^128^ | 14 | CIS-Fatigue severity | - Mean (SD): 43.0 (5.3) |  |
| Kalkman 2005^131^ |  | CIS | - Fatigue severity mean (SD): 36.5 (12.5) - Activity, mean (SD): 10.1 (5.5) - Concentration, mean (SD): 12.3 (8.2) - Motivation, mean (SD): 12.0 (5.9) |  |
| Koopman 2017^132^ | 24 | CIS- Fatigue severity | - Mean (SD): 43.1 (5.8) |  |
| Leclair-Visonneau 2013^113^ | NR | FSS | Instrument cited in Methods, but no data presented. |  |
| Minis 2010^134^ | 1349 | CIS | Data reported by subgroup of employed / not employed |  |
| van der Kooi 2007^143^ | 65 | CIS-Fatigue Severity | Overall population, baseline:   - Mean (SD): 28.8 (12.1)   Placebo, no training (n=16):   - Baseline mean (range): 27.1 (21.1 – 33.0) - Week 52 mean (range): 26.1 (20.0 – 31.8)   Placebo, training (n=19)   - Baseline mean (range): 25.3 (19.8 – 30.8) - Week 52 mean (range): 27.4 (22.1 – 32.6)   Albuterol, no training (n=15)   - Baseline mean (range): 30.5 (24.3 – 36.6) - Week 52 mean (range): 28.0 (22.1 – 33.9)   Albuterol, training (n=15)   - Baseline mean (range): 33.3 (27.1 – 39.4) - Week 52 mean (range): 33.8 (27.9 – 39.7) | Baseline subgroup data available for overall population by pain / no pain, not severely fatigued / severely fatigued.  DOF was significantly different between the training and no training groups. |
|  | 65 | DOF* | Overall population, baseline:   - Mean (SD): 3.4 (2.3)   Placebo, no training (n=16):   - Baseline mean (range): 3.6 (2.5 – 4.7) - Week 52 mean (range): 3.7 (2.7 – 4.6)   Placebo, training (n=19)   - Baseline mean (range): 2.7 (1.8 – 3.6) - Week 52 mean (range): 2.7 (1.8 – 3.6)   Albuterol, no training (n=15)   - Baseline mean (range): 2.7 (1.7 – 3.8) - Week 52 mean (range): 3.1 (2.1 – 4.1)   Albuterol, training (n=15)   - Baseline mean (range): 4.2 (3.1 – 5.4)   Week 52 mean (range): 3.6 (2.6 – 4.6) |  |
| van de Geest-Buit 2023^142^ | 138 | CIS-Fatigue severity | - Mean (SD; range): 34.2 (12.3; 9 – 56) - % scoring ≥cut-off score of 35 indicating severe fatigue: 5.1% |  |
| Voet  2014^144^ | 57 | CIS-Fatigue severity | Period 1  Usual care (n=24):   - Pretreatment, median (range): 42 (35-51) - 16 week, median (range): 42 (32-56) - 28 week, median (range): 42 (35-56)   Aerobic exercise training:   - Pretreatment (n=20), median (range): 41 (35-54) - 16 week (n=19), median (range): 38 (18-53) - 28 week (n=19), median (range): 39 (11-53)   Cognitive behavioral therapy (n=13):   - Pretreatment, median (range): 43 (35-56) - 16 week, median (range): 24 (12-56) - 28 week, median (range): 28 (16-56)   Period 2  Aerobic exercise training (n=8):   - Pretreatment, median (range): 41 (36-47) - 16 week, median (range): 29 (20-42) - 28 week, median (range): 29 (24-39)   Cognitive behavioral therapy (n=12):   - Pretreatment, median (range): 41 (35-51) - 16 week, median (range): 30 (8-53) - 28 week, median (range): 30 (20-42) | Significant differences were observed between aerobic exercise and cognitive behavioral therapy versus usual care . |

*No further information available on how this was derived; a higher DOF score indicates more experienced fatigue

DOF, Daily Observed Fatigue; FSS, Fatigue Severity Scale; NeuroQoL, Quality of Life in Neurological Disease; NR, not reported; SD, standard deviation; VAS, visual analogue scale

Table 32. Outcomes in studies using neurological / psychological instruments, for the topic “Humanistic Burden and PROs”

| **Author, year** | **n** | **Instrument** | **Results** | **Comment** |
| --- | --- | --- | --- | --- |
| Bungener 1998^121^ | 11 | Abrams scale | - Mean (SD): 1.6 (1.8) |  |
|  | 11 | PAS | - Mean (SD): 17.8 (8.8) |  |
|  | 11 | SAS | - Mean (SD): 12.8 (7.5) |  |
| Dos Santos 2021^100^ | 13 | F-A-S-categorical | - Mean (SD; range): 18.23 (4.41; 13 – 28) - Proportion impaired: 0% |  |
|  | 13 | F-A-S | - Mean (SD; range): 31.84 (10.69; 17 – 50) - Proportion impaired: 15.38% |  |
|  | 13 | MMSE | - Mean (SD; range): 26.76 (2.45; 23 – 30) - Proportion impaired: 46.15% |  |
|  | 13 | MoCA | - Mean (SD; range): 23.0 (3.51; 18 – 29) - Proportion impaired: 76.92% |  |
|  | 13 | RAVLT | - Total, mean (SD; range): 34.53 (12.09; 20 – 61) - Total, proportion impaired: 61.53% - Short-term, mean (SD; range): 6.76 (3.05; 4 – 15) - Short-term, proportion impaired: 76.92% - Long-term, mean (SD; range): 6.30 (3.42; 3 – 15) - Long-term, proportion impaired: 61.53% |  |
|  | 13 | TMT | - A, median (IQR; range): 80 (58.5 – 138; 44 – 182) - A, proportion impaired: 100% - B, median (IQR; range): 123 (95.5 – 263; 57 – 291) - B, proportion impaired: 69.23% |  |
| Gallais 2015^123^ | 19 | LARS | - Apathy: 21.1% - Global score, mean (SD): -22.8 (6.3) - Intellectual curiosity, mean (SD): -2.45 (0.7) - Emotion, mean (SD): -2.66 (1.2) - Action initiation, mean (SD): -2.73 (1.1) - Self-awareness, mean (SD): -2.3 (1.6) |  |
|  | 19 | MMSE | - Mean (SD): 28.9 (0.9) |  |
|  | 19 | Stroop test | - Word T score, mean (SD): 51.6 (8.9) - Color T score, mean (SD): 46.56 (9.8) - Color-word T score, mean (SD): 49.2 (7.9) - Interference score, mean (SD): 49.68 (6.5) |  |
|  | 19 | TMT | - A z-score, mean (SD): 0.9 (0.4) - B z-score, mean (SD): 0.4 (0.6) |  |
|  | 19 | FAB | - Mean (SD): 15.9 (1.4) |  |
| Jacques 2019^128^ | 14 | GSE | - Mean (SD): 30.7 (7.5) |  |
| Koopman 2017^132^ | 23 | SES | - Mean (SD): 20.4 (3.1) |  |
|  | 23 | J-FCS | - Mean (SD): 1.7 (0.6) |  |
|  | 20 | PCS | - Mean (range): 3.0 (0 – 10) |  |
|  | 24 | SSI | - Mean (SD): 9.8 (2.6) |  |
| O’Dowd 2021^136^ | NR | RSES | - Mean (SD): 21.8 (6.3) |  |
|  | NR | PSPP | - Mean (SD): 11.5 (3.7) |  |
| van der Kooi 2007^143^ | 65 | SCL | Overall population, baseline:   - Mean (SD): 117 (29)   Placebo, no training (n=16):   - Baseline mean (range): 117 (104 - 131) - Week 52 mean (range): 111 (101 - 121)   Placebo, training (n=19)   - Baseline mean (range): 112 (99 - 124) - Week 52 mean (range): 106 (97 - 116)   Albuterol, no training (n=15)   - Baseline mean (range): 116 (102 - 130) - Week 52 mean (range): 110 (100 - 121))   Albuterol, training (n=15)   - Baseline mean (range): 124 (110 - 138) - Week 52 mean (range): 123 (112 - 133) | No significant differences between training / no training and albuterol / placebo over time.  Subgroup data available for overall by pain/no pain, not severely fatigued/fatigued. |

FAB, Frontal Assessment Battery; GSE, General Self-efficacy Scale; J-FCS, Jacobsen-fatigue catastrophizing scale; LARS, Lille Apathy Rating Scale; MMSE, Mini Mental State Evaluation; NR, not reported; PAS, Physical Anhedonia Scale; PCS, Pain Catastrophizing Scale; PROs, patient-reported outcomes; PSPP, Physical Self-Perception Profile; RAVLT, Ray Auditory Verbal Learning Test; RSES, Rosenberg Self-Esteem Scale; SAS, Social Anhedonia Scale; SCL, Symptom Check List; SES, Self-Efficacy Scale; SSI, Social Support Inventory; TMT, Trail Making Test

Table 33. Outcomes in studies using instruments to assess symptoms, for the topic “Humanistic Burden and PROs”

| **Author, year** | **n** | **Instrument** | **Results** |
| --- | --- | --- | --- |
| Aprile 2013^118^ | NR | LSQ | Data only reported by subgroup of shoes with/without orthoses. |
| Koopman 2017^132^ | 24 | ICQ-acceptance | - Mean (SD): 17.4 (5.5) |
|  | 23 | IES | - Mean (range): 6.0 (0 – 37) |
|  | 24 | IMQ | - Mean (SD): 3.1 (1.1) |
| Rose 2012^139^ | 49 | IPQ-R | - Illness, mean (SD): 4.6 (2.2) - Timeline acute/chronic, mean (SD): 4.7 (0.6) - Consequences, mean (SD): 3.9 (0.8) - Personal control, mean (SD): 2.7 (0.9) - Treatment control, mean (SD): 2.3 (0.9) - Illness coherence, mean (SD): 4.0 (1.1) - Timeline cyclical, mean (SD): 3.1 (0.8) - Emotional, mean (SD): 3.1 (0.8) |

ICQ, Illness Cognitions Questionnaire; IES, Impact of Event Scale; IMQ, Illness Management Questionnaire; IPQ-R, Illness Perception Questionnaire-Revised; LSQ, Lumbar Spine Questionnaire; PROs, patient-reported outcomes

Table 34. Qualitative outcomes for the topic “Humanistic Burden and PROs”

| **Author, year** | **n** | **Domain** | **Results** | **Comment** |
| --- | --- | --- | --- | --- |
| Bakker 2017^119^ | 25 | Illness perception | The following intra-individual themes emerged:   - Feelings evoked when hearing the diagnosis - Integrating the functional consequences of the diagnosis into one’s life - Handling fatigue - Wrestling with heredity and progeny - Changes in the relationship with a partner   The following extra-individual themes emerged:   - Reponses of the social environment to the diagnosis - Postponing the use of assistive devices - The difficulty of (giving up) work |  |
| Dijkstra 2021^122^ | 20 | Pain | Prevalence of pain self-reported by participants:   - Baseline, n (%): 12 (60) - 2-year follow-up, n (%): 15 (75) | Subgroup data available by early and classic onset |
|  |  | Fatigue | Prevalence of fatigue, n (%): 12 (60) |  |
| Gergev 2013^110^ | 4 | Cognitive | Cognitive profile varied between normal abilities and mild intellectual disability. All patients showed deficit in verbal and working memory with reduced learning capacity. |  |
| Goselink 2018^124^ | 24 | Pain | Pain was a frequent complaint, n (%): 15 (63%). |  |
|  |  | Fatigue | Fatigue was reported by 83% (n=80) of respondents. |  |
| Guy-Coichard 2008^125^ | 119 | Pain | 91 (75.8%) patients reported pain in the previous 3 months. |  |
| Hamel 2019^126^ | 304 | Symptoms | The following symptomatic themes were reported:   - Problems with shoulders or arms: 96.9% - Limitations with mobility or walking: 93.6% - Inability to do activities: 94.7% - Back, chest or abdomen weakness: 93.8% - Changed body image due to disease: 91.6% - Fatigue: 93.8% - Problems with physical health: 75.4% - Pain: 87.7% - Problems with hands or fingers: 68.6% - Decreased performance in social situations: 76.9% - Decreased satisfaction in social situations: 71.9% - Emotional issues: 75.4% - Difficulty thinking: 30.3% - Problems eating: 41.8% - Communication difficulties: 32.9% | Subgroup data available by duration of symptoms, education, pain level, age and sex. |
| Heatwole 2012^111^ | 328 | Symptoms | Problems with shoulders or arms (96.9%); inability to do activities (94.7%); fatigue (93.8%); back, chest, or abdomen weakness (93.8%); and limitations with mobility or walking (93.6%) were the symptomatic themes with the highest frequency in FSHD. |  |
| Hodge 2016^112^ | 48 | Disability | Patients reported past (n=22; 45.8%) and current (n=10; 20.8%) concerns about speech; 19 (39.6%) had received speech therapy. |  |
| Johnson 2012^130^ | 20 | Symptoms | 1,375 direct quotes were coded identifying 251 symptoms of importance. These symptoms represented 14 symptomatic themes and 4 major domains: physical health (617), mental health (267), social health (332), and FSHD specific issues (122).  Most frequently identified symptoms were “impaired walking” (n=16; 51 quotes), “difficulty with stairs” (n=15; 29 quotes), “reliance on family members” (n=15; 26 quotes), “difficulty lifting objects” (n=14; 25 quotes), “limitations physically on what one can do” (n=13; 24 quotes), and “difficulty getting places without handicap accessibility” (n=10; 24 quotes). |  |
| Schipper 2017^140^ | 25 | Fatigue | Patients described fatigue as:   - An overwhelming and unpredictable experience - A complicated combination of causes - The far-reaching influences in their lives - An ongoing process of adaptation |  |
| Sezer 2022^141^ | 16 | Disability | Among 7 participants with moderate facial weakness 9 participants with severe facial weakness, the following themes emerged:   - Facial weakness affects multiple aspects of life - Trying to stay down to earth - Focus on self-management - Avoiding discussing facial weakness - Thoughts on expert support |  |
| Shirilla 2019^115^ | 390 | Disability | Most common factors affecting quality of life “very much” were foot/leg weakness (54%) and arm/shoulder weakness (49%) |  |
| Statland 2012^116^ | 343 | Disability | Baseline: The average self-reported total impairment was 10.4 (moderately impaired).  Follow-up: The self-reported total impairment score increased by 0.78. | Time period of follow-up not reported |

#### 7.5 Description of instruments for “Humanistic Burden and PROs”

##### 7.5.1 Abilhand

The Abilhand questionnaire is an interview-based test assessing a patient’s perceived difficulty in using their hand to perform manual activities, by measuring the ability to perform bimanual tasks.^248^ The 56 manual activities are rated on a four-level scale of 0 (impossible) to 3 (easy); lower scores indicate greater difficulty in performing manual activities.

##### 7.5.2 Abrams scale for emotional blunting

The Abrams scale assesses emotional blunting.^249^ Scores range from 0 – 30.

##### 7.5.3 Barthel index

The Barthel index is a 10-item scale that measures functional independence in the domains of personal care and mobility.^250^ For each item, patients receive a score of 0 if they are unable to carry out the activity; totals range from 0 to 100. Scores of 0 – 20 represent total dependent, while a score of 100 represents totally independent.

##### 7.5.4 BDI: Beck Depression Inventory

The BDI is a self-administered 21-item self-report scale, where each item is rated, ranging from the absence of a symptom to an intense level.^251^ Ratings are summed linearly to create a score which ranges from 0 to 63, with higher scores indicating greater degrees of depression. The BDI short form produces a binary outcome of “not depressed” or “depressed” for patients with a cutoff score above 4.^252^

##### 7.5.5 BPI: Brief Pain Inventory

The BPI assesses the severity of pain and its impact on functioning by measuring both the intensity of pain (sensory dimension) as well as the interference of pain in a patient’s life (reactive dimension).^253^ Scores range from 0 to 10, and the total pain severity score is an average of the items. Higher scores indicate greater pain.

##### 7.5.6 BUE: Brooke Upper Extremity scale

The BUE is a six-point scale that allows for the classification of upper extremity function and also helps document progression.^254^ The BUE score ranges from 1 to 6, where 1 indicates normal arm and hand function and 6 indicates that no arm and hand function is left.

##### 7.5.7 CIS: Checklist Individual Strength

The CIS measures four dimensions of fatigue: fatigue severity, concentration problems, reduced motivation and activity.^255^ The 20 statements on fatigue-related problems are scored using a 7-point Likert scale; higher scores indicate greater fatigue.

##### 7.5.8 Covi brief anxiety scale

The Covi scale is a scale designed to determine the intensity of anxiety and is determined by three parameters: patient complaints, his/her behavior and somatic manifestations.^256^ Total score ranges from 0 – 9, with higher scores indicating more intense anxiety.

##### 7.5.9 CUE: Capabilities of Upper Extremity scale

The CUE measures functional limitation and assesses the amount of difficult experienced in performing specific actions with one or both arms and hands with the 32-items across 7 domains.^257^ The CUE is scored on a 7-point scale from 1 (totally limited, can’t do it at all) to 7 (not at all limited); scores range from 32 to 224 with higher scores indicating greater function.

##### 7.5.10 DOP: Daily Observed Pain score

The DOP was used to assess the intensity of pain on a daily self-observation list during a two-week period.^234^ Pain severity was rated four times a day on a scale of 0 (no pain) to 4 (severe pain). The DOP score can range from 0 to 16, and the 14 daily pain scores were averaged into one DOP score.

##### 7.5.11 EHD: Depressive mood scale

The EHD is a 20-item scale, with 10 items assessing emotional changes expressed by the patient and 10 items assessing the emotional state as perceived by the investigator observing the patients’ facial reactions, speech and motor expressiveness. Each item is rated from 0 (absent) to 4 (severe).^258^

##### 7.5.12 EQ-5D: EuroQoL 5-dimension

The EQ-5D is a simple health state measure consisting of five questions that relate to five dimensions of health: mobility, self-care, usual activities, pain/discomfort and anxiety/depression.^259^ Each dimension is divided into three levels of severity, resulting in 243 possible combinations in total.

##### 7.5.13 ESS: Epworth Sleepiness Scale

The ESS is a self-administered questionnaire to provide a measurement of the subject’s general level of daytime sleepiness.^260^ The ESS lists eight situations, where the tendency to become sleepy is rated from 0 (no chance of dozing) to 3 (high chance of dozing); scores range from 0 to 24, and higher scores indicate higher levels of sleepiness

##### 7.5.14 FAB: Front Assessment Battery

The FAB is a brief tool that is used to assess frontal lobe function.^261^ Total score is from a maximum of 18; higher scores indicate better performance.

##### 7.5.15 FaCE: Facial Clinimetric Evaluation scale

The FaCE scale measures both impairment and disability associated with facial dysfunction.^262^ There are 15 items rated on a 5-point Likert scale and items are categorized into 6 domains: facial movement, facial comfort, oral function, eye comfort, lacrimal control and social function. Each domain is scored out of a total of 100, where 0 is the worst and 100 is the best; the mean derived from all domains determines the total score (out of 100).

##### 7.5.16 F-A-S test

The F-A-S test measures phonemic word fluency, as part of cognitive function, by requesting individuals to orally produce as many words as possible that being with the letters F, A and S within a time frame.^263^ Scores are determined by adding all the words spoken with the three letters; score is affected by education and age.

##### 7.5.17 F-A-S-categorial test

The F-A-S categorical test is like the F-A-S test but asks for words within a given category. When used with animals, a score of 9 or greater named animals for subjects with up to 8 years of formal education and 13 or greater named animals for those with over 9 years of formal education is the cutoff.^264^

##### 7.5.18 FSHD-HI: Facioscapulohumeral Dystrophy Health Index

The FSHD-HI is a 11-6item disease-specific patient reported outcome measure for designed to assess the QOL of FSHD patients across four major domains (physical, mental, social health and FSHD-specific issues) and 14 subdomains.^130^

##### 7.5.19 FSHD-HI Peds: Facioscapulohumeral Dystrophy Health Index Pediatric

The FSHD-HI Peds is a disease-specific patient reported outcome measure that measures change across 14 areas of disease-specific health that was formulated based on the original scale.^130^ A total of 109 items are included, divided into 14 subscales assessing a child’s perception of the shoulder and arm function, mobility and ambulation, fatigue, cognitive function, activity limitation, trunk strength and function, gastrointestinal function, social performance, body image, hand/finger function, social satisfaction, pain, emotional health and communication. A score of 0 – 100 is given for each of the subscales, with higher scores representing higher burden of disease.

##### 7.5.20 FSHD-RODS: Facioscapulohumeral Dystrophy Rasch-built Overall Disability Scale

The FSHD-RODS is a disease-specific measure that assesses facial function and is suitable for detecting activity and participation restrictions in patients with FSHD.^69^ It has a total of 38 items.

##### 7.5.21 FSS: Fatigue Severity Scale

The FSS is a 9-item instrument that assesses the effect of fatigue on daily functioning.^265^ Respondents use a scale of 1 (completely disagree) to 7 (completely agree), along with a VAS, to rate each statement; higher scores indicate more severe fatigue.

##### 7.5.22 Grading of chronic pain scale

The grading of chronic pain scale asks respondents to rate pain on a scale of 0 (no pain) to 10 (pain as bad as can be).^266^

##### 7.5.23 GSE: General Self-efficacy Scale

The GSE is a 10-item scale designed to assess optimistic self-beliefs to cope with a variety of difficult demands in life.^267^ Statements are rated on a 4-item scale and items are summed for a total score; higher scores indicate higher self-efficacy.

##### 7.5.24 HADS: Hospital Anxiety and Depression Scale

The HADS is a self-report rating scale of 14 item son a 4-point Likert scale (range 0 – 30), and is designed to measure anxiety and depression with 7 items for each subscale.^268^ The total score is the sum of the 14 items, and for each subscale, the score is the sum of the respective seven items (ranging from 0 – 21), with higher scores indicating more anxiety and/or depression.

##### 7.5.25 HAQ: Health Assessment Questionnaire

The HAQ is a self-reported functional measure which assesses function in 8 activity domains, giving scores for each domain as well as a total score; a high score indicates greater disability.^269^

##### 7.5.26 HDRS: Hamilton Depression Rating Scale

The HDRS is a 17-item depressive rating scale, ranging from 0 – 51.^270^ Higher scores indicate greater severity of depression.

##### 7.5.27 ICQ: Illness Cognitions Questionnaire

The ICQ is an 18-item questionnaire that contains three 6-item scales related to helplessness, acceptance and perceived benefits, each with a score range of 6 – 24.^271^ Higher scores represent a higher degree of acceptance of the disease.

##### 7.5.28 ID Pain

ID pain is a 6-item pain questionnaire asking participants regarding the nature of their pain.^272^ Higher scores suggest a neuropathic pain component.

##### 7.5.29 IES: Impact of Event Scale

The IES is a 15-ite questionnaire that assesses subjective distress caused by traumatic events.^273^ Each item is rated on a 4-point Likert scale, with a total range of scores from 0 – 75. Higher scores represent higher impact on being diagnosed with the disease.

##### 7.5.30 IMQ: Illness Management Questionnaire

The IMQ is a questionnaire designed to assess coping with fatigue.^274^ There are 9 items, each rated on a 6-point Likert scale. Higher scores represent more focus on fatigue symptoms.

##### 7.5.31 INQoL: Individualized Neuromuscular Quality of Life Questionnaire

The INQoL is a neuromuscular disease-specific measure of QoL.^275^ The final score of each section, and total INQoL score, is presented as a percentage of the maximum detrimental impact where 100 is the greatest impact on QoL.

##### 7.5.32 IPQ-R: Illness Perception Questionnaire – Revised

The IPQ-R provides a quantitative assessment divided into three sections: identity subscale, causal subscale and a third section containing seven subscales (consequences, timeline acute/chronic and cyclical, personal and treatment control/cure, illness coherence and emotional representations).^276^ It is an 84-item instrument; some items are answered with yes or no, other others using a 5-point Likert scale. All items for each of the subscales are summed to give an overall score. Higher scores on the identify, consequences, timeline acute/chronic and cyclical subscales represent strongly held beliefs; high scores on the personal and treatment control and coherence subscales represent positive beliefs.

##### 7.5.33 J-FCS: Jacobsen-Fatigue Catastrophizing Scale

The J-FCS is a scale designed to measure the relationship of catastrophizing to fatigue.^277^ There are a total of 10-tiems, each rated on a 5-point Likert scale (range 1 – 5); higher scores indicate more fatigue-related catastrophizing.

##### 7.5.34 KIDSCREEN

The European KIDSCREEN-52 is a generic health-related quality of life questionnaire to assess the subjective health and the psychological, mental and social well-being for children and between the ages of 8 and 18.^278^ The questionnaire can be completed both by the children themselves or by a proxy assessment for parents. The KIDSCREEN covers 10 domains: physical well-being; psychological well-being; mood and emotions; self-perception; autonomy; parent relation and home life; financial resources; social support and peers; school environment; and social acceptance (bullying).

##### 7.5.35 LARS: Lille Apathy Rating Scale

The LARS assesses apathy via 33-items, divided into 9 domains; eight of the domains concern the main clinical manifestations of apathy and the ninth domain refers to self-awareness.^279^ Responses are either on a 5-point Likert scale or via yes/no. Each of the nine domains can be evaluated separately, or equally weighted to contribute to a global score. The global score ranges from -36 to +36 with higher scores representing a greater degree of apathy.

##### 7.5.36 Low Back Pain Rating scale

Low Back Pain Rating scale is an index scale which includes measurements of pain intensity, disability and physical impairment.^280^ The scale includes 21 self-reported items and 4 performance-based measures, and back and leg pain, disability and physical impairment. The higher the score on the scale, the greater the level of disability and impairment.

##### 7.5.37 LSQ: Lumbar Spine Questionnaire

The LSQ was designed by the North American Spine Society and includes items on comorbidity, general well-being, symptoms of pain aner nerve compression, functional capacity and occupational status.^281^ It contains 62-items; higher scores indicate greater pain and functional limitation and more frequent and intense neurologic symptoms.

##### 7.5.38 MADRS: Montgomery and Asperg Depression Rating Scale

The MADRS stratifies the severity of depressive episodes in adults based on clinical interviews with patients.^282^ Clinical judgment is used to determine whether the rating lies on the defined scale steps, or is between them (denoted as “Worsening symptoms”). The MADRS consists of 10 items, with higher ratings indicating worse depression.

##### 7.5.39 MoCA: Montreal Cognitive Assessment

The MoCA is a rapid screening test to detect mild cognitive impairment.^283^ The cutoff is 26 points; individuals with 12 or less years of education receive an extra point.

##### 7.5.40 MINI: Mini-International Neuropsychiatric Interview

The MINI is a short, structured, diagnostic interview and assesses 10 psychiatric disorders.^284^

##### 7.5.41 MMSE: Mini Mental State Evaluation

The MMSE is designed to be used to detect the presence of cognitive impairment.^285^ It consists of 11 questions that look at various functions including arithmetic, memory and orientation. Scoring is based on the number of correct items, with a total score of 30; scores of 23 or less indicate the presence of cognitive impairment corresponding to mild (18-23) or severe (0-17).

##### 7.5.42 MPQ: McGill Pain Questionnaire

The MPQ consists is used by patients to specify the subjective pain experience by using three classes of word descriptors: sensory, affective and evaluative.^286^ Scores range from 0 to 78, with higher scores indicating greater pain.

##### 7.5.43 NEADL: Nottingham Extended Activities of Daily Living

The NEADL is a questionnaire that monitors the level of activities of daily living disability in patients, across four categories: mobility, kitchen activities, domestic activities and leisure activities.^287^ The instrument includes 22 items, all of which are scored on a four-point scale ranging from 0 (not at all) to 3 (alone easily); lower scores indicate greater impairment.

##### 7.5.44 NeuroQoL: Quality of Life in Neurological Diseases

The NeuroQoL measures the physical, mental and social effects experienced by adults and children living with neurological conditions.^288^ The NeuroQoL fatigue domain 19-items for the adult version and 11-items for the child version. NeuroQoL scores have a mean of 50 and a SD of 10 in a referent population. Scores of 0.5 – 1.0 SD worse than the mean equate mild symptom/impairment; scores of 1.0 – 2.0 SD worse than the mean equate moderate symptom/impairment; and scores of 2.0 SD or more worse than the mean equate severe symptom/impairment.

##### 7.5.45 NHP: Nottingham Health Profile

The NHP is a 38-item questionnaire that assesses the domains of physical mobility, pain, sleep, social isolation, emotional reactions and energy level.^289^ Items are rated in terms of relative importance and then rescaled to vary between 0 and 100 within each section

##### 7.5.46 PAS: Physical Anhedonia Scale

The PAS measures physical anhedonia (the inability to feel pleasure from physical or sensory stimuli), through a 61-item measure using true/false response items.^290^ High scores indicate less ability to experience pleasure from physical and sensory stimuli.

##### 7.5.47 PCS: Pain Catastrophizing Scale

The PCS is a 13-item scale, with three subscales to assess the different aspects of pain catastrophizing.^291^ Each item is rated on a 5-point Likert scale, with higher scores representing more pain-related catastrophic thoughts and feelings.

##### 7.5.48 PedsQL NMM: Pediatric Quality of Life Neuromuscular Module

The PedsQL NMM measures health-related quality of life in children with neuromuscular disease. Problems experienced in the past month in relation to neuromuscular disease symptoms, ability to communicate and family resources are rated. Scoring is based on a 0 – 4 ordinal scale (range 0=never and 4=almost always). Scores are then linearly transformed from a 0 -11 scale to a 0 – 100 scale, where higher scores indicate better health-related quality of life.^292^

##### 7.5.49 PGIC: Patient Global Impression of Change

The PGIC reflects a patient’s belief about the efficacy of a treatment, related to their chronic pain.^293^ Respondents are asked how treatment has impacted their painful condition using a 7-point Likert scale.

##### 7.5.50 PSPP: Physical Self-Perception Profile

The PSPP is a scale that assesses four subdomains theorized to comprise the physical self (body attractiveness, sport competence, physical strength and sport conditioning) with a fifth subscale assessing global physical self-worth.^294^ The 30-item inventory results in a score from 6 – 24 with higher scores indicating higher perceived competence or adequacy.

##### 7.5.51 PSQI: Pittsburg Sleep Quality Index

The PSQI is a self-rated questionnaire which assesses sleep quality and disturbances over a 1-month time interval.^295^ The seven component scores are scored from 0 (no difficulty) to 3 (severe difficulty); the component scores are summed to produce a global score (range 0 to 21). Higher scores indicate worse sleep quality.

##### 7.5.52 RAVLT: Rey-Auditory Verbal Learning Test

The RAVLT is a word learning test where five presentations of a 15-word list are given and measures the rate of learning.^296^ Scoring is assessed by age (20 to 59 and over 60) and gender (male and female).

##### 7.5.53 RSES: Rosenberg Self-Esteem Scale

The 10-item RSS measures self-esteem with a 4-point Likert scale.^297^ Scores range from 0 – 30, with scores below 15 suggesting low self-esteem.

##### 7.5.54 SAAS: Social Appearance Anxiety Scale

The SAAS is a 16-item self-report measure that assesses fear of situations in which one’s appearance will be evaluated.^298^ Items are rated on a 5-point Likert scale with higher scores indicated higher social appearance anxiety.

##### 7.5.55 SAS: Social Anhedonia Scale

The SAS measures social anhedonia (the inability to find pleasure experienced when interacting with others and interest in having close social connections) through a 40-item self-report measure using true/false responses.^290^ Higher scores indicate less ability to experience pleasure from social and interpersonal experiences.

##### 7.5.56 SCL: Symptom Checklist

The SCL Is a 90-item questionnaire used to assess psychological problems.^299^ Each item is graded from 1 to 5 according to severity (none to extreme); the scores are summed to create a total score. Symptoms are regarded as positive when the total score exceeds 160 points.

##### 7.5.57 SES: Self-Efficacy Scale

The SES is a scale designed to measure the sense of control over fatigue.^300^ There are a total of 7 items, each of which is measured on a 4-point Likert scale, resulting in a score ranging from 4 – 28. Higher scores represent a higher sense of control over fatigue.

##### 7.5.58 SF-36: Short-Form 36

The SF-36 consists of 36 questions that cover the general health status of patients.^301^ The questionnaire assesses health-related quality of life across eight domains: physical functioning; role-physical; bodily pain; general health; vitality; social functioning; role-emotional; and mental health. These domains can be summarized in two main scores as a physical composite score and a mental composite score; these component scores are calculated such that for each score, there is a mean of 50 (SD of 10). Lower scores indicate worse quality of life. Results may be compared with those of the general population.

##### 7.5.59 SIAS: Social Interaction Anxiety Scale

The SIAS measures social interaction anxiety through 20 items that are rated on a 5-point Likert scale, from “not at all characteristic of me” to “extremely characteristic of me”.^302^ A total score is generated by summing the ratings with higher scores indicating higher social interaction anxiety.

##### 7.5.60 SIP: Sickness Impact Profile

The SIP is a measure of health status and measures patients dysfunction through everyday behavior.^303^ The general score of general disability (total score) is calculated by adding the weights of items in the 12 categories or subscales: sleep / rest; emotional behavior; body care / movement; household management; mobility; social interactions; ambulation; alertness behavior; communication; work limitations; recreation / pastimes; and eating. The total SIP score ranges from 0 to 10,298; higher scores mean more disability.

##### 7.5.61 SSI: Social Support Inventory

The SSI is a scale to assess the perceived adequacy of social support.^304^ There are a total of 8 items on the scale, with each rated using a 4-point Likert scale, resulting in a range of scores from 0 – 24. Higher scores represent more discrepancies between the received amount of social support and the desired amount of social support.

##### 7.5.62 STAI: State-Trait Anxiety Inventory

The STAI is a psychological inventory consisting of 40 self-report items on a 4-point Likert scale, measures both state anxiety and trait anxiety.^305^ Higher scores correlate with higher levels of anxiety

##### 7.5.63 Stroop test

The Stroop test is a neuropsychological test used to assess the inability to inhibit cognitive interference that occurs when the processing of a specific stimulus feature impedes the simultaneous processing of a second stimulus attribute, known as the Stroop effect.^306^ World, color and color-word T-scores of 40 or less are considered “low”.

##### 7.5.64 TMT: Trail Making Test

The TMT is widely used to assess executive function, assessing mental abilities such as letter and number recognition, mental flexibility, visual scanning and motor function.^307^ Performance is evaluated using two different parts: Part A involves connecting numbers and Part B involves connecting numbers and letters. Scoring is based on timing with normal scores for Part A ranging from 1 – 39 seconds (40 or more seconds indicates brain damage) and normal scores for Part B ranging from 1 – 91 seconds (92 or more seconds indicates brain damage).

##### 7.5.65 Tyrer anxiety scale

The Tyrer anxiety scale is suitable for recording anxious symptoms and is a subdivision of the Comprehensive Psychopathological Rating Scale.^308^ It comprises 10 items, all of which are rated on a 7-point scale, resulting in a total score ranging from 0 - 60.

##### 7.5.66 Universal pain assessment tool

The universal pain assessment tool is a visual assessment scale of 0 – 10, with verbal descriptors, facial grimace scales and activity tolerance prompts to assess pain according to individual patient needs.^309^ Higher scores indicate greater pain, with a 10 equivalent to the worst pain possible.

##### 7.5.67 VAS: Visual Analogue Scale

A VAS can be used to measure health-related quality of life, pain or other domains, by asking respondents to rate their current status on a continuous line between 0 to 100, or 0 cm to 10 cm. Higher scores indicate worse outcomes.

##### 7.5.68 VLE: Vignos lower extremity scale

The VLE scale assess functional performance for lower extremetities.^310^ The scale ranges from 0 to 10, with higher scores indicate worse functional performance. A score of 1 equals walking and climbing stairs without assistance and 10 means the patient is bed-bound.

##### 7.5.69 WFS: Walton Functional Scale

The WFS assesses functional disability.^311^ Scores range from 0 to 10, with a higher score representing a higher degree of disability.

#### 7.6 Detailed tables for “Economic Burden”

Table 35. Study and population characteristics for “Economic Burden” studies

| **Author, year** | **Country** | **Study design** | **FSHD population type: n (%)** | **% pediatric** | **Age (years)** | **Female,**  **n (%)** | **Age at onset (years)** | **Disease duration (years)** | **CSS** | **D4Z4 pairs (kb)** |
| --- | --- | --- | --- | --- | --- | --- | --- | --- | --- | --- |
| CONFERENCE ABSTRACTS | | | | | | | | | | |
| Konersman 2022a^154^ | US | Retrospective cohort | FSHD NS: 79 (100%) | --- | --- | --- | --- | --- | --- | --- |
| Konersman 2022b^312^ | US | Retrospective cohort | FSHD NS: 290 (100%) | --- | --- | --- | --- | --- | --- | --- |
| Statland 2012^116^ | --- | Retrospective cohort | FSHD NS: 343 (100%) | NS | 51 (NR) | 51% | 22 (NR) | --- | --- | 25^a^ |
| Winter 2009^152^ | Germany | Prospective cohort | FSHD NS: 20 (100%) | --- | --- | --- | --- | --- | --- | --- |
| Wood 2015^147^ | UK | Retrospective cohort | Type 1: 416 (100%) | NS | 47.6 ±16.5  (4-83) | 48.1% | --- | --- | --- | --- |
| Zielman-Blokhuis 2020^153^ | Netherlands | Cross-section survey | FSHD NS: 172 (100%) | --- | --- | --- | --- | --- | --- | --- |
| FULL TEXT ARTICLES | | | | | | | | | | |
| Bakker 2017^119^ | Netherlands | Qualitative | FSHD NS: 25 (100%) | 0% | 56 (24 – 77) | 44% | --- | 16 (1 – 49) | --- | --- |
| Blokhuis 2021^150^ | Netherlands | Cross-sectional survey | FSHD: 172 (31% Type 1; 4% Type 2; 1% Type 1 and 2; 64% NS) | 0% | 56 (45-66)^b^ | 55% | 20 (12-40)^b^  Range: 0-70 | 30 ±16 (0-75) | --- | --- |
| Fitzgerald 2016^157^ | US | Prospective survey | FSHD NS: 193 (100%) | 0% | 57.5 ±13.6 | 44% | 22.6 ±15.0 | 35.0 ±15.7 | --- | --- |
| Fowler 1997^156^ | US | Cross-sectional survey | FSHD NS: 16 (100%) | NS | 42.4 ±11.2 | --- | --- | 23.9 ±13.1 | --- | --- |
| Hilbert 2012^155^ | US | Retrospective cohort | FSHD NS: 574 (100%) | 0% | 46.1 ±15.9 | 52.3% | 21.5 ±15.0 | --- | --- | --- |
| Hoffmann 2022^127^ | US | Prospective survey | FSHD: 690 (52% Type 1; 7% Type 2) | 0% | NS | 50% | NR (20 – 39) | --- | --- | --- |
| Jensen  2008^129^ | US | Cross-sectional survey | FSHDH NS: 127 (100%) | 0% | 51.8 ±13.8 | 52% | --- | --- | --- | --- |
| Katz 2021^148^ | US | Prospective cohort | Type 1: 578 (100%) | NS | 59 (46, 70)^b^ | 47.9 | 17 (12, 29.5)^b^ | --- | --- | Repeat length:  1-3: 60 (10.4%)  4-7: 340 (58.8%)  8-10: 119 (20.6%) |
| Moris 2018^135^ | UK | Prospective cohort | Type 1: 398 (100%) | NS | 47 (60.6)^b^ | 50.4% | 17.0 (21.2)^b^ | 26.0 (27.0)^b^ | --- | 25.0 (10.0)^b^ |
| Schepelmann 2010^151^ | Germany | Cross-sectional survey | FSHD NS: 20 (100%) | NS | ---^c^ | 65% | --- | --- | --- | --- |
| Sezer 2022^141^ | Netherlands | Qualitative | FSHD NS: 16 (100%) | 0% | 47 (24 – 77) | 56.3% | --- | --- | --- | --- |
| Statland 2014^149^ | US | Retrospective cohort | Type 1: 313 (100%) | NS | 51.5 ±17.0 | 53% | 21.1 ±15.0 | --- | --- | Contraction size: 24.8 ±5.7 |

Data are presented as mean ± SD or ranges, unless otherwise specified. --- represents data not reported.

FSHD, facioscapulohumeral muscular dystrophy; IQR, interquartile range; NR, not reported; NS, not specified; SD, standard deviation; UK, United Kingdom; US, United States.

^a^ Median with or without (range)

^b^ Median (IQR)

^c^ Reported categorically

Table 36. Outcomes of studies reporting “Economic Burden”

| **Author, year** | **Treatment arm** | **Follow-up** | **Cost and value** | **HRCU and value** | **Other** |
| --- | --- | --- | --- | --- | --- |
| CONFERENCE ABSTRACTS | | | | | |
| Konersman 2022a^154^ | FSHD (n=79) | Two yrs pre-dx vs two yrs post-dx | Post vs pre-dx medical costs (PMPY) (yr NS $): ↑ 5,646*  Post vs pre-dx costs (yr NS $):   - “other lower respiratory disease”: ↑153 - “immunizations”: ↑24 - malaise and fatigue”: ↓30 - other connective tissue disease”: ↑1,477 - “respiratory failure/insufficiency/arrest”: ↑1,169 - “cardiac dysrhythmias”: ↑969 | Post vs pre-dx care days (PMPY): ↑ 6*  Post vs pre-dx ED care (PMPY): ↑ 16.5%* | --- |
| Konersman 2022b^312^ | FSHD (n=290) | 1 yr pre-diagnosis | Cost of prescriptions (yr NS $):   - Sex hormones and modulators of the genital system: 202* - Analgesics: 126* - Psychoanaleptics: 155* | Number of prescriptions:   - Sex hormones and modulators of the genital system: 2.62* - Analgesics: 1.64* - Psychoanaleptics: 2.83* | --- |
|  | MC (n=1,450) | 1 yr pre-index | Cost of prescriptions (yr NS $):   - Sex hormones and modulators of the genital system: 72 - Analgesics: 20 - Psychoanaleptics: 155 | Number of prescriptions:   - Sex hormones and modulators of the genital system: 1.03 - Analgesics: 1.22 - Psychoanaleptics: 0.67 | --- |
| Statland 2012^116^ | FSHD (n=343) | Mean: 6 yrs (range: 1-9 yrs) | --- | At baseline:   - Use of ankle brace, leg brace, cane, or walker: 41% - Use of wheelchair at some point in the day: 24% - Use of breathing machine: 4% - Use of hearing aid: 7%   At follow-up, among device-free patients at baseline:   - Use of ankle brace, leg brace, cane, or walker: 11% - Use of wheelchair at some point in the day: 22% | --- |
| Winter 2009^152^ | FSHD (n=20) | --- | Total per-patient annual cost from the societal perspective: (2009 €): 26,240 (95% CI: 17,770-37,940) | --- | --- |
| Wood 2015^147^ | FSHD (n=416) | --- | --- | Full-time wheelchair use: 18%  Part-time wheelchair/ other assistive device use: 40%  Ventilator use (at least part-time): 8.9% | --- |
| Zielman-Blokhuis 2020^153^ | --- | --- | Per-patient annual direct costs (yr NS €): 14,282  Informal care and transport (yr NS €): 6,974  Loss of productivity (yr NS €): 5,066  Total socioeconomic burden per FSHD patient per year (yr NS €): 40,850 | --- | Monetary value of the loss in QoL (yr NS €): 14,528  The per-patient annual direct costs of illness are three times higher than the per-person health expenditures in the Netherlands. |
| FULL TEXT ARTICLES | | | | | |
| Bakker 2017^119^ | FSHD (n=25) | --- | --- | Ventilator use at night: 2 (8%)  Dependent on wheelchair: 2 (8%)  Wheelchair for long distances only: 6 (24%) | --- |
| Blokhuis 2021^150^ | FSHD (n=172) | --- | Total annual per-patient cost of illness (2018€): 26,322   - Per-patient direct costs of illness: 21,256   - Annual direct medical costs: 12,077   - Annual direct non-medical costs: 9,179 - Indirect costs: 5,066   Total socioeconomic burden per FSHD patient per year (2018€): 40,850 | In the preceding 3 months:   - Physical therapist visits: 58% - GP visits: 53% - Rehabilitation physician visits: 26% - Occupational therapist visits: 20% - Neurologist visits: 19% - Psychologist visits: 10% - Hospital admissions: 3%   In the past year:   - Purchase of aids and devices: 36% - House adaptations: 21%   Mean hours of informal care: 22h per week (SD 18h, range 1–56h per week  Employment:   - In paid work: 42%   - Absenteeism in the past 4 weeks: 12%   - Median absence duration: 152 days   - Presenteeism: 48% - Retired: 23% - Permanently disabled: 24% | Mean total annual costs by mobility level (2018 €):   - Without mobility aid: 14,008 - With mobility aid: 29,389 - Unable to walk: 63,765   The per-patient direct costs of illness are approximately five times higher than the mean per-capita health expenditures in the Netherlands. |
| Fitzgerald 2016^157^ | FSHD (n= 130) | --- | --- | Pill burden and employment:   - Participants taking 6+ were more likely to be unemployed (75.4%) than those taking 0 - 5 medications (41.9%) ** | 7.7% (23 patients) indicated that costs inference with the medication adherence. |
| Fowler 1997^156^ | FSHD NS (n=16) | --- | --- | Employed: 9  Total unemployed: 7  Employed in past: 6 | --- |
| Hilbert 2012^155^ | FSHD (n=487) | Mean: 4.7 yrs | --- | Wheelchair use:   - Baseline: 29.6% - Follow-up: +28.6%   Wheelchair use always:   - Baseline: 8.3% - Follow-up: 18.1%   Leg brace use:   - Baseline: 11.3% - Follow-up: +2%   Cane use:   - Baseline: 0% - Follow-up: +5% | The majority of FSHD members who reported increased wheelchair use reported a change from no wheelchair use at enrollment to wheelchair use for long distances at follow-up (13.9%).  At baseline, 50% of all adult members reported reduced employment opportunities. |
| Hoffmann 2022^127^ | FSHD (n=690) | --- | --- | Wheelchair use: 25.5%  Scooter: 16.2%  Walking stick or walker: 41.0%  Spinal or leg brace: 22.8% | 38.2% of patients used no mobility aides. |
| Jensen  2008^129^ | FSHD NS (n=127) | --- | --- | Cane/crutches: 32.3%  Walker: 17.3%  Scooter: 18.1%  Manual wheelchair: 11.0%  Electric wheelchair: 17.3% | No assisted device use: 35.4% |
| Katz 2021^148^ | Type 1 (n=578) | Mean: 9 years | --- | Wheelchair use:   - Baseline: 5%   Use of a breathing machine   - Baseline: 4.8% | --- |
| Moris 2018^135^ | Type 1 (n=398) | --- | --- | Wheelchair use:   - No use: 244 (62.2%) - Part-time: 94 (24.0%) - Full-time: 54 (13.8%) | --- |
| Schepelmann 2010^151^ | FSHD NS (n=20) | --- | Total annual cost (2009€): 26,240 (95% CI: 17,770 – 37,940)  Total direct annual cost (2009€): 9,270 (95% CI: 6,610 – 14,420)  Total indirect annual cost (2009€): 16,980 (95% CI: 10,440 – 24,980)  Mean annual costs of NMD-specific drugs (2009€): 2,400 (95% CI: 1,450 – 3,510) | Formal care:   - Formal care required by 14% (n=3) FSHD patients, for 5 hours per week. | --- |
| Sezer 2022^141^ | FSHD NS (n=16) | --- |  | Wheelchair use:   - Yes: 12 (75%) - Yes, only for long distances: 3 (25%) |  |
| Statland 2014^149^ | FSHD1 (n=313) | Mean: 6.2 yrs |  | Prevalence of:   - Wheelchair use at any time: 22.7% (95% CI: 18.0 – 27.3) - Wheelchair bound: 6.4% (95%% CI: 3.7-9.1) - Ankle brace: 18.5% (95% CI: 14.2 – 22.8) - Cane: 22.4% (95% CI: 17.7 – 27.0) - Walker: 9.3% (95% CI: 6.1 – 12.5) - Breathing machine: 3.5% (95% CI: 1.5 – 5.6) - Hearing aids: 7.7% (95% CI: 4.7 – 10.6) |  |
|  | Large contractions (≤18 kb) (n=52) |  |  | Prevalence of:   - Wheelchair use at any time: 42.3% (95% CI: 28.9-55.7) - Wheelchair bound: 19.2% (8.5 – 29.9) - Ankle brace: 11.5% (95% CI: 2.9 – 20.2) - Cane: 11.5% (95% CI: 2.9 – 20.2) - Walker: 7.7% (95% CI: 0.2 – 14.9) - Breathing machine: 5.9% (95% CI: 0 – 12.3) - Hearing aids: 21.6% (95% CI: 10.3 – 32.9) | Participants with D4Z4 contractions ≤18 kb started using wheelchairs earlier than participants with contractions between 19 and 28 kb (24.1 yrs, 95% CI, 17.0–31.3 vs. 48.1 yrs, 95% CI, 44.0–52.3; P < 0.001) or participants with contractions >28 kb (58.6 yrs, 95% CI, 52.2–  64.9; P < 0.001). |
|  | Medium or small contractions (>18 kb) (n=261) |  |  | Prevalence of:   - Wheelchair use at any time: 18.8% (95% CI: 14.0-23.5) - Wheelchair bound: 3.8% (1.5 – 6.2) - Ankle brace: 11.5% (95% CI: 2.9 – 20.2) - Cane: 19.9% (95% CI: 15.1 – 24.8) - Walker: 9.6% (95% CI: 6.0 – 13.1) - Breathing machine: 3.1% (95% CI: 1.0 – 5.2) - Hearing aids: 5.0% (95% CI: 2.3-7.6) |  |

* Statistically significant compared to MC
** Statistically significant at p<0.001
↑, increased by; ↓, decreased by; Dx, diagnosis; FSHD, facioscapulohumeral muscular dystrophy; HCRU, health care resource use; MC, matched controls; NMD, neuromuscular disease; NS, not specified; PMPY, per-member-per-year; QoL, quality of life; SD, standard deviation; yr(s), year(s)

#### 7.7 Detailed tables for “Disease Classification”

Table 37. Study and population characteristics for “Disease Classification” studies

| **Author, year** | **Country** | **Study design** | **FSHD population type: n (%)** | **% pediatric** | **Age (years)** | **% female** | **Age at onset (years)** | **Disease duration (years)** | **CSS** | **D4Z4 pairs (kb)** |
| --- | --- | --- | --- | --- | --- | --- | --- | --- | --- | --- |
| CONFERENCE ABSTRACTS | | | | | | | | | | |
| Alobaidan 2021^158^ | UK | Cross-sectional/ survey | FSHD NS: 125 (100%) | 0% | --- | 40% | 31.0 (10–61) | --- | --- | --- |
| Banerji 2021a^161^ | UK  US | Retrospective cohort | FSHD NS: 512 (100%) | 0% | --- | --- | --- | --- | --- | --- |
| Bettio 2021^189^ | Italy | Retrospective cohort | FSHD NS and their relatives: >4,000 | 0% | --- | --- | --- | --- | --- | --- |
| Chen 2010^173^ | NR | Prospective cohort | FSHD NS: 7 (100%) | NS^*^ | 20.0 (14–33) | 29% | NR (0.4–6) | --- | --- | --- |
| Duong 2015^174^ | Multinational | Retrospective cohort | FSHD NS: 53 (100%) | NS^*^ | 23.0 ± 14.7 | 59% | --- | --- | --- | 18 ± 7 |
| Liu 2019^190^ | China | Cross-sectional/ survey | Type 1: 54 (100%) | 0% | 33.2 | 52% | 10.4 | --- | --- | 6 repeats |
| Mah 2013^175^ | Canada | Cross-sectional/ survey | FSHD NS: 11 (100%) | NS^*^ | 19.5 (8.7–50.9) | 64% | Facial: 1.0 Scapular: 5.8  Hip: 6.5  Foot: 8.0 | --- | --- | 22 (15–38) |
| Mah 2014^176^ | Multinational | Retrospective cohort | FSHD NS: 20 (100%) | NS^*^ | 16.7 (13.1–25.4)^†^ | 50% | Facial:  4.0 (1–6)^†^  Shoulder:  7.0 (5–8)^†^ | --- | --- | 15 (12–19)^†^  75% pts: <4 repeats |
| Mele 2019^191^ | Italy | Retrospective cohort | FSHD NS: 846 (100%) | 0% | --- | --- | --- | --- | --- | NR (1–10) repeats |
| Mongini 2015^192^ | Italy | Retrospective cohort | FSHD NS: >500 (100%) | 0% | --- | --- | --- | --- | --- | ≤8 repeats |
| Ricci 2021a^193^ | Italy | Cross-sectional/ survey | FSHD NS: 460 (100%) | 0% | --- | --- | --- | --- | --- | --- |
| Ruggiero 2017^194^ | Italy | Retrospective cohort | FSHD NS: 252 (100%) | 0% | --- | --- | --- | --- | --- | NR (33–35) |
| Ruggiero 2019^163^ | Italy | Retrospective cohort | Type 1: 187 (100%) | 0% | --- | --- | --- | --- | --- | --- |
| Ruggiero 2021^195^ | Italy | Retrospective cohort | FSHD NS: 176 (100%)  Relatives: 22 (100%) | 0% | --- | 51% | --- | --- | Patients: 6.2 ± 2.8^‡^  Relatives: 5.3 ± 2.7^‡^ | --- |
| Tabata 2021^184^ | --- | Retrospective cohort | FSHD NS: 32 (100%) | 0% | --- | --- | Early onset:  3.0 (0–4)^§^  Classic onset: 9.0 (3–17)^§^ | --- | --- | Early onset: NR (1–3) repeats  Classic onset: NR (2–4) repeats |
| FULL TEXT ARTICLES | | | | | | | | | | |
| Alavi 2018^162^ | Iran | Prospective cohort | FSHD NS: 43 (100%) Type 1: 40 (93%) | 0% | 34.0 ± 12.1 | 49% | 15.1 ± 11.6 | --- | 7.8 ± 5.1 | 93% pts: <32  (2–7 repeats) |
| Banerji 2020^172^ | UK | Retrospective cohort | Type 1: 643 (100%) | NS^*^ | 49.8 (8–87) | 51% | --- | 10.5 (0–39) | --- | 18% pts: >28  18% pts: 21–28  19% pts: <21  46% pts: unknown |
| Brouwer 1994^177^ | Netherlands | Case series | FSHD NS: 9 (100%) | 0% | --- | --- | --- | --- | --- | --- |
| Dijkstra 2021^122^ | Netherlands | Prospective cohort | Type 1: 28 (100%) | 100% | 10.8 ± 4.6 | 60% | 6.9 ± 5.1 | --- | Early onset: 40 | 5 ± 2 repeats |
| Dorobek 2015^178^ | Poland | Prospective cohort | Type 1: 22 (100%) | NS^*^ | NR (3–38) | --- | NR (1–10) | --- | NR (0.5–3.5) / NR (285–1750)^‖^ | NR (9–19)  (1–4 repeats) |
| Eren 2020^73^ | --- | Prospective cohort | FSHD NS: 29 (100%) | NS^*^ | 34.5 (13–73) | 48% | --- | --- | --- | --- |
| Evangelista 2016^169^ | UK | Narrative summary | FSHD NS: 518 (100%) Type 1: 475 (92%)  Type 2: 9 (2%) | 0% | 47.8 ± 16.1 | 50% | --- | --- | --- | --- |
| Felice 2005^170^ | US | Retrospective cohort | FSHD NS: 41 (100%) | 0% | --- | --- | --- | --- | --- | --- |
| Giacomucci 2020^168^ | Denmark  Italy  Netherlands  Spain  UK | Prospective cohort | Type 2: 34 (100%) | 0% | 49.0 ± 13.2 | 32% | --- | --- | 3.3 (0.5–5.0) | --- |
| Goselink 2018a^124^ | Netherlands | Cross-sectional/ survey | FSHD NS: 32 (100%) | 100% | 12.0 (4–17) | 6% | 6.7 | --- | 2.5 (0.0–6.0) | --- |
| Goselink 2018b^180^ | Netherlands | Cross-sectional/ survey | Type 1: 7 (78%)  Type 2: 2 (22%) | NS^*^ | 17.0 ± 9.0^§^ | --- | --- | --- | BL: 6.3  FU: 8.0 ± 2.0 (Max. 10) / 12^‡^ | 3 (2–5) repeats |
| Goselink 2019^179^ | Netherlands | Prospective cohort | Type 1: 83 (100%) | 0% | ---*** | ---*** | ---*** | ---*** | ---*** | ---*** |
| Guien 2018^167^ | France | Retrospective cohort | Type 1: 606 (95%)  Type 2: 32 (5%) | 0% | --- | --- | --- | --- | --- | --- |
| Hassan 2012^187^ | US | Case series | FSHD NS: 7 (100%) | 0% | --- | 43% | 37.0 (18–63) | --- | --- | --- |
| Korf 1985^181^ | US | Case series | FSHD NS: 6 (100%) | 100% | --- | 67% | --- | --- | --- | --- |
| Mah 2018^182^ | Australia  Canada  Europe  US | Cross-sectional/ survey | Type 1: 52 (100%) | NS^*^ | 22.9 ± 14.7 | 60% | Facial: 3.8 ± 2.9  Shoulder: 7.3 ± 4.2 | 18.9 ± 14.5 | 7.1 ± 4.1 | 18 ± 7  (3 ± 2 repeats) |
| Mair 2017^166^ | Germany | Retrospective cohort | Type 1: 6 (55%)  Type 2: 5 (45%) | 0% | NR (20–68) | 73% | NR (20–58) | NR (0.5–30) | --- | NR (20–38) |
| Moreira 2017^185^ | UK | Retrospective cohort | Type 1: 97 (97%)  Type 2: 3 (3%) | 0% | 48.0 ± 16.7 | 46% | FSHD1:  23.8 ± 15.7 FSHD2:  39.0 ± 21.8 | 22.8 ± 15.0 | 46 pts: <3.5  54 pts: ≥3.5 | --- |
| Qiu 2020^188^ | China | Case-control | FSHD NS: 35 (100%) | NS^*^ | 45.0 (15–65)^§^ | --- | 17.0 (8–35)^§^ | 20.0 (3–50)^§^ | 0 (0–467)^§‖^ | 3 (2–5)^§^ repeats |
| Ricci 2016^196^ | Italy | Methodological study | FSHD NS: 106 (100%) | 0% | --- | --- | --- | --- | --- | NR (11–38)  (1–9 repeats) |
| Ricci 2019^171^ | UK | Retrospective cohort | Type 1: 642 (100%) | 0% | --- | --- | --- | --- | --- | --- |
| Rieken 2021^165^ | Multinational Canada  US | Retrospective cohort | Type 1: 664 (94%)  Type 2: 39 (6%) | 0% | --- | --- | --- | --- | --- | FSHD1: 6 (4–7) repeats^†^  FSHD2: 15 (12-22) repeats^†^ |
| Sacconi 2019^164^ | France | Non-randomized trial  NCT01970735 | FSHD NS: 103 (100%) Type 1: 57 (55%)  Type 2: 20 (19%) | 0% | 56.2 ± 14.8 | 48% | 32.2 ± 16.9 | 24.0 ± 15.3 | --- | --- |
| Statland 2015^160^ | US | Cross-sectional | Type 1: 74 (100%) | 0% | --- | --- | --- | --- | --- | --- |
| Steel 2019^183^ | UK | Case series | Type 1: 18 (100%) | 100% | 13.8 (8–19) | 61% | 9 pts: <5  1 pt: 5–10  8 pts: 10–15 | --- | --- | ≤14 or ≥21 |
| Tawil 1996^186^ | US | Cross-sectional | FSHD NS: 157 (100%) | 0% | --- | --- | --- | --- | --- | --- |
| Vercelli 2021^159^ | Italy | Retrospective cohort | FSHD NS: 141 (100%) index cases and 105 carrier relatives | 0% | Index cases: 46.1 ± 14.2  Carrier relatives:  38.3 ± 15.6 | Index cases:  56 (40%) Carrier relatives: 54 (51%) | Female:  29.6 ± 14.3  Male:  24.2 ± 16.4 | Index cases: 20.5 ± 14.3  Carrier relatives: 10.0 ± 11.5 | Index cases: 6.3 ± 3.3^‡^  Carrier relatives: 1.8 ± 2.5^‡^ | Index cases:  12 pts: 1–3 repeats  37 pts: 4–5 repeats  41 pts: 6 repeats  29 pts: 7–8 repeats  22 pts: 9–10 repeats |
| Yasukohchi 1988^197^ | Japan | Case study | FSHD NS: 2 (100%) | 100% | --- | 1 (50%) | --- | --- | --- | --- |

Data are presented as mean ± SD or mean (range), unless otherwise specified.

BL, baseline; CCS, clinical severity score; FSHD, facioscapulohumeral muscular dystrophy; FU, follow-up; IQR, interquartile range; MMT, manual muscle testing; MRC, Medical Research Council; NR, not reported; NS, not specified; pts, patients; SD, standard deviation; UK, United Kingdom; US, United States.

^*^Not explicitly reported.

^†^Data presented as median (IQR).

^‡^FSHD clinical score.

^§^Data presented as median ± SD or median (range).

^‖^Age-corrected CSS.

^¶^MRC sum score (0–50).

^**^MMT score.

***Reported by subgroup only.

#### 7.8 Detailed tables for “Diagnosis”

Table 38. Study and population characteristics of “Diagnosis” studies

| **Author, year** | **Country** | **Study design** | **FSHD population type: n (%)** | **% pediatric** | **Age (years)** | **% female** | **Age at onset (years)** | **Disease duration (years)** | **CSS** | **D4Z4 pairs (kb)** |
| --- | --- | --- | --- | --- | --- | --- | --- | --- | --- | --- |
| CONFERENCE ABSTRACTS | | | | | | | | | | |
| Guruju 2022^199^ | --- | Prospective cohort | --- | NS^*^ | 42.0 (3.5–66)^†^ | 44% | --- | --- | --- | --- |
| Larsen 2014a^208^ | --- | Prospective cohort | FSHD NS: 95 (100%) | 0% | --- | --- | --- | --- | --- | --- |
| Larsen 2014b^209^ | --- | NR | FSHD NS: NR (100%) | 0% | --- | --- | --- | --- | --- | --- |
| Skoblov 2019^205^ | --- | Prospective cohort | FSHD NS: 7 (100%) | 0% | --- | --- | --- | --- | --- | --- |
| Storch 2017^211^ | --- | Case study | FSHD NS: 1 (100%) | 0% | --- | --- | --- | --- | --- | --- |
| Walker 2022^218^ | UK | Retrospective cohort | Type 1: 1,042 (97%)  Type 2: 32 (3%) | 0% | --- | --- | --- | --- | --- | --- |
| Zernov 2020^206^ | Russia | Prospective cohort | FSHD NS: 64 (100%) | 0% | --- | --- | --- | --- | --- | --- |
| FULL TEXT ARTICLES | | | | | | | | | | |
| Bakker 1996^215^ | Netherlands | Narrative | FSHD NS: 90 (100%) | 0% | --- | --- | --- | --- | --- | --- |
| Fabry 2022^212^ | France | Retrospective cohort | Type 1: 19 (100%) | 0% | 47.7 ± 18.0 (20–72) | 53% | --- | 13.1 ± 9.1 | 1.2 ± 0.4^‡^ | --- |
| Goto 2006^204^ | Japan | Cross-sectional | FSHD NS: 105 (100%) | 0% | --- | --- | --- | --- | --- | --- |
| Larsen 2015^210^ | Germany | Retrospective cohort | FSHD NS: 95 (100%) | 0% | --- | --- | --- | --- | --- | --- |
| Lemmers 2012^203^ | --- | Narrative summary | --- | 0% | --- | --- | --- | --- | --- | --- |
| Monforte 2022^213^ | Italy | Retrospective cohort | Type 1: 187 (100%) | 0% | 45.5 ± 15.5 | 48% | --- | --- | --- | --- |
| Nicolau 2021^207^ | --- | Narrative summary | --- | 0% | --- | --- | --- | --- | --- | --- |
| Orrell 1999^200^ | US | Case-control | FSHD NS: 113 (100%) | NS^*^ | NR (11–88) | 46% | --- | --- | --- | --- |
| Padberg 1991^216^ | -- | Narrative | --- | 0% | --- | --- | --- | --- | --- | --- |
| Parisien 2020^198^ | US | Case study | FSHD NS: 1 (100%) | 100% | 13.0 | 100% | --- | --- | --- | --- |
| Podnar 2009^214^ | Slovenia | Case-control | FSHD NS: 31 (100%) | 0% | 43.0 (22–77)^†^ | 45% | --- | --- | --- | --- |
| Qiu 2020^188^ | China | Case-control | FSHD NS: 35 (100%) | NS^*^ | 45.0 (15–65)^†^ | --- | 17.0 (8–35)^†^ | 20.0 (3–50)^†^ | NR (0–467)^§^ | 3 (2– 5)^†^ repeats |
| Rieken 2021^165^ | Multinational  Canada  US | Retrospective cohort | Type 1: 664 (94%)  Type 1: 39 (6%) | 0% | --- | --- | --- | --- | --- | FSHD1: 6 (4–7)^‖^ repeats  FSHD2: 15 (12 -22)^‖^ |
| Rodrigues 2015^217^ | New Zealand | Narrative summary | FSHD NS: 24 (100%) | 0% | --- | --- | --- | --- | --- | --- |
| Steel 2019^183^ | UK | Case series | Type 1: 18 (100%) | 100% | 13.8 (8–19) | 61% | 9 pts: <5  1 pt: 5–10  8 pts: 10–15 | --- | --- | ≤14 or ≥21 |
| Upadhyaya 1997^202^ | --- | Case-control | FSHD NS: 130 (100%) | 0% | --- | --- | --- | --- | --- | --- |
| Yanoov-Sharav 2012^201^ | Israel | Narrative summary | FSHD NS: 32 (100%) | 0% | --- | 28% | --- | --- | --- | --- |

Data are presented as mean ± SD or mean (range), unless otherwise specified.

CCS, clinical severity score; FSHD, facioscapulohumeral muscular dystrophy; IQR, interquartile range; NR, not reported; NS, not specified; pt, patient; SD, standard deviation; UK, United Kingdom; US, United States.

^*^Not explicitly reported.

^†^Data presented as median (range).

^‡^Modified Rankin scale.

^§^Age-corrected CSS.

^‖^Data presented as median (IQR).
